# A planetary health solution for disease, sustainability, food, water, and poverty challenges

**DOI:** 10.1101/2022.08.02.22278196

**Authors:** Jason R Rohr, Sidy Bakhoum, Christopher B Barrett, Andrew J Chamberlin, David J Civitello, Molly J Doruska, Giulio A De Leo, Christopher J E Haggerty, Isabel Jones, Nicolas Jouanard, Amadou T. Ly, Raphael A Ndione, Justin V Remais, Gilles Riveau, Alexandra Sack, Anne-Marie Schacht, Simon Senghor, Susanne H Sokolow, Caitlin Wolfe

## Abstract

Global health and development communities lack sustainable, cost-effective, mutually beneficial solutions for infectious disease, food, water, and poverty challenges despite their regular interdependence worldwide^1–7^. Here, we show that agricultural development and fertilizer use in west Africa increase the devastating tropical disease schistosomiasis by fueling the growth of submerged aquatic vegetation that chokes out water access points and serves as habitat for snails that transmit *Schistosoma* parasites to >200 million people globally^8–10^. In a randomized control trial where we removed invasive submerged vegetation from water points, control sites had 124% higher fecal *Schistosoma* reinfection rates in schoolchildren and lower open water access than removal sites without any detectable long-term adverse effects of the removal on local water quality or freshwater biodiversity. The removed vegetation was as effective as traditional livestock feed but 41-179 times cheaper and converting the vegetation to compost yielded private crop production and total (public health plus private benefits) benefit-to-cost ratios as high as 4.0 and 8.8, respectively. Thus, we provide an economic incentive – with important public health co-benefits – to maintain cleared waterways and return nutrients captured in aquatic plants back to agriculture with great promise of breaking poverty-disease traps. To facilitate targeting and scaling of this intervention, we lay the foundation for using remote sensing technology to detect snail habitat. By offering a rare, profitable, win-win innovation for food and water access, poverty, infectious disease emergence, and environmental sustainability, we hope to inspire the interdisciplinary search for other planetary health solutions^11^ to the numerous and formidable, co-dependent global grand challenges of the 21st century.

---

Infectious diseases and insufficient access to water and food resources burden poor communities worldwide. For example, (*i*) ∼264 million people are undernourished in Africa^12^, (*ii*) Sub-Saharan Africa loses 40 billion hours per year collecting water^13^, and (*iii*) 75% of deaths in low-income countries can be attributable to infectious disease^5^. Disease, food, and water challenges intersect in many ways, such as through widespread poverty-disease traps, whereby disease exacerbates malnutrition, stunts physical and cognitive development, and reduces educational attainment, labor supply, and incomes, all of which impede economic growth that can improve sanitation and access to energy, water, education, and health care^1–4^. Despite their interdependence, disease, food, and water challenges are typically addressed independently by governmental and non-governmental organizations, practitioners, and researchers. Consequently, few convincing examples exist of sustainable win-win planetary health (a transdisciplinary field that searches for sustainably beneficial actions for natural systems and human health^11^) interventions with real promise of breaking poverty-disease traps^3,4^. Even more rarely are the economic costs and benefits of win-win planetary health interventions quantified to demonstrate their cost-effectiveness and thus potential for widespread adoption.

An example of a neglected tropical disease with clear links to water and food production is human schistosomiasis^5–7^, the world’s second most burdensome parasitic human disease after malaria with >800 million people at risk of infection^8–10^. Schistosomiasis is caused by snail-transmitted flatworms that penetrate human skin, reinforces poverty, devastates childhood health, and defies control efforts, because even when humans are provided drugs to treat these infections, they quickly get re-infected when they return to snail-infested waterbodies^8–10^. In Senegal, the site of our experiment, schistosomiasis prevalence in children often rebounds to 70-90% within a year after drug treatment, and >99% of host snails are captured in *Ceratophyllum demersum*, an aquatic plant that (*i*) has a mutualistic relationship with snails, (*ii*) is found throughout Africa, Southeast Asia, and Latin America where schistosomiasis is endemic, and, along with other invasive aquatic plants, (*iii*) chokes out waterways impeding access to open water needed for washing clothes, irrigation, and cooking^14–16^. Additionally, *C. demersum* is well documented to proliferate in the presence of fertilizer runoff^17–19^.

We hypothesized that agricultural development in west Africa and associated fertilizer use has led to increases in schistosomiasis by increasing submerged aquatic vegetation and attached algae that are the habitat and food for snails, respectively (see Table S1 for evidence to support this pathway, Fig. 1a,b). Based on this hypothesis, we predicted that we could decrease human schistosomiasis and increase open water access by removing this vegetation from water access points. We also predicted that we could increase food production by returning the nutrients captured in this vegetation back to agriculture. This would help reduce nitrogen and phosphorus pollution by closing the nutrient loop^20^, while simultaneously providing an economic incentive to maintain cleared waterways. This would provide a mutually beneficial planetary health solution for sustainability, disease reduction, water access, agricultural development, and poverty that could facilitate adoption of this innovation.

**Fig. 1.**
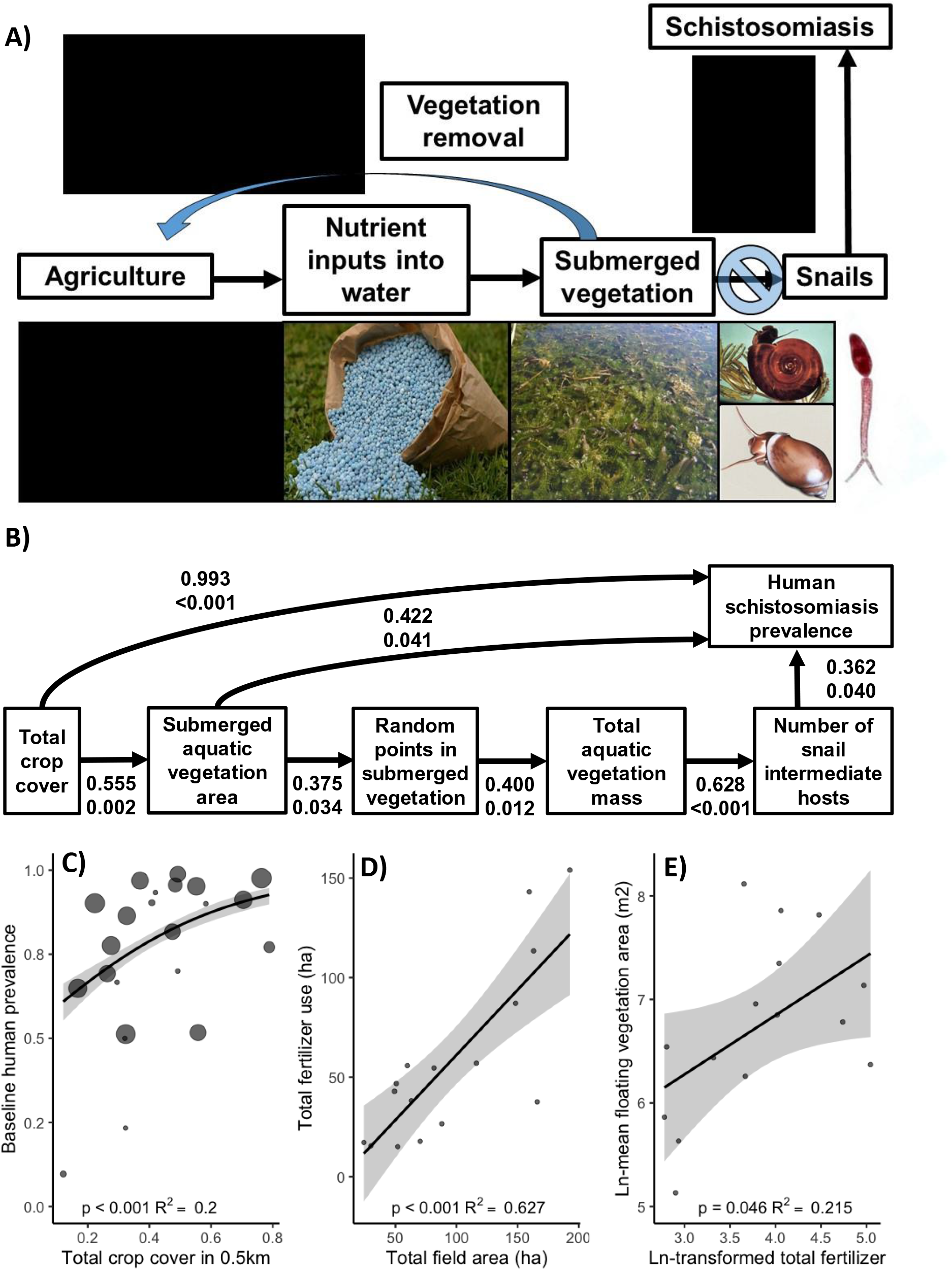
Hypothesized and observed associations between agriculture and human schistosomiasis in Senegal. **(A)** Hypothesized pathway by which agriculture affects schistosomiasis and the proposal to return nutrients captured in aquatic vegetation back to agriculture to disrupt the pathway promoting high levels of human schistosomiasis. **(B)** Best fitting path model for the observed associations between agriculture and human schistosomiasis infection prevalence in schoolchildren (*n*=23 sites; see Methods). Positive effects are shown as black arrows and the standardized effect sizes followed by probability values are provided under each path. The distribution of sampling points among different habitat types (open water, submerged or floating vegetation, and emergent vegetation) at water access points was proportional to the surface area of each habitat type (see Methods). Thus, total surface area of submerged or floating vegetation increased sampling effort of this habitat. This, in turn, increased estimates of its mass, which represents the primary three-dimensional habitat for snails. The final path model was a good fit to the data (Test of directed separation: *C*16 = 25.2, *p* = 0.067; SI Appendix Text S1). Agriculture was positively associated with the **(C)** schistosomiasis prevalence in children (the size of points is proportional to the number of children tested, the denominator in the binomial regression model) and **(D)** fertilizer use. **(E)** Although fertilizer use was positively associated with the amount of aquatic vegetation (*Rs* = 0.56, *p* = 0.034), this pathway was not included in the path model because fertilizer use was only available for a subset of sites (*n*=16). Faces were covered or darkened to protect identities. Black boxes cover photographs of people, which are not allowed on medRxiv. Contact the corresponding authors if you would like the complete figure.

To test for associations among agriculture and schistosomiasis, we conducted collaborative research with 23 communities in the St. Louis-Richard Toll region of Senegal (Fig. S1-S2) and quantified the amount of (*i*) agricultural fields, fertilizer, and other agrochemical applications within a 0.5-km radius from the center of each site (Table S2-5), (*ii*) submerged aquatic vegetation in water access points (Fig. S3), (*iii*) snails that transmit human *Schistosoma* worms (Table S6), and (*iv*) human *Schistosoma* infections in ∼1,700 schoolchildren (i.e., using a combination of cross-sectional surveys and cohort studies; see Methods and Tables S7-8). Regression analyses (Table S9-14) revealed that each km^2^ of agricultural cover around a site was associated with an increase of 3.1 schistosomiasis cases in schoolchildren per community (95% CI: 2.4-3.9, Fig. 1c), and this pattern was robust to different methods of quantifying agriculture and controlling for freshwater habitat around sites and area of water access points (see SI Appendix Text S1). The best fitting path model provided support for our hypothesized indirect pathway (Table S15-19, Fig. 1a). Total area of crops was linked positively with fertilizer use (Fig. 1d), which was positively associated with aquatic vegetation at water access points (Fig. 1e). Submerged aquatic vegetation was itself positively correlated with snail abundance (Fig. 1b), which in turn was positively associated with the prevalence of *Schistosoma* in schoolchildren (Table S18-19, Fig. 1b). The pathway from agriculture directly to prevalence was also significantly positive (Table S18-19, Fig. 1b), indicating that agriculture is somehow associated with schistosomiasis beyond this identified indirect pathway.

Given the positive association between agriculture and schistosomiasis that seems to be mediated by aquatic vegetation, we predicted that we could disrupt this relationship and increase open water access by removing this vegetation from water access points. To test this hypothesis, we implemented a three-year randomized controlled trial in 16 communities in Senegal (Fig. S4-5), quantifying effort to remove vegetation and abundance of snails, aquatic vegetation, open water, and *Schistosoma* infections in >1,400 schoolchildren before and after vegetation removal in half of the sites (Table S20). All infected schoolchildren received praziquantel annually to treat *Schistosoma* infections (Table S21); thus, we tracked reinfection rates (see Methods; Fig. S5).

We removed an estimated 433 metric tons of aquatic vegetation and removed significantly less vegetation when revisiting sites semi-quarterly relative to the first visit (*p <* 0.001, Table S22-S23, Fig. 2a). Consequently, the labor costs of removing vegetation dropped significantly relative to the first removal (*p* < 0.001) and remained relatively constant thereafter (Fig. S6). We sampled 7,833 snails and vegetation removal was associated with an 8-fold reduction in snails in the following year (time*treatment: *p <* 0.001, Table S24-25, Fig. 2c; see Tables S26-S28 for details on each snail species). The more vegetation removed, the more *Biomphalaria* and *Bulinus* snails we removed (*bivariate correlation rho =* -0.46, *p* < 0.001, Fig. 2b) and the greater the area of open water access we provided to community members (time*treatment: *p =* 0.026). We encourage future studies to quantify how much water collection time is saved as a result of this increased open water.

**Fig. 2.**
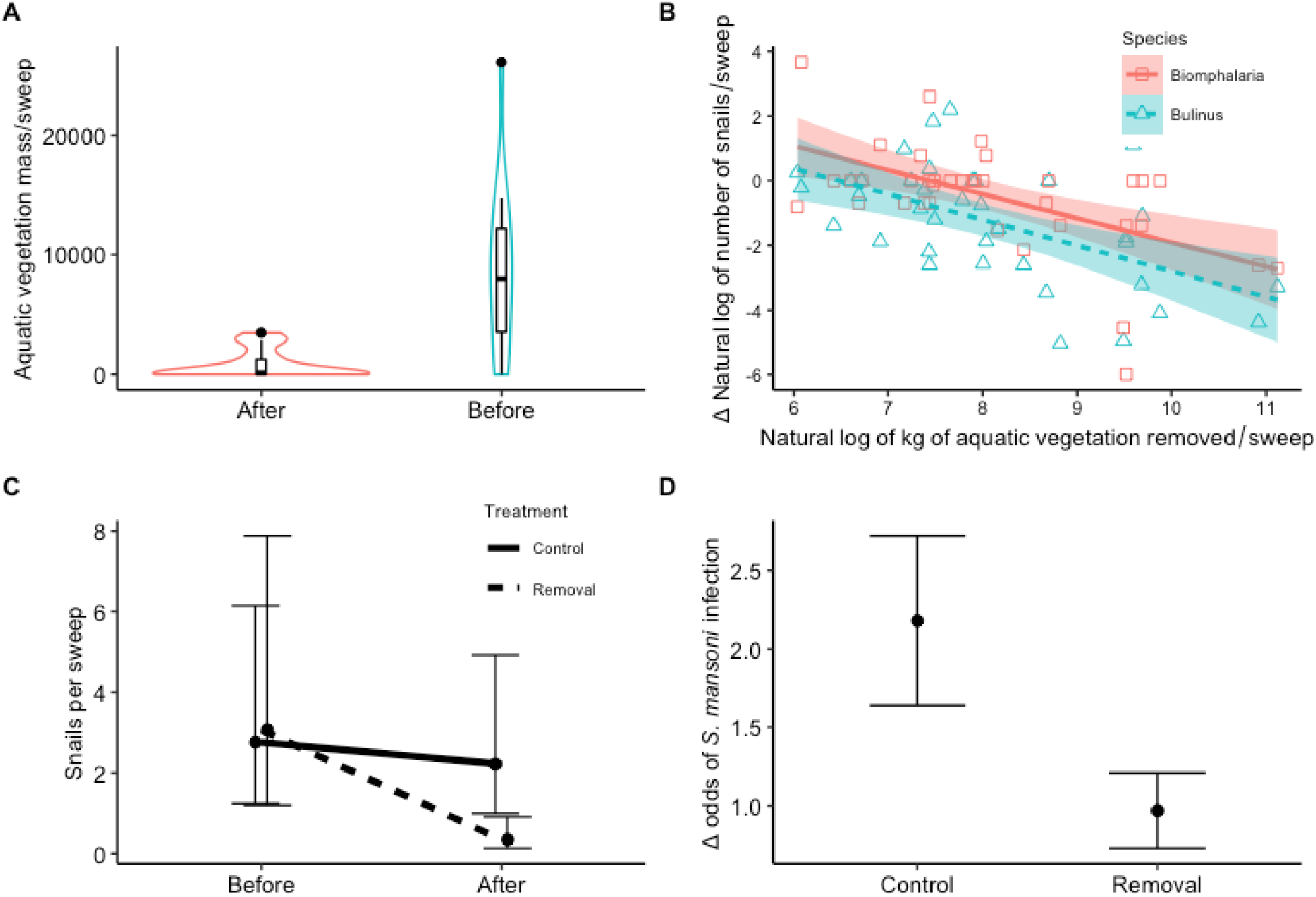
The relationship between vegetation removal, snails, and probability of infection. **(A)** Mean (95% CIs) non-emergent vegetation mass (g) per sweep removed the first and subsequent quarterly vegetation removal events at 16 water access points located in 8 sites. **(B)** Association between aquatic vegetation removed (ln(kg)) at water access points and the change (Δ) in *Bulinus* spp. and *Biomphalaria* spp. snails (ln(snails +1)) collected per sweep before and after the initial vegetation removal. **(C)** Mean (95% CIs) snails per sweep sampled at the same locations before and after the initial removal event. **(D)** The change (Δ) in the community-school mean (+SE) probability of infection with *S. mansoni* in removal and control sites from before as compared to after the intervention.

The baseline fecal *Schistosoma* average egg count among infected schoolchildren was 161 eggs per gram of stool (Tables S29), characterized as moderate intensity by the World Health Organization^21^. After the intervention, the percent of schoolchildren infected with fecal schistosomiasis in vegetation-removal sites was 23.5% (19.8-27.2%), compared to 31.5% (27.2-35.8%) in control sites (Tables S29). Hence, vegetation removal significantly reduced schistosomiasis reinfection rates (*p =* 0.02), amounting to a 124% higher odds (i.e., odds ± SE = 2.24 ± 0.79) of being infected in control than vegetation-removal sites (Table S31-32, Fig. 2d). Additionally, we found no evidence of changes in human-water contact following vegetation removal (Tables S33-S38). Note that any induced increase in human-water contact should attenuate estimated effects on infection rates.

In only three vegetation-removal sites was the total difference in vegetation mass sampled after versus before the removal <3kg per sweep, because these sites either had little vegetation to remove, substantial regrowth between visits, or vegetation blown in from deeper waters. Only in these three intervention sites did fecal schistosomiasis not decline, further corroborating the benefit of aquatic vegetation removal, but also suggesting that shorter removal intervals may be necessary when vegetation regrowth is particularly rapid or *Schistosoma* transmission is extremely high^22,23^ (see Table S39-40 and SI Appendix Text S2 for results and discussion on urinary schistosomiasis). Our results are consistent with the results of smaller-scale vegetation removal studies^22,24^ that focused only on *Schistosoma*-harboring snails but not human infections^25^ or were confounded with molluscicide applications^22^, and corroborate predictions from mathematical models that schistosomiasis prevalence should decline faster with vector control than drug treatments alone^26–28^.

Importantly, there were no adverse effects of removing this overgrowth of invasive vegetation on water quality or chemistry (Table S41). Given that these water access points are contiguous with the river or lake, the reduction in vegetation, which serves as habitat for freshwater biodiversity (such as snails), was highly localized and temporary, indicating that the vegetation clearing we practiced and promote likely has no long-term adverse effects on the aquatic ecosystem as a whole. We hypothesize that the limited scale of invasive vegetation removal – only at water access points – may obviate any ecosystem effects.

Because aquatic plant overgrowth appears to be at least partially caused by runoff from agriculture, we hypothesized that we could profitably improve food production by returning the nutrients captured in the removed plants back to agriculture, thus reducing nitrogen and phosphorus pollution by closing the nutrient loop^20^ (Fig. 1). To test this hypothesis, we collaborated with local farmers and ranchers to evaluate whether aquatic vegetation could be cost-effectively converted to compost to increase crop yields (Fig. S7, Fig. 3a-d) and/or used as livestock feed (Fig. 3e-g) (see Methods). The compost was reasonably high in moisture, nitrogen, and phosphorous (Table S42), increased both onion and pepper production (total weight) independent of whether the compost was tilled in the soil or applied with fertilizer (Tables S43-45, Fig. 3c, d), and onion rot was significantly greater in plots with both fertilizer and compost than compost alone, further supporting the use of compost as a substitute for fertilizer (*p* < 0.001). Depending upon the compost treatment arm, crop, wage estimates, and crop valuation, conservative private benefit-to-cost ratios ranged from 2.7-4.0 and were always significantly greater than one, indicating that conversion of removed vegetation to compost was highly profitable (Table S46, see SI Appendix Text S3 for additional details on the economic analyses).

**Fig. 3.**
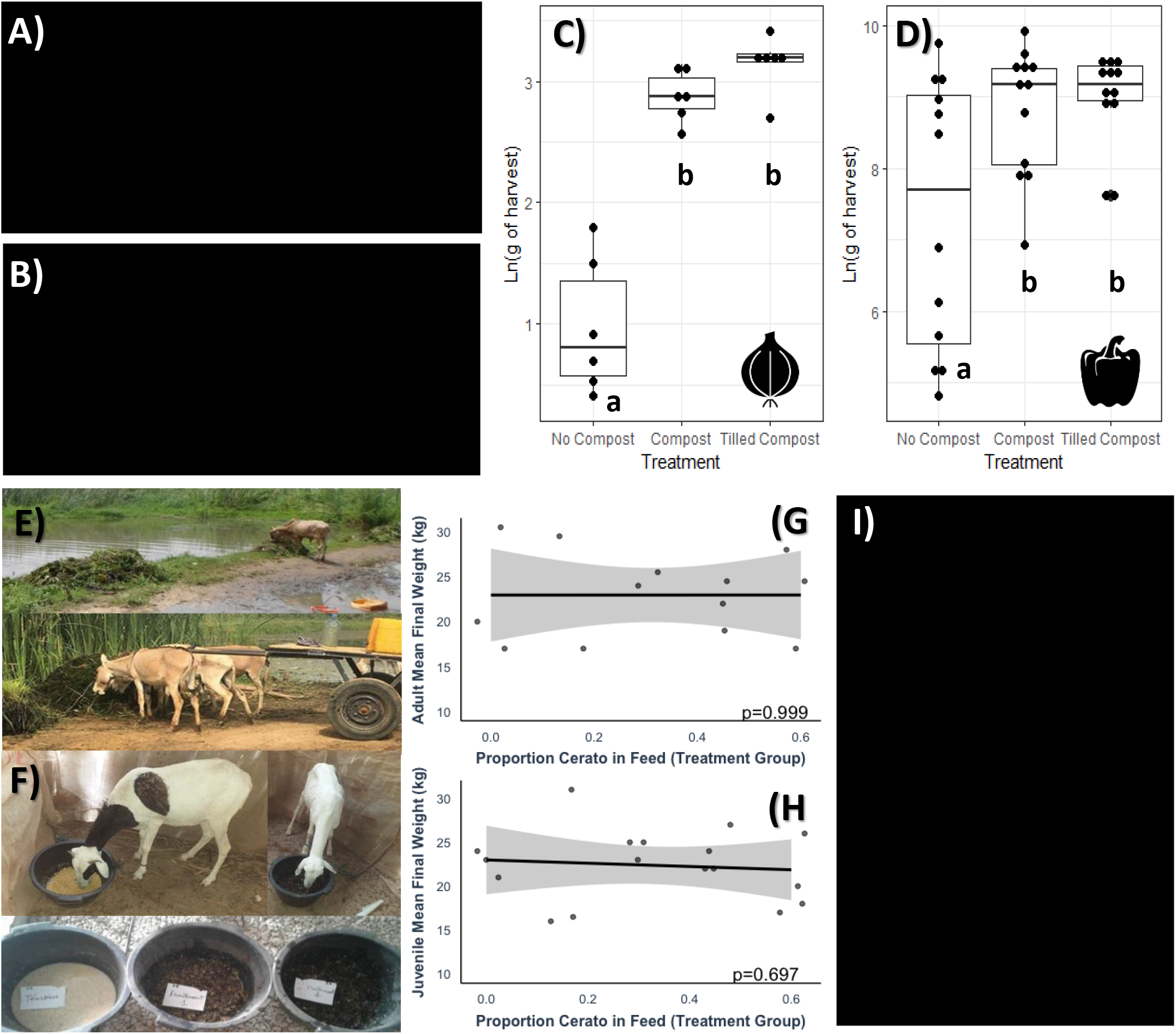
Use of nuisance aquatic vegetation as compost and livestock feed to increase food production and profits. **(A)** Photographs of aquatic vegetation and compost piles. **(B)** Photographs of pepper plots receiving compost, fertilizer, and tilling treatments and the farmer collecting data for the project. The compost increased **(C)** onion and **(D)** pepper production independent of the cross-randomized tilling or fertilizer treatments (data shown averages across fertilizer treatment because there was no effect beyond that of compost). Treatments that share letters are not significantly different from one another (*p*>0.05) based on a Tukey’s multiple comparison test. The middle line of the box is the median, and the box represents the 25th to 75th percentile, and the whiskers extend to 1.5 the distance of the interquartile range. **(E)** Photographs of cattle and donkey readily consuming the removed aquatic vegetation, which is almost exclusively *Ceratophyllum demersum*. Goats refused to eat *C. demersum*. **(F)** Sheep and feeds used in our sheep trials. Final weights of **(G)** adult and **(H)** juvenile sheep were similar to controls when their normal diet was substituted iso-calorically with *C. demersum* up to 60%. Shown are regression lines, jittered points, and 95% confidence bands. Age class of the sheep had no significant effect on sheep weight (*F*_1,28_=0.14, *p*=0.71). **(I)** Voluntary community engagement in the vegetation removal process. Personal protective equipment was refused by members of the community. Black boxes cover photographs of people, which are not allowed on medRxiv. Contact the corresponding authors if you would like the complete figure.

Sheep, donkeys, and cattle, but not goats, readily consumed the removed aquatic vegetation suggesting that it might be useful as livestock feed (Fig. 3e-g). Using 17 female juvenile sheep and 13 female adult sheep, we iso-calorically substituted their typical purchased feed (a mix of peanut straw and pellets for juveniles, cornmeal for adults) with the removed aquatic vegetation (*C. demersum*) and tracked growth rates (Table S47). The vegetation was first dried and ground with a pestle in an effort to kill any helminth eggs or cysts and then reconstituted with water (see Methods). We found no significant effect of any feed substitution level, up to the 60% maximum, on sheep weight gain or growth in either age group (Tables S48-49, Fig. 3f). The cost of aquatic vegetation per calorie is 0.10 – 0.20 FCFA ($0.0002 – $0.0004 USD), whereas the cost per calorie of peanut straw and cornmeal are 13.39 – 17.86 FCFA ($0.02 – $0.03 USD) and 8.29 FCFA ($0.01 USD), respectively. Thus, feeding the sheep aquatic vegetation is 41 to 179 times cheaper than purchasing traditional feed for sheep (Tables S50-51). Using the removed vegetation as compost had a higher private benefit-to-cost ratio than using it as livestock feed (Table S46, S50-53), consistent with the observation that sheep graze freely, especially when forage is available, obviating the need to purchase feed. Including a highly conservative estimate for the public health benefit of reducing schistosomiasis (see Methods, Table S54) adds 15-22% to the gross private benefits of using the vegetation as compost, resulting in total benefit-to-cost ratios of 3.1-8.8 (Table S52), while using vegetation as livestock feed sharply reduces costs (without loss of benefits) during those occasional periods when farmers supplement grazing with purchased feed.

Although the benefit-to-cost ratios are already high, we expect them to increase with time as intervention costs decline and benefits rise. Average total intervention costs should fall when fixed costs are spread to produce more compost or feed over time. Additionally, the benefits should increase through time as future studies evaluate (*i*) economic benefits of increased open water access (not included in our calculations), (*ii*) health gains to non-school-aged people, (*iii*) gains in educational attainment and cognitive development (for which DALYs do not account), and (i*v*) reduced transmission outside the community. Additionally, by 2100, global population is projected to reach 10-11 billion people^6^, and most of this population growth will occur in Africa and Southeast Asia^6^, increasing the total number of people at risk of schistosomiasis. To meet projected global food demand, agricultural production must intensify^29^. Sharp expansion of inorganic fertilizer use, however, could fuel further increases in aquatic vegetation and schistosomiasis risk^6^, hence the importance of introducing circularity through returning nutrients captured in aquatic vegetation back to soil in place of fertilizers.

We observed prosocial behavior in the form of regular, voluntary community engagement in the vegetation removal process after we educated community members on the expected public health benefits of this intervention (Fig. 3h). Nonetheless, relying purely on voluntary labor to clear the vegetation will almost surely result in suboptimal vegetation clearing and higher schistosomiasis prevalence relative to the condition where they perceive and act on private benefits. By converting the public nuisance, aquatic vegetation, into a profitable agricultural input, we provide a private economic incentive to maintain cleared waterways, generating public health co-benefits along with higher private incomes. Although it remains to be tested, we hypothesize that communicating these private benefits to community members will increase vegetation removal relative to communicating the public health benefits alone, which could help marginalized communities escape poverty-disease traps^4^. Importantly, competition for private benefits of a public resource (aquatic vegetation) will likely require communities to manage access to the vegetation and thus prevent conflict, a potential adverse side effect of this innovation.

Given that our intervention offers a mutually beneficial solution for several societal challenges (Fig. 2-3) and there was some level of interest in adopting the intervention (Fig. 3h), we began developing a scaling plan. We hypothesized that we could use satellite imagery to identify snail habitat and thus potential schistosomiasis hotspots, which would then facilitate targeting our intervention to where it is needed most. Although there is a substantial amount of work left to hone our remote sensing approach, our preliminary analyses allowed us to reliably discriminate submerged vegetation from emergent vegetation and open water (see Methods and^30^; Fig. 4). This suggests that there is considerable potential to use remote sensing to facilitate targeting and scaling of this intervention. Scaling of this intervention could also be further assisted by investigations of (*i*) vegetation removal intervals, spatial scales, compliance, and mechanization, (*ii*) other economic uses of the vegetation, and (*iii*) the efficacy of the intervention in other parts of the world.

**Fig. 4.**
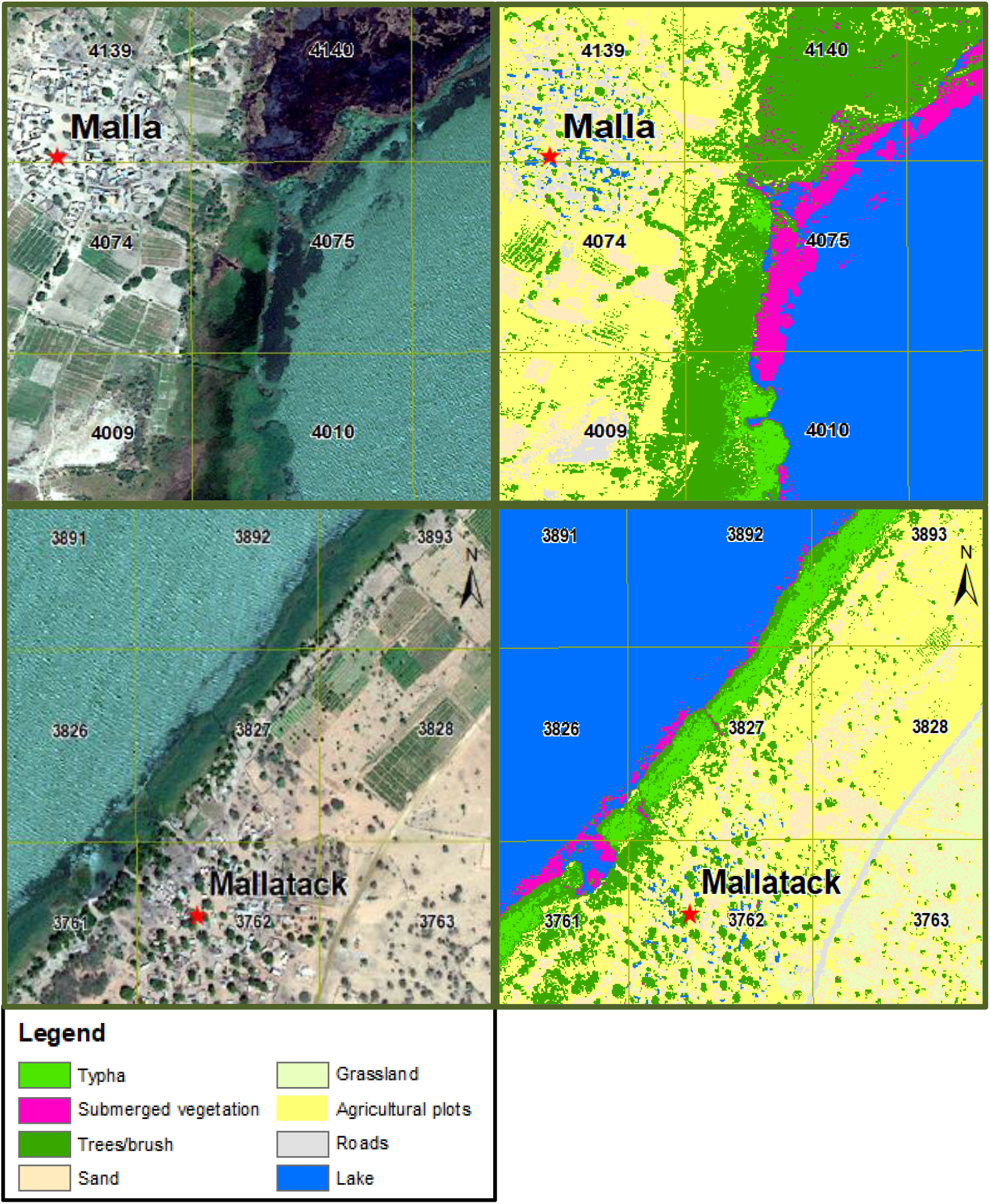
Ability to discriminate among submerged vegetation, emergent vegetation (*Typha*), and open water with adjustments to red, green, and blue levels only. Fuchsia = high densities of *Ceratophyllum demersum*, which is snail habitat.

We provide support for the hypothesis that agricultural development in west Africa and associated fertilizer use are increasing the devastating human disease schistosomiasis by fostering the growth of aquatic plants that function as snail habitat. By removing this vegetation and returning the nutrients captured in it back to agriculture, we offer a profitable and environmentally responsible solution to several of the most formidable, co-dependent global grand challenges of the 21st century – environmental sustainability, food and water access, poverty, and infectious disease emergence. Our solution aligns with the United Nations Sustainable Development Goals^31^ and synergistically leveraged principles of ecology, and the social, environmental, and agricultural sciences. Identifying similar planetary health innovations must be a priority and will likely require capitalizing on interdisciplinary, systems-based approaches to beneficially modifying the built and natural world and designing incentives for community-led maintenance of innovations with both public and private benefits. We hope that this project offers a prototype planetary health solution, not only for similar agricultural, health, water, and sustainability challenges, but for other co-dependent grand challenges as well.

## Data and code availability

All data and code accompany this manuscript in the supplementary materials and an RMarkdown file.

## Supporting information

Veg Removal RMarkdown

Ag. Correlation and Path Model RMarkdown

## Data Availability

All data produced in the present study are available upon reasonable request to the authors

## Acknowledgements

This research was supported by a National Institutes of Health grant (R01GM109499) to JRR, JVR, SHS, GAD, NJ, and GR. Additionally, this research was supported by grants from the National Science Foundation (EF-1241889, DEB-2109293, DEB-2017785), National Institutes of Health (R01TW010286), and the Indiana Clinical and Translational Sciences Institute to JRR; the National Institutes of Health (K01AI091864) and the National Science Foundation (EAR-1646708, EAR-1360330) to JVR, and the National Science Foundation (CNH grant # 1414102), Bill and Melinda Gates Foundation, National Institutes of Health (R01TW010286-01), Stanford GDP SEED (grant # 1183573-100-GDPAO), and SNAP-NCEAS (working group “Ecological levers for health: Advancing a priority agenda for Disease Ecology and Planetary Health in the 21st century”) to SHS and GAD.

## Author contributions

JRR conceptualized the experiments, directed the project and analyses, and wrote the majority of manuscript. CJEH and AS contributed to figure development, statistical analyses, and helped write sections of the manuscript. CJEH, CW, SB, and RAN performed field sampling, data collection, and curation of field data. AJC and IS provided the drone imagery. CW acquired the DigitalGlobe grant for the satellite imagery and conducted the remote sensing analyses. CBB and MJD performed economic analyses and contributed to writing those sections of the manuscript and to general editing. NJ, SS, and GR directed the human sampling. ATL collected human infection samples. AMS performed human data curation. NJ oversaw the livestock feed trials. DC contributed to the original idea development. JRR, CJEH, CW, AJC, and IS collected the data to compare the quadrat and sweep net sampling protocols. JRR, GAD, NJ, JVR, GR, and SHS developed the grant that funded much of this research. All co-authors contributed to manuscript editing.

## Additional information

**Supplementary information** is available for this paper at

**Correspondence and requests for materials** should be addressed to Jason Rohr.

**Peer review information** *Nature* thanks the anonymous reviewers for their contribution to the peer review of this work. Peer reviewer reports are available.

**Reprints and permissions information** is available at www.nature.com/reprints.

## Methods

### Study area and village selection

For the study testing for an association between agriculture land use and schistosomiasis, we selected 23 sites from a list of >700 sites as fully described by Wood et al.^15^. Briefly, sites were located along the Senegal and Lampsar Rivers and the shore of Lac de Guiers ([16+15′N 15+50′W](https://geohack.toolforge.org/geohack.php?pagename=Lac_de_Guiers&params=16_15_N_15_50_W_type:waterbody_region:SN)), in northern Senegal (Fig. S1). Sites were selected that had access to permanent freshwater without large-scale drainage or vegetation clearing projects that could impact snail or plant distributions, a small human population (<3,000 persons), an active school, ≤4 water points accessed by community members, and permission from the community leader for us to sample the water access points. For the vegetation removal randomized control trial, we selected 16 (Fig. S4) of these 23 sites (i.e., 7 sites were only sampled in 2017, whereas the others were sampled repeatedly from 2016-2018). The primary source of water for the communities was the local (open) water access points, as only two communities had a functional water well, and only one of those had electricity.

### Agricultural land use study

Within the 23 sites selected for the correlational study exploring the relationship between agricultural land use and schistosomiasis, we created sampling polygons at each water access point that encompassed all open water and submerged and floating vegetation, extending laterally several meters into the dense emergent vegetation border (Fig. S3). The outer limit of each sampling polygon was determined by water depths at which technicians could safely sample wearing chest waders. Technicians sampling snails or removing vegetation wore both chest waders and shoulder length gloves as personal protective equipment (PPE). During removal, PPE was worn by adult male community members who were paid an hourly rate to remove aquatic vegetation. To quantify vegetation within the polygon, we collected aerial photographs using an unmanned drone with a 12.4 megapixel camera and geo-referenced using QGIS 3.2 (https://qgis.org/en/site/) to estimate the area (m^2^) of submerged aquatic vegetation or snail habitat (Fig. S3). We summed vegetation area (m^2^) per village because the human infection data are only available at the village-level (see below). All of these procedures are also published in Wood et al.^15^.

In addition to using QGIS to map the water access points, we used QGIS to quantify agricultural cover and freshwater habitat within a 0.5, 1 and 2 km radius buffer from the center of each village applying the Food and Agriculture Organization of the United Nations (FAO) produced LANDSAT ETM (remote) images of Senegal with a spatial resolution of 30 m (Table S2-S3). The FAO data were verified by an accuracy assessment using 10m resolution Google Earth imagery and ground truthing in Senegal^32^.

Because FAO data did not include agrochemical application rates, we also conducted a household survey based on the children who were enrolled in the parasitological study. In the summer of 2016 in the 16 sites used in our randomized control trials, we conducted surveys of the extended family households (i.e., concessions) where the children lived (*n* = 663 households). The number of households per village varied with the size of the village from 16 to > 60 households. All data were self-reported by the head of the household who was asked to estimate the size of all parcels of land that were held by someone in the concession. For each parcel, we also asked whether the land was fallow/uncultivated and whether fertilizers were being used (Tables S3-S5).

### Aquatic vegetation removal randomized control trial

The 16 sites selected for the randomized control trial had a total of 32 water access points used by community members (1-4 per site), within which we created sampling polygons encompassing all open water and submerged and floating vegetation, as described above for the agricultural land use study. During each vegetation removal round, we removed submerged vegetation from all sampling polygons of the eight removal sites, with three sites starting in 2017 and eight sites in 2018 (Fig. S5). We removed vegetation quarterly for one year within a 1-2-week period for all sites (Fig. S5-S6). We estimated the mass of vegetation removed by comparing the vegetation piles on the shoreline of water points (vegetation shaken to remove water) to standardized volumes of each plant species to convert field measured volume to a total estimated weight removed (kg) (Table S22-23, Fig. S6). We also recorded the number of person hours required for each semi-quarterly vegetation removal.

### Snail sampling protocols

To sample snails in the 16 sites used in our agriculture land use assessment and randomized control trial, we created 15 random sampling points within each polygon described above, with points stratified across three potential snail habitats (open water, non-emergent vegetation, emergent vegetation) based on their proportional cover found by aerial photography and a visual estimation by technicians on the date of sampling. We exhaustively sampled each quadrat (76.2-cm length × 48.26-cm width × 48.26-cm height; area, 0.3677 m^2^) placed at these points using a 2.5 mm mesh aquatic dipnet, all as described in Wood et al.^15^. We took sample contents to the shore where we shook all non-emergent vegetation to remove the snails, weighed the vegetation using a spring scale, and identified and counted *Bulinus truncatus/globosus* spp. and *Biomphalaria pfeifferi*, which are intermediate hosts to the human schistosomes *S. haematobium,* and *S. mansoni*, respectively. We sampled for snails annually at the same GPS locations both before and after a vegetation removal at water access points of each village.

At the seven additional sites used in the agriculture land use assessment study, we used identical snail-sampling methods except that we performed a 1-m sweep at each random sampling point using a 45 x 40 cm aquatic dipnet with a 2.5 mm mesh, instead of exhaustively sampling a quadrat with a 2.5 mm mesh net. Given this minor difference in the snail sampling methodology, we performed a separate study using the quadrat and sweep net methods side-by-side at 218 random points. We compared snail counts between the two sampling methods, by performing a negative binomial regression with village as a random intercept to account for re-sampling the same village. These analyses demonstrated that both methods captured similar numbers of snails (χ^2^ = 0.76, df = 1, *p* = 0.383; Fig S8).

### Human sampling

A total of 1,479 school-aged children were treated with praziquantel at the 16 sites in April 2016 and 211 school-aged children were treated at the additional seven sites added for the agricultural land use study (Table S7-8, S29-30). Parasite identification was performed by Espoir Pour La Santé (EPLS) in Saint Louis, Senegal, using urine and fecal samples from these children to determine human infection by *S. haematobium* and *S. mansoni*, respectively. *Schistosoma haematobium* in children was quantified by urine filtration using standardized methods^33^. Two 10 mL samples were separately filtered and analyzed by different observers. *Schistosoma mansoni* infection levels were determined by collecting stool samples and using the Kato-Katz assay^34^. For the randomized control trial, the same children were then tested and treated annually from 2017-2018 to quantify re-infection rates each winter season at the village-level (Fig. S2). New children were recruited to the study as needed when children left the school, and thus we could not always track the same children throughout the study. Thus, we could examine baseline infection at all 23 sites in 2016, but infection post-treatment for control villages in 2017 and 2018 for only 16 sites.

To determine if human water contact was influenced by vegetation removal, we made one-hour observations of the number people visiting the water, and recorded individual-level variables including: age group, the level of water immersion, as parasite exposure risk rises with body surface area immersed, and the duration of the water contact. We twice assessed water contact at water access points at each of two control and manipulation sites for two time points both before and after vegetation removal, holding constant the time of day of visits to each site so as to control for diurnal variation (Tables S34-39).

Human sampling, drug administration, and informed consent from the parents or guardians of child participants were all overseen by the Biomedical Research Center EPLS (Espoir Pour La Santé), approved by the National Committee of Ethics for Health Research from the Republic of Senegal (Comité National d’Ethique de la Recherche en Santé; CNERS, Dakar, Senegal; Protocol #SEN14/33), and conducted in accordance with the Declaration of Helsinki III and with the International Ethical Guidelines for Biomedical Research Involving Human Subjects as set forth by the World Health Organization guidelines for good clinical practice ^35^. Additionally, this study was also approved by the University of South Florida Internal Review Board (Protocol #MS7_Pro00017473) and the University of Notre Dame (18-10-4951), and is registered on ClinicalTrials.gov (Record R01 TW010286). Any infected children found during data collection were offered the anti-schistosome drug praziquantel (40 mg/kg) and all human data were anonymized prior to analysis to protect the identity of participants. A full description of participant demographics and numbers of children tested in each site is presented in Table S7-S8, S29.

### Crop compost experiments

Following the first round of vegetation removal, we dried vegetation along the shoreline of two sites for several days and then placed it into an earthen pit (approximately 3.5 x 3.5 x 0.5m) at each village, along with an aliquot of animal manure to seed the pile with bacteria that could initiate the decomposition. The pits were covered with a layer of soil in June 2017, but periodically uncovered to monitor its moisture and texture until June 2018, when we randomly sampled 10 locations within each compost pit and then mixed each pit to create a final 1kg composite sample for lab analyses of its nutrient and moisture content (Table S42).

In June 2018, we performed a two factor split-plot experiment wherein we divided a field into six equal rows or blocks, half of which were assigned the whole-plot treatment of fertilizer addition (U = 8 g urea per plant, NU = no urea), with each row divided equally into three randomly assigned split-plot treatments (C = compost added to soil surface, TC = compost hand tilled into the soil, and NC = no compost) (Fig. S7). We applied compost at approximately 7 kg/m^2^ in C and TC plots and planted a local stock of 1,100 pepper seedlings spaced 0.5m apart in each plot. We quantified the number of peppers and their weight (kg) produced per plot when harvested from the field (every 10 days) until the end of the harvest period. This same experiment was then repeated in 2019 for onion production on a novel field using the same planting density and spacing as above. We used an Orient-F1 variant of *Allium cepa* or red onion (LOT: 1257085) using seeds that were pre-treated with the fungicide metalaxyl-M. We planted Orient-F1 seeds on February 6 2019, with community members applying equal amounts of water to all plots beginning on April 18, 2019. We then harvested all onions on June 7, 2019. In each trial, the farmer weighed and counted the peppers and onions from each plot and recorded any evidence of infections on the crops, such as onion rot.

### Livestock feed experiment

Sheep livestock are commonly raised by rural families due to their adaptation to the harsh environment of sub-Saharan Africa, their short production cycle, and low upkeep costs. Farmers can generate income or benefits including milk, meat, offspring (lambs), and manure for fertilizer. Sheep can be grazed on natural pasture but may be housed in shelters, due to their value, and fed a variety of grain or forage (i.e., cornmeal, peanut straw, or pelletized composite feeds).

To determine if aquatic vegetation removed from waterways provides a viable livestock feed supplement, we purchased a total of 30 female sheep in two groups of similar sized ewes and randomly assigned each to one of five treatments. Sheep were stratified by weight so that all treatment groups had a comparable initial mean weight at the start of the two phases of the experiment. An ANOVA revealed that the starting weights among the five treatment groups did not differ (Phase I: *F1,11=*1.624*, p=*0.229, Phase II: *F1,15 =* 0.47*, p =* 0.750; Table S47). The five treatments were 0, 15%, 30%, 45%, and 60% replacement of their standard feed, cornmeal (adults) or peanut straw (juveniles), with *C. demersum* that was dried (2 weeks) in to kill helminth eggs and cysts, ground with a pestle, reconstituted with water (1000g), and mixed to create a relatively homogenous mixture of cornmeal or peanut straw and *C. demersum*. The Phase I trial involved adult ewes, whereas the Phase II trial started with six-month old juvenile ewes. All sheep were maintained individually in enclosures so we could control what they ate and were fed daily at the caloric equivalent of 1000 g of cornmeal. Weight was assessed weekly. A veterinarian technician was on staff to monitor animal welfare and weight gain/loss during the experiment.

### Remote sensing analyses

We received a grant from the DigitalGlobe Foundation that provided sub-meter, visible and near-infrared imagery of northern Senegal (taken by WorldView-2 and -3 commercial satellites) that we coupled with fine scale maps of open water, *C. demersum*, other submerged vegetation, and emergent vegetation at our water access points developed from our drone imagery. We then adjusted the red, green, and blue balance of the satellite imagery and assessed qualitatively whether this enhanced our abilities to discriminate the predominant snail habitat, *C. demersum*, from open water, other submerged vegetation, and emergent vegetation (Fig. 4). Additional details can be found in^30^.

### Data analysis

All non-economic statistical analyses were conducted in *R-3.4.2 and R-4.1.1* (https://www.r-project.org/). The non-economic data and R code for this study are provided as csv and RMarkdown files, respectively, that accompany this paper. The economic analyses were conducted in Stata 16.

#### Agricultural land use study

Even though 16 of the 23 sites used in this study were also part of the vegetation removal study, only baseline data across all sites and non-manipulated control sites were used in our analyses to ensure that these manipulations were not confounded with our agricultural-based analyses (Fig. S2). To determine if village-level agricultural land use predicted the number of infected children, while controlling for other site attributes, we began with a full regression model of all covariates, including: agricultural land use, freshwater habitat cover, water access area, and village population. We also assessed whether crop and freshwater habitat coverage at 0.5km, 1km, or 2km radii was the best predictor of human infection. We performed model selection by Akaike Information Criteria (AIC) using the *AICcmodavg* package for all possible subsets of the site-level predictors (Table S10-12). We performed the above model selection for two separate binomial models in the *MASS* package^36^ to predict the baseline number of children infected with schistosomiasis or fecal schistosomiasis (conducted separately; Table S10-12) at all 23 sites. Between 2016 and 2018, 180 children left the study (88% retention rate), with drop outs occurring as children left the school and/or emigrated to another site.

We examined whether agricultural cover was positively associated with fertilizer use reported in the household survey and whether the use of fertilizer was positively associated with aquatic vegetation cover, using linear regression models. Outliers were tested for using residuals and q-q plots when warranted. We also used linear regression to test whether baseline human prevalence was positively correlated with post-treatment prevalence because our path model combined these data (Table S9, Fig. S9), and thus assumed a strong and consistent local infection risk even after drug treatment. The association between schistosomiasis prevalence in children and total crop cover within 0.5 km was tested using a binomial regression, using the number of positive and negative children tested per a site. McFadden’s psedo-R^2^ was also calculated (Table S13-14).

To quantify the relationships among agricultural cover, aquatic habitat, snail hosts, and human infection, we performed path analyses using the *piecewiseSEM* package^37^. We created an initial global path model incorporating all data from all baseline infection and infection post-treatment rounds that followed snail data collection (Fig. S2, Table S15). We included all hypothesized causal pathways between agriculture and human infection that were supported by available literature and our study design (Tables S1^15^). We could not perform a child-level path model, because *piecewisesem*() required a single input dataset where each response variable can vary for each observation (row) and thus we aggregated each response variable in the dataset per year at the village-level (but see the child-level analyses below for consideration of individual-level effects). All pathways had a random effect of village to account for repeated sampling. All variables that functioned as independent variables in the path analyses were summarized at the village-level, standardized using the *scale*() function, and modeled with a Gaussian error distribution. Before scaling, all count variables were natural-log transformed, as was vegetation mass because it was positively skewed. For infection prevalence, which only functioned as a dependent variable, we used a binomial error distribution. We performed model selection by dropping non-significant predictors from our initial global and compared nested models using AIC until all predictors were significant (Tables S16-18). Given that our path analysis used mixed models to account for repeated sampling of sites, we report both marginal and conditional *R*^2^ values (Table S19), which are based on only fixed effects or both fixed and random effects, respectively. Although the full path model displayed in Fig. 1b has never been tested, many of the individual paths in the model are well-established in the literature (e.g.^15^, Table S1), providing clear *a priori* directional hypotheses (i.e., support for one-tailed tests) for the paths in our site-level path model.

Given that our path model could only be performed at the hierarchical level of the site, we also performed a separate mixed effects binomial model at the child level to determine whether predictors identified as important by a previous study^15^, such as child gender and age, influenced our findings. The results of this child-level regression generally agreed with our site-level path model, but with reduced statistical power; thus these results were relegated to Supplement (Table S55-58).

#### Aquatic vegetation removal randomized control trial

We estimated average and 95% CIs for the vegetation mass (g) sampled at the same random points before/after the first removal using the *plyr* package^38^ (Table S22-23, Fig. S6). We used the *glmmTMB* package^39^ to evaluate how the log-transformed quantity of vegetation removed (kg) at the water access point-level changed over the study using fixed effects for removal round (1-10), water access area (m^2^), an interaction between round and area, and the number of labor hours used during removal. We also included a random term for removal round nested within water access point to account for re-sampling.

To determine whether total snail counts in 2017 and 2018 were impacted by vegetation removal, we used the *glmmTMB* package^39^ to compare a Poisson, negative binomial, and a zero-inflated negative binomial model (ZINB; Table S24), each with the same fixed effects for vegetation removal treatment (hereafter ‘treatment’), manipulation time (a factor for before or after treatment), an interaction term between treatment and time, and a nested random term for sampling visit within point location within water point to account for re-sampling the same points before/after removal (Table S25). Model selection using AICc values favored a ZINB model (Table S24), likely because sampling points without snail habitat generally had no snails during the study. To calculate the effects in our ZINB model, we used the *effects* package^40^ and plotted the interaction between time and treatment using *ggplots2* package^41^ (Fig. 2c). Finally, we performed a Tukey’s pairwise post-hoc contrast on the ZINB model above using the *lsmeans* package^42^. We also used a Pearson’s Correlation to test for the association between the log-transformed quantity of vegetation (kg) removed and the change in snail counts for time points before and after the removal (Fig. 2b).

To determine if child-level probability of schistosomiasis infection was negatively associated with the vegetation removal intervention, we performed a generalized linear mixed-effects models (GLMM) using the *lme4* package. Only children with both urine and feces samples tested were included in the models. We predicted infection probability for each schistosome species separately at the child-level using fixed predictors for treatment (removal or control), time (before or after), and an interaction term between treatment and time. We also included fixed predictors for child gender, school class, and a factor for whether sites were on the lake or river, and included a random intercept term for time nested within site to account for temporal sampling (Table S30-31, S39). We also used the same predictors and random terms above to perform negative binomial GLMMs to separately predict child egg burden (natural log transformed egg counts) for each schistosome species (Table S30, S32, S40). For all models, we selected human re-infection data per site for the year preceding the start of vegetation removal (2017 for 3 site pairs and 2018 for 5 site pairs) and for one year after vegetation removal to follow methods provided for a before-after-control-impact (BACI) analysis with multiple sites^43^ The ‘dredge’ function in the *MuMIn* package^44^ was used to fit all subsets of the primary model and to compute model-averaged regression coefficients, standard errors (SEs), and cumulative AIC weights as a measure of variable importance (Table S31-32, S39-40). We averaged all models with a delta AIC less than two using the model.avg function in the *MuMIn* package. Estimated marginal means and standard errors were calculated for both control and removal sites in the before and after time periods and the means before and after were compared (Table S29), using the *emmeans* package^45^.

To determine how human behavior at water access points was influenced by vegetation removal, we used the *ordinal* package^46^ to perform a mixed model using the ordinal response level of water contact immersion as the dependent variable (Table S33), and with the following fixed predictors: intervention time, intervention, the interaction between intervention time and intervention, age group, gender of the individual, and time of day of each observation (Table S34-38). We included a random term for time nested within site to account for correlation within observation rounds. We then used the same fixed and random terms above but in a Gaussian mixed model to predict the ln-transformed duration of water contact (minutes) of each observed individual (Table S37-38).

#### Crop compost experiments

To determine whether compost improved crop yield, we performed negative binomial regression to separately predict the total number of pepper and onions at the end of our split-plot experiments with fixed effects for compost treatment (C, TC, or NC), fertilizer treatment (U, NU), site, and an interaction term between the compost treatment and fertilizer (Table S44-45). A mixed effect model was used to account for harvest round for peppers and site was included as a fixed effect. We then used a Gaussian error distribution using the same fixed and random terms above to separately predict natural log-transformed pepper weight, onion weight, and onion diseases (Onion rot) (Table S44-45).

#### Livestock feed experiment

To analyze sheep growth rates among treatments, we performed two linear regression analyses for each sheep age group (juveniles and adults), each with percent of *C. demersum* in the feed as the continuous independent variables and one with percent change in sheep weight and another with final weight as the dependent variables (Fig. 3g, h; Table S48). A separate mixed-effect model with Sheep ID as the random intercept, sheep mass as the dependent variable, and interactions among age of the ewes (juvenile or adult), date, and percent of *C. demersum* in the feed was conducted to test for interactions among time and treatments.

#### Private economic benefits of using aquatic vegetation as compost

To determine economic value of compost before accounting for the costs of vegetation removal, transport, and compost application, we first estimated the marginal physical product (MPP) by separately regressing the (natural) log-transformed fruit yield (kg/ha) of pepper and onion using binary indicator variables for each compost treatment (Table S59). We also included a factor for site for pepper because pepper crop trials were performed in two sites. We also generated kernel density plots and stochastic dominance testes of yield by compost treatment for both peppers and onions, to determine variation across the whole yield distribution. We then calculated the expected marginal revenue product (MRP) of compost by multiplying the MPP estimates from above by the expected price of each crop and then converting the MPP to per metric ton of compost. To determine the marginal cost of the compost, we multiplied the sum of the labor time spent removing vegetation and labor time creating and applying the compost per compost treatment by the estimated average cost of labor for horticultural work in our study region provided by KU Leuven and the Institut Sénégalais de Recherches Agricoles Bureau d’Analyses Macro Economiques (Table 47). We also added the cost of cart rental to transport the compost to the field to produce the marginal cost per metric ton of surface and tilled compost. For additional details on the economic analyses, see SI Appendix Text S3.

#### Private economic benefits of using aquatic vegetation as livestock feed

To estimate economic costs of *C. demersum* as a livestock supplement, we used the cost per calorie of each food type. We used 159 calories/100g, 210 calories/100g and 350 calories/100g, as the nutritional content of *C. demersum*, peanut straw, and cornmeal, respectively^47^ (Table S50). The cost of one kg bag of cornmeal is 290 FCFA ($0.50 USD) and the cost of one kg of peanut straw is 281 – 375 FCFA ($0.49 – $0.65 USD) while it costs less than 0.01 FCFA to produce one kg of *C. demersum* (Table S50). Thus, a single calorie of cornmeal costs 8.28 FCFA and a single calorie of peanut straw costs 13.39 – 17.85 FCFA while a single calorie of *C. demersum* costs 0.02 FCFA (Table S50). Given that the sheep are fed *C. demersum* in calorie equivalence to the cornmeal feed or peanut straw, the expectation is that there should not be significant differences in weight gain across the different treatment groups, which is consistent with what we found in our experiment (*p*=0.86, Fig. 3g, h). For additional details on the economic analyses, see SI Appendix Text S3.

#### Public economic benefits of aquatic vegetation removal

To calculate the public health benefit of the vegetation removal, we began by calculating the Disability-Adjusted Live Years (DALYs) averted due to the reduction in *S. mansoni* infection prevalence among school-aged children. Disease burden in DALYs is calculated by summing the Years of Life Lost (YLL) and Years Lost to Disability (YLD) from a disease or condition. Given that the WHO data indicate that schistosomiasis causes very few deaths in Senegal since 2016, and no deaths in the 5 – 29 years old age range, we assume that the change in YLL is zero. So, we focus on the reduction in DALYs through a reduction in YLD. The induced change in YLD per person can be calculated by multiplying the estimated reduction in *S. mansoni* infection prevalence (ΔRI) by the estimated disability weight (DW) for schistosomiasis. Since the intervention was done at the site level, we then multiply by the average village population (POP) to calculate the DALYs gained for the community. We then use the conservative valuation of each DALY at the gross national income (GNI) per capita for Senegal in US dollars (GNIpc) to calculate the monetary value of the averted DALYs. Finally, we divide the value of the averted DALYs by the total amount of compost (C) that would have been produced if all vegetation removed was composted, using high- and low-end estimates of the vegetation loss in the composting process. This yields the money metric public health (DALY) benefit per metric ton of compost. This estimate follows the equation:

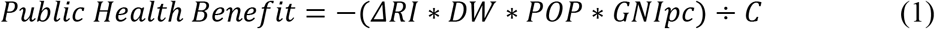

Table S56 describes the parameters and their data sources. We estimate the public health benefit per metric ton of compost to be $17.36 on the low end, with a high-end estimate of $22.21 while the public health benefit of one kg vegetation used in animal feed is 0.01.

The expected public health benefit of $19.79 adds 15-22% to the gross private benefits from composting the harvested aquatic vegetation. Applying this public health gain to the private gains associated with onion and pepper yields results in an inclusive estimated benefit-to-cost ratio of removing vegetation and converting it to compost that now ranges from 3.1 to 8.8. For animal feed, the expected public health benefit adds 1 – 25% to the gross private benefits. Adding this public health gain to the private gain yields benefit-to-cost ratios of -0.39 to 0.46. For additional details on the economic analyses, see SI Appendix Text S3.

## Supplementary Information

### SI Appendix Text S1. More detailed results from the agricultural land use study

#### Counts of infected individuals

A total of 1,479 children were screened for infection at 16 sites in 2016, and 211 additional children at 7 sites were screened in 2017 (average = 92 and 30 children per site, respectively). The number of participants is shown in Table S7, with infection rates by age, gender, and schistosome species in Table S8. Both sets of sites had a similar distribution in agricultural cover within 0.5km and human infection rates, which were positively associated (Figure 1C). *S. mansoni* baseline prevalence was 23% (0 to 93%). *Schistosoma haematobium* was found in school-aged children at all 23 sites with a baseline prevalence of 72% (range 10 to 99%). Despite drug treatment of the same children at 16 sites in 2016, prevalence in 2017 and 2018 remained as high as 99% (Figure S9).

Regression analyses revealed that each km^2^ of agricultural cover around a site was associated with an increase in 1.12 schistosomiasis cases (95% CI: 1.05-1.20), when controlling for the amount of freshwater habitat cover and the area of water access points (see Tables S10-S12 for model selection results).

#### Correlations among land uses and infection observations

We found that baseline infection rates at the 16 sites in 2016 were positively associated with reinfection post-treatment in both 2017 (*R^2^*=0.806, *p*<0.001; Figure S9a) and 2018 (*R^2^* = 0.587, *p* <0.001; Figure S9b). Social surveys found that fertilizers were applied to 61% of the 1,515 ha of total cropped fields at the 16 sites (Table S3-4). Although herbicide use has been associated with increases in snails in mesocosms^7^, herbicides were used too sparsely at these sites (mean of 22 ha/site) to justify analyses (see Table S5 for herbicide use estimates). Given that we only had fertilizer use data for 16 of the 23 sites, the significant positive relationship between fertilizer use and the area of non-emergent vegetation per site was positive (*p*=0.046, Fig. 1e) but not included in our path analyses. Non-emergent vegetation was largely *C. demersum* by mass, which has previously been shown to be the preferred aquatic habitat of schistosome-harboring snails in this region^14,15^. Only 3/23 sites had zero *Bulinus truncatus/globosus* snails and were sites with <1.5kg of sampled, non-emergent aquatic vegetation.

Non-emergent vegetation was largely *C. demersum* by mass, which has previously been shown to be the preferred aquatic habitat of schistosome-harboring snails in this region^14,15^. Only 3/23 sites had zero *Bulinus truncatus/globosus* snails and were sites with <1.5kg of sampled, non-emergent aquatic vegetation.

#### Path analyses of site-level infection

After model selection (Table S16-18), our path analysis suggested that agricultural cover was the strongest positive predictor of human infection prevalence (*Std. coeff.* = 0.993; Table S18). When controlling for snail habitat and snail density, a direct positive pathway between agricultural cover and human infection prevalence identified by d-separation tests (Table S17) suggested a pathway 60% more influential to human schistosomiasis prevalence than snail habitat (*Std. coeff.* = 0.422) or snail density (*Std. coeff.* = 0.362). Further, agricultural cover within 0.5 km from the center of the 23 sites was associated positively with human schistosomiasis infections via a significant indirect pathway whereby total area of crops was positively associated with area of aquatic vegetation at sites, aquatic vegetation was positively associated with snail abundance, and snail abundance was positively associated with *S. haematobium* prevalence in schoolchildren (Table S18, Fig. 1b). More specifically, the total area of vegetation at individual water points increased sampling effort in vegetated points, because the distribution of points within different habitats is proportional to habitat cover. Sampling more points in snail habitat increased estimates of snail habitat mass, which itself increased the total snail abundance found at sites (Fig. 1b).

#### Regression of individual-level infection

We also performed a binomial mixed effects regression model on infection prevalence at the level of the child rather than site, because it offered the maximal statistical power for effects of gender and age that had to be aggregated in the site-level path model. Overall, the child-level model identified that child gender influences upon human infection were significant but did not change the relative strength of three significant positive predictors of human infection revealed by our path model (total crop cover within 0.5 km > aquatic vegetation > snails), indicating that the results of the path model were robust to the inclusion of these individual-level traits (Tables S52-S54).

### SI Appendix Text S2. Discussion of urinary schistosomiasis in clinical trials

We predicted that the vegetation removal intervention would be less effective for urinary (*S. haematobium/S. haematobium-S. bovis* hybrids; we did not discriminate hybrids from non-hybrids) than fecal schistosomiasis (*Schistosoma mansoni*) for several reasons. First, the parasites that cause urinary schistosomiasis have more reservoir^8,9^ and intermediate hosts (Table S6) than *Schistosoma mansoni*, which should reduce the effectiveness of the intervention and increase the rate of snail recovery after the intervention, respectively. Second, *Schistosoma* eggs are likely to enter waterbodies more frequently through urine than fecal contamination. This should also increase the rate of urinary schistosomiasis recovery after the intervention relative to fecal schistosomiasis. Consistent with this, a recent study showed that the spatial scale of influence was much more localized for *S. mansoni* than *S. haematobium*^48^, urinary schistosomiasis regularly returns to 70-90% prevalence in schoolchildren in this region a year after praziquantel treatments (Fig. S9), and several studies have shown that schistosomiasis interventions are less effective in extremely high than moderate to low transmission settings^49^, particularly in West Africa^50^. As hypothesized, the vegetation removal intervention resulted in a weaker reduction of *S. haematobium* than *S. mansoni*, but importantly did not cause harm (Table S29, S30). Given that this intervention successfully reduced *Bulinus* snails, future studies should explore whether there are conditions where this intervention does reduce *S. haematobium*.

### SI Appendix Text S3. Additional Details on the Economic Analyses

#### Marginal Revenue Product Calculations for Compost

To calculate the expected marginal revenue product (MRP) of compost, we first estimated the expected marginal physical product (MPP) for each treatment arm of the crop trials using a multiple regression (Table S59). We regressed the natural log of the fruit yield in kilograms per hectare on one binary indicator variable for each treatment arm separately for both peppers and onions.^1^ The control condition – represented by the regression constant – is no compost and no urea, and we include a control for site in the pepper regressions.^2^ We then performed t-test of the null hypotheses that treatments with compost (either surface or tilled), urea, or urea plus compost (either surface or tilled) generate no yield gains, along with Wald tests of the null of equal yield impacts between combinations of treatment arms (Table S59).

Surface compost and tilled compost both showed statistically significant increases in expected yield relative to the control, with the estimated yield gains from tilled compost slightly, but not significantly higher (Table S59). Surface compost is expected to increase yield by 207.7% (se = 32.4%) for onions and 126.4% (se = 58.7%) for peppers. Tilled compost is expected to increase yield by 229.3% (se = 32.4%) for onions and 126.5% (se = 58.7%) for peppers. The treatments involving urea, either on its own or in combination with compost, generated no statistically significant expected yield gains in either onions or peppers. Compost clearly generates an expected physical productivity gain in both crops, while urea fertilizer does not.

We also generated kernel density plots of yield by treatment arm for both peppers and onions (Fig. S10), so as to reflect variation across the whole yield distribution, not just regression-based testing for differences in mean yield effects for each treatment. We then conducted first and second order stochastic dominance tests. Stochastic dominance is a less restrictive method than regression analysis for comparing among stochastic outcomes, as inference based on regression analysis concerns differences in conditional means between treatment and control, while stochastic dominance compares the entire distribution of outcomes. Under stochastic dominance, one outcome is ranked as superior to another option for a broad class of decision-makers with minimal assumptions. The reasonably innocuous assumptions include decision-makers preferring higher to lower returns (first order dominance), being returns risk averse (second order dominance), and being downside risk averse (third order dominance)^51^. With experimental results on the distribution of returns under control and each treatment arm, stochastic dominance tests thereby allow for quite general inference about which option would be preferable without imposing excessive structural assumptions about people’s preferences.

Stochastic dominance tests revealed that surface compost, surface compost and urea, and tilled compost and urea first order stochastically dominate both the control and the urea-only treatment for onions.^3^ Tilled compost and urea also first order stochastically dominate the compost and compost and urea treatments. First order dominance is a strong criterion; any farmer who prefers higher to lower yields will therefore favor surface compost, surface compost and urea, and tilled compost and urea over the control or urea-only in physical productivity terms. For peppers, surface compost, surface compost and urea, and tilled compost and urea second order stochastically dominate the control condition.

Both surface compost and urea and tilled compost and urea also second order stochastically dominate the urea-only treatment. Second order stochastic dominance suggests that famers who favor higher yields to lower yields and who are risk-averse (i.e., prefer lower to higher variance in yields) should favor tilled compost and urea over surface compost and surface compost and urea. Furthermore, tilled compost and urea third order dominates tilled compost applied to onions, and for peppers, tilled compost third order dominates the control condition while tilled compost and urea third order dominates surface compost, surface compost and urea and tilled compost. If farmers exhibit decreasing absolute risk aversion, meaning that wealthier individuals are less averse to risking a fixed amount of money than are poorer individuals – and most economic evidence supports the decreasing absolute risk aversion hypothesis – then tilled compost and urea applied to onions will be preferred to just applying tilled compost. Based on both the regression and stochastic dominance analysis, we focus the economic valuation on the surface compost and tilled compost treatments.

To calculate the expected MRP, we had to multiply the expected MPP estimates generated from the regression analysis described above by the expected price of the product. To estimate expected prices, we used monthly price data from January 2017 to December 2019 for peppers and onions from Dakar, Senegal as reported by the Agence National de la Statisitque et de la Demographie (Fig. S11).^4^ Dakar is the destination market for these crops grown in the study area, so we need to adjust Dakar prices for the spatial marketing margin.^5^ We estimate the spatial marketing margin using the marketing margin between Saint Louis and Dakar in monthly rice prices between October 2017 and December 2019 (Figure S12). We combine the Dakar onion and pepper prices with the estimated mean marketing margin – and as a robustness check, one standard deviation above and below the mean marketing margin as well.

We then converted the estimated expected marginal revenue product of the compost treatment from FCFA to USD using the mean exchange rate, January 2017 to December 2019, by converting FCFA to Euros by the fixed exchange rate (1 FCFA = 0.0015 Euro) and then converting Euros to USD using Federal Reserve Economic Data.^6^ Finally, we converted the expected MRP estimate to per metric tons of compost.

We estimated an expected MRP per ton of surface compost is $102.05 for onions ($95.07 and $109.04 are the lower and upper estimates, respectively, from using the estimated marketing margin plus or minus one standard deviation to adjust the price of pepper and onions from Dakar prices to Saint Louis prices) and $132.88 for peppers ($123.79 and $141.98). The expected MRP per ton of tilled compost is $130.18 for onions ($121.28 and $139.09, Table SX) and $133.07 for peppers ($123.96 and $142.17).

#### Marginal Cost Calculations for Compost

To find the marginal cost of the compost treatments, we start by summing the labor time spent^7^ removing the vegetation (about 12.5-person days),^8^ digging a compost pit (1.5 person days)^9^, moving harvest vegetation to the compost pit (1 person day), moving compost to the fields (9 person days), spreading the compost (7.5 person days), and burying compost for the tilled compost treatment only (7.5 person days). Then, using estimates of the rural wage rate from a 2018 survey of 27 horticulture workers from 7 sites in the Senegal Delta region^10^, we estimated the average cost of labor, displayed in Figure S13, using both the mean and median daily wage rate (2092 FCFA/day and 2000 FCFA/day, respectively), multiplied these by the labor time needed for each treatment, and added in the cost of renting a cart to move compost from pits to fields (FCFA 2000) to arrive at the marginal cost of the surface compost and tilled compost. The marginal cost is estimated at $35.43 per metric ton of surface compost and $44.05 per metric ton of tilled compost. We also performed the marginal cost calculations using rural wage rates obtained from a larger horticulture survey undertaken by Le Project d’Appui aux Politiques Agricoles (PAPA) that contains information on 1096 horticulture laborers in 57 sites in the Saint Louis region, of which 266 reported a monthly wage^11^. The mean wage rate was 1066 FCFA/day while the median was 1042 FCFA/day and a distribution is displayed in Figure S13. Using these estimates of the rural wage rate, the marginal cost is estimated at $18.60 per metric ton of surface compost and $22.65 per metric ton of tilled compost, so roughly half the estimates using the smaller sample’s estimated wage rates.

We then compared this marginal cost to the MRP calculated earlier. The expected marginal benefit/marginal cost ratio falls between 2.7 and 4.0 (5.1 and 7.6 using the PAPA data), indicating that compost application to these horticultural crops is highly profitable at the margin, even accounting for risk premia. Surface compost application exhibits comparable or superior profitability to tilling compost into soils on these two crops.

For overall profitability measurements, we added in the fixed costs of the equipment required for aquatic vegetation removal: waders (84.99 USD/48822 FCFA and 14.99 USD/8611 FCFA for a high end and low-end price estimate, respectively), shoulder gloves (18.99 USD/10909 FCFA and 7.99 USD/4590 FCFA), 12-teeth rakes (1750 FCFA/rake), round rakes (1500 FCFA/rake) and shovel (7000 FCFA/shovel)^12^.

The resulting low (high) end estimate of the average cost per metric ton of surface compost is $50.76 or $33.93 ($74.58 or $57.76) and $59.39 or $37.98 ($83.21 or $61.80) for tilled compost depending on which wage rates one uses for cost labor (Table S47). Even accounting for fixed costs, compost application on horticultural products is clearly profitable in these communities. Moreover, average total cost will fall as more compost is produced, so these estimates are conservative estimates on the overall cost of surface compost and tilled compost.

#### Marginal Revenue Calculations for Animal Feed

To calculate the benefit of animal feed, i.e., the marginal revenue product of the input, we first to value weight gain in sheep. Based local estimates of 50000 – 55000 FCFA for 28 – 30 kg sheep, we estimate the value of one kg of sheep to be 1667 – 1964 FCFA ($2.90 – $3.42 USD). Then, we calculate the average weight gain of the sheep during the feeding trial for each treatment arm. Finally, we multiply the average weight gain by the estimated value of one kg of sheep to get the total benefit of raising sheep throughout the course of the trial using each feed. We emphasize that the per kg price of sheep varies considerably based on lots of other attributes of the sheep, general market conditions, etc. so these estimates are necessarily coarse. Our resulting estimates of the benefit of raising sheep range from $-7.98 USD to $15.39 USD per 1kg of peanut straw isocalorically equivalent. The lower bound is negative because while there was no statistical difference in weight gain across treatment arms for sheep, some arms had negative average weight gain.

#### Marginal Cost Calculations for Animal Feed

To calculate the cost of animal feed, first we use local prices in Saint Louis to calculate the price of peanut straw, cornmeal, or pellets in the sheep feed. The price of peanut straw is 4500 – 6000 FCFA / 16 kg bag, pellets are 8000 – 10500 FCFA / 40 kg bag,^13^ and cornmeal costs 290 FCFA for one kg. Then, we calculate the cost of using aquatic vegetation as animal feed. The cost of the aquatic vegetation is labor needed to remove it requiring a daily wage rate. Using daily wage rates of 2092 FCFA/day and 1066 FCFA/day, we calculate that one kg of vegetation costs 1.65 – 3.23 FCFA to produce as one worker can remove a large volume of vegetation during one workday. Using the iso-caloric substitution feed quantities per day, we calculated the cost of animal feed per day. Then, we scaled that value by the number of days in the trial and converted prices from FCFA to USD to get the total feed cost for the entire trial. The resulting feed costs range from $20.12 USD to $60.14 USD. The lowest feed costs come from the highest amounts of aquatic vegetation in the feed.

To get from individual costs and benefits to benefit-cost ratios, we divided the total benefits of raising sheep, which is the value from their additional weight, by the total cost of their feed. The benefit-cost ratios range from -0.396 to 0.454. Again, the lower bound is negative because while there was no statistical difference in weight gain across treatment arms for sheep, some arms had negative average weight gain. These results suggest that on average it was not profitable to raise sheep strictly by providing feed, no matter the level of vegetation substitution. This is not surprising given that sheep graze freely when forage is available preventing many households from needing to continuously purchase feed. Conditional on a livestock owner finding that forage or other conditions necessitate providing sheep with supplemental feed so as to prevent excessive weight loss, the *Ceratohyllum demersum*-rich blends were far cheaper relative to feed available on the market presently. While complete reliance on supplemental feed of any sort is on average unprofitable in this system, relative to other feed options, the harvested vegetation provides farmers an equally beneficial but far lower cost option.

## Supplemental Tables

**Table S1.** List of hypotheses between predictor and response variables used in the starting full piecewise-path model, prior to model selection, based on *a priori* pathways from the literature or our sampling design. Model selection was conducted by individually dropping non-significant pathways and incorporating any significant missing pathways in d-separation tests until the final model fit the observed data. All variables are summarized at the site-level and all variables except for infection prevalence (a binomial error distribution) were scaled and modeled with a Gaussian error distribution. Prior to scaling, all count variables were natural-log transformed, as was vegetation mass because it was heterogeneous within water points and positively skewed. Significant paths after model selection are shown in Fig. 1b.

| Response | Predictor | Hypotheses and/or literature |
| --- | --- | --- |
| Total area of non-emergent vegetation across water access sites of each site (m<sup>2</sup>) | Total crop area within 0.5km (km<sup>2</sup>) | Total crop cover was expected to increase aquatic vegetation, especially *Ceratophyllum demersum* that uptakes dissolved nutrients quickly (particularly inorganic N) and grows to form dense sub-surface mats<sup>17-19</sup> |
| Total area of non-emergent vegetation across water access sites of each site (m<sup>2</sup>) | Total fertilizer use (ha) | Fertilizer use near water points was expected to increase aquatic vegetation, especially *Ceratophyllum demersu*<sup>17-19</sup>, but was only available at a subset of 16 sites and thus has less power as a predictor than total crop cover (*n* = 23 sites) |
| Frequency of non-emergent vegetation in sampled quadrats/dipnets (natural-log transformed) | Total area of non-emergent vegetation across water access sites of each site (m<sup>2</sup>) | We expected a positive relationship because we stratified the number of random points that were sampled based on vegetation cover at the water access point (sampling design) |
| Total non-emergent vegetation mass sampled at each site (g) (natural-log transformed) | Frequency of non-emergent vegetation in sampled quadrats/dipnets (natural-log transformed) | Total vegetation mass sampled at a site was related to the frequency of sampling points falling within non-emergent vegetation, which by design, was selected to match the visual estimates of total non-emergent vegetation cover at a site; however, total vegetation mass could also have been affected by the depth of the non-emergent vegetation mats at those points, which was found to be relatively uniform at these sites, especially for *C. demersum* that forms dense mats that can extend to 6 m in depth<sup>52</sup> |
| Total non-emergent vegetation mass sampled at each site (g)<br>(natural-log transformed) | Total fertilizer use (ha) | Fertilizer use (ha) near water points was expected to increase aquatic vegetation, especially *Ceratophyllum demersum* that forms dense mats that can extend to 6 m in depth<sup>52</sup> |
| Total snail count at site<br>(natural-log transformed) | Total non-emergent vegetation mass sampled at each site (g)<br>(natural-log transformed) | Total non-emergent vegetation mass at a site, especially *C. demersum*, was previously shown to correlate with *Bulinus* spp. snail counts using the same human and environmental (snail and habitat) data as presented in this paper for quadrat sampling<sup>15</sup> and for dipnet sampling |
| Human infection prevalence | Total snail count at site<br>(natural-log transformed) | Snail abundance was already demonstrated to be positively associated with human infection post-treatment in the following season, using the same human and environmental (snail and habitat) data as presented in this paper for quadrat sampling, when aquatic vegetation abundance is properly accounted for in statistical modeling<sup>15</sup> |
| Human infection prevalence | Total area of non-emergent vegetation across water access sites of each site (m<sup>2</sup>) | Non-emergent vegetation may increase parasite production in snails by increasing food resources and therefore energy available to snails and parasites for parasite asexual reproduction<sup>14</sup>, or by increasing the probability of snail presence through by providing more habitat to colonize<sup>53</sup> |
| Human infection prevalence | Village population | The number of definitive human hosts was expected to increase disease prevalence given findings by<sup>15</sup>. |
| Human infection prevalence | Total crop area within 0.5km (km<sup>2</sup>) | The positive relationship between total area of crops and floating vegetation was expected to increase not only snail abundance but also potentially increase parasite production per snail given findings by Haggerty et al.<sup>14</sup> |

**Table S2.** List of site-level variables used in separate negative binomial models to predict human baseline infection (*n=23*) and infection post-treatment (*n=16*) one year after drug treatment. All negative binomial models included an offset term for the log-transformed number of children tested to account for the fact that the number of children tested varied by site.

| Human data | $n$ | Predictor | Scale | Source (year) |
| --- | --- | --- | --- | --- |
| Baseline infection | 23 | Total crop cover | 0.5km | FAO (2005) |
|  |  | Total freshwater habitat | 0.5km | FAO (2005) |
|  |  | Village population | Site | Household survey (2016) |
|  |  | Water access area | Site | Aerial imagery (2017) |
| Infection post-treatment | 16 | Total crop cover | Site | Household survey (2016) |
|  |  | Total freshwater habitat | 0.5km | FAO (2005) |
|  |  | Village population | Site | Household survey (2016) |
|  |  | Water access area | Site | Aerial imagery (2017) |

**Table S3.** Site characteristics including village population, sq. km of total crop and freshwater habitat within 0.5 km (remote imagery), hectares of total field area and fertilizer or insecticide use (household survey), average non-emergent vegetation area and water access point area during the study period (drone imagery). Ag = one of the seven additional sites used in the agricultural land use study. Removal = is a vegetation removal site. Control = is a control, non-removal site.

| Site ID | Inter-<br>vention | Village<br>pop-<br>ulation | Sq. km<br>total crop<br>0-5km | Sq. km<br>water<br>0-5km | Field<br>area<br>(ha) | Total<br>fertilizer<br>(ha) | Total<br>insecticide<br>(ha) | Mean non-emergent<br>vegetation<br>area (m<sup>2</sup>) | Mean<br>access site<br>area (km<sup>2</sup>) |
| --- | --- | --- | --- | --- | --- | --- | --- | --- | --- |
| 1 | Ag | 165 | 0.30 | 0.00 | NA | NA | NA | 1921.00 | 12450.00 |
| 2 | Ag | 91 | 0.58 | 0.01 | NA | NA | NA | 4920.00 | 1035.00 |
| 3 | Removal | 1500 | 0.22 | 0.06 | 159.60 | 143.1 | 85.6 | 1255.49 | 3452.76 |
| 4 | Removal | 1300 | 0.79 | 0.03 | 116.30 | 57.0 | 65.5 | 2584.98 | 5613.69 |
| 5 | Control | 107 | 0.41 | 0.00 | 63.00 | 38.2 | 39.4 | 520.99 | 2953.02 |
| 6 | Removal | 459 | 0.48 | 0.00 | 88.00 | 26.6 | 38.5 | 623.63 | 1683.94 |
| 7 | Removal | 400 | 0.47 | 0.14 | 81.55 | 54.6 | 58.1 | 944.26 | 2511.67 |
| 8 | Removal | 1623 | 0.28 | 0.06 | 59.79 | 55.8 | 22.6 | 1553.70 | 2918.33 |
| 9 | Removal | 609 | 0.37 | 0.20 | 192.80 | 154.0 | 74.0 | 583.36 | 1783.95 |
| 10 | Control | 648 | 0.49 | 0.02 | 50.76 | 46.8 | 29.5 | 81.46 | 481.16 |
| 11 | Control | 833 | 0.55 | 0.27 | 148.35 | 87.1 | 100.3 | 2481.98 | 6413.00 |
| 12 | Removal | 1120 | 0.17 | 0.30 | 51.90 | 15.1 | 9.4 | 351.00 | 1373.80 |
| 13 | Removal | 1852 | 0.33 | 0.08 | 163.15 | 113.4 | 100.8 | 882.07 | 1940.32 |
| 14 | Control | 1316 | 0.32 | 0.03 | 24.15 | 17.2 | 9.5 | 168.54 | 663.71 |
| 15 | Control | 761 | 0.70 | 0.08 | 70.25 | 17.8 | 14.3 | 278.57 | 3204.23 |
| 16 | Ag | 1146 | 0.49 | 0.05 | NA | NA | NA | 3775.00 | 46700.00 |
| 17 | Ag | 2200 | 0.32 | 0.00 | NA | NA | NA | 1905.00 | 29220.00 |
| 18 | Ag | 517 | 0.42 | 0.11 | NA | NA | NA | 2135.00 | 9300.00 |
| 19 | Control | 1542 | 0.26 | 0.09 | 29.90 | 15.5 | 16.7 | 692.73 | 1217.24 |
| 20 | Control | 771 | 0.56 | 0.01 | 49.19 | 42.9 | 25.1 | 1050.38 | 2571.77 |
| 21 | Ag | 237 | 0.12 | 0.00 | NA | NA | NA | 2464.00 | 19015.00 |
| 22 | Control | 866 | 0.76 | 0.01 | 166.05 | 37.6 | 75.6 | 3347.20 | 6786.36 |
| 23 | Ag | 521 | 0.32 | 0.00 | NA | NA | NA | 1565.00 | 33410.00 |

**Table S4.** Field surface area (ha) of fertilizer application for individual crops per site found via household surveys in 2016 at 16 sites, the percent of the individual crop area where fertilizer was applied (right column), and the percent of crop fields where fertilizers were used (bottom row). DAP = Diammonium phosphate

| Crop | No<br>fertilizer<br>(ha/site) | Urea<br>(ha/site) | NPK<br>(ha/site) | DAP<br>(ha/site) | Manure<br>(ha/site) | Other<br>(ha/site) | Crop area<br>with fertilizer<br>use (%) |
| --- | --- | --- | --- | --- | --- | --- | --- |
| Water melon | 135.1 | 61.0 | 24.0 | 8.5 | 11.5 | 2.0 | 44.2 |
| Goundnut | 39.8 | 100.8 | 40.5 | 7.0 | 3.0 | 7.1 | 79.9 |
| Rice | 2.3 | 194.5 | 28.6 | 4.7 | 0.0 | 0.5 | 99.0 |
| Local bean | 109.0 | 29.0 | 9.5 | 1.3 | 6.0 | 1.0 | 30.0 |
| Onions | 14.5 | 75.8 | 17.3 | 3.3 | 5.8 | 4.5 | 88.0 |
| Sweet potato | 0.3 | 78.5 | 7.0 | 6.7 | 4.0 | 2.5 | 99.7 |
| Cassava | 3.8 | 52.0 | 11.8 | 2.5 | 0.0 | 3.0 | 94.8 |
| Tomato | 2.0 | 10.7 | 9.4 | 7.0 | 2.0 | 0.5 | 93.7 |
| Other crops | 285.9 | 36.6 | 16.5 | 6.0 | 11.0 | 7.8 | 21.4 |
| Total cropped area with<br>fertilizer use (%) | 39.1 | 42.2 | 10.9 | 3.1 | 2.9 | 1.9 | - |

**Table S5.** Field surface area (ha) of herbicide application for individual crops found via household surveys in 2016 at 16 sites, the percent of individual crop area where herbicide was applied (right column), and the percent of crop fields where herbicides were used (bottom row).

| Crop | No herbicide<br>(ha/site) | Propanil<br>(ha/site) | Londax<br>(ha/site) | 2,4-D<br>(ha/site) | Other<br>(ha/site) | Crop area with<br>herbicide use (%) |
| --- | --- | --- | --- | --- | --- | --- |
| Water melon | 233.6 | 0.0 | 4.0 | 1.0 | 3.5 | 3.5 |
| Rice | 25.0 | 109.9 | 79.5 | 2.3 | 13.9 | 89.2 |
| Groundnut | 169.0 | 1.0 | 4.0 | 0.5 | 23.5 | 14.6 |
| Local bean | 138.6 | 1.0 | 0.3 | 2.0 | 14.0 | 11.0 |
| Onion | 73.2 | 15.8 | 7.9 | 5.5 | 18.8 | 39.6 |
| Sweet potato | 92.9 | 0.0 | 1.0 | 0.0 | 5.0 | 6.1 |
| Tomatoes | 26.6 | 0.0 | 4.0 | 0.0 | 1.0 | 15.8 |
| Other | 421.3 | 3.5 | 2.7 | 0.5 | 8.7 | 3.5 |
| Total cropped area with<br>herbicide use (%) | 77.9 | 8.7 | 6.8 | 0.8 | 5.8 | - |

**Table S6.** Snail abundance and vegetation mass (g) sampled at each site (aquatic sampling in 2017). Ag = one of the seven additional sites used in the agricultural land use study. Removal = is a vegetation removal site. Control = is a control, non-removal site.

| Site ID | Intervention | *Bulinus*<br>spp. snail<br>counts | *Biomphalaria*<br>*glabrata* snail<br>counts | Vegetation<br>mass (g) |
| --- | --- | --- | --- | --- |
| 1 | Ag | 123 | 1 | 12450 |
| 2 | Ag | 0 | 0 | 1035 |
| 3 | Removal | 59 | 9 | 13260 |
| 4 | Removal | 1 | 0 | 7005 |
| 5 | Control | 0 | 0 | 450 |
| 6 | Removal | 5 | 21 | 200 |
| 7 | Removal | 47 | 16 | 25261 |
| 8 | Removal | 91 | 4 | 8420 |
| 9 | Removal | 135 | 1 | 11208 |
| 10 | Control | 0 | 9 | 5 |
| 11 | Control | 10 | 9 | 3810 |
| 12 | Removal | 1091 | 32 | 8351 |
| 13 | Removal | 18 | 7 | 12858 |
| 14 | Control | 2 | 0 | 1470 |
| 15 | Control | 0 | 0 | 1340 |
| 16 | Ag | 341 | 34 | 62500 |
| 17 | Ag | 39 | 0 | 44780 |
| 18 | Ag | 48 | 0 | 9300 |
| 19 | Control | 38 | 4 | 4127 |
| 20 | Control | 22 | 11 | 6200 |
| 21 | Ag | 29 | 2 | 19915 |
| 22 | Control | 12 | 0 | 5766 |
| 23 | Ag | 25 | 24 | 43070 |

**Table S7.**
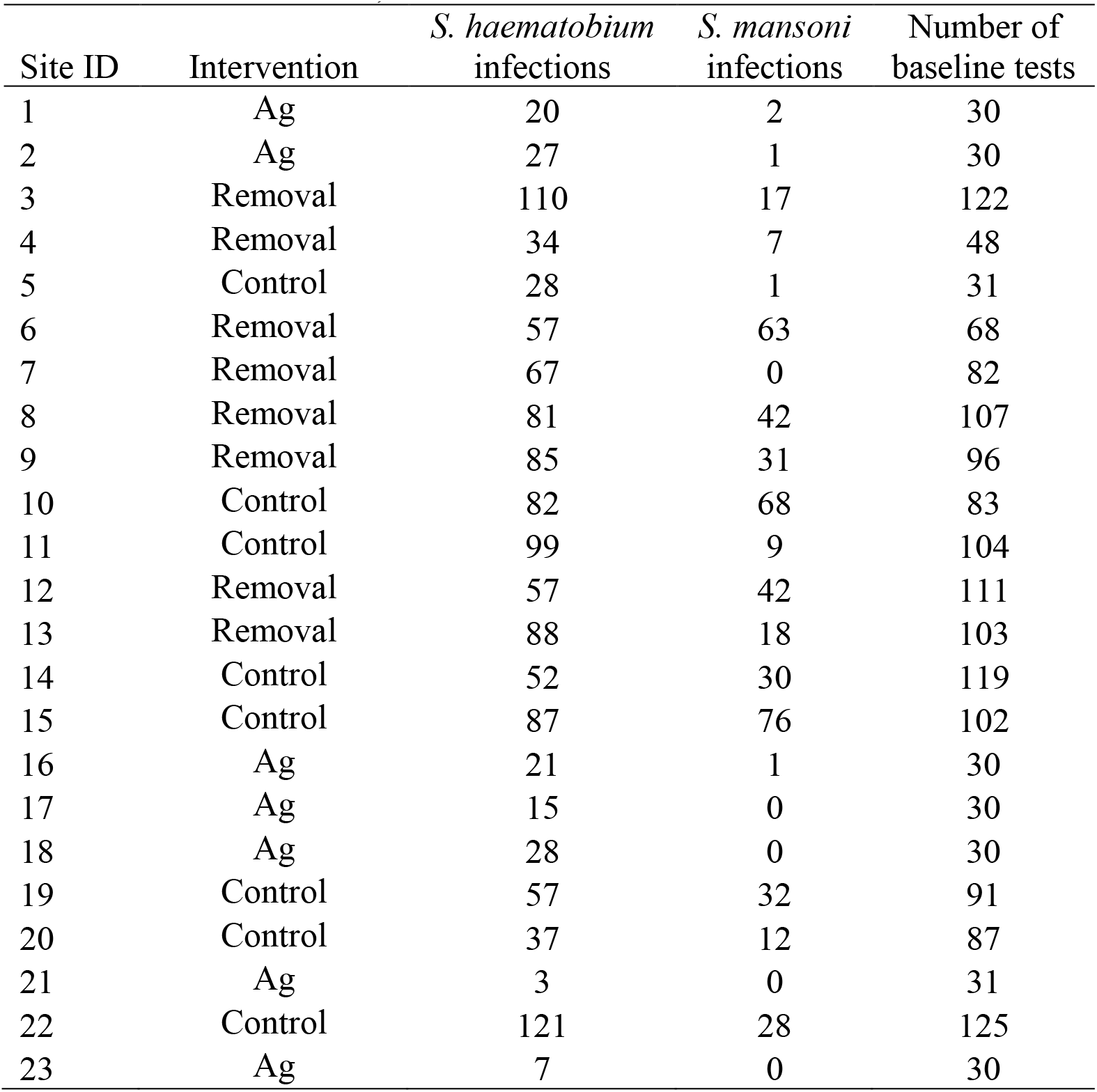
Site characteristics including the number of baseline infections by each schistosome species along with number of individuals tested for baseline infection (human sampling in 2016 or 2017). Ag = one of the seven additional sites used in the agricultural land use study. Removal = is a vegetation removal site. Control = is a control, non-removal site.

| Site ID | Intervention | *S. haematobium*<br>infections | *S. mansoni*<br>infections | Number of<br>baseline tests |
| --- | --- | --- | --- | --- |
| 1 | Ag | 20 | 2 | 30 |
| 2 | Ag | 27 | 1 | 30 |
| 3 | Removal | 110 | 17 | 122 |
| 4 | Removal | 34 | 7 | 48 |
| 5 | Control | 28 | 1 | 31 |
| 6 | Removal | 57 | 63 | 68 |
| 7 | Removal | 67 | 0 | 82 |
| 8 | Removal | 81 | 42 | 107 |
| 9 | Removal | 85 | 31 | 96 |
| 10 | Control | 82 | 68 | 83 |
| 11 | Control | 99 | 9 | 104 |
| 12 | Removal | 57 | 42 | 111 |
| 13 | Removal | 88 | 18 | 103 |
| 14 | Control | 52 | 30 | 119 |
| 15 | Control | 87 | 76 | 102 |
| 16 | Ag | 21 | 1 | 30 |
| 17 | Ag | 15 | 0 | 30 |
| 18 | Ag | 28 | 0 | 30 |
| 19 | Control | 57 | 32 | 91 |
| 20 | Control | 37 | 12 | 87 |
| 21 | Ag | 3 | 0 | 31 |
| 22 | Control | 121 | 28 | 125 |
| 23 | Ag | 7 | 0 | 30 |

**Table S8.** Site characteristics during testing in 2017 and 2018 including prevalence of infections by each schistosome species along with number of individuals tested for infection post-treatment (*n*= 23 sites).

| Year of sampling | Child gender | School grade | Children tested | *S. haematobium* prevalence | Average *S. haematobium* eggs per 10 mL urine (1 SE) | *S. mansoni* prevalence | Average *S. mansoni* eggs per 1 g stool (1 SE) |
| --- | --- | --- | --- | --- | --- | --- | --- |
| 2017 | Female | 1 | 258 | 0.60 | 50.2 (8.69) | 0.15 | 20.8 (6.17) |
| 2017 | Female | 2 | 260 | 0.62 | 23.9 (3.27) | 0.19 | 51.7 (21.7) |
| 2017 | Female | 3 | 156 | 0.60 | 29.9 (5.02) | 0.09 | 10.5 (4.54) |
| 2017 | Female | 4 | 80 | 0.69 | 28.1 (6.08) | 0.18 | 107. (70.6) |
| 2017 | Male | 1 | 250 | 0.60 | 37.2 (4.80) | 0.18 | 28.1 (9.67) |
| 2017 | Male | 2 | 272 | 0.67 | 40.1 (4.51) | 0.20 | 34.0 (11.7) |
| 2017 | Male | 3 | 159 | 0.67 | 26.4 (4.67) | 0.17 | 14.1 (5.67) |
| 2017 | Male | 4 | 91 | 0.75 | 29.6 (6.45) | 0.17 | 25.9 (9.32) |
| 2018 | Female | 1 | 236 | 0.66 | 56.3 (11.7) | 0.21 | 19.9 (7.20) |
| 2018 | Female | 2 | 189 | 0.60 | 32.2 (5.56) | 0.21 | 47.4 (15.9) |
| 2018 | Female | 3 | 139 | 0.62 | 21.7 (5.14) | 0.14 | 27.2 (12.1) |
| 2018 | Female | 4 | 74 | 0.69 | 32.6 (9.55) | 0.27 | 33.5 (15.4) |
| 2018 | Male | 1 | 236 | 0.72 | 65.3 (10.1) | 0.27 | 58.8 (16.3) |
| 2018 | Male | 2 | 191 | 0.75 | 39.5 (5.00) | 0.32 | 49.3 (22.3) |
| 2018 | Male | 3 | 139 | 0.72 | 15.9 (2.67) | 0.23 | 27.2 (9.85) |
| 2018 | Male | 4 | 94 | 0.86 | 27.3 (5.19) | 0.23 | 73.3 (52.6) |

**Table S9.** Comparison of radius around center of site used to calculate the effect of agriculture on baseline schistosomiasis prevalence in children at 23 sites using all subsets selection ranked by Akaike’s Information Criteria corrected for small sample sizes (AICc), including model name (predictor combination), number of estimated parameters (K), AICc, ΔAIC, AICc weight (AICcWt), cumulative AICcWt (Cum.Wt), and log-likelihood (LL).

| Model | K | AICc | Delta<br>AICc | AICc<br>Wt | Cum.<br>Wt | LL |
| --- | --- | --- | --- | --- | --- | --- |
| sqkmtotalcrop0.5km | 4 | 372.91 | 0.00 | 1 | 1 | -181.35 |
| Sqkmtotalcrop2.0km | 4 | 438.14 | 65.22 | 0 | 1 | -213.96 |
| Sqkmtotalcrop1.0km | 4 | 469.95 | 97.04 | 0 | 1 | -229.86 |

**Table S10.**
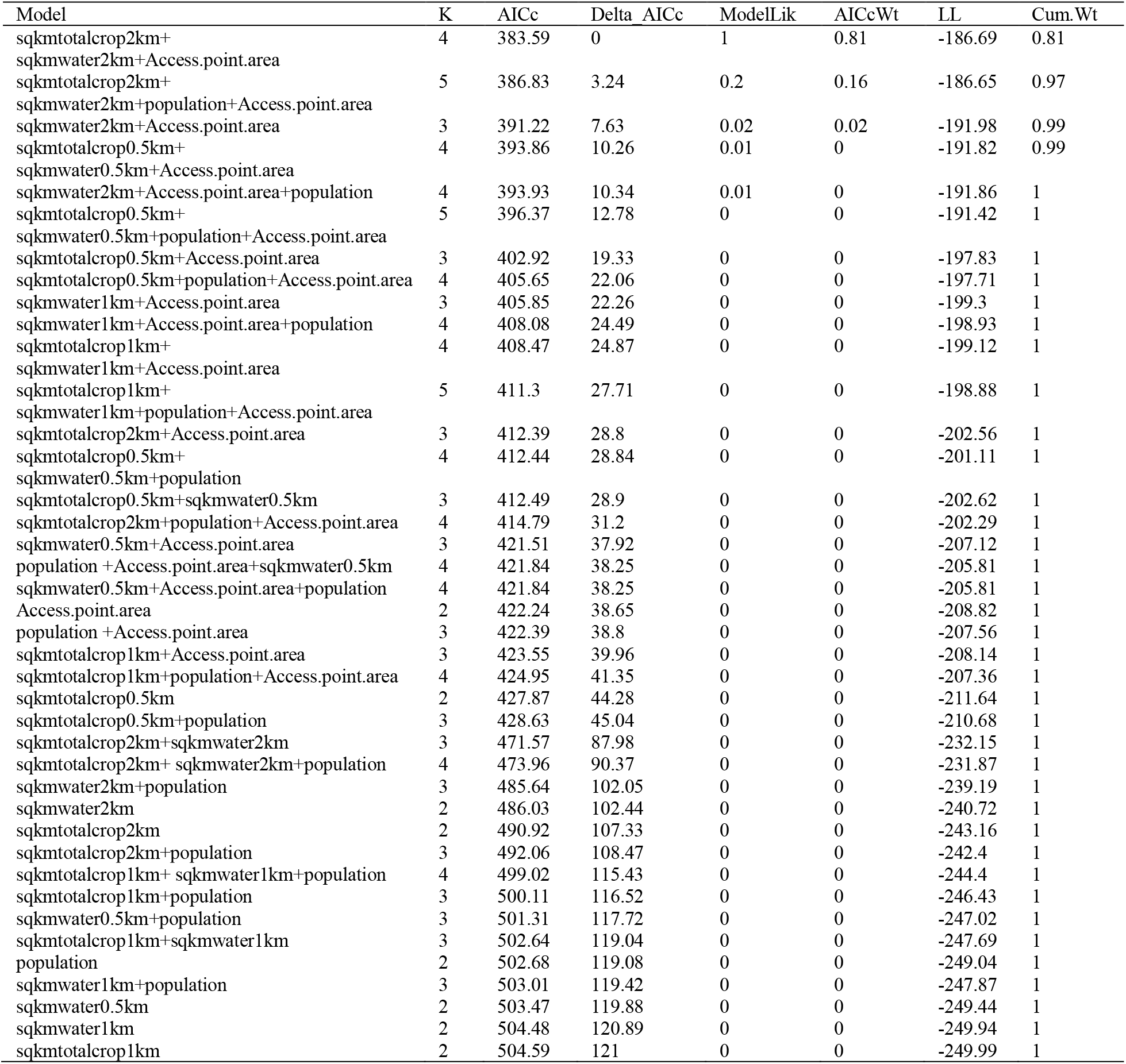
Predictors of baseline human schistosomiasis infection at 23 sites using all subsets selection ranked by Akaike’s Information Criteria corrected for small sample sizes (AICc), including model name (predictor combination), number of estimated parameters (K), AICc, ΔAIC, AICc weight (AICcWt), cumulative AICcWt (Cum.Wt), and log-likelihood (LL).

**Table S11.**
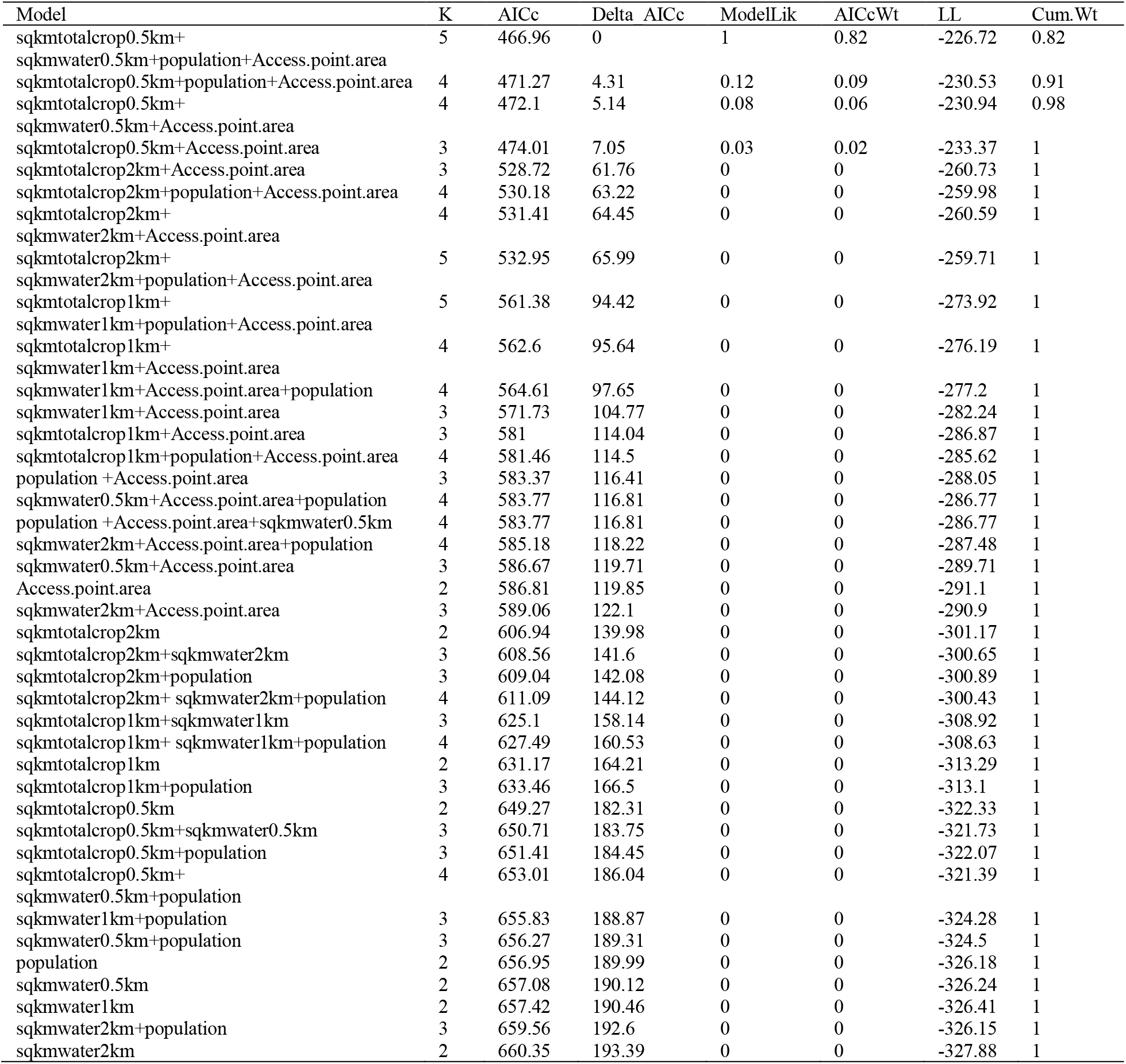
Predictors of baseline human *Schistosoma mansoni* infection at 23 sites using all subsets selection ranked by Akaike’s Information Criteria corrected for small sample sizes (AICc), including model name (predictor combination), number of estimated parameters (K), AICc, ΔAIC, AICc weight (AICcWt), cumulative AICcWt (Cum.Wt), and log-likelihood (LL).

**Table S12.**
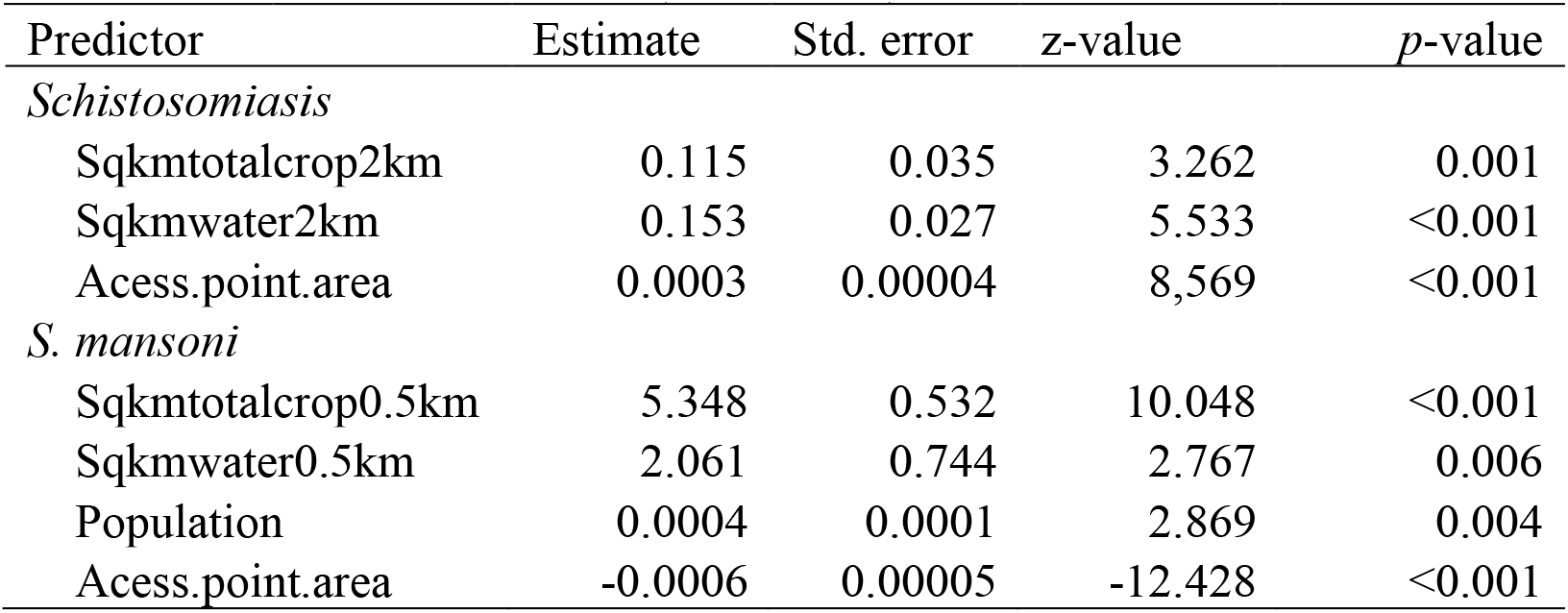
Results of the binomial regression predicting site-level *Schistosoma* prevalence in humans at baseline (*n* = 23 sites).

**Table S13.** Binomial regression results of the association between infected children with schistosomiasis at baseline and total area of crops within 0.5 km of the site center point at 23 sites.

| Predictor | Estimate | Std. error | z-value | p-value |
| --- | --- | --- | --- | --- |
| (Intercept) | 0.067 | 0.152 | 0.444 | 0.657 |
| Sqkmtotalcrop0.5km | 3.126 | 0.377 | 8.292 | <0.001 |

**Table S14.** Linear regression results of the association between agriculture inputs.

| Response | Predictor | Estimate | Std. error | z-value | p-value |
| --- | --- | --- | --- | --- | --- |
| Total fertilizer used | Total area of fields | 0.653 | 0.128 | 5.123 | <0.001 |
| Natural log of the mean<br>of floating vegetation | Natural log of the<br>total fertilizer used | 0.571 | 0.260 | 2.199 | 0.047 |

**Table S15.** Starting initial piecewise-path model results prior to data exclusion and model selection, including all *a priori* pathways from the literature or our sampling design (Table S1). All variables are summarized at the site-level and all variables except infection prevalence (a binomial error distribution) were scaled and modeled with a Gaussian error distribution. Prior to scaling, all count variables were natural-log transformed, as was vegetation mass because it was heterogeneous within water points and positively skewed. One-tailed significance values are reported (Table S1).

| Response | Predictor | Estimate | Std.<br>error | df | Crit.<br>value | *p*-<br>value |
| --- | --- | --- | --- | --- | --- | --- |
| Total area of non-emergent vegetation across water access sites of each site (m<sup>2</sup>) | Total crops (km<sup>2</sup>) within 0.5km of each site centroid | 0.5983 | 0.2257 | 13 | 2.6508 | 0.010 |
| Total area of non-emergent vegetation across water access sites of each site (m<sup>2</sup>) | Total fertilizer use (ha) | 0.2440 | 0.2291 | 13 | 1.0654 | 0.153 |
| Frequency of non-emergent vegetation in sampled quadrats/dipnets | Total area of non-emergent vegetation across water access sites of each site (m<sup>2</sup>) | 0.3750 | 0.1857 | 11 | 2.0192 | 0.034 |
| Total non-emergent vegetation mass sampled at each site (g) | Frequency of non-emergent vegetation in sampled quadrats/dipnets | 0.3921 | 0.1551 | 15 | 2.5289 | 0.012 |
| Total non-emergent vegetation mass sampled at each site (g) | Total fertilizer use (ha) | 0.2702 | 0.1804 | 14 | 1.4981 | 0.078 |
| Total snail counts at each site | Total non-emergent vegetation mass sampled at each site (g) | 0.6282 | 0.1467 | 10 | 4.2832 | <0.001 |
| Human infection prevalence | Total snail counts at each site | 0.3791 | 0.2064 | 27 | 1.8369 | 0.033 |
| Human infection prevalence | Total area of non-emergent vegetation across water access sites of each site (m<sup>2</sup>) | 0.4905 | 0.3123 | 27 | 1.9537 | 0.025 |
| Human infection prevalence | Village population | -0.2389 | 0.2357 | 27 | -1.0135 | 0.155 |
| Human infection prevalence | Total crops (km<sup>2</sup>) within 0.5km of each site centroid | 0.9116 | 0.3123 | 27 | 2.9191 | 0.002 |

**Table S16.** AIC model selection by dropping predictors that were non-significant in the initial global path model prior to any data exclusion. All predictors retained as significant were used in a global path model on the dataset excluding site-by-time points with potential manipulation impacts (e.g. vegetation removal).

| Selection steps | Pathway(s) of predictor(s) dropped | Predictor(s) dropped | AIC | BIC |
| --- | --- | --- | --- | --- |
| Initial global model | None | None | 90.07 | 121.17 |
| Selection 1 | Human infection | Village population | 67.16 | 94.37 |
|  | Total area of non-emergent vegetation (m<sup>2</sup>) | Total fertilizer (ha) |  |  |
|  | Total non-emergent vegetation mass (g) | Total fertilizer (ha) |  |  |

**Table S17.** Directed separation tests^54^ for potential missing pathways in the final path model.

| Response | Predictor | Estimate | Std.<br>error | df | Crit.<br>value | p-value |
| --- | --- | --- | --- | --- | --- | --- |
| Frequency of non-emergent vegetation in sampled quadrats/dipnets | Total crops within 0.5km of each site centroid (km<sup>2</sup>) | -0.2487 | 0.2511 | 14 | -0.9905 | 0.339 |
| Total non-emergent vegetation mass sampled at each site (g) | Total crops within 0.5km of each site centroid (km<sup>2</sup>) | -0.2239 | 0.1765 | 14 | -1.2685 | 0.225 |
| Total snail counts at each site | Total crops within 0.5km of each site centroid (km<sup>2</sup>) | -0.3916 | 0.132 | 21 | -2.9656 | 0.007 |
| Total non-emergent vegetation mass sampled at each site (g) | Total area of non-emergent vegetation (m<sup>2</sup>) | 0.3225 | 0.2006 | 10 | 1.6074 | 0.139 |
| Total snail counts at each site | Total area of non-emergent vegetation (m<sup>2</sup>) | 0.2250 | 0.1689 | 6 | 1.3321 | 0.231 |
| Human infection prevalence | Frequency of non-emergent vegetation in sampled quadrats/dipnets | 0.2749 | 0.2792 | NA | 0.9847 | 0.325 |
| Total snail counts at each site | Frequency of non-emergent vegetation in sampled quadrats/dipnets | 0.1170 | 0.2172 | 5 | 0.5386 | 0.613 |
| Total non-emergent vegetation mass sampled at each site (g) | Human infection prevalence | 0.0545 | 0.7787 | 1 | 0.0700 | 0.956 |

**Table S18.**
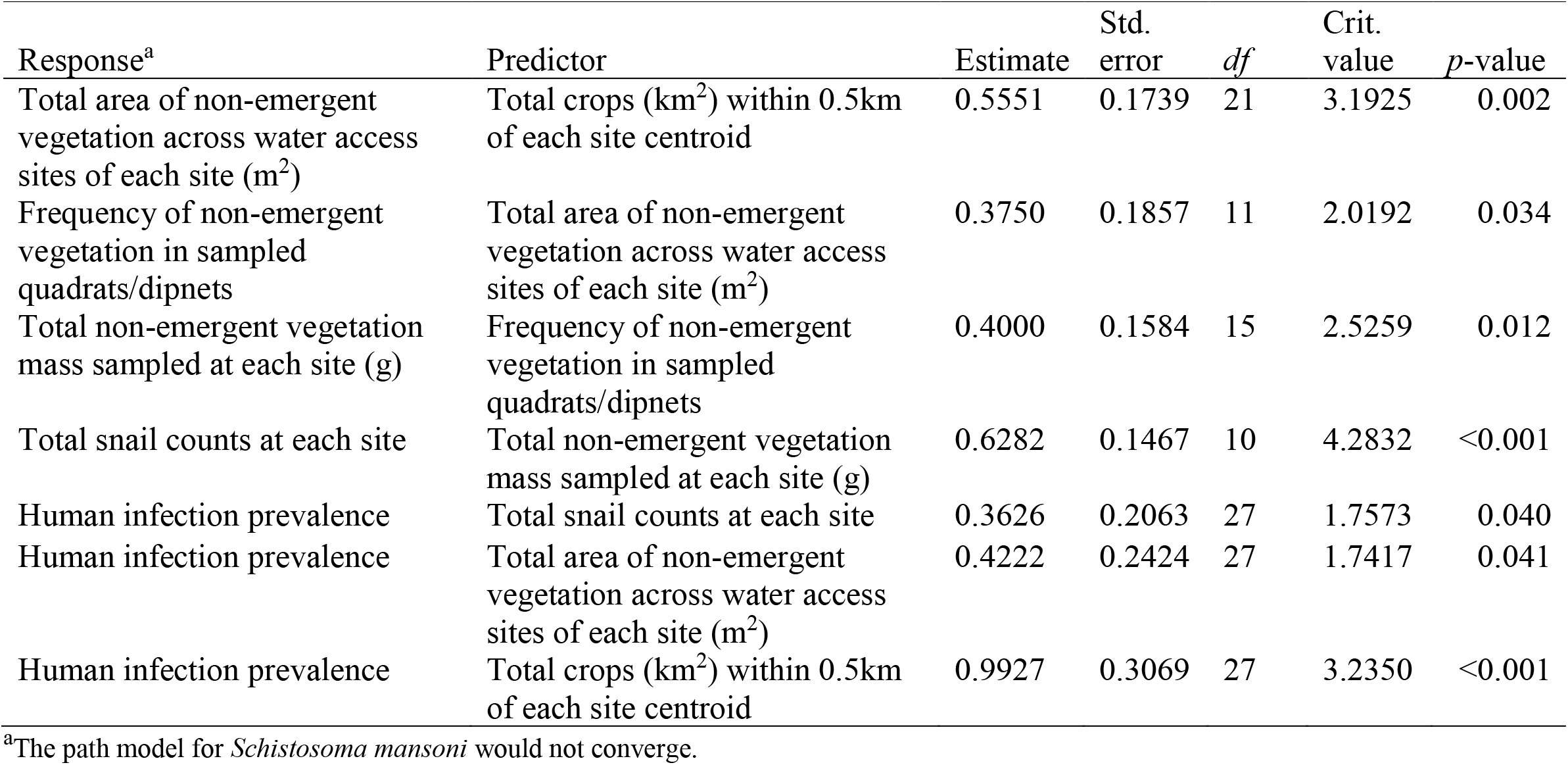
Final piecewise-path model results for infection prevalence using significant predictors from the initial global model selection. All pathways were significant (one-tailed significance testing) without model selection and the final model fit the observed data (*C*16 = 25.2, *p* = 0.067). All variables are summarized at the site-level and all variables except infection prevalence (a binomial error distribution) were scaled and modeled with a Gaussian error distribution. Prior to scaling, all count variables were natural-log transformed, as was vegetation mass because it was heterogeneous within water points and positively skewed. Significant paths after model selection are shown in Figure 1b.

**Table S19.** Marginal and conditional coefficients of determination values (*R*^2^) from the path model relating human infection to agricultural cover near sites. Marginal *R*^2^ shows variance of path model response variables explained by fixed effects only, whereas conditional *R*^2^ includes the variance explained by both fixed predictors and the random effect of site.

| Response | Marginal $R^2$ | Conditional $R^2$ |
| --- | --- | --- |
| Total area of non-emergent vegetation across water access sites of each site (m<sup>2</sup>) | 0.31 | 0.85 |
| Frequency of non-emergent vegetation in sampled quadrats/dipnets | 0.14 | 0.80 |
| Total non-emergent vegetation mass sampled at each site (g) | 0.19 | 0.68 |
| Average snail abundance across water access points of site | 0.40 | 0.72 |
| Human infection prevalence | 0.22 | 0.37 |

**Table S20.** Dates of human parasitology testing for each site by year. Two visits are made to each site represented by P1 and P2.

| Sites | 2016 |  | 2017 |  | 2018 |  | 2019 |  |
| --- | --- | --- | --- | --- | --- | --- | --- | --- |
|  | P1 | P2 | P1 | P2 | P1 | P2 | P1 | P2 |
| ME | 3/14/2016 | 3/25/2016 | 3/2/2017 | 3/9/2017 | 2/15/2018 | 2/22/2018 | 11/28/2018 | 12/4/2019 |
| FS | 3/16/2016 | 3/25/2016 | 2/28/2017 | 3/7/2017 | 2/14/2018 | 2/21/2018 | 11/20/2019 | 11/27/2019 |
| MT | 3/8/2016 | 3/18/2016 | 2/16/2017 | 2/23/2017 | 2/12/2018 | 2/19/2018 | 11/19/2019 | 11/26/2019 |
| GK | 2/23/2016 | 2/29/2016 | 2/15/2017 | 2/22/2017 | 2/13/2018 | 2/20/2018 | 11/18/2019 | 11/25/2019 |
| ST | 2/24/2016 | 3/3/2016 | 3/3/2017 | 3/8/2017 | 2/1/2018 | 2/8/2018 | 11/14/2019 | 11/21/2019 |
| GG | 2/15/2016 | 2/19/2016 | 1/10/2017 | NA | 1/31/2018 | 2/7/2018 | 11/6/2019 | 11/13/2019 |
| MG | 2/25/2016 | 3/2/2016 | 1/11/2017 | NA | 1/30/2018 | 2/6/2018 | 11/5/2019 | 11/12/2019 |
| DF | 2/12/2016 | 2/17/2016 | 2/14/2017 | 2/21/2017 | 1/29/2018 | 2/5/2018 | 11/4/2019 | 11/11/2019 |
| MA | 2/11/2016 | 2/18/2016 | 1/19/2017 | NA | 2/26/2018 | 3/5/2018 | 12/3/2019 | 12/10/2019 |
| DT | 2/10/2016 | 2/21/2016 | 1/24/2017 | NA | 2/27/2018 | 3/6/2018 | 12/2/2019 | 12/9/2019 |
| NE | 4/15/2016 | 4/21/2016 | 4/11/2017 | 4/19/2017 | 3/15/2018 | 3/22/2018 | 1/8/2020 | 1/21/2020 |
| MB | 4/27/2016 | 5/5/2016 | 3/17/2017 | 3/22/2017 | 3/13/2018 | 3/22/2018 | 1/10/2020 | 1/22/2020 |
| MO | 4/22/2016 | 4/28/2016 | 4/13/2017 | 4/21/2017 | 3/12/2018 | 3/21/2018 | 1/20/2020 | 1/24/2020 |
| LR | 4/12/2016 | 4/19/2016 | 4/20/2017 | 4/25/2017 | 3/1/2018 | 3/8/2018 | 1/14/2020 | 1/23/2020 |
| NM | 4/14/2016 | 4/20/2016 | 3/15/2017 | 3/24/2017 | 2/28/2018 | 3/7/2018 | 1/13/2020 | 1/16/2020 |
| MD | 4/25/2016 | 5/2/2016 | 3/14/2017 | 3/23/2017 | 3/14/2018 | 3/23/2018 | 1/9/2020 | 1/15/2020 |

**Table S21.** Dates of human drug treatment for schistosomiasis infection with praziquantel for each site by year.

| Sites | 2016 | 2017 |  | 2018 |  | 2019 |
| --- | --- | --- | --- | --- | --- | --- |
|  | T1 | T1 | T2 | T1 | T2 | T1 |
| ME | 3/25/2016 | 3/10/2017 | 08/06/2017 | 4/16/2018 | 10/23/2018 | 12/13/2019 |
| FS | 3/25/2016 | 3/10/2017 | 08/06/2017 | 4/13/2018 | 10/31/2018 | 12/13/2019 |
| MT | 3/21/2016 | 3/3/2017 | 07/06/2017 | 4/15/2018 | 10/22/2018 | 12/14/2019 |
| GK | 3/16/2016 | 3/1/2017 | 07/06/2017 | 4/14/2018 | 10/24/2018 | 12/14/2019 |
| ST | 3/22/2016 | 3/9/2017 | 09/06/2017 | 4/13/2018 | 10/20/2018 | 12/12/2019 |
| GG | 3/17/2016 | 1/5/2017 | 09/06/2017 | 4/12/2018 | 10/22/2018 | 12/16/2019 |
| MG | 3/24/2016 | 1/6/2017 | 09/06/2017 | 4/12/2018 | 10/23/2018 | 12/16/2019 |
| DF | 3/19/2016 | 2/28/2017 | 6/11/2017 | 4/11/2018 | 10/22/2018 | 12/12/2019 |
| MA | 3/15/2016 | 1/10/2017 | 6/8/2017 | 4/17/2018 | 10/22/2018 | 12/11/2019 |
| DT | 3/21/2016 | 2/14/2017 | 6/8/2017 | 4/17/2018 | 11/6/2018 | 12/11/2019 |
| NE | 5/31/2016 | 4/24/2017 | 6/19/2017 | 4/24/2018 | 10/30/2018 | 1/21/2020 |
| MB | 5/31/2016 | 3/23/2017 | 6/19/2017 | 4/23/2018 | 11/6/2018 | 1/22/2020 |
| MO | 5/30/2016 | 4/25/2017 | 6/16/2017 | 4/26/2018 | 11/7/2018 | 1/24/2020 |
| LR | 5/31/2016 | 4/25/2017 | 6/16/2017 | 4/19/2018 | 11/8/2018 | 1/23/2020 |
| NM | 6/1/2016 | 3/24/2017 | 6/15/2017 | 4/19/2018 | 11/6/2018 | 1/16/2020 |
| MD | 5/25/2016 | 3/23/2017 | 6/15/2017 | 4/18/2018 | 10/30/2018 | 1/15/2020 |

**Table S22.** Descriptive statistics for ln-transformed non-emergent vegetation mass (g) at the sweep-level for each year, time (before or after vegetation manipulation), and manipulation treatment.

| Year | Time | Treatment | N | Average | Std.<br>dev. | Std.<br>error | Median | Lower<br>95%CI | Upper<br>95%CI |
| --- | --- | --- | --- | --- | --- | --- | --- | --- | --- |
| 2017 | Before | Control | 74 | 6.10 | 2.44 | 0.28 | 6.96 | 5.54 | 6.66 |
| 2017 | Before | Removal | 101 | 5.00 | 2.94 | 0.29 | 6.48 | 4.43 | 5.58 |
| 2017 | After | Control | 73 | 6.63 | 2.25 | 0.26 | 7.33 | 6.11 | 7.15 |
| 2017 | After | Removal | 119 | 0.67 | 1.57 | 0.14 | 0.00 | 0.38 | 0.95 |
| 2018 | Before | Control | 240 | 2.51 | 3.06 | 0.20 | 0.00 | 2.12 | 2.90 |
| 2018 | Before | Removal | 240 | 2.97 | 3.14 | 0.20 | 2.09 | 2.57 | 3.37 |
| 2018 | After | Control | 225 | 2.33 | 2.56 | 0.17 | 1.79 | 1.99 | 2.66 |
| 2018 | After | Removal | 225 | 2.01 | 2.35 | 0.16 | 0.00 | 1.70 | 2.32 |
| 2019 | Before | Control | 225 | 2.61 | 2.83 | 0.19 | 2.40 | 2.24 | 2.99 |
| 2019 | Before | Removal | 240 | 2.55 | 3.15 | 0.20 | 0.00 | 2.15 | 2.95 |
| 2019 | After | Control | 239 | 2.82 | 3.07 | 0.20 | 2.40 | 2.43 | 3.21 |
| 2019 | After | Removal | 240 | 2.66 | 3.05 | 0.20 | 0.00 | 2.27 | 3.05 |

**Table S23.** Descriptive statistics for aquatic vegetation mass (kg) removed at the water access point-level for each removal round (1-10).

| Removal | N | Average | Std. dev. | Std. error | Median | Lower 95%CI | Upper 95%CI |
| --- | --- | --- | --- | --- | --- | --- | --- |
| 1 | 20 | 14212 | 188867.7 | 4576.1 | 6788 | 4511 | 23914 |
| 2 | 15 | 1450 | 1416.7 | 378.6 | 891 | 632 | 2268 |
| 3 | 16 | 2110 | 2010.5 | 502.6 | 1513 | 1039 | 3181 |
| 4 | 15 | 2563 | 2541.5 | 679.3 | 1439 | 1096 | 4031 |
| 5 | 16 | 2192 | 1616.7 | 448.4 | 1591 | 1215 | 3169 |
| 6 | 7 | 1457 | 1144.3 | 432.5 | 827 | 399 | 2516 |
| 7 | 7 | 1357 | 1139.9 | 430.8 | 1181 | 303 | 2411 |
| 8 | 7 | 2628 | 1537.7 | 581.2 | 2110 | 1207 | 4051 |
| 9 | 7 | 2597 | 1598.5 | 604.2 | 2453 | 1119 | 4076 |
| 10 | 8 | 2012 | 2578.9 | 974.7 | 978 | -372 | 4397 |

**Table S24.** AICc comparison of a zero-inflated negative binomial (ZINB), negative binomial (NB), and Poisson (P) mixed effects model of total snail counts.

| Model | K | AICc | ΔAICc |
| --- | --- | --- | --- |
| ZINB | 10 | 3841 | 0.00 |
| NB | 8 | 3923 | 82 |
| P | 7 | 5504 | 1663 |

**Table S25.** Zero-inflated negative binomial model results for total snail counts one year after versus one year before vegetation removal. Fixed effects include treatment (removal or control), time (before or after), an interaction term between treatment and time, and a random term for point location nested within water access point within year to account for resampling.

| Predictor | $X^2$ | *df* | *p*-value |
| --- | --- | --- | --- |
| Treatment | 1.47 | 1 | 0.22 |
| Time | 55.38 | 1 | <0.001 |
| Treatment : Time | 40.96 | 1 | <0.001 |

**Table S26.** Descriptive statistics for *Bulinus* snail counts at the sweep-level for each year, time (before or after vegetation manipulation), and manipulation treatment.

| Year | Time | Treatment | N | Average | Std.<br>dev. | Std.<br>error | Median | Lower<br>95%CI | Upper<br>95%CI |
| --- | --- | --- | --- | --- | --- | --- | --- | --- | --- |
| 2017 | Before | Control | 74 | 6.03 | 10.52 | 1.22 | 2 | 3.61 | 8.45 |
| 2017 | Before | Removal | 101 | 11.39 | 29.38 | 2.92 | 0 | 5.60 | 17.17 |
| 2017 | After | Control | 73 | 4.41 | 13.80 | 1.62 | 0 | 1.21 | 7.61 |
| 2017 | After | Removal | 119 | 0.24 | 1.16 | 0.11 | 0 | 0.03 | 0.46 |
| 2018 | Before | Control | 240 | 1.70 | 5.23 | 0.34 | 0 | 1.03 | 2.37 |
| 2018 | Before | Removal | 240 | 2.70 | 7.42 | 0.48 | 0 | 1.76 | 3.65 |
| 2018 | After | Control | 225 | 1.68 | 3.95 | 0.26 | 0 | 1.15 | 2.20 |
| 2018 | After | Removal | 225 | 0.67 | 2.27 | 0.15 | 0 | 0.37 | 0.97 |
| 2019 | Before | Control | 225 | 2.51 | 12.56 | 0.84 | 0 | 0.85 | 4.17 |
| 2019 | Before | Removal | 240 | 3.03 | 11.59 | 0.75 | 0 | 1.54 | 4.51 |
| 2019 | After | Control | 239 | 2.37 | 8.82 | 0.57 | 0 | 1.24 | 3.50 |
| 2019 | After | Removal | 240 | 2.00 | 6.02 | 0.39 | 0 | 1.23 | 2.77 |

**Table S27.** Descriptive statistics for *Biomphalaria* snail counts at the sweep-level for each year, time (before or after vegetation manipulation), and manipulation treatment.

| Year | Time | Treatment | N | Average | Std.<br>dev. | Std.<br>error | Median | Lower<br>95%CI | Upper<br>95%CI |
| --- | --- | --- | --- | --- | --- | --- | --- | --- | --- |
| 2017 | Before | Control | 74 | 0.57 | 2.30 | 0.27 | 0 | 0.04 | 1.10 |
| 2017 | Before | Removal | 101 | 1.75 | 5.18 | 0.52 | 0 | 0.73 | 2.77 |
| 2017 | After | Control | 73 | 0.79 | 2.54 | 0.30 | 0 | 0.21 | 1.38 |
| 2017 | After | Removal | 119 | 0.03 | 0.29 | 0.03 | 0 | 0.00 | 0.09 |
| 2018 | Before | Control | 240 | 0.55 | 2.41 | 0.16 | 0 | 0.24 | 0.86 |
| 2018 | Before | Removal | 240 | 0.41 | 1.95 | 0.13 | 0 | 0.16 | 0.66 |
| 2018 | After | Control | 225 | 0.21 | 0.90 | 0.06 | 0 | 0.10 | 0.33 |
| 2018 | After | Removal | 225 | 0.27 | 1.11 | 0.07 | 0 | 0.12 | 0.42 |
| 2019 | Before | Control | 225 | 0.41 | 1.94 | 0.13 | 0 | 0.15 | 0.67 |
| 2019 | Before | Removal | 240 | 2.14 | 11.67 | 0.75 | 0 | 0.65 | 3.63 |
| 2019 | After | Control | 239 | 1.09 | 5.09 | 0.33 | 0 | 0.44 | 1.74 |
| 2019 | After | Removal | 240 | 1.99 | 8.32 | 0.54 | 0 | 0.93 | 3.06 |

**Table S28.** Descriptive statistics for total snail counts at the sweep-level for each year, time (before or after vegetation manipulation), and manipulation treatment.

| Year | Time | Treatment | N | Average | Std.<br>dev. | Std.<br>error | Median | Lower<br>95% CI | Upper<br>95% CI |
| --- | --- | --- | --- | --- | --- | --- | --- | --- | --- |
| 2017 | Before | Control | 74 | 6.59 | 11.01 | 1.28 | 3 | 4.06 | 9.13 |
| 2017 | Before | Removal | 101 | 13.14 | 31.37 | 3.12 | 0 | 6.96 | 19.32 |
| 2017 | After | Control | 73 | 5.21 | 15.82 | 1.85 | 0 | 1.54 | 8.87 |
| 2017 | After | Removal | 119 | 0.28 | 1.21 | 0.11 | 0 | 0.06 | 0.50 |
| 2018 | Before | Control | 240 | 2.25 | 5.99 | 0.39 | 0 | 1.48 | 3.02 |
| 2018 | Before | Removal | 240 | 3.11 | 7.76 | 0.50 | 0 | 2.12 | 4.10 |
| 2018 | After | Control | 225 | 1.89 | 4.11 | 0.27 | 0 | 1.35 | 2.43 |
| 2018 | After | Removal | 225 | 0.94 | 2.76 | 0.18 | 0 | 0.58 | 1.31 |
| 2019 | Before | Control | 225 | 2.92 | 13.12 | 0.87 | 0 | 1.19 | 4.65 |
| 2019 | Before | Removal | 240 | 5.17 | 18.03 | 1.16 | 0 | 2.86 | 7.47 |
| 2019 | After | Control | 239 | 3.46 | 10.87 | 0.70 | 0 | 2.06 | 4.85 |
| 2019 | After | Removal | 240 | 3.99 | 10.67 | 0.69 | 0 | 2.63 | 5.35 |

**Table S29.** Estimated marginal means for the probability of infection and egg burden by *S. mansoni and S. haematobium* at the child level for each time (before or after vegetation manipulation) and treatment (control or vegetation removal).

| Endpoint | Intervention | Time | N | Average | Std. error | Lower 95% CI | Upper 95% CI |
| --- | --- | --- | --- | --- | --- | --- | --- |
| *Schistosoma mansoni* |  |  |  |  |  |  |  |
| Probability of infection | control | Before | 638 | 0.11 | 0.05 | 0.04 | 0.27 |
|  | control | After | 457 | 0.21 | 0.09 | 0.08 | 0.45 |
|  | removal | Before | 660 | 0.19 | 0.09 | 0.07 | 0.41 |
|  | removal | After | 511 | 0.18 | 0.08 | 0.07 | 0.40 |
| Egg burden (natural log transformed) | control | Before | 638 | 0.65 | 0.49 | -0.04 | 1.70 |
|  | control | After | 457 | 0.49 | 0.49 | 0.07 | 2.18 |
|  | removal | Before | 660 | 0.49 | 0.49 | -0.06 | 2.05 |
|  | removal | After | 511 | 0.49 | 0.49 | -0.05 | 2.06 |
| *Schistosoma haematobium* |  |  |  |  |  |  |  |
| Probability of infection | control | Before | 638 | 0.76 | 0.08 | 0.57 | 0.89 |
|  | control | After | 457 | 0.71 | 0.10 | 0.50 | 0.86 |
|  | removal | Before | 660 | 0.68 | 0.10 | 0.47 | 0.83 |
|  | removal | After | 511 | 0.68 | 0.10 | 0.47 | 0.84 |
| Egg burden (natural log transformed) | control | Before | 638 | 1.87 | 0.34 | 1.16 | 2.58 |
|  | control | After<sup>a</sup> | 457 | 1.87 | 0.35 | 1.16 | 2.58 |
|  | removal | Before | 660 | 1.82 | 0.34 | 1.11 | 2.53 |
|  | removal | After<sup>a</sup> | 511 | 2.02 | 0.34 | 1.31 | 2.73 |

<sup>a</sup>Includes 31 samples from after ( $n=18$ controls, $n=13$ removals) that were censored at 200 eggs.

**Table S30.** BACI one year before/after vegetation removal at 16 sites for *S. mansoni* and *S. haematobium* egg burden (negative binomial GLMM) and prevalence (binomial GLMM) with school by intervention time as random effect (*n*=2266).

| Statistical model | *Schistosoma mansoni* |  |  |  | *Schistosoma haematobium* |  |  |  |
| --- | --- | --- | --- | --- | --- | --- | --- | --- |
|  | Estimate | Std. error | z/t-value | *p*-value | Estimate | Std. error | z/t-value | *p*-value |
| Parasite prevalence |  |  |  |  |  |  |  |  |
| Intervention.time | 0.78 | 0.25 | 3.11 | 0.04 | -0.27 | 0.46 | -0.58 | 0.74 |
| Intervention.type | 0.63 | 0.78 | 0.82 | 0.75 | -0.44 | 0.63 | -0.69 | 0.60 |
| Sex (Male) | 0.21 | 0.12 | 1.76 | 0.08 | 0.44 | 0.11 | 3.98 | <0.001 |
| River (as compared to lake) | -1.02 | 0.79 | -1.31 | 0.19 | -2.50 | 0.57 | -4.36 | <0.001 |
| School Class | -0.003 | 0.07 | -0.04 | 0.97 | -0.13 | 0.06 | -1.97 | 0.049 |
| Intervention.time : Intervention.type | -0.81 | 0.35 | -2.30 | 0.02 | 0.31 | 0.64 | 0.48 | 0.63 |
| Parasite egg counts (natural log transformed) |  |  |  |  |  |  |  |  |
| Intervention.time | 0.47 | 0.21 | 2.28 | 0.10 | -0.002 | 0.35 | -0.01 | 0.68 |
| Intervention.type | 0.34 | 0.61 | 0.56 | 0.84 | -0.06 | 0.44 | -0.12 | 0.90 |
| Sex (Male) | 0.15 | 0.06 | 2.51 | 0.01 | 0.30 | 0.07 | 4.41 | <0.001 |
| River (as compared to lake) | -0.85 | 0.62 | -1.37 | 0.17 | -1.64 | 0.37 | -4.41 | <0.001 |
| School Class | -0.01 | 0.03 | -0.30 | 0.76 | -0.27 | 0.04 | -7.19 | <0.001 |
| Intervention.time : Intervention.type | -0.46 | 0.29 | -1.58 | 0.11 | 0.20 | 0.50 | 0.42 | 0.67 |

**Table S31.** Model selection results using Akaike Information Criteria (AIC) for all possible models evaluating *S. mansoni* prevalence. A “*” in the variable columns indicates that the variable was included in that model. K is the number of variables included in each model. All GLMM models included the random term of School by intervention type. Table includes the top ten models by AIC.

| # | Intervention.time | Intervention.type | Intervention.time:<br>Intervention.type | Sex<br>(M) | River | Class<br>Number | K | AIC | Δ AIC | weight |
| --- | --- | --- | --- | --- | --- | --- | --- | --- | --- | --- |
| 1 | * | * | * | * |  |  | 4 | 1861.5 | 0.00 | 0.09 |
| 2 | * | * | * | * | * |  | 5 | 1861.9 | 0.40 | 0.08 |
| 3 | * |  |  | * |  |  | 2 | 1862.4 | 0.85 | 0.06 |
| 4 | * | * | * |  |  |  | 3 | 1862.5 | 1.00 | 0.06 |
| 5 |  |  |  | * |  |  | 1 | 1862.7 | 1.13 | 0.05 |
| 6 | * |  |  | * | * |  | 2 | 1862.8 | 1.23 | 0.05 |
| 7 | * | * | * |  | * |  | 4 | 1863.0 | 1.44 | 0.05 |
| 8 |  |  |  | * | * |  | 2 | 1863.0 | 1.47 | 0.04 |
| 9 | * |  |  |  |  |  | 1 | 1863.4 | 1.84 | 0.04 |
| 10 | * | * | * | * |  | * | 5 | 1863.5 | 2.01 | 0.03 |
| Sum of<br>weights | 0.70 | 0.54 | 0.37 | 0.62 | 0.45 | 0.27 |  |  |  |  |
| Average<br>estimates<br>from<br>models <sup>a</sup> | 0.51 | 0.34 | -0.42 | 0.15 | -0.43 | NI |  |  |  |  |

<sup>a</sup> - averaging all models with Delta AIC < 2; NI=not included

**Table S32.** Model selection results using Akaike Information Criteria (AIC) for all possible models evaluating *S. mansoni* egg count. A “*” in the variable columns indicates that the variable was included in that model. K is the number of variables included in each model. All NB GLMM models included the random term of School by intervention type. Table includes the top ten models by AIC.

| # | Intervention.time | Intervention.type | Intervention.time:<br>Intervention.type | Sex<br>(M) | River | Class<br>Number | K | AIC | Δ AIC | weight |
| --- | --- | --- | --- | --- | --- | --- | --- | --- | --- | --- |
| 1 | * |  |  | * |  |  | 2 | 7987.7 | 0.00 | 0.11 |
| 2 |  |  |  | * |  |  | 1 | 7987.9 | 0.22 | 0.10 |
| 3 | * |  |  | * | * |  | 3 | 7987.9 | 0.27 | 0.10 |
| 4 |  |  |  | * | * |  | 2 | 7988.1 | 0.48 | 0.09 |
| 5 | * | * | * | * |  |  | 4 | 7989.3 | 1.65 | 0.05 |
| 6 | * | * | * | * | * |  | 5 | 7989.6 | 1.91 | 0.04 |
| 7 | * |  |  | * |  | * | 3 | 7989.6 | 1.93 | 0.04 |
| 8 | * | * |  | * |  |  | 3 | 7989.6 | 1.98 | 0.04 |
| 9 |  |  |  | * |  | * | 2 | 7989.8 | 2.13 | 0.04 |
| 10 | * |  |  | * | * | * | 4 | 7989.8 | 2.17 | 0.04 |
| Sum of<br>weights | 0.59 | 0.38 | 0.14 | 0.89 | 0.47 | 0.28 |  |  |  |  |
| Average<br>estimates<br>from<br>models<sup>a</sup> | 0.20 | 0.06 | -0.07 | 0.15 | -0.34 | -0.001 |  |  |  |  |

<sup>a</sup> - averaging all models with Delta AIC < 2; NI=not included

**Table S33.**
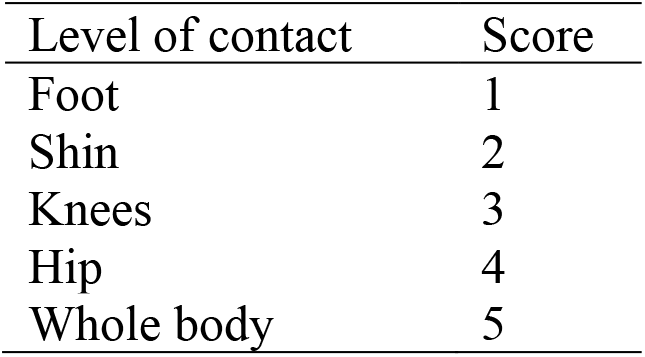
Ordinal multinomial level of water contact recorded during human water contact observations.

| Level of contact | Score |
| --- | --- |
| Foot | 1 |
| Shin | 2 |
| Knees | 3 |
| Hip | 4 |
| Whole body | 5 |

**Table S34.** Date and time of observations of human water access contact behaviors at two control (Ndiawdoune, Mbarigot) and manipulation sites (Maka Diama, Mbakhana) in relation to the date of the most recent vegetation removal rounds. Manipulation sites are shown in bold font. Observation time was held constant for each site for all observations.

| Date | Observation time | Site | Date of the last veg. removal |
| --- | --- | --- | --- |
| 10/3/2018 | 10:00 am -11:00 am | Ndiawdoune |  |
| 10/3/2018 | 11:30 am-12:30 pm | **Maka Diama 1** |  |
| 10/3/2018 | 3:30 pm-4:30 pm | **Mbakhana** |  |
| 10/3/2018 | 5:00 pm-6:00 pm | Mbarigo 1 |  |
| 10/4/2018 | 10:00 am -11:00 am | Ndiawdoune |  |
| 10/4/2018 | 11:30 am-12:30 pm | **Maka Diama 1** |  |
| 10/4/2018 | 3:30 pm-4:30 pm | **Mbakhana** |  |
| 10/4/2018 | 5:00 pm-6:00 pm | Mbarigo 1 |  |
| 10/29/2018 | 10:00 am -11:00 am | Ndiawdoune |  |
| 10/29/2018 | 11:30 am-12:30 pm | **Maka Diama 1** | 10/6/2018 |
| 10/29/2018 | 3:30 pm-4:30 pm | **Mbakhana** | 10/14/2018 |
| 10/29/2018 | 5:00 pm-6:00 pm | Mbarigo 1 |  |
| 10/30/2018 | 10:00 am -11:00 am | Ndiawdoune |  |
| 10/30/2018 | 11:30 am-12:30 pm | **Maka Diama 1** | 10/6/2018 |
| 10/30/2018 | 3:30 pm-4:30 pm | **Mbakhana** | 10/14/2018 |
| 10/30/2018 | 5:00 pm-6:00 pm | Mbarigo 1 |  |
| 5/6/2019 | 10:00 am -11:00 am | Ndiawdoune |  |
| 5/6/2019 | 11:30 am-12:30 pm | **Maka Diama 1** | 1/30/2019 |
| 5/6/2019 | 3:30 pm-4:30 pm | **Mbakhana** | 1/26/2019 |
| 5/6/2019 | 5:00 pm-6:00 pm | Mbarigo 1 |  |
| 5/7/2019 | 10:00 am -11:00 am | Ndiawdoune |  |
| 5/7/2019 | 11:30 am-12:30 pm | **Maka Diama 1** | 1/30/2019 |
| 5/7/2019 | 3:30 pm-4:30 pm | **Mbakhana** | 1/26/2019 |
| 5/7/2019 | 5:00 pm-6:00 pm | Mbarigo 1 |  |
| 5/30/2019 | 10:00 am -11:00 am | Ndiawdoune |  |
| 5/30/2019 | 11:30 am-12:30 pm | **Maka Diama 1** | 5/16/2019 |
| 5/30/2019 | 3:30 pm-4:30 pm | **Mbakhana** | 5/20/2019 |
| 5/30/2019 | 5:00 pm-6:00 pm | Mbarigo 1 |  |
| 5/31/2019 | 10:00 am -11:00 am | Ndiawdoune |  |
| 5/31/2019 | 11:30 am-12:30 pm | **Maka Diama 1** | 5/16/2019 |
| 5/31/2019 | 3:30 pm-4:30 pm | **Mbakhana** | 5/20/2019 |
| 5/31/2019 | 5:00 pm-6:00 pm | Mbarigo 1 |  |

**Table S35.** Count of water contact observations by level of water immersion (ordinal score 1-5), child gender and class (1-3) for each manipulation treatment and time.

| Gender | Time | Class | Treatment | Level of water immersion (score 1-5) |  |  |  |  |
| --- | --- | --- | --- | --- | --- | --- | --- | --- |
|  |  |  |  | 1 | 2 | 3 | 4 | 5 |
| F | after | 1 | control | 2 | 6 | 7 | 2 | 3 |
| F | after | 1 | removal | 1 | 3 | 5 | 0 | 27 |
| F | after | 2 | control | 0 | 9 | 7 | 0 | 4 |
| F | after | 2 | removal | 0 | 7 | 22 | 0 | 4 |
| F | after | 3 | control | 0 | 9 | 6 | 1 | 1 |
| F | after | 3 | removal | 1 | 9 | 7 | 0 | 11 |
| F | before | 1 | control | 8 | 0 | 9 | 0 | 6 |
| F | before | 1 | removal | 3 | 3 | 6 | 3 | 23 |
| F | before | 2 | control | 7 | 3 | 6 | 0 | 5 |
| F | before | 2 | removal | 3 | 4 | 6 | 1 | 13 |
| F | before | 3 | control | 2 | 2 | 7 | 0 | 2 |
| F | before | 3 | removal | 2 | 5 | 8 | 3 | 6 |
| M | after | 1 | control | 0 | 11 | 2 | 1 | 24 |
| M | after | 1 | removal | 1 | 5 | 0 | 0 | 51 |
| M | after | 2 | control | 0 | 5 | 1 | 0 | 1 |
| M | after | 2 | removal | 0 | 1 | 1 | 0 | 3 |
| M | after | 3 | control | 0 | 6 | 2 | 1 | 0 |
| M | after | 3 | removal | 0 | 3 | 4 | 0 | 1 |
| M | before | 1 | control | 1 | 1 | 4 | 6 | 8 |
| M | before | 1 | removal | 1 | 5 | 6 | 0 | 37 |
| M | before | 2 | control | 2 | 2 | 7 | 0 | 0 |
| M | before | 2 | removal | 0 | 0 | 2 | 0 | 5 |
| M | before | 3 | control | 0 | 1 | 2 | 1 | 2 |
| M | before | 3 | removal | 0 | 3 | 0 | 0 | 4 |

**Table S36.** Average duration of water contact (min) by level of water immersion (ordinal score 1-5), child gender and class (1-3) for each manipulation treatment and time.

| Gender | Time | Class | Treatment | Level of water immersion<br>(scores 1-5) |  |  |  |  |
| --- | --- | --- | --- | --- | --- | --- | --- | --- |
|  |  |  |  | 1 | 2 | 3 | 4 | 5 |
| F | after | 1 | control | 13.0 | 14.5 | 27.6 | 25.0 | 29.0 |
| F | after | 1 | removal | 1.0 | 48.7 | 23.0 | 0.0 | 26.1 |
| F | after | 2 | control | 0.0 | 12.9 | 16.1 | 0.0 | 21.5 |
| F | after | 2 | removal | 0.0 | 27.7 | 19.2 | 0.0 | 27.5 |
| F | after | 3 | control | 0.0 | 12.9 | 17.7 | 20.0 | 46.0 |
| F | after | 3 | removal | 1.0 | 28.0 | 17.3 | 0.0 | 26.1 |
| F | before | 1 | control | 8.1 | 0.0 | 14.0 | 0.0 | 25.2 |
| F | before | 1 | removal | 2.0 | 28.3 | 21.7 | 4.0 | 22.7 |
| F | before | 2 | control | 16.7 | 28.3 | 15.7 | 0.0 | 24.2 |
| F | before | 2 | removal | 3.0 | 21.0 | 27.0 | 1.0 | 24.5 |
| F | before | 3 | control | 5.0 | 22.0 | 17.4 | 0.0 | 35.5 |
| F | before | 3 | removal | 33.0 | 6.6 | 20.1 | 27.0 | 20.8 |
| M | after | 1 | control | 0.0 | 10.6 | 20.0 | 38.0 | 16.6 |
| M | after | 1 | removal | 1.0 | 21.0 | 0.0 | 0.0 | 19.7 |
| M | after | 2 | control | 0.0 | 1.4 | 3.0 | 0.0 | 5.0 |
| M | after | 2 | removal | 0.0 | 6.0 | 22.0 | 0.0 | 32.7 |
| M | after | 3 | control | 0.0 | 6.7 | 6.0 | 23.0 | 0.0 |
| M | after | 3 | removal | 0.0 | 2.0 | 5.3 | 0.0 | 12.0 |
| M | before | 1 | control | 42.0 | 9.0 | 32.5 | 10.2 | 30.1 |
| M | before | 1 | removal | 4.0 | 11.6 | 11.7 | 0.0 | 33.6 |
| M | before | 2 | control | 1.5 | 1.5 | 1.3 | 0.0 | 0.0 |
| M | before | 2 | removal | 0.0 | 0.0 | 3.5 | 0.0 | 6.8 |
| M | before | 3 | control | 0.0 | 5.0 | 7.5 | 4.0 | 7.5 |
| M | before | 3 | removal | 0.0 | 4.7 | 0.0 | 0.0 | 10.8 |

**Table S37.** BACI before/after vegetation removal at 16 sites for the level of water immersion (an ordinal variable 1-5).

| Predictor | Estimate | Std. error | z-value | *p-value* |
| --- | --- | --- | --- | --- |
| Intervention.type | 1.20 | 0.26 | 4.60 | <0.001 |
| Intervention.time | -0.11 | 0.28 | -0.38 | 0.71 |
| Time of Day | 0.04 | 0.04 | 1.01 | 0.31 |
| Sex (M) | 0.74 | 0.19 | 3.90 | <0.001 |
| 15-35 years old as compared to <15 | -1.12 | 0.21 | -5.29 | <0.001 |
| >35 years old as compared to <15 | -1.12 | 0.23 | -4.85 |  |
| Interventionremoval:Intervention.time | 0.02 | 0.38 | 0.05 | 0.96 |

**Table S38.** BACI before/after vegetation removal at 16 sites for the natural log transformed duration of water immersion (minutes).

| Predictor | Estimate | Std. error | t-value | *p-value* |
| --- | --- | --- | --- | --- |
| Intervention.type | 0.15 | 2.21 | 4.25 | 0.96 |
| Intervention.time | 0.02 | 0.13 | 0.14 | 0.81 |
| Time of Day | -0.53 | 0.13 | -4.03 | <0.001 |
| Sex (M) | -0.41 | 0.09 | -4.75 | <0.001 |
| 15-35 years old as compared to <15 | -0.47 | 0.10 | -4.52 | <0.001 |
| >35 years old as compared to <15 | -0.44 | 0.11 | -3.98 |  |
| Interventionremoval:Intervention.time | -0.06 | 0.17 | -0.37 | 0.71 |

**Table S39.**
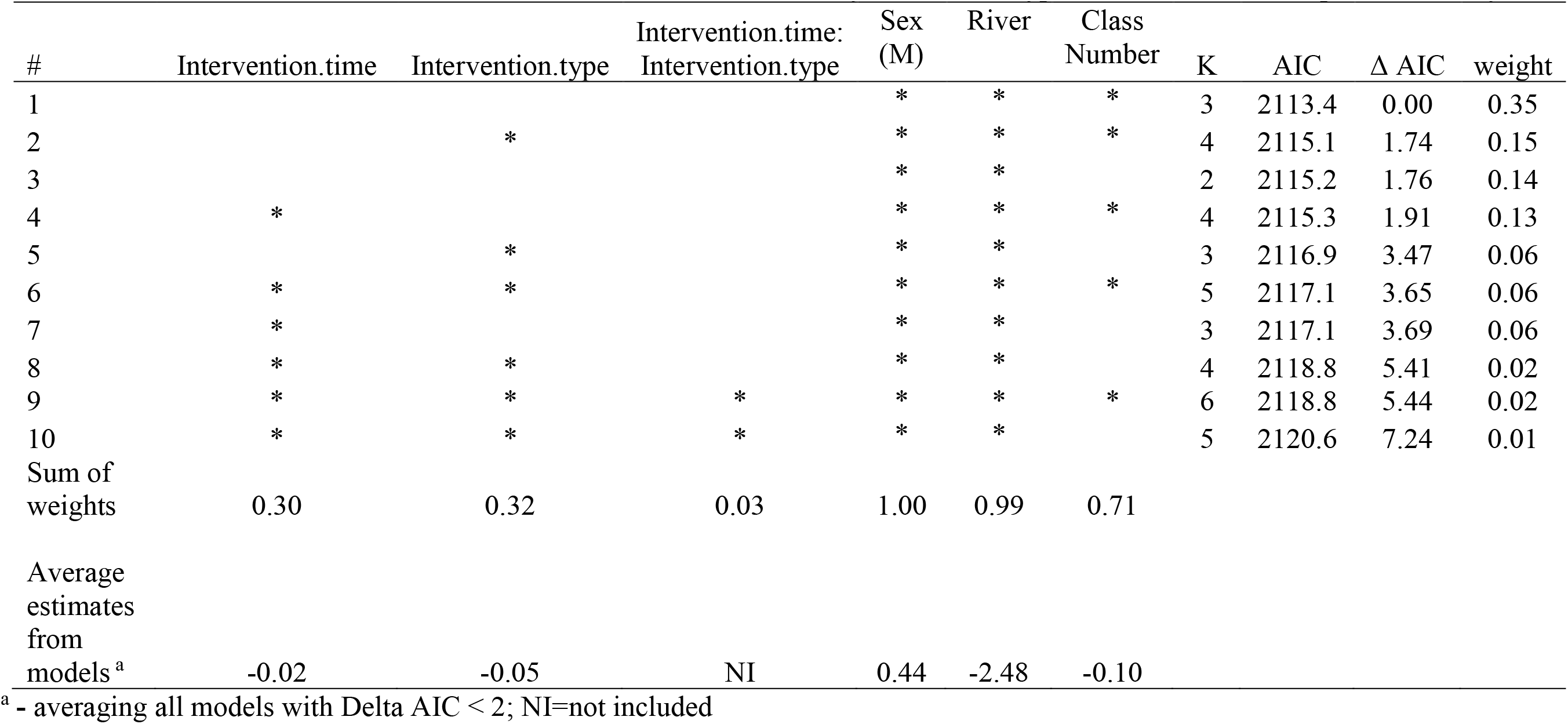
Model selection results using Akaike Information Criteria (AIC) for all possible models evaluating *S. haematobium* prevalence. A “*” in the variable columns indicates that the variable was included in that model. K is the number of variables included in each model. All GLMM models included the random term of School by intervention type. Table includes the top ten models by AIC.

**Table S40.**
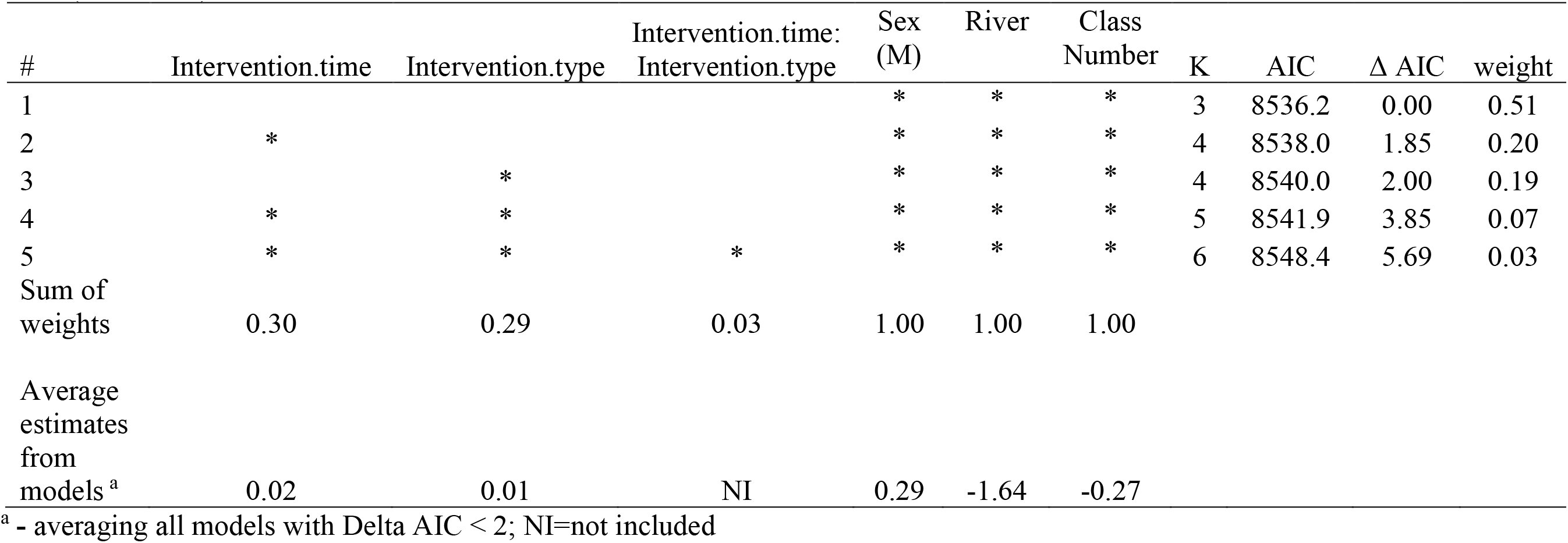
Model selection results using Akaike Information Criteria (AIC) for all possible models evaluating *S. haematobium* egg count. A “*” in the variable columns indicates that the variable was included in that model. K is the number of variables included in each model. All NB GLMM models included the random term of School by intervention type. Table includes the top five models by AIC (Δ AIC<10).

**Table S41.** Effects of vegetation removal and time on water quality and water chemistry in the before-after-control-impact experiment. Shown are mean values across water access sites with standard error in parentheses. Also shown are the *p*-values for the intervention-by-time interactions from mixed effect models with site included as a random intercept. No *p-*values are <0.05, indicating that there is little evidence to support the hypothesis that the vegetation removal significantly affected water quality or chemistry.

| Intervention | Time | N | Temperature | Conductivity | Salinity | Dissolved<br>Oxygen ppt | pH | Nitrate | Phytoplankton<br>Ft value | Periphyton<br>Ft value |
| --- | --- | --- | --- | --- | --- | --- | --- | --- | --- | --- |
| Control | Before | 16 | 29.3<br>(1.0) | 206.9<br>(142.9) | 0.10<br>(0.07) | 4.8<br>(3.5) | 7.1<br>(0.5) | 0.55<br>(0.88) | 351.1<br>(260.0) | 2476.1<br>(3790.6) |
| Control | After | 16 | 28.8<br>(1.9) | 180.1<br>(110.3) | 0.08<br>(0.05) | 5.6<br>(2.2) | 6.8<br>(0.4) | 0.63<br>(0.80) | 358.8<br>(283.7) | 3862.4<br>(3153.1) |
| Removal | Before | 23 | 29.5<br>(1.5) | 194.0<br>(77.5) | 0.09<br>(0.04) | 3.4<br>(2.5) | 7.5<br>(1.1) | 0.26<br>(0.27) | 260.6<br>(275.9) | 1869.2<br>(2381.4) |
| Removal | After | 23 | 27.7<br>(3.4) | 179.8<br>(75.7) | 0.13<br>(0.21) | 2.4<br>(2.3) | 7.3<br>(0.5) | 0.42<br>(0.58) | 372.5<br>(420.4) | 3085.3<br>(3289.7) |
| *p*-value for<br>Intervention<br>* time |  |  | 0.09 | 0.54 | 0.36 | 0.08 | 0.95 | 0.92 | 0.43 | 0.95 |

**Table S42.** Lab analyses of compost samples collected from three compost pits dug after the initial removal round.

| Sample ID | 1 | 2 | 3 |
| --- | --- | --- | --- |
| Moisture (%) | 11.8 | 19.3 | 12.5 |
| pH | 6.6 | 7.2 | 6.3 |
| Total Kjeldahl N (mg/L) | 10600 | 8693 | 11905 |
| Ammonia N (mg/L) | 69 | 18 | 56 |
| Elemental P (mg/L) | 1740 | 913 | 1164 |
| Elemental K (mg/L) | 13018 | 8525 | 4802 |
| Total Solids (mg/L) | 881876 | 807125 | 875297 |
| Total Ash (mg/L) | 537208 | 391640 | 398156 |
| Cu (mg/kg) | 20.1 | 12 | 15.5 |
| Mn (mg/kg) | 191 | 552.45 | 327.3 |
| Zn (mg/kg) | 50.1 | 21.5 | 43.6 |

**Table S43.** Descriptive statistics for pepper and onion counts and yields (kg) by compost treatment (NC= No compost, C = compost, TC = compost tilled into the soil), and fertilizer treatment (NU = No urea, U = Urea application), at the site-level.

| Site | Crop | Endpoint | Compost | Fertilizer | N | Average | Median | Std. dev. | Std. error | Lower 95%CI | Upper 95%CI |
| --- | --- | --- | --- | --- | --- | --- | --- | --- | --- | --- | --- |
| Lampsar | Pepper | Counts | C | NU | 3 | 168.00 | 112.00 | 165.28 | 95.42 | 0.00 | 356.94 |
| Lampsar | Pepper | Counts | C | U | 3 | 245.67 | 87.00 | 279.16 | 161.17 | 0.00 | 564.79 |
| Lampsar | Pepper | Counts | NC | NU | 3 | 14.67 | 6.00 | 15.01 | 8.67 | 0.00 | 31.83 |
| Lampsar | Pepper | Counts | NC | U | 3 | 11.33 | 7.00 | 8.39 | 4.84 | 1.75 | 20.92 |
| Lampsar | Pepper | Counts | TC | NU | 3 | 175.00 | 84.00 | 177.01 | 102.20 | 0.00 | 377.35 |
| Lampsar | Pepper | Counts | TC | U | 3 | 320.67 | 344.00 | 60.48 | 34.92 | 251.53 | 389.80 |
| Mbane | Pepper | Counts | C | NU | 3 | 424.00 | 425.00 | 105.50 | 60.91 | 303.39 | 544.61 |
| Mbane | Pepper | Counts | C | U | 3 | 352.67 | 347.00 | 74.66 | 43.11 | 267.32 | 438.02 |
| Mbane | Pepper | Counts | NC | NU | 3 | 374.33 | 380.00 | 132.59 | 76.55 | 222.76 | 525.91 |
| Mbane | Pepper | Counts | NC | U | 3 | 258.67 | 296.00 | 80.75 | 46.62 | 166.35 | 350.98 |
| Mbane | Pepper | Counts | TC | NU | 3 | 391.33 | 366.00 | 77.18 | 44.56 | 303.10 | 479.57 |
| Mbane | Pepper | Counts | TC | U | 3 | 364.33 | 339.00 | 80.06 | 46.23 | 272.81 | 455.86 |
| Lampsar | Onion | Counts | C | NU | 3 | 484.33 | 492.00 | 54.90 | 31.70 | 421.57 | 547.10 |
| Lampsar | Onion | Counts | C | U | 3 | 208.67 | 211.00 | 40.55 | 23.41 | 162.31 | 255.02 |
| Lampsar | Onion | Counts | NC | NU | 3 | 190.00 | 165.00 | 56.79 | 32.79 | 125.08 | 254.92 |
| Lampsar | Onion | Counts | NC | U | 3 | 83.33 | 79.00 | 15.95 | 9.21 | 65.10 | 101.56 |
| Lampsar | Onion | Counts | TC | NU | 3 | 497.67 | 493.00 | 43.19 | 24.94 | 448.29 | 547.04 |
| Lampsar | Onion | Counts | TC | U | 3 | 286.00 | 281.00 | 17.06 | 9.85 | 266.50 | 305.50 |
| Lampsar | Pepper | Yields | C | NU | 3 | 1.54 | 1.36 | 0.94 | 0.54 | 0.47 | 2.61 |
| Lampsar | Pepper | Yields | C | U | 3 | 1.92 | 1.43 | 1.00 | 0.58 | 0.78 | 3.06 |
| Lampsar | Pepper | Yields | NC | NU | 3 | 0.34 | 0.17 | 0.30 | 0.17 | 0.00 | 0.68 |
| Lampsar | Pepper | Yields | NC | U | 3 | 0.25 | 0.25 | 0.13 | 0.08 | 0.10 | 0.40 |
| Lampsar | Pepper | Yields | TC | NU | 3 | 1.61 | 1.13 | 0.88 | 0.51 | 0.61 | 2.61 |
| Lampsar | Pepper | Yields | TC | U | 3 | 2.50 | 2.58 | 0.23 | 0.13 | 2.24 | 2.76 |
| Mbane | Pepper | Yields | C | NU | 3 | 2.60 | 2.63 | 0.18 | 0.11 | 2.39 | 2.80 |
| Mbane | Pepper | Yields | C | U | 3 | 2.30 | 2.35 | 0.26 | 0.15 | 2.00 | 2.60 |
| Mbane | Pepper | Yields | NC | NU | 3 | 2.44 | 2.42 | 0.45 | 0.26 | 1.92 | 2.95 |
| Mbane | Pepper | Yields | NC | U | 3 | 2.13 | 2.18 | 0.35 | 0.20 | 1.73 | 2.54 |
| Mbane | Pepper | Yields | TC | NU | 3 | 2.48 | 2.44 | 0.21 | 0.12 | 2.24 | 2.71 |
| Mbane | Pepper | Yields | TC | U | 3 | 2.28 | 2.19 | 0.27 | 0.16 | 1.97 | 2.60 |
| Lampsar | Onion | Yields | C | NU | 3 | 2.94 | 2.92 | 0.16 | 0.09 | 2.77 | 3.12 |
| Lampsar | Onion | Yields | C | U | 3 | 2.92 | 2.94 | 0.27 | 0.16 | 2.61 | 3.23 |
| Lampsar | Onion | Yields | NC | NU | 3 | 1.20 | 0.99 | 0.43 | 0.25 | 0.71 | 1.70 |
| Lampsar | Onion | Yields | NC | U | 3 | 1.43 | 1.25 | 0.45 | 0.26 | 0.92 | 1.95 |
| Lampsar | Onion | Yields | TC | NU | 3 | 3.15 | 3.24 | 0.35 | 0.20 | 2.75 | 3.55 |
| Lampsar | Onion | Yields | TC | U | 3 | 3.23 | 3.22 | 0.04 | 0.02 | 3.18 | 3.28 |

**Table S44.** Mixed model results for pepper counts (negative binomial error) and ln-transformed yields (Gaussian error) for tilled compost and compost treatment (as compared to no compost), fertilizer treatment (Urea), and site (Mbane and Lampsar) with mixed effect for harvest round

| Predictor | Estimate | Std. error | z-value | *p-value* |
| --- | --- | --- | --- | --- |
| Counts |  |  |  |  |
| Compost (as compared to. no compost) | 0.91 | 0.17 | 5.10 | <0.001 |
| Tilled Compost (as compared to no compost) | 0.92 | 0.18 | 5.16 |  |
| Urea.Treatment | -0.31 | 0.17 | -1.82 | 0.46 |
| Site Mbane | 1.46 | 0.12 | 12.32 | <0.001 |
| Compost : Urea.Treatment | 0.47 | 0.24 | 1.91 | 0.01 |
| Tilled Compost : Urea.Treatment | 0.70 | 0.24 | 2.87 |  |
| Yields (weight) |  |  |  |  |
| Compost (as compared to. no compost) | 1.96 | 0.30 | 6.54 | <0.001 |
| Tilled Compost (as compared to no compost) | 2.05 | 0.30 | 6.86 |  |
| Urea.Treatment | 0.006 | 0.30 | 0.02 | 0.54 |
| Site Mbane | 2.24 | 0.18 | 12.46 | <0.001 |
| Compost : Urea.Treatment | 0.05 | 0.42 | 0.13 | 0.79 |
| Tilled Compost : Urea.Treatment | 0.57 | 0.42 | 1.34 |  |

**Table S45.** Model results for onion counts (negative binomial error) and ln-transformed yields (Gaussian error) for tilled compost and compost treatment (as compared to no compost) and fertilizer treatment (Urea). Onions were grown in only one site and one round of harvest.

| Predictor | Estimate | Std. error | z/t-value | *p-value* |
| --- | --- | --- | --- | --- |
| Counts |  |  |  |  |
| Compost (as compared to. no compost) | 0.93 | 0.11 | 8.70 | <0.001 |
| Tilled Compost (as compared to no compost) | 0.96 | 0.11 | 8.96 |  |
| Urea.Treatment | -0.82 | 0.12 | -6.75 | <0.001 |
| Compost : Urea.Treatment | -0.01 | 0.16 | -0.11 | 0.10 |
| Tilled Compost : Urea.Treatment | 0.27 | 0.16 | 1.68 |  |
| Yields (weight) |  |  |  |  |
| Compost (as compared to. no compost) | 1.74 | 0.26 | 6.65 | <0.001 |
| Tilled Compost (as compared to no compost) | 1.94 | 0.26 | 7.45 |  |
| Urea.Treatment | 0.23 | 0.26 | 0.87 | 0.54 |
| Compost : Urea.Treatment | -0.25 | 0.37 | -0.68 | 0.79 |
| Tilled Compost : Urea.Treatment | -0.14 | 0.37 | -0.40 |  |
| Disease (Onion Rot) |  |  |  |  |
| Compost (as compared to. no compost) | -5.12e-16 | 1.09 | 0.00 | 1.00 |
| Tilled Compost (as compared to no compost) | -1.02e-15 | 1.09 | 0.00 |  |
| Urea.Treatment | -1.68e-15 | 1.09 | 0.00 | 1.00 |
| Compost : Urea.Treatment | 8.50 | 1.55 | 5.50 | <0.001 |
| Tilled Compost : Urea.Treatment | 7.00 | 1.55 | 14.53 |  |

**Table S46.** Estimates of the expected ratio of marginal revenue product (MRP, including private gains and public health benefit) to marginal cost (MC) for private gains of a metric ton of compost. Mean estimates are produced using the mean estimated marketing margin between Saint Louis and Dakar. Upper MRP estimates use the mean plus (minus) one standard deviation. All prices have been converted into USD.

| Wage rate | Crop | Treatment | Mean MRP/MC (metric ton compost) | Upper MRP/MC (metric ton compost) | Lower MRP/MC (metric ton compost) |
| --- | --- | --- | --- | --- | --- |
| High<sup>a</sup> | Onion | Surface Compost | 2.881 | 3.078 | 2.684 |
| High<sup>a</sup> | Onion | Tilled Compost | 2.955 | 3.158 | 2.753 |
| High<sup>a</sup> | Pepper | Surface Compost | 3.751 | 4.008 | 3.494 |
| High<sup>a</sup> | Pepper | Tilled Compost | 3.021 | 3.228 | 2.814 |
| Low<sup>b</sup> | Onion | Surface Compost | 5.487 | 5.863 | 5.112 |
| Low<sup>b</sup> | Onion | Tilled Compost | 5.749 | 6.142 | 5.356 |
| Low<sup>b</sup> | Pepper | Surface Compost | 7.145 | 7.634 | 6.656 |
| Low<sup>b</sup> | Pepper | Tilled Compost | 5.876 | 6.278 | 5.474 |

<sup>a</sup>Calculations using a mean wage rate of 2092 FCFA/day and a median wage rate of 2000 FCFA/day
<sup>b</sup>Calculations using a mean wage rate of 1066 FCFA/day and a median wage rate of 1042 FCFA/day

**Table S47.** Descriptive statistics of initial sheep weight (kg) in Phase I and Phase II.

| Treatment | N | Mean | St. dev. | Min. | Max. |
| --- | --- | --- | --- | --- | --- |
| Phase I<sup>a</sup> |  |  |  |  |  |
| All cornmeal (control) | 3 | 17.7 | 4.9 | 13.5 | 23.0 |
| 15% *C. demersum* | 2 | 18.8 | 4.6 | 15.5 | 22.0 |
| 30% *C. demersum* | 2 | 17.8 | 2.5 | 16.0 | 19.5 |
| 45% *C. demersum* | 3 | 20.3 | 0.3 | 20.0 | 20.5 |
| 60% *C. demersum* | 3 | 21.0 | 5.4 | 16.5 | 27.0 |
| Phase II<sup>b</sup> |  |  |  |  |  |
| All peanut straw (control) | 3 | 18.3 | 0.9 | 17.0 | 20.0 |
| 15% *C. demersum* | 3 | 19.0 | 4.1 | 14.5 | 27.0 |
| 30% *C. demersum* | 3 | 19.8 | 0.2 | 19.5 | 20.0 |
| 45% *C. demersum* | 4 | 20.5 | 0.9 | 19.0 | 23.0 |
| 60% *C. demersum* | 4 | 17.5 | 1.4 | 15.0 | 21.5 |

<sup>a</sup>There were no significant differences among groups ( $F_{1,11} = 1.624$ , $p = 0.229$ ).
<sup>b</sup>There were no significant differences among groups ( $F_{1,15} = 0.47$ , $p = 0.750$ ).

**Table S48.** Descriptive statistics of sheep weight (kg) during all weeks in Phase I and Phase II.

| Treatment | N | Mean | St. dev. | Min. | Max. |
| --- | --- | --- | --- | --- | --- |
| Phase I<sup>a</sup> |  |  |  |  |  |
| All cornmeal (control) | 3 | 22.2 | 6.0 | 13.5 | 33.0 |
| 15% *C. demersum* | 2 | 22.0 | 5.7 | 14.5 | 30.0 |
| 30% *C. demersum* | 2 | 22.8 | 3.0 | 16.0 | 26.5 |
| 45% *C. demersum* | 3 | 21.2 | 1.8 | 17.0 | 24.5 |
| 60% *C. demersum* | 3 | 23.4 | 5.2 | 16.0 | 31.5 |
| Phase II<sup>b</sup> |  |  |  |  |  |
| All peanut straw (control) | 3 | 19.5 | 3.4 | 12.0 | 24.0 |
| 15% *C. demersum* | 3 | 19.9 | 6.6 | 12.0 | 31.0 |
| 30% *C. demersum* | 3 | 22.5 | 1.8 | 18.0 | 26.0 |
| 45% *C. demersum* | 4 | 22.4 | 2.4 | 17.5 | 27.0 |
| 60% *C. demersum* | 4 | 18.6 | 2.6 | 14.5 | 26.0 |

<sup>a</sup>There were no significant differences among groups ( $F_{1,11}=0.438$ , $p=0.509$ ).
<sup>b</sup>There were no significant differences among groups ( $F_{1,15}=0.575$ , $p=0.686$ ).

**Table S49.** Daily and weekly livestock feed costs, cost per calorie of feed types, and calories/kg in feed across the five treatments.

| Treatment | Daily<br>feed cost<br>(USD) | Weekly<br>feed cost<br>(USD) | Cost per<br>calorie<br>(USD) | Calories<br>/kg in<br>feed |
| --- | --- | --- | --- | --- |
| Phase I |  |  |  |  |
| All cornmeal (control) | 0.505 | 3.534 | 0.0144 | 21.000 |
| 15% *C. demersum* | 0.429 | 3.000 | 0.0134 | 19.281 |
| 30% *C. demersum* | 0.353 | 2.474 | 0.0121 | 17.562 |
| 45% *C. demersum* | 0.278 | 1.944 | 0.0105 | 15.843 |
| 60% *C. demersum* | 0.202 | 1.414 | 0.0086 | 14.124 |
| Phase II |  |  |  |  |
| All peanut straw (control) | 0.54 | 3.76 | 0.0269 | 19.98 |
| 15% *C. demersum* | 0.47 | 3.29 | 0.0269 | 17.46 |
| 15% *C. demersum* | 0.40 | 2.81 | 0.0269 | 14.94 |
| 45% *C. demersum* | 0.33 | 2.34 | 0.0270 | 12.42 |
| 60% *C. demersum* | 0.27 | 1.87 | 0.0270 | 9.900 |

**Table S50.** Estimates of the expected ratio of weight gain to feed cost for a kg of livestock feed. High-cost estimates use the high estimate in the price of livestock feed (peanut straw, pellets, corn) while low-cost estimates use the low price of the same livestock feed prices. Prices follow seasonal fluctuations that align with the agricultural season in the region. All prices have been converted into USD. Cornmeal only has one price so there is no difference between high and low cost in Phase I.

| Wage rate | Treatment | High-Cost Weight Gain to Feed Cost (kg / USD) | Low-Cost Weight Gain to Feed Cost (kg / USD) |
| --- | --- | --- | --- |
| Phase I |  |  |  |
| High<sup>a</sup> | Control | 0.013 |  |
| High<sup>a</sup> | 15% Cerato | -0.012 |  |
| High<sup>a</sup> | 30% Cerato | -0.062 |  |
| High<sup>a</sup> | 45% Cerato | -0.048 |  |
| High<sup>a</sup> | 60% Cerato | -0.114 |  |
| Low<sup>b</sup> | Control | 0.013 |  |
| Low<sup>b</sup> | 15% Cerato | -0.012 |  |
| Low<sup>b</sup> | 30% Cerato | -0.062 |  |
| Low<sup>b</sup> | 45% Cerato | -0.049 |  |
| Low<sup>b</sup> | 60% Cerato | -0.116 |  |
| Phase II |  |  |  |
| High<sup>a</sup> | Control | 0.072 | 0.096 |
| High<sup>a</sup> | 15% Cerato | 0.041 | 0.055 |
| High<sup>a</sup> | 30% Cerato | 0.100 | 0.132 |
| High<sup>a</sup> | 45% Cerato | 0.062 | 0.081 |
| High<sup>a</sup> | 60% Cerato | 0.093 | 0.120 |
| Low<sup>b</sup> | Control | 0.072 | 0.096 |
| Low<sup>b</sup> | 15% Cerato | 0.041 | 0.055 |
| Low<sup>b</sup> | 30% Cerato | 0.100 | 0.133 |
| Low<sup>b</sup> | 45% Cerato | 0.062 | 0.082 |
| Low<sup>b</sup> | 60% Cerato | 0.093 | 0.122 |

<sup>a</sup>Calculations using a mean wage rate of 2092 FCFA/day
<sup>b</sup>Calculations using a mean wage rate of 1066 FCFA/day

**Table S51.**
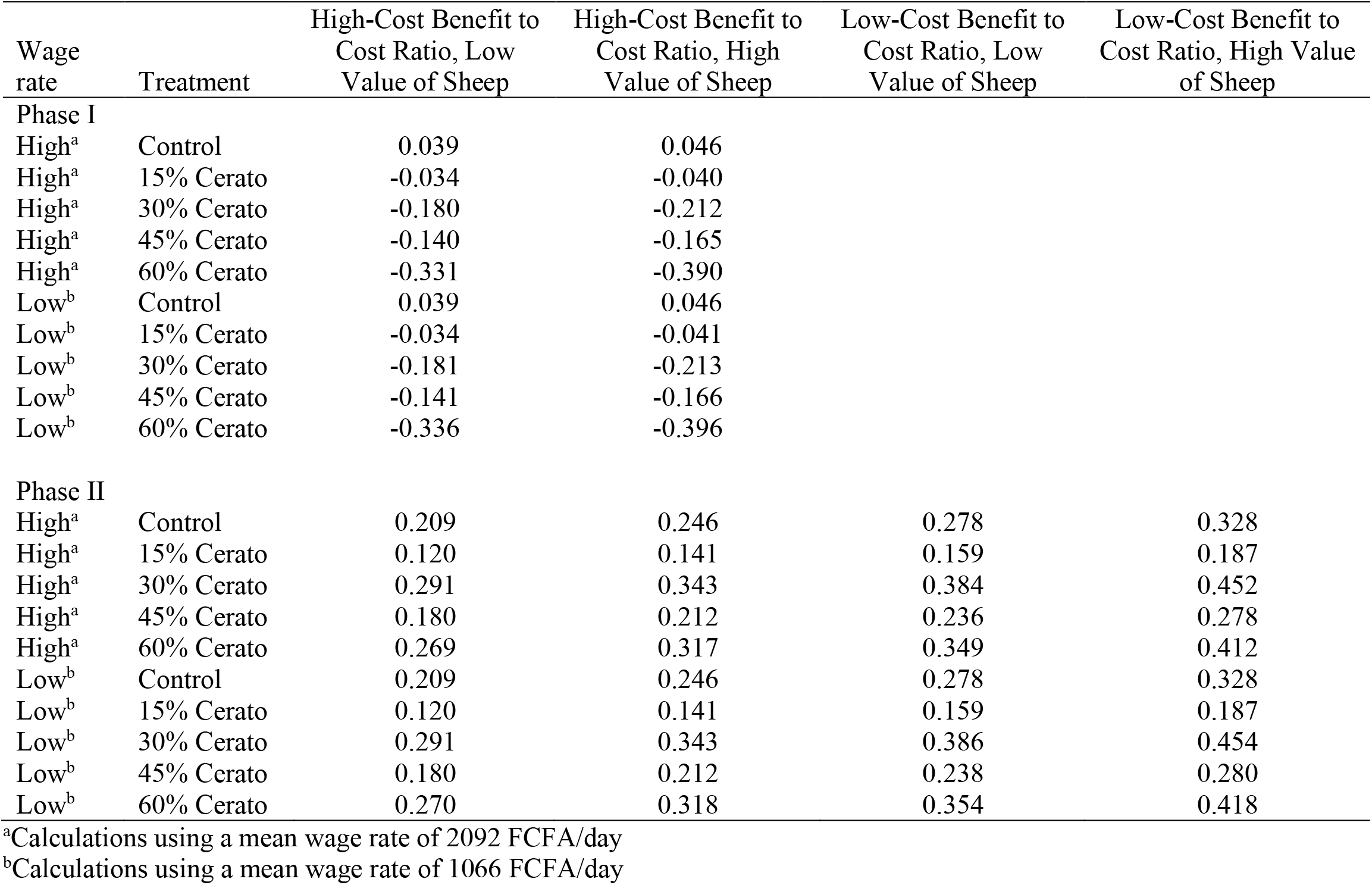
Estimates of the expected ratio of marginal revenue product (MRP, including only private gains) to marginal cost (MC) of a kg of livestock feed. The high value of a sheep is calculated by dividing 55000 FCFA by 28 kg and converting to USD while the low value of a sheep uses 50000 FCFA and 30 kg. High-cost estimates use the high estimate in the price of livestock feed (peanut straw, pellets, corn) while low-cost estimates use the low price of the same livestock feed prices. Prices follow seasonal fluctuations that align with the agricultural season in the region. All prices have been converted into USD. Corn only has one price so there is no difference between high and low cost in Phase I.

**Table S52.** Estimates of the expected ratio of marginal revenue product (MRP, including private gains and public health benefit) to marginal cost (MC) of a metric ton of compost. Mean estimates are produced using the mean estimated marketing margin between Saint Louis and Dakar and the mean public health benefit. Upper MRP estimates use the mean plus (minus) one standard deviation and the high estimate of the public health benefit. All prices have been converted into USD.

| Wage rate | Crop | Treatment | Mean MRP/MC<br>(metric ton<br>compost) | Upper<br>MRP/MC<br>(metric<br>ton<br>compost) | Lower<br>MRP/MC<br>(metric<br>ton<br>compost) |
| --- | --- | --- | --- | --- | --- |
| High<sup>a</sup> | Onion | Surface Compost | 3.439 | 3.705 | 3.174 |
| High<sup>a</sup> | Onion | Tilled Compost | 3.405 | 3.662 | 3.147 |
| High<sup>a</sup> | Pepper | Surface Compost | 4.310 | 4.635 | 3.984 |
| High<sup>a</sup> | Pepper | Tilled Compost | 3.470 | 3.732 | 3.208 |
| Low<sup>b</sup> | Onion | Surface Compost | 6.551 | 7.057 | 6.045 |
| Low<sup>b</sup> | Onion | Tilled Compost | 6.623 | 7.123 | 6.122 |
| Low<sup>b</sup> | Pepper | Surface Compost | 8.209 | 8.828 | 7.589 |
| Low<sup>b</sup> | Pepper | Tilled Compost | 6.750 | 7.259 | 6.241 |

<sup>a</sup>Calculations using a mean wage rate of 2092 FCFA/day and a median wage rate of 2000 FCFA/day
<sup>b</sup>Calculations using a mean wage rate of 1066 FCFA/day and a median wage rate of 1042 FCFA/day

**Table S53.** Estimates of the expected ratio of marginal revenue product (MRP, including private gains and public health benefits) to marginal cost (MC) of a kg of livestock feed. The high value of a sheep is calculated by dividing 55000 FCFA by 28 kg and converting to USD while the low value of a sheep uses 50000 FCFA and 30 kg. High-cost estimates use the high estimate in the price of livestock feed (peanut straw, pellets, corn) while low-cost estimates use the low price of the same livestock feed prices. Prices follow seasonal fluctuations that align with the agricultural season in the region. All prices have been converted into USD. Corn only has one price so there is no difference between high and low cost in Phase I.

| Wage rate | Treatment | High-Cost Benefit to<br>Cost Ratio, Low<br>Value of Sheep | High-Cost Benefit to<br>Cost Ratio, High<br>Value of Sheep | Low-Cost Benefit to<br>Cost Ratio, Low<br>Value of Sheep | Low-Cost Benefit to<br>Cost Ratio, High Value<br>of Sheep |
| --- | --- | --- | --- | --- | --- |
| Phase I |  |  |  |  |  |
| High<sup>a</sup> | Control | 0.039 | 0.046 |  |  |
| High<sup>a</sup> | 15% Cerato | -0.034 | -0.040 |  |  |
| High<sup>a</sup> | 30% Cerato | -0.162 | -0.194 |  |  |
| High<sup>a</sup> | 45% Cerato | -0.105 | -0.130 |  |  |
| High<sup>a</sup> | 60% Cerato | -0.325 | -0.384 |  |  |
| Low<sup>b</sup> | Control | 0.039 | 0.046 |  |  |
| Low<sup>b</sup> | 15% Cerato | -0.034 | -0.040 |  |  |
| Low<sup>b</sup> | 30% Cerato | -0.162 | -0.195 |  |  |
| Low<sup>b</sup> | 45% Cerato | -0.106 | -0.131 |  |  |
| Low<sup>b</sup> | 60% Cerato | -0.330 | -0.390 |  |  |
| Phase II |  |  |  |  |  |
| High<sup>a</sup> | Control | 0.209 | 0.246 | 0.278 | 0.328 |
| High<sup>a</sup> | 15% Cerato | 0.122 | 0.143 | 0.161 | 0.189 |
| High<sup>a</sup> | 30% Cerato | 0.295 | 0.347 | 0.389 | 0.458 |
| High<sup>a</sup> | 45% Cerato | 0.187 | 0.220 | 0.246 | 0.288 |
| High<sup>a</sup> | 60% Cerato | 0.282 | 0.330 | 0.366 | 0.428 |
| Low<sup>b</sup> | Control | 0.209 | 0.246 | 0.278 | 0.328 |
| Low<sup>b</sup> | 15% Cerato | 0.122 | 0.143 | 0.161 | 0.190 |
| Low<sup>b</sup> | 30% Cerato | 0.295 | 0.347 | 0.391 | 0.460 |
| Low<sup>b</sup> | 45% Cerato | 0.188 | 0.220 | 0.248 | 0.290 |
| Low<sup>b</sup> | 60% Cerato | 0.283 | 0.331 | 0.371 | 0.435 |

<sup>a</sup>Calculations using a mean wage rate of 2092 FCFA/day
<sup>b</sup>Calculations using a mean wage rate of 1066 FCFA/day

**Table S54.**
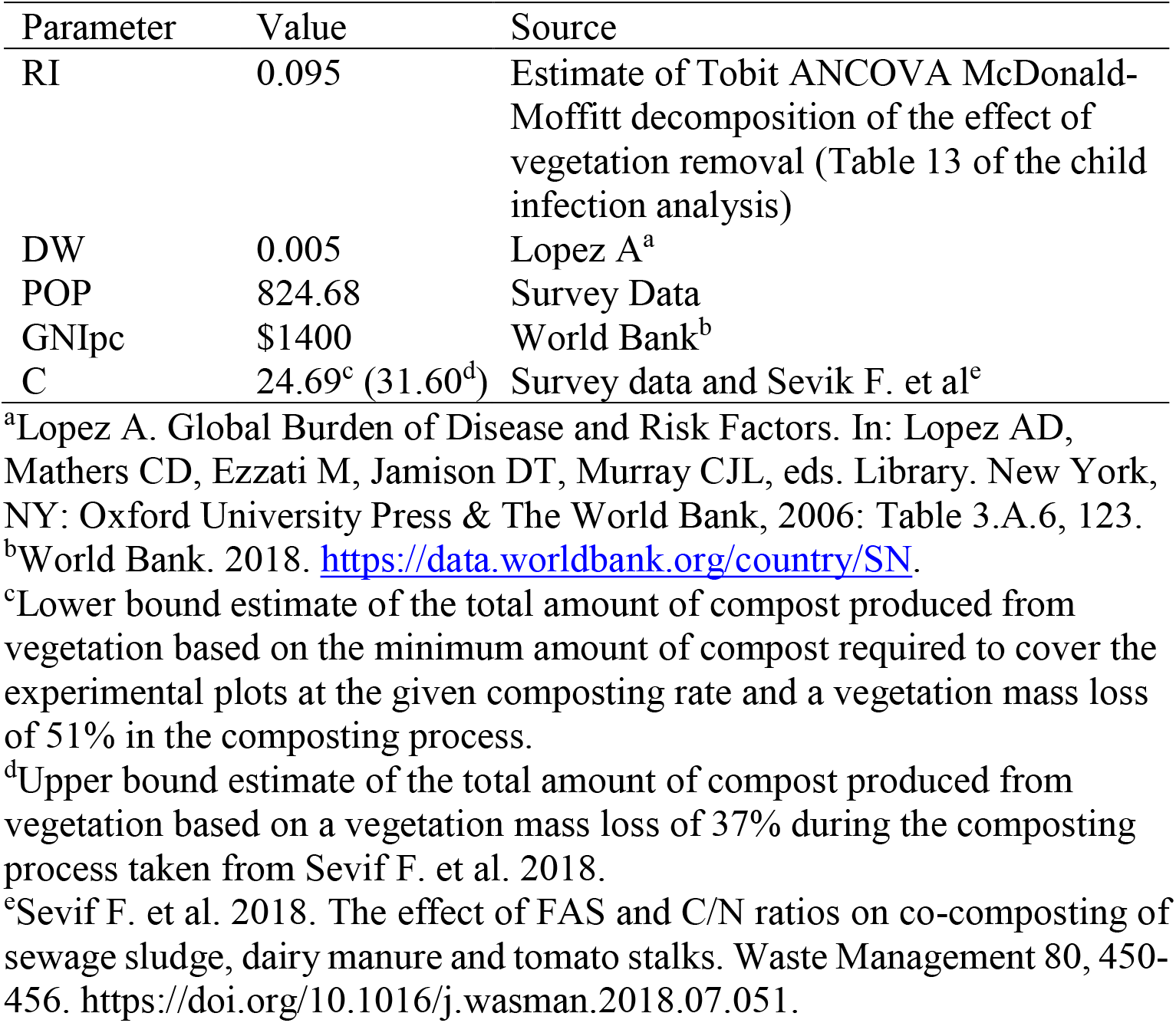
Parameters and data sources used in public health benefit calculation.

**Table S55.** Results of the global mixed effects binomial regression predicting child-level *S. haematobium* prevalence described in SI Appendix Text S4. The child-level global model began with the same predictors as our path model but also controlled for predictors that were identified as important by^15^ in their dataset, including water access area per site (m^2^) and individual-level effects of child gender and age. Our hypothesized pathways to human infection that were significant in the final path model remained significant in the child-level regression after controlling for the above predictors (Table S18).

| Fixed effects | Estimate | Std. error | z-value | p-value |
| --- | --- | --- | --- | --- |
| (Intercept) | 0.93448 | 0.28217 | 3.312 | <0.001 |
| Scale(accessarea) | 0.13716 | 0.48049 | 0.285 | 0.775 |
| Child gender | 0.36463 | 0.12023 | 3.033 | 0.002 |
| Class | -0.02713 | 0.06609 | -0.411 | 0.681 |
| Scale(population) | -0.42953 | 0.21303 | -2.016 | 0.044 |
| Scale(vegetation area) | 0.57334 | 0.38288 | 1.497 | 0.134 |
| Scale(sqkmtotalcrop0.5km) | 1.01248 | 0.35747 | 2.832 | 0.005 |
| Scale(log-totalsnails) | 0.48730 | 0.21432 | 2.274 | 0.023 |

**Table S56.** Results of the final mixed effects binomial regression predicting child-level *S. haematobium* prevalence after model selection.

| Fixed effects | Estimate | Std. error | z-value | p-value |
| --- | --- | --- | --- | --- |
| (Intercept) | 0.8748 | 0.2453 | 3.566 | <0.001 |
| Child gender | 0.3619 | 0.1199 | 3.019 | 0.003 |
| Scale(population) | -0.4471 | 0.1992 | -2.245 | 0.025 |
| Scale(vegetation area) | 0.6506 | 0.2788 | 2.334 | 0.020 |
| Scale(sqkmtotalcrop0.5km) | 1.0448 | 0.3349 | 3.120 | 0.002 |
| Scale(log-totalsnails) | 0.4811 | 0.2134 | 2.255 | 0.024 |

**Table S57.**
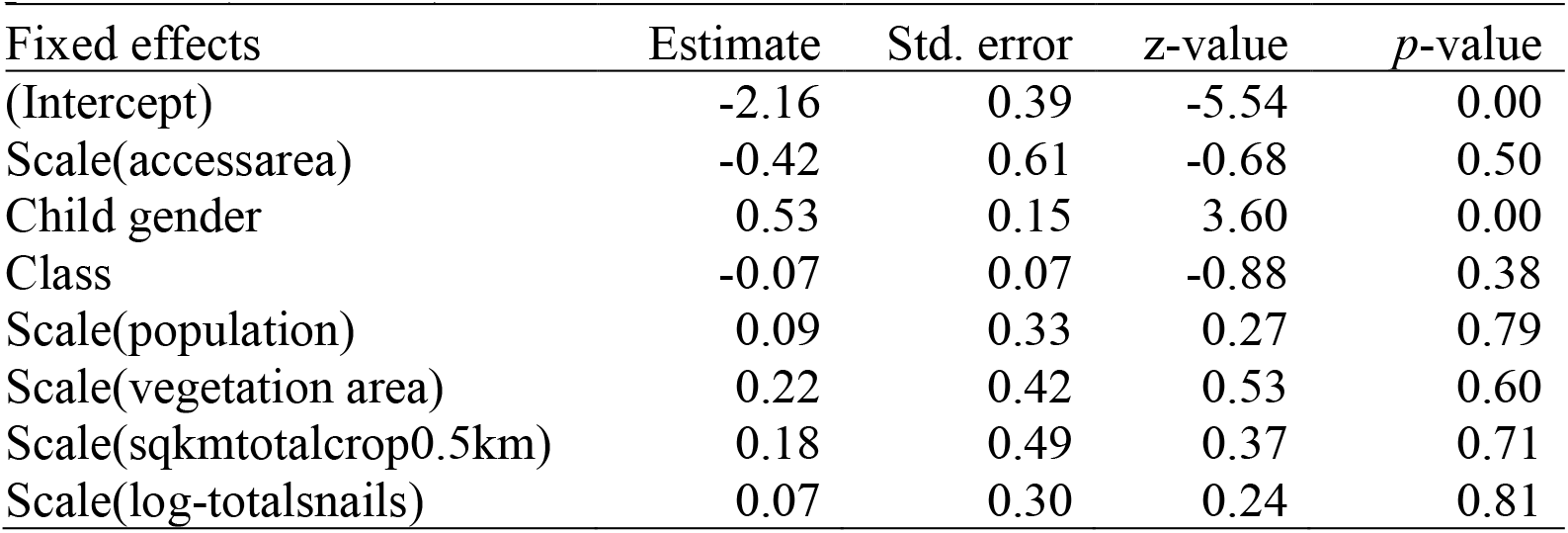
Results of the global mixed effects binomial regression predicting child-level *S. mansoni* prevalence. The child-level global model began with the same predictors as our path model but also controlled for predictors that were identified as important by^15^ in their dataset, including water access area per site (m^2^) and individual-level effects of child gender and age. Our hypothesized pathways to human infection that were significant in the final path model remained significant in the child-level regression after controlling for the above predictors (Table S18).

**Table S58.** Results of the final mixed effects binomial regression predicting child-level *S. mansoni* prevalence after model selection.

| Fixed effects | Estimate | Std. error | z-value | *p*-value |
| --- | --- | --- | --- | --- |
| (Intercept) | -2.17 | 0.32 | -6.68 | <0.001 |
| Child gender | 0.40 | 0.13 | 2.98 | 0.003 |

**Table S59.** Results of linear regression of ln-transformed yield (kg/hectare) of onions or peppers. Ln-transformed yield is regressed on indicator variables for the five crop trial treatment arms (surface compost, tilled compost, urea, surface compost and urea, and tilled compost and urea) and an additional control for the site of the trial. Table S52 shows the mean marginal physical product effects by treatment. The control treatment is no compost and no urea. The pepper trials were conducted in two sites with different plot sizes. We also include a dummy variable for the site of Mbane to control for plot size and other site-specific but treatment-independent unobservables. Thus, the control group mean is for no compost and no urea in Lampsar. Treatments that share letters have estimated coefficients that are not significantly different from one another (*p*>0.05) based on a Wald tests of equity.

| Effect | Estimate | Std. Error | t-value | p-value | Wald test of equity |
| --- | --- | --- | --- | --- | --- |
| Onions ($N=18$, adjusted $R$-squared of 0.858) | | | | | |
| Intercept (control group mean) | 7.252 | 0.229 | 31.65 | 0.000 | a |
| Surface Compost | 2.077 | 0.324 | 6.41 | 0.000 | b |
| Tilled Compost | 2.293 | 0.324 | 7.08 | 0.000 | b |
| Urea | 0.320 | 0.324 | 0.99 | 0.342 | c |
| Surface Compost x Urea | -0.347 | 0.458 | -0.76 | 0.463 | c |
| Tilled Compost x Urea | -0.235 | 0.458 | -0.51 | 0.617 | c |
| Peppers ($N=36$, adjusted $R$-squared of 0.621) | | | | | |
| Intercept (control group mean) | 6.232 | 0.448 | 13.90 | 0.000 | a |
| Surface Compost | 1.264 | 0.587 | 2.15 | 0.040 | b |
| Tilled Compost | 1.265 | 0.587 | 2.16 | 0.040 | b |
| Urea | -0.280 | 0.587 | -0.48 | 0.637 | c |
| Surface Compost x Urea | 0.375 | 0.830 | 0.45 | 0.655 | bc |
| Tilled Compost x Urea | 0.719 | 0.830 | 0.87 | 0.393 | bc |

**Table S58.**
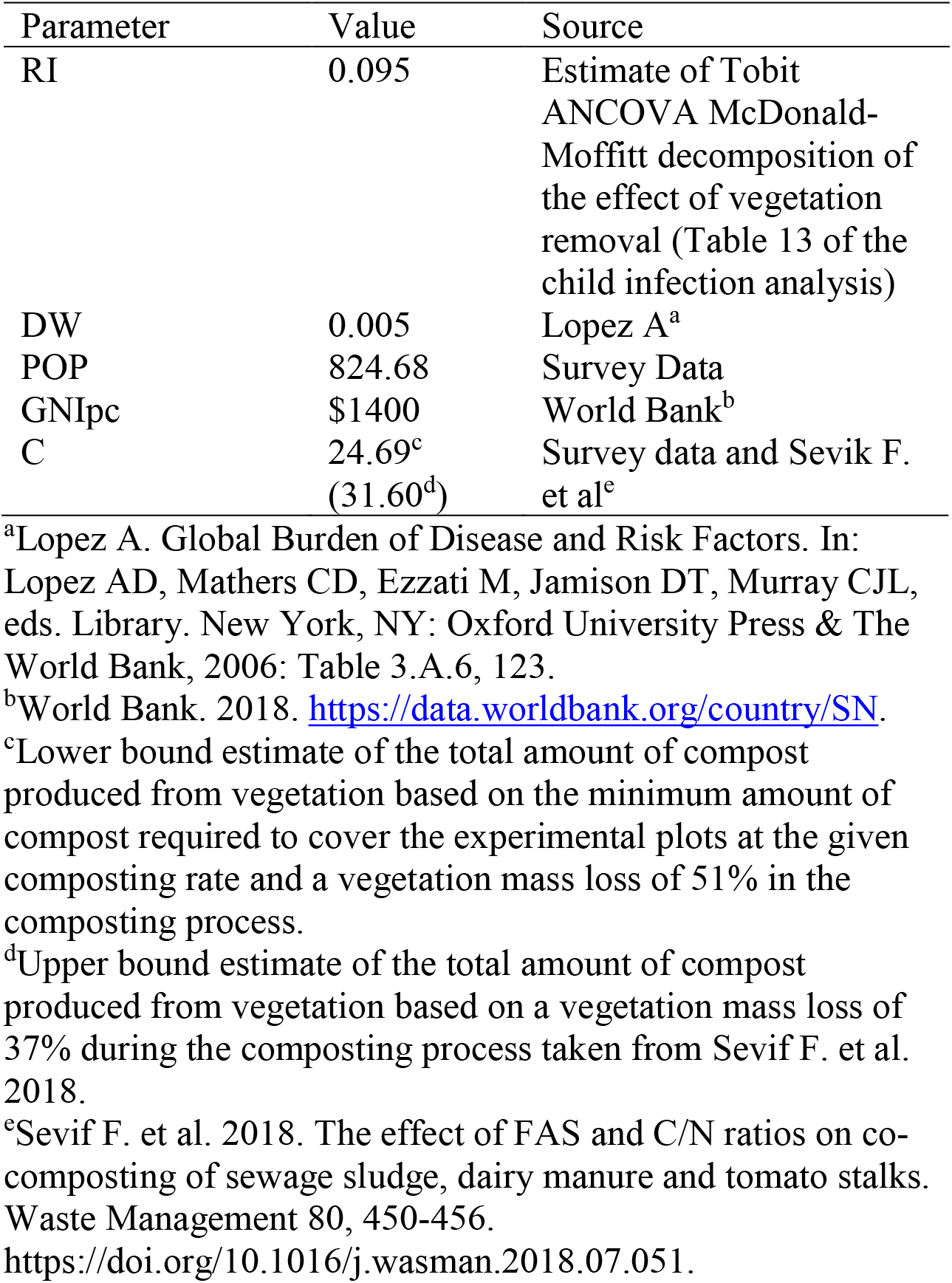
Parameters and data sources used in public health benefit calculation.

## Supplemental Figures

**Fig. S1.**
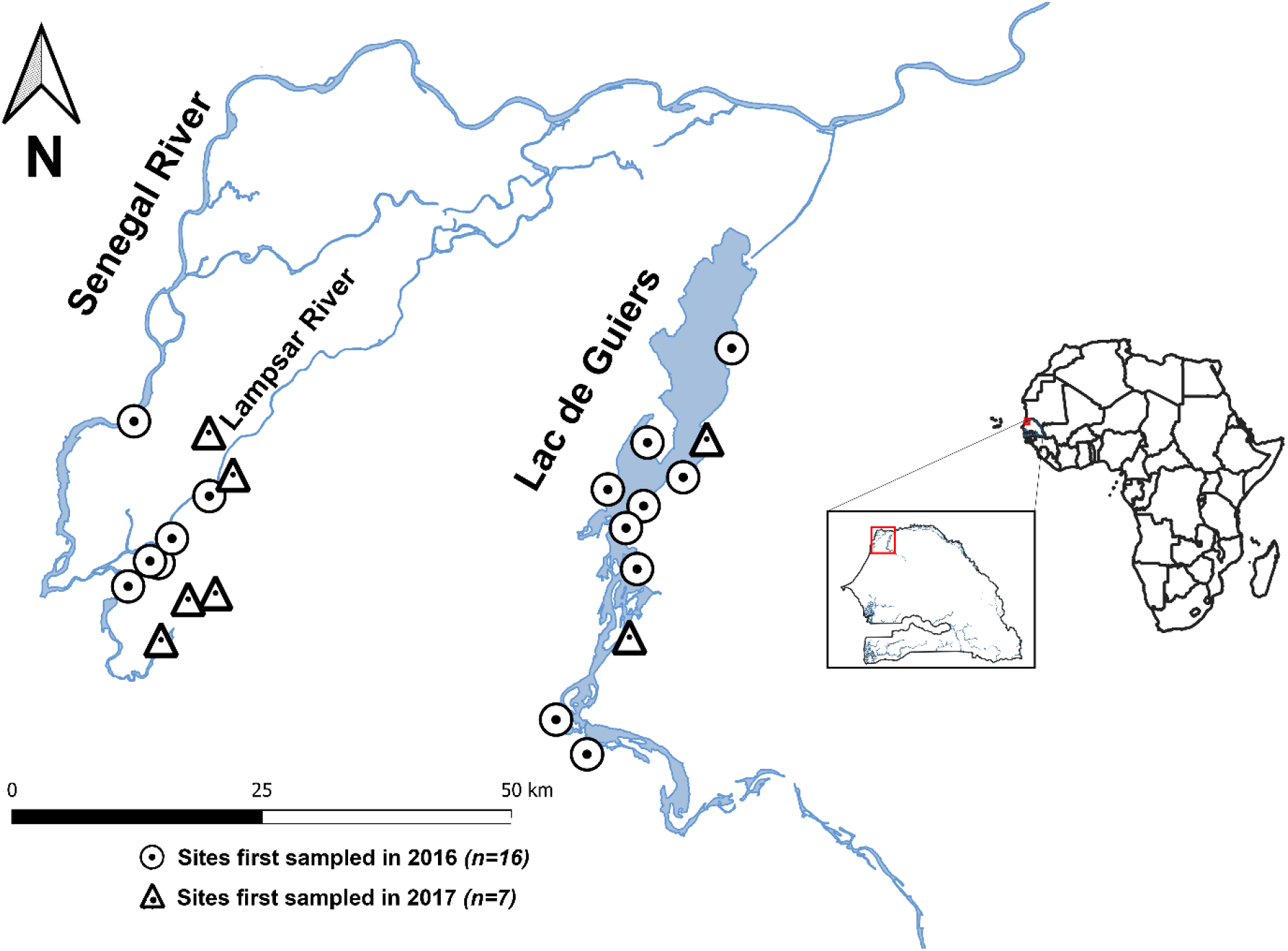
Map of 23 sampled sites located along the lower Senegal River (left site cluster) and Lac de Guiers regions (right site cluster) of northern Senegal. All site point locations are displayed on a single map enlarged from red polygon of the inset map on the right showing the country of Senegal within Africa. Circular symbols denote sites first sampled for baseline infection in 2016 and tested for infection post-treatment in both 2017 and 2018 (*n=16*), while triangles denote sites sampled only for baseline infection in 2017 (*n=7*).

**Fig. S2.**
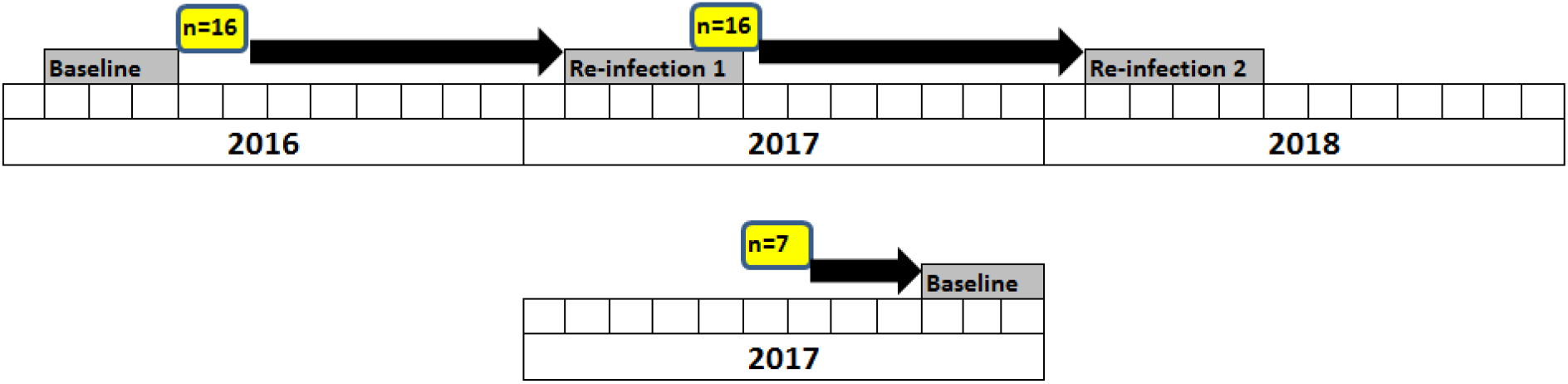
Human infection (gray) and snail/vegetation sampling (yellow) dates (each box is a month of the year), with black arrows showing which snail data was used to predict each human infection round in the path model of human infection. Text above black arrows describes how data was excluded to limit manipulation impacts upon our findings. Only baseline data for all sites (i.e. before any manipulations) and non-manipulated controls (i.e. no vegetation removed) were used to test hypotheses in the agricultural land use study. The top row is the sampling regime for the 16 sites first sampled in 2016 (circle symbols in Fig. S1) and the bottom row is the sampling regime for the 7 sites first sampled in 2017 (triangle symbols in Fig. S1).

**Fig. S3.**
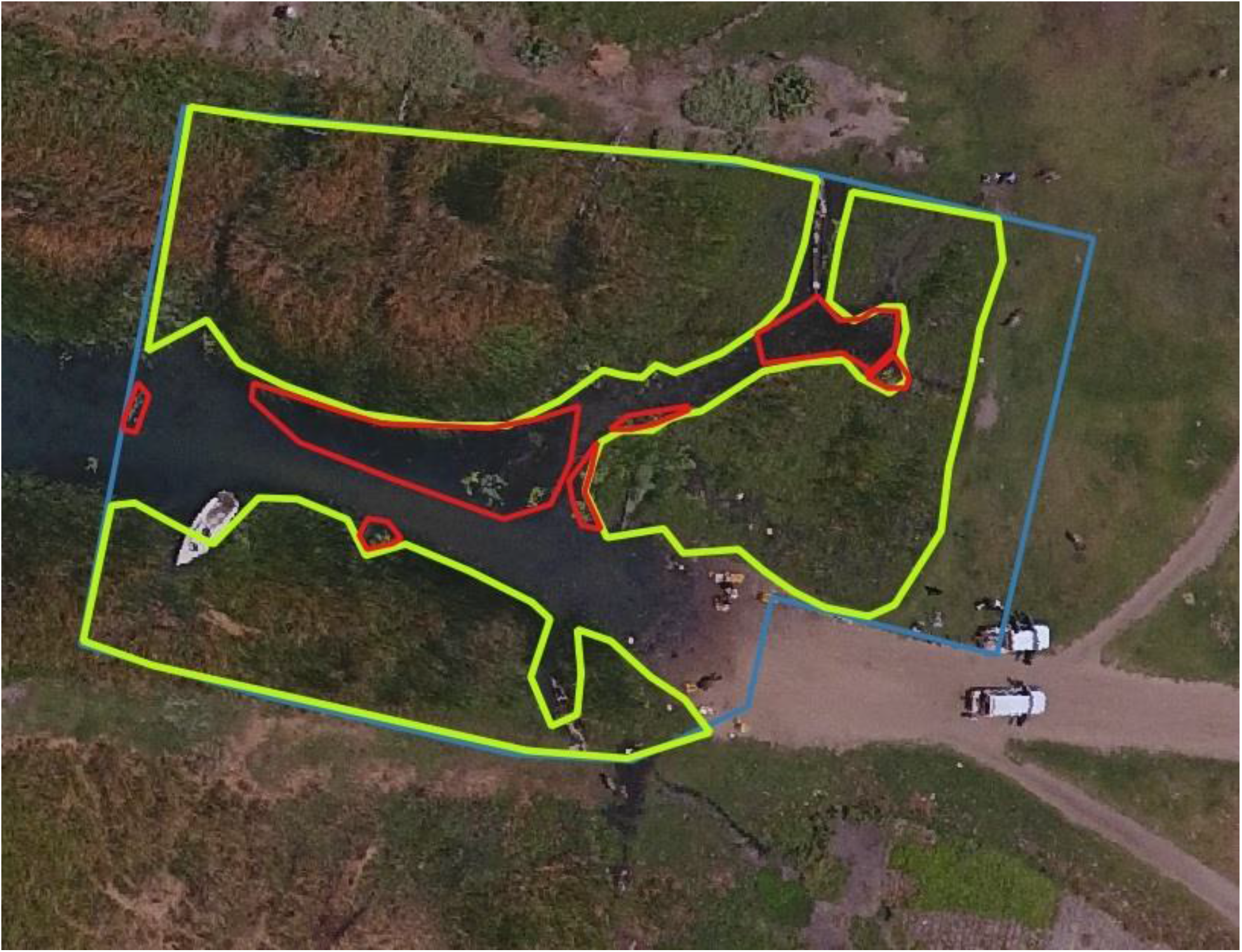
Water access point 1 of 2 at the site of Diokhor showing QGIS 3.2 workflow to estimate area (m^2^) of the water access point (blue), emergent vegetation (yellow), and non-emergent aquatic vegetation (red).

**Fig. S4.**
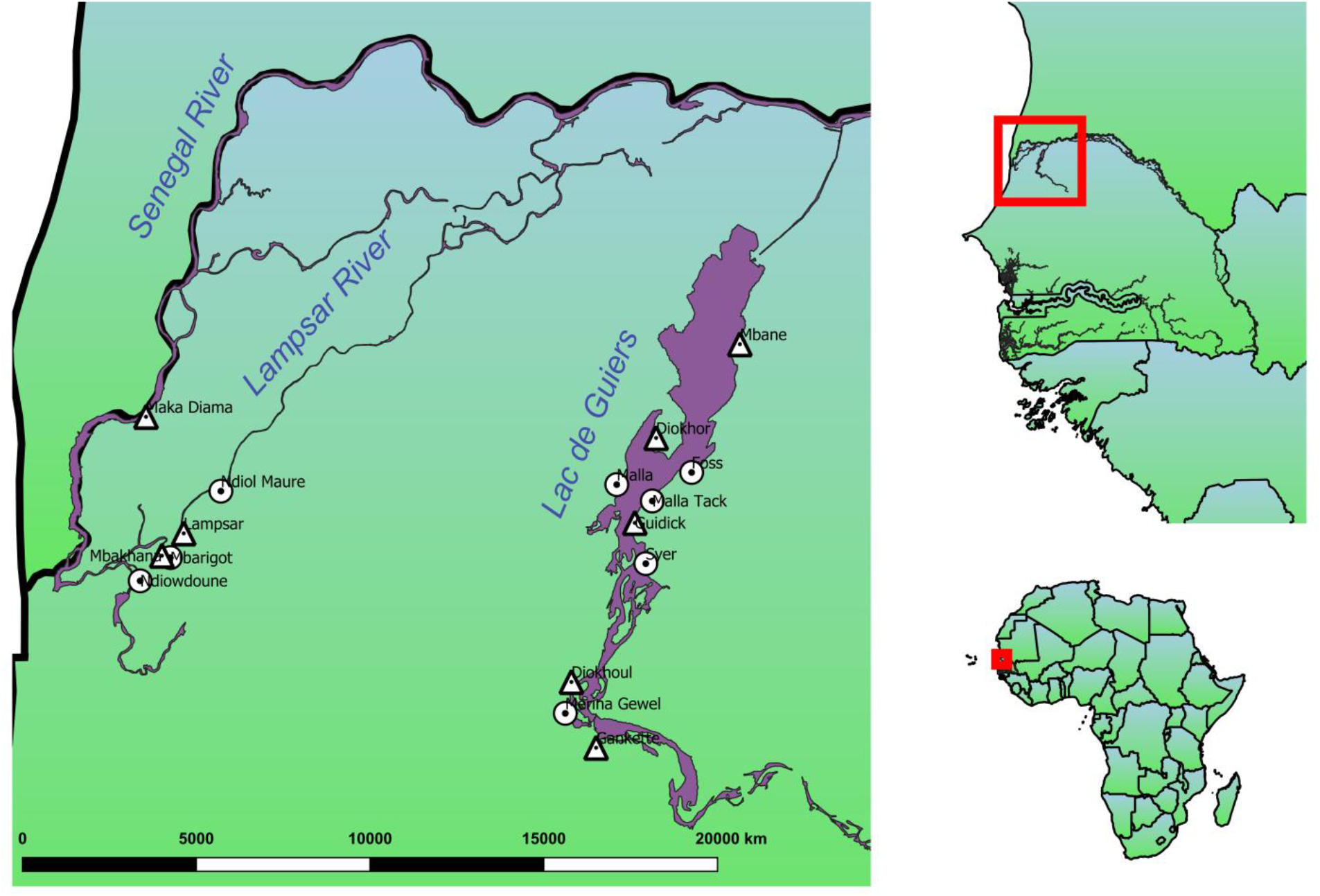
Map of 16 sampled sites located along the lower Senegal and Lampsar Rivers (left site cluster) and Lac de Guiers regions (right site cluster) of northern Senegal, Africa. All site point locations are displayed on a single map enlarged from red polygon of the inset map on the right showing the country of Senegal within Africa. Circular and triangular symbols denote control and manipulation sites, respectively.

**Fig. S5.**
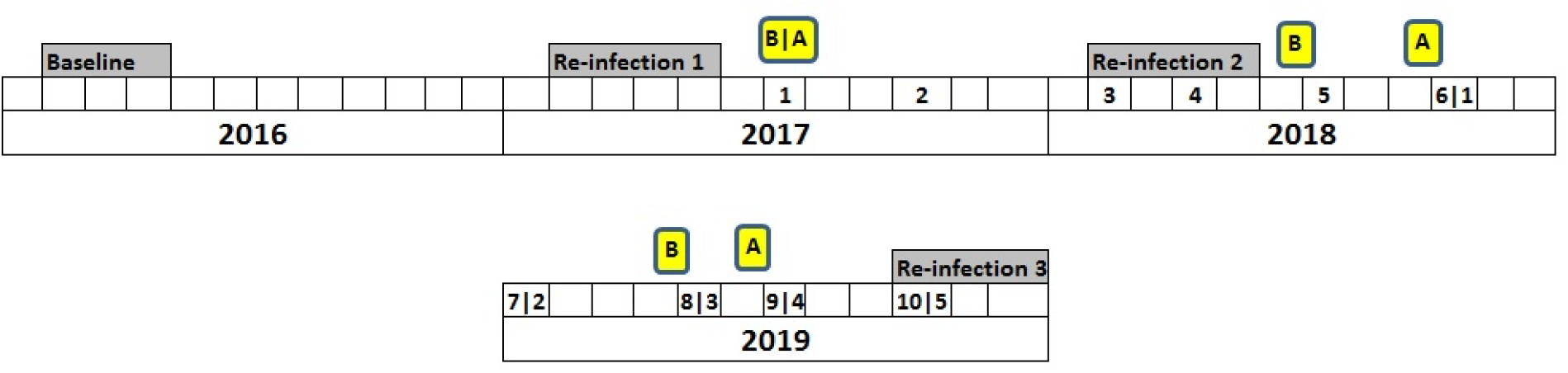
Timing of human infection testing (gray), snail sampling (yellow), and vegetation removal rounds per year (numeric values 1-10). For each year of vegetation removal, we performed snail sampling both before (B) and after (A) a vegetation removal round. We used the symbol “|” to divide removal rounds by sites started in 2017 (left of “|”) from those that started in 2018 (right of “|”).

**Fig. S6.**
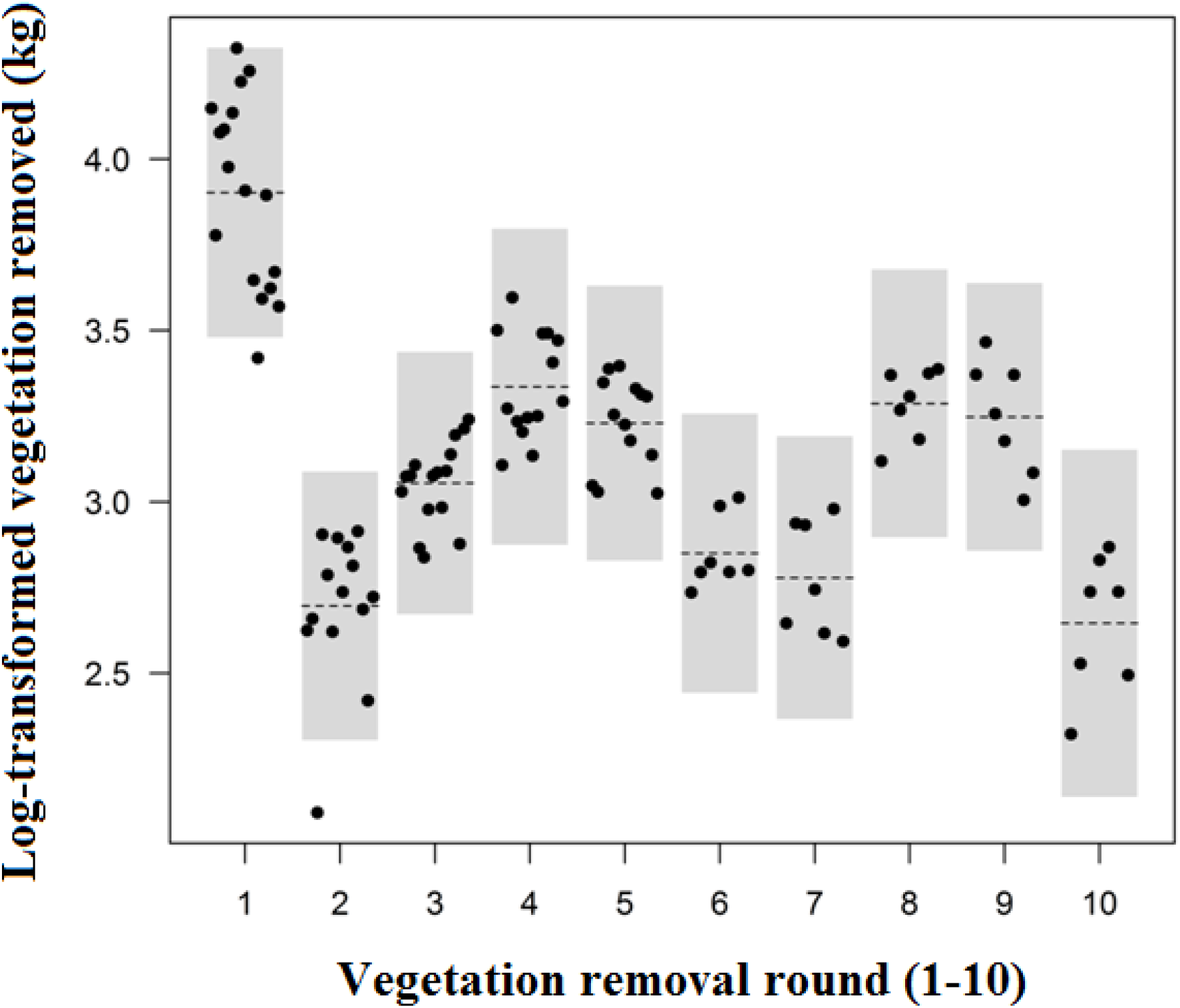
Log-transformed estimated quantity of vegetation removed (kg) for each removal round (1-10) during the study.

**Fig. S7.**
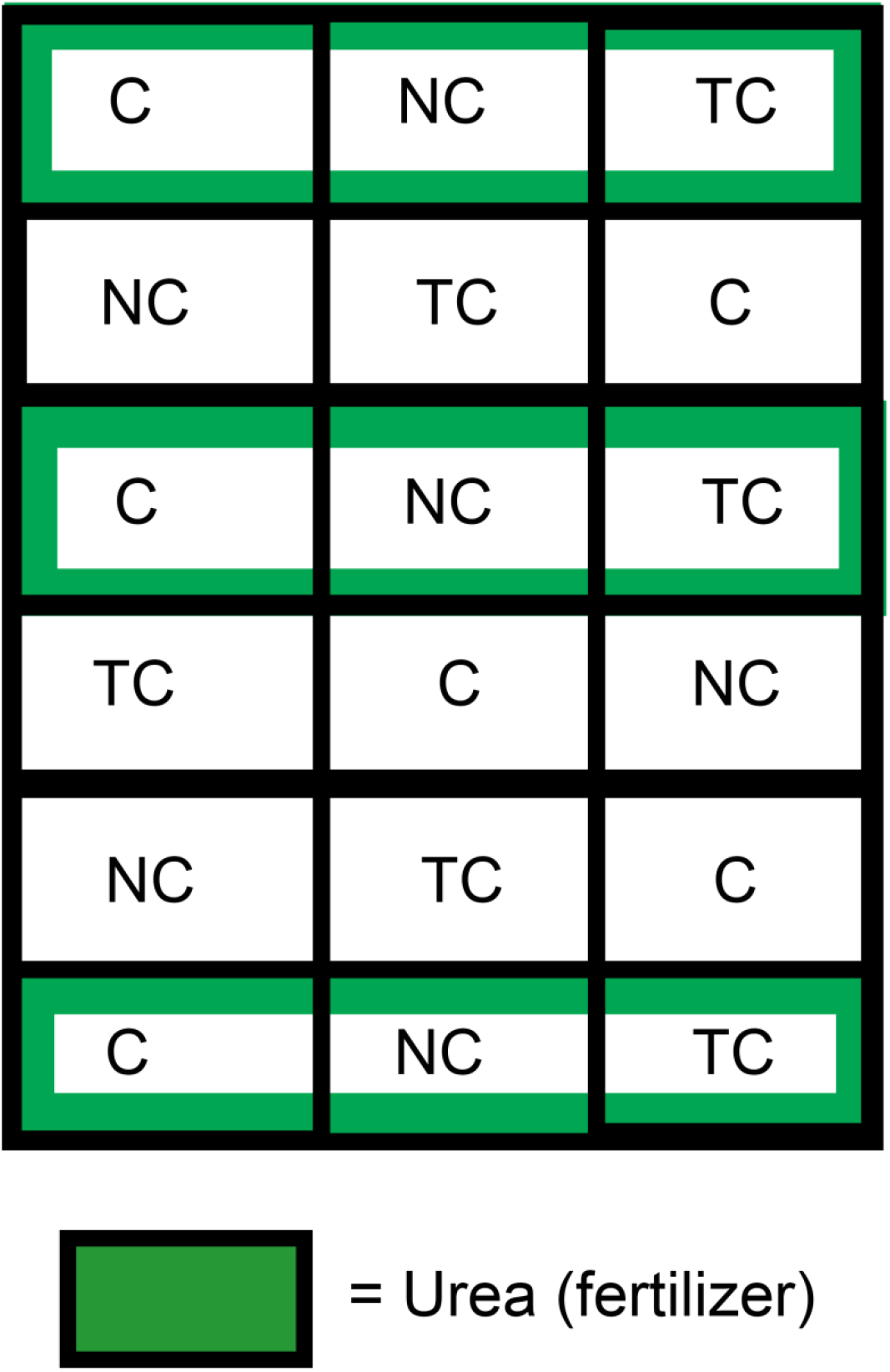
Split-plot experiment field layout showing the random assignment of whole-plot (Green = urea or fertilizer) and split-plot treatments (C= compost, TC = tilled compost, NC = no compost). Seedlings were planted at equal densities in all plots and compost was applied at standardized quantities at both C and TC plots.

**Figure S8.**
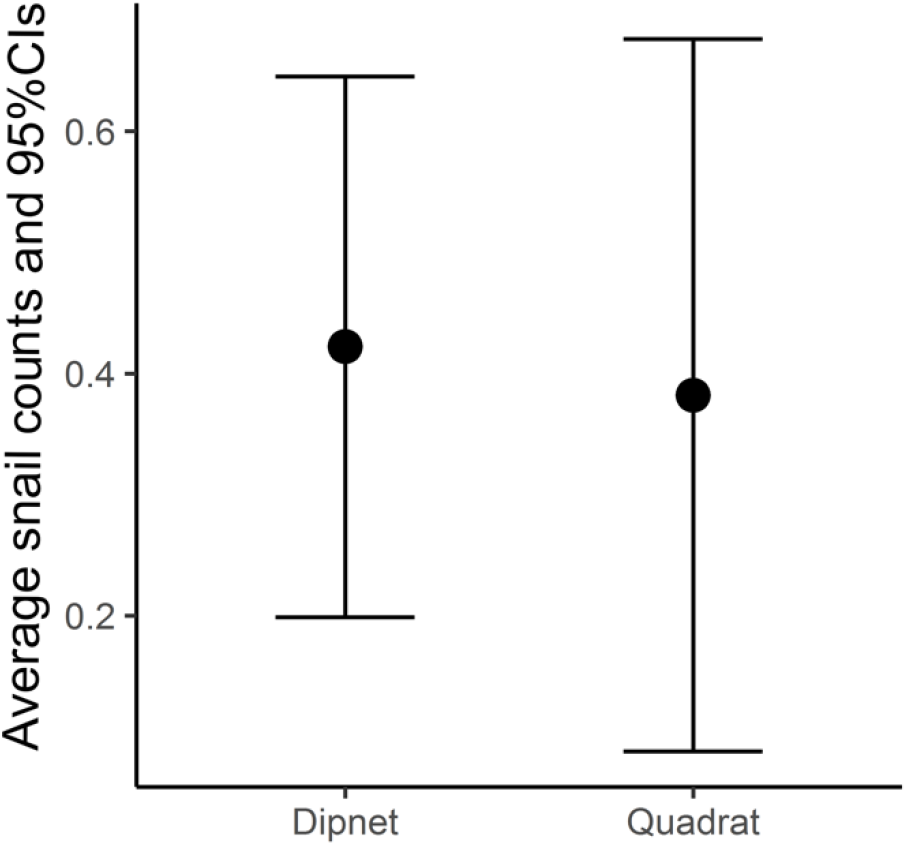
Average *B. truncatus/globosus* counts and 95% CIs found using side-by-side sampling with both dipnet and quadrat methods.

**Fig. S9.**
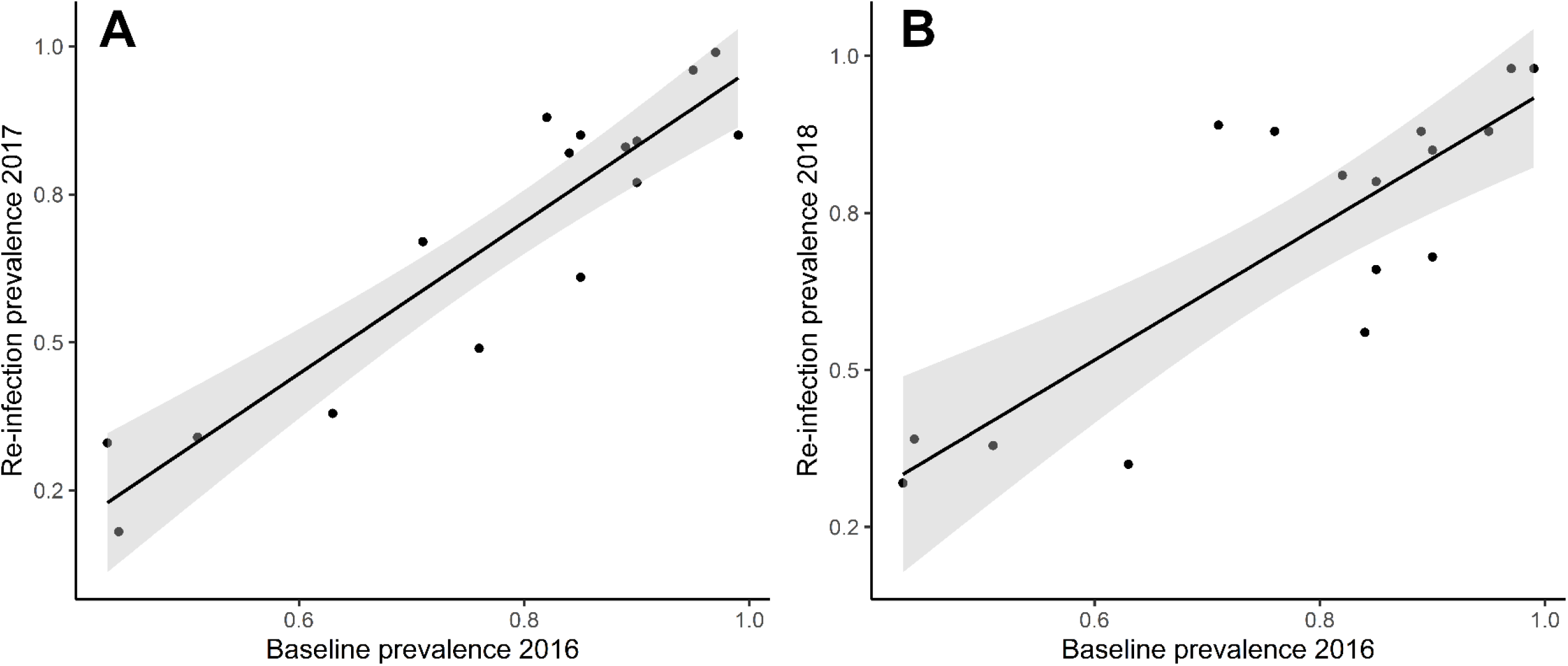
Bi-variate scatterplots of human baseline prevalence at 16 sites sampled in 2016 versus both infection post-treatment in 2017 **(A)** and in 2018 **(B)**.

**Fig. S10.**
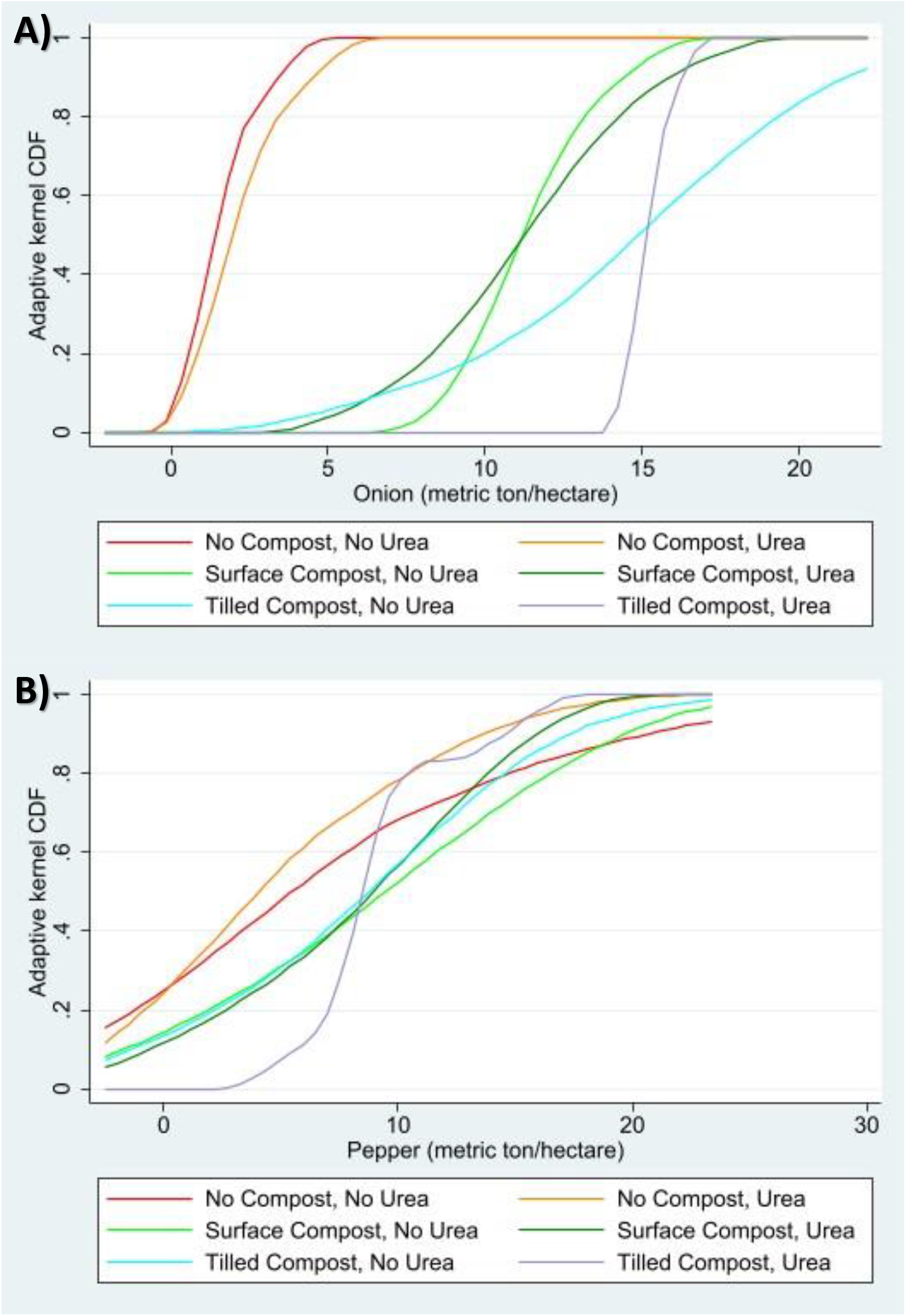
Kernel density plot of **A)** onion and **B)** pepper yield in metric ton/hectare by treatment arm.

**Fig. S11.**
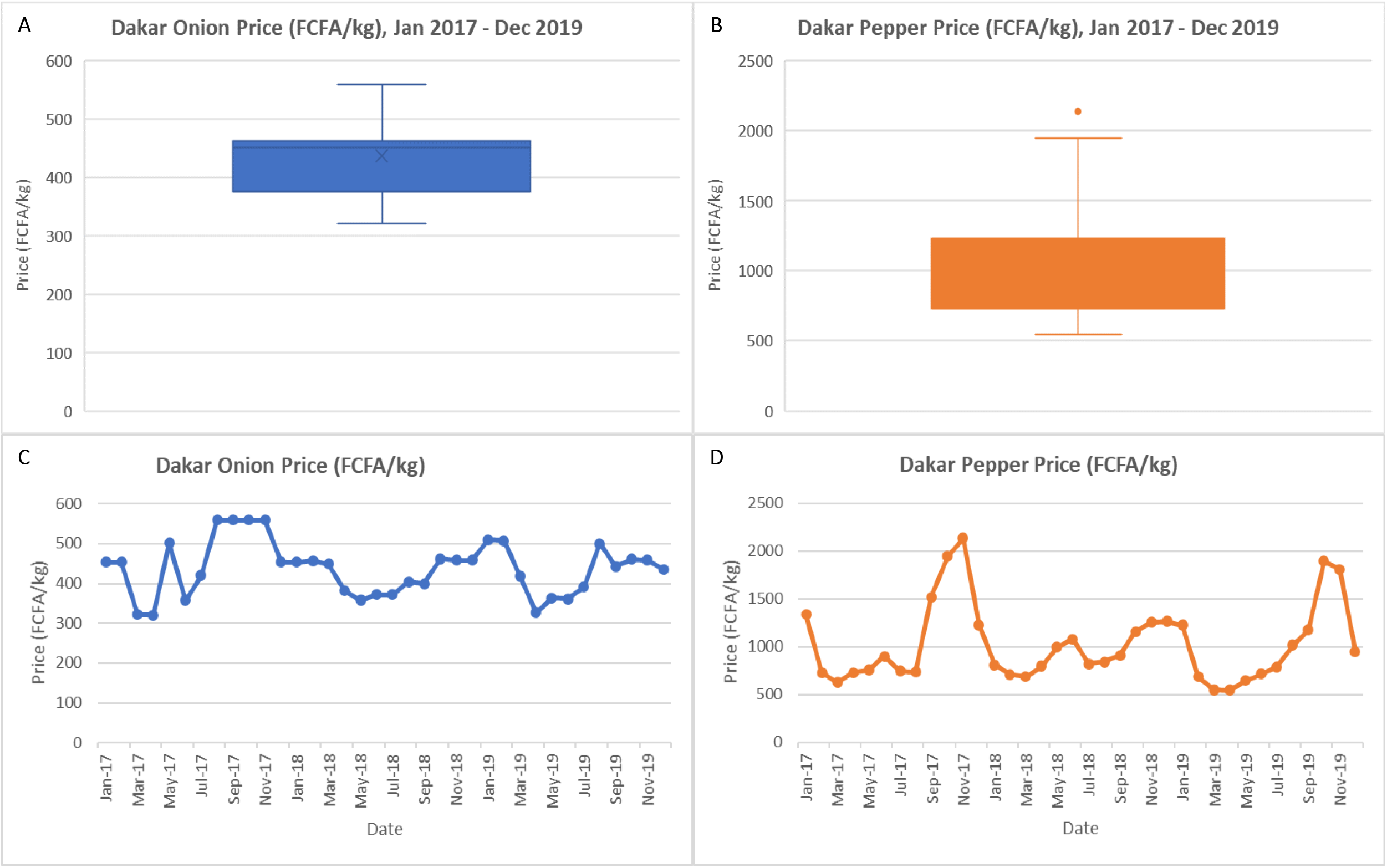
Distribution of mean **A)** onion and **B)** price (FCFA/kg) and time series of mean **C)** onion and **D)** pepper price (FCFA/kg) in Dakar reported monthly from January 2017 – December 2019 (Data Source: Agence National de la Statisitque et de la Demographie).

**Fig. S12.**
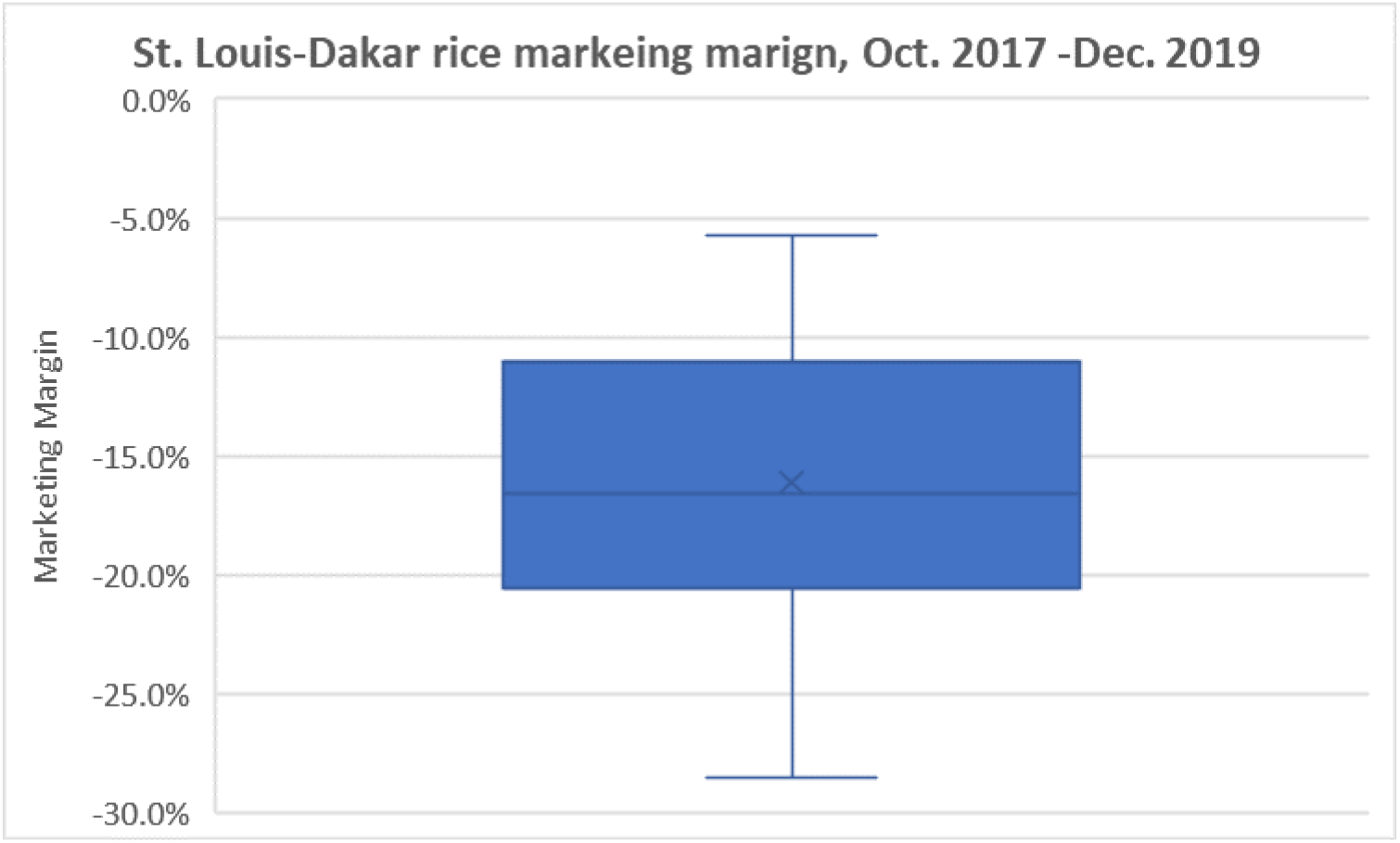
Distribution of the marketing margin between Saint Louis and Dakar (Saint Louis price – Dakar Price) using local rice prices from October 2017 – December 2019.

**Fig. S13.**
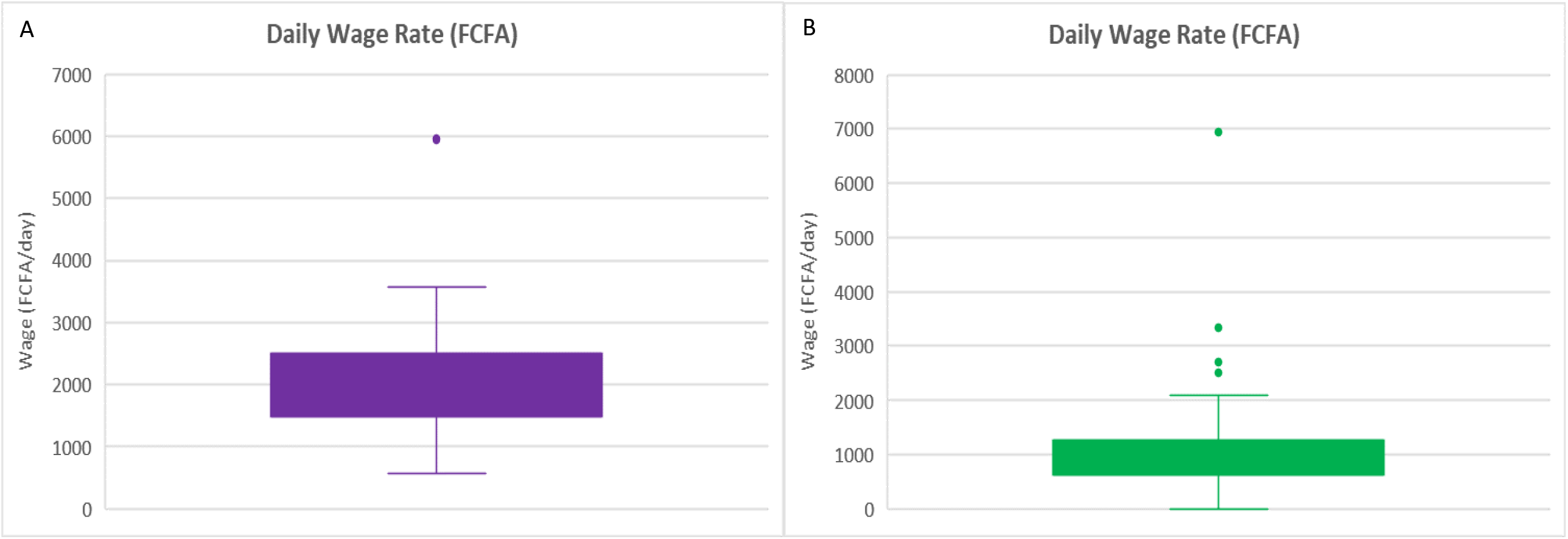
Distribution of the daily wage rate for male agricultural workers using **A)** a 2018 survey of 27 horticulture workers in seven sites in the Senegal Delta region and **B)** a 2017-18 survey of 266 horticulture workers in the Saint Louis region.

## Footnotes

1 The logarithmic specification fits the data better than a linear specification. The resulting coefficient estimates can be interpreted roughly as percentage changes due to that treatment relative to the control group (omitted).

2 Pepper crop trials were done in two villages, Lampsar and Mbane. A village binary indicator variable is included to control for plot size differences across villages and other site-specific (but treatment-independent) unobserved variation.

3 Kolmogorov-Smirnov tests were used for first-order stochastic dominance tests, with p-values < 0.050 for all of them. Second order and third order stochastic dominance tests use the *domsvy* program for Stata written by David Stifel (https://sites.lafayette.edu/stifeld/misc/stata-tests-of-stochastic-dominance/). All reported stochastic dominance results hold at the 95% confidence level for second-order stochastic dominance.

4 https://www.ansd.sn/index.php?option=com_ansd&view=titrepublication&id=9&Itemid=287. Each report contains the current and previous three months’ price, and the price from a year ago. We complied the time series from 24 separate reports covering this period.

5 For fresh horticultural products, storage or preservation are unlikely options, so the contemporaneous spatial marketing margin (i.e., price difference) is the relevant adjustment.

6 The noon buying rates in New York City for cable transfers payable in foreign currencies was used. Board of Governors of the Federal Reserve System (US), U.S. / Euro Foreign Exchange Rate [DEXUSEU], retrieved from FRED, Federal Reserve Bank of St. Louis; https://fred.stlouisfed.org/series/DEXUSEU, June 12, 2020.

7 A person day is eight hours. Reported vegetation harvest time is from the survey data, other time estimates come from local project management staff.

8 The labor time required for vegetation removal was calculated using the mean and the median vegetation removal rate for the survey data. The total amount of vegetation required to make the compost was calculated by taking the amount of compost required to cover the plots based on the reported composting rates (3400 kg of compost) and then adjusting for vegetation loss during the composting process (36 – 51% mass loss). Based on the required 5450 to 6973 kg of vegetation, the mean labor time required is 12.81-person days while the median labor time required is 12.42 person days.

9 One might consider the cost of digging the pit a fixed cost. We treat it as a variable cost for three reasons: (i) the size of the pit and thus the cost of creating it varies with the volume of compost created; (ii) even if reused for subsequent rounds of compost production, there will be some required maintenance effort (e.g., to remove erosive fill); and (iii) we prefer to err on the conservative side, so that, if anything, our estimates understate the returns to compost production and application from the harvested aquatic vegetation.

10 All workers were male. We therefore value daily labor per male rural wage rates from survey data from the region kindly provided by Prof. Miet Maertens (KU Leuven).

11 All workers were male. Data were graciously made available by the Bureau d’analyses macro-économiques (BAME) of the Institute sénégalais de recherches agricoles (ISRA).

12 Fixed costs were calculated by first converting all prices into USD. Three 12-teeth rakes, three round rakes, and one shovel were used throughout the vegetation removal and composting process. The costs of these six rakes and one shovel were added to either the high-end or low-end cost of waders and gloves to produce the high-end and low-end fixed cost estimates.

13 Since peanut straw and pellets are agricultural products, their prices exhibit significant seasonality. We use both the high and low prices for benefit cost calculation to look at how the benefit cost ratio changes throughout the year.

