## Supplementary material for "A planetary health solution for disease, sustainability, food, water, and poverty challenges": Veg Removal RMarkdown

### Vegetation removal ms Rmarkdown

Chris and Lexi

7/28/22

#This code for the vegetation removal ms #Analyses will include before/after comparisons of snails and vegetation… #And similar analyses for human infection

```
#Packages:
library(car)
```

```
## Loading required package: carData
```

```
library(nlme)
library(lme4)
```

```
## Loading required package: Matrix
```

```
##
## Attaching package: 'lme4'
```

```
## The following object is masked from 'package:nlme':
##
##     lmList
```

```
library(visreg)
library(MASS)
library(MuMIn)
library(lattice)
library(rmarkdown)
```

```
## Warning: package 'rmarkdown' was built under R version 4.1.2
```

```
library(ggplot2)
library(sjPlot)
```

```
## #refugeeswelcome
```

```
library(cowplot)
```

```
##
## Attaching package: 'cowplot'
```

```
## The following objects are masked from 'package:sjPlot':
##
##     plot_grid, save_plot
```

```
library(plyr)
library(glmmTMB)
```

```
## Warning in checkDepPackageVersion(dep_pkg = "TMB"): Package version inconsistency detected.
## glmmTMB was built with TMB version 1.7.21
## Current TMB version is 1.7.22
## Please re-install glmmTMB from source or restore original 'TMB' package (see '?reinstalling' for more
```

```
library(lsmeans)
```

```
## Loading required package: emmeans
```

```
## The 'lsmeans' package is now basically a front end for 'emmeans'.
## Users are encouraged to switch the rest of the way.
## See help('transition') for more information, including how to
## convert old 'lsmeans' objects and scripts to work with 'emmeans'.
```

```
library(effects)
```

```
## Warning: package 'effects' was built under R version 4.1.2
```

```
## Use the command
##     lattice::trellis.par.set(effectsTheme())
##   to customize lattice options for effects plots.
## See ?effectsTheme for details.
```

```
library(PerformanceAnalytics)
```

```
## Loading required package: xts
```

```
## Loading required package: zoo
```

```
##
## Attaching package: 'zoo'
```

```
## The following objects are masked from 'package:base':
##
##     as.Date, as.Date.numeric
```

```
##
## Attaching package: 'PerformanceAnalytics'
```

```
## The following object is masked from 'package:graphics':
##
##     legend
```

```
library(ordinal)
```

```
##
## Attaching package: 'ordinal'

## The following objects are masked from 'package:glmmTMB':
##
##     ranef, VarCorr

## The following objects are masked from 'package:lme4':
##
##     ranef, VarCorr

## The following objects are masked from 'package:nlme':
##
##     ranef, VarCorr
```

```
library(VGAM)
```

```
## Loading required package: stats4

## Loading required package: splines

##
## Attaching package: 'VGAM'

## The following objects are masked from 'package:ordinal':
##
##     dgumbel, dlgamma, pgumbel, plgamma, qgumbel, rgumbel, wine

## The following object is masked from 'package:glmmTMB':
##
##     betabinomial

## The following object is masked from 'package:MuMIn':
##
##     AICc

## The following object is masked from 'package:car':
##
##     logit
```

```
library(tidyverse)
```

```
## -- Attaching packages --------------------------------------- tidyverse 1.3.1 --

## v tibble  3.1.6     v dplyr   1.0.7
## v tidyr   1.1.4     v stringr 1.4.0
## v readr   2.1.2     v forcats 0.5.1
## v purrr   0.3.4
```

```
## Warning: package 'readr' was built under R version 4.1.2

## -- Conflicts ------------------------------------------ tidyverse_conflicts() --
## x dplyr::arrange()   masks plyr::arrange()
## x dplyr::collapse()  masks nlme::collapse()
## x purrr::compact()   masks plyr::compact()
## x dplyr::count()     masks plyr::count()
## x tidyr::expand()    masks Matrix::expand()
## x dplyr::failwith()  masks plyr::failwith()
## x tidyr::fill()      masks VGAM::fill()
## x dplyr::filter()    masks stats::filter()
## x dplyr::first()     masks xts::first()
## x dplyr::id()        masks plyr::id()
## x dplyr::lag()       masks stats::lag()
## x dplyr::last()      masks xts::last()
## x dplyr::mutate()    masks plyr::mutate()
## x tidyr::pack()      masks Matrix::pack()
## x dplyr::recode()    masks car::recode()
## x dplyr::rename()    masks plyr::rename()
## x dplyr::select()    masks MASS::select()
## x dplyr::slice()     masks ordinal::slice()
## x purrr::some()      masks car::some()
## x dplyr::summarise() masks plyr::summarise()
## x dplyr::summarize() masks plyr::summarize()
## x tidyr::unpack()    masks Matrix::unpack()
```

```
library(GLMMadaptive)
```

```
## Warning: package 'GLMMadaptive' was built under R version 4.1.2

##
## Attaching package: 'GLMMadaptive'

## The following objects are masked from 'package:VGAM':
##
##     coef, confint, fitted, formula, model.frame, model.matrix, nobs,
##     predict, residuals, terms

## The following objects are masked from 'package:stats4':
##
##     coef, confint, nobs

## The following object is masked from 'package:ordinal':
##
##     ranef

## The following object is masked from 'package:MASS':
##
##     negative.binomial

## The following object is masked from 'package:lme4':
##
##     negative.binomial
```

```r
library(emmeans)
```

```r
#####################################################
########  VEGETATION CHANGE    ####################
#####################################################
#Plot vegetation mass from first before/after incident each village:
#veg and snail totals per access-level:
baciveg<-read.csv("vegbaci1.csv")

names(baciveg)
```

```
##  [1] "village"           "site"              "Time"
##  [4] "emergent"          "floating"          "areasite"
##  [7] "propfloatingcover" "totalveg"          "totalbulinus"
## [10] "totalbiom"
```

```r
levels(baciveg$Time)
```

```
## NULL
```

```r
baciveg2=baciveg[baciveg$Time == "After" | baciveg$Time == "Before", ]
baciveg2$Time <- baciveg2$Time[ , drop=TRUE]
reportv3 =ddply(baciveg2,c("Time"),summarize,
               N=length(totalveg),
               avg=mean(totalveg,na.rm=TRUE),
               sd=sd(totalveg,na.rm=TRUE),
               se=sd/sqrt(N),
               ucl=avg+1.98*se,
               lcl=avg-1.98*se)
reportv3$Time2 <- factor(reportv3$Time,levels = c("Before", "After"))
#plot of total veg before/after removal with 95%CI:
profileplotv3 <- ggplot(data=reportv3, aes(x=Time2, y=avg))+
  ggtitle("")+ ylab("Aquatic vegetation mass/sweep")+ xlab("")+
  geom_point(position=position_dodge(w=0.1), size=2)+ geom_line(position=position_dodge(w=1), size=1)+
  geom_errorbar(aes(ymax=ucl, ymin=lcl), width=0.4, position=position_dodge(w=0.1))+
  theme_bw() +
  theme(axis.line = element_line(colour = "black"),
        panel.grid.major = element_blank(),
        panel.grid.minor = element_blank(),
        panel.border = element_blank(),
        panel.background = element_blank()) +theme(axis.text=element_text(size=10,colour = "black"),
                                                   axis.title=element_text(size=10, colour = "black"),)


#total veg mass sig lower after removal at 8 villages
profileplotv3
```

```
## geom_path: Each group consists of only one observation. Do you need to adjust
## the group aesthetic?
## geom_path: Each group consists of only one observation. Do you need to adjust
## the group aesthetic?
```

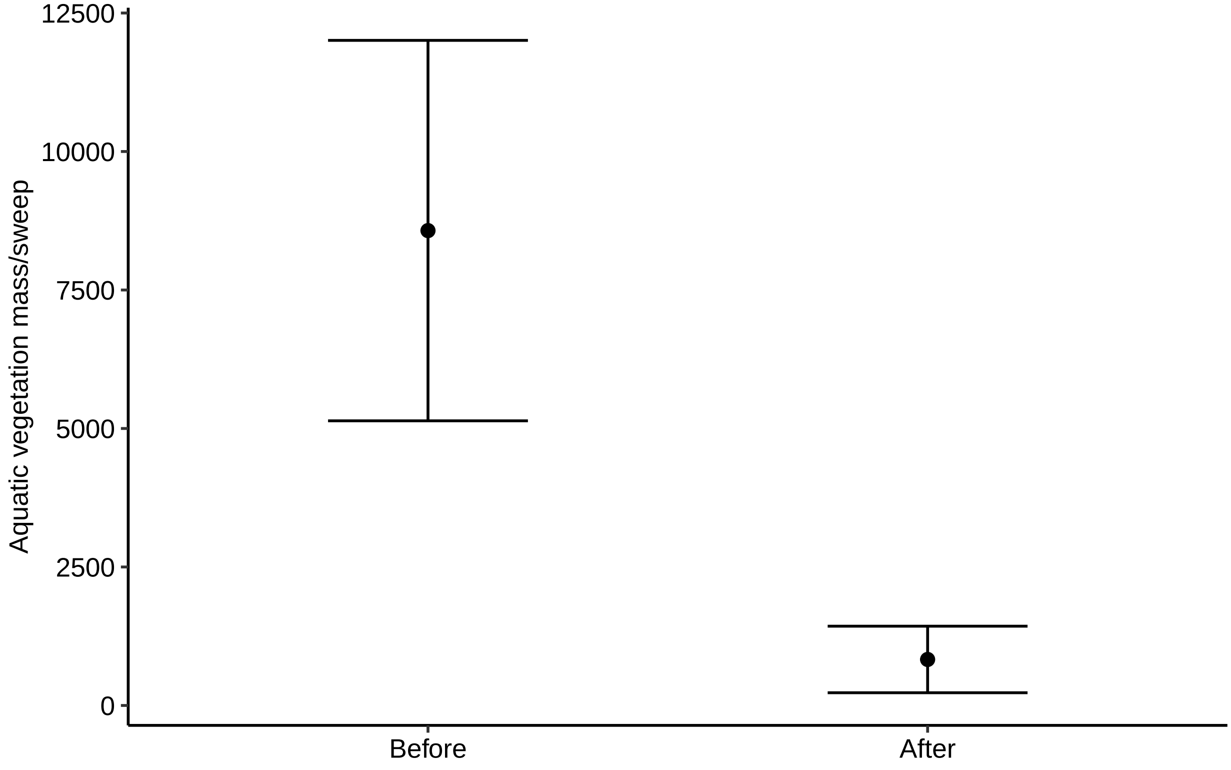


```
#Figure 2A
#plot of all aquatic veg kg removed vs. loss in aquatic veg area m2  after removal:
#This is the data of vegetation cover before/after from all time points from when
#drone imagery is available:
bacikg<-read.csv("Table_vegetation.csv")
bacikg$Kg.removed<-as.numeric(bacikg$Kg.removed)
bacikg<-subset(bacikg,is.na(bacikg$Kg.removed)==FALSE)
bacikg$log.kg.
```

```
##   [1] 4.743141 3.427648 4.209193 4.166874 3.778079 4.205448 4.829818 4.121429
##   [9] 3.582291 4.275519 3.990161 2.768638 3.466719 3.831742 2.905256 3.662191
##  [17] 3.230193 2.757396 2.759668 1.518514 3.682326 3.219585 2.382017 3.426349
##  [25] 2.752048 3.404492 3.081707 3.513484 2.665581 1.785330 3.216957 3.524396
##  [33] 3.206556 3.151370 3.025715 2.702431 2.922725 2.658011 3.003891 2.639486
##  [41] 3.229938 3.206826 3.491081 3.808953 3.113943 3.253338 3.854367 3.950657
##  [49] 2.971276 2.753583 3.728516 2.989895 2.751279 2.959041 2.657056 2.492760
##  [57] 3.502837 3.697229 3.649627 3.278982 3.374565 3.075547 2.515874 3.480151
##  [65] 3.494433 3.061452 3.458940 3.650890 3.134496 3.723948 3.201670 3.522053
##  [73] 2.496930 2.643453 2.868056 2.917506 2.788875 3.227630 3.381296 3.544936
##  [81] 2.622214 2.778151 3.262925 3.072250 2.849419 3.125156 3.561221 2.311754
##  [89] 3.245759 3.188647 3.766562 3.439648 3.145507 3.324282 3.476252 3.761326
##  [97] 3.412629 3.389698 3.178689 3.358886 2.805501 3.467756 2.454845 2.779596
## [105] 2.990339 3.293141 3.877487 3.390759 2.409933 3.381296
```

```r
RoundVegSum<- bacikg %>%
  group_by(Removal.round) %>%
  summarise(
    n=n(),
    mean=mean(Kg.removed),
    median=median(Kg.removed),
    sd=sd(Kg.removed)
  ) %>%
  mutate(se = sd / sqrt(n),
         lower.95ci = mean - qt(1 - (0.05 / 2), n - 1) * se,
         upper.95ci = mean + qt(1 - (0.05 / 2), n - 1) * se)
```

```
## Warning in qt(1 - (0.05/2), n - 1): NaNs produced

## Warning in qt(1 - (0.05/2), n - 1): NaNs produced
```

```r
RoundVegSum
```

```
## # A tibble: 11 x 8
##    Removal.round     n   mean median     sd    se lower.95ci upper.95ci
##            <int> <int>  <dbl>  <dbl>  <dbl> <dbl>      <dbl>      <dbl>
##  1             1    17 14213.   6788 18868. 4576.      4512.     23914.
##  2             2    14  1450.    891  1417.  379.       632.      2268.
##  3             3    16  2110.   1513  2011.  503.      1039.      3182.
##  4             4    14  2564.   1439  2542.  679.      1096.      4031.
##  5             5    13  2192.   1591  1617.  448.      1215.      3169.
##  6             6     7  1457.    827  1144.  433.       399.      2516.
##  7             7     7  1357.   1181  1140.  431.       303.      2411.
##  8             8     7  2629.   2110  1538.  581.      1207.      4051.
##  9             9     7  2597.   2453  1599.  604.      1119.      4076.
## 10            10     7  2012.    978  2579.  975.      -373.      4397.
## 11            NA     1  2406    2406    NA    NA        NaN        NaN
```

```r
names(bacikg)
```

```
##  [1] "Removal.round"                  "village1"
##  [3] "site1"                          "sitearea1"
##  [5] "logsitearea1"                   "Date"
##  [7] "Nbre.de.personne"               "Nbre.de.jour"
##  [9] "manhr"                          "Kg.removed"
## [11] "log.kg."                        "X..Ceratophyllum"
## [13] "X..Typha"                       "X..Ludwigia"
## [15] "X..Potamogetum"                 "X..Nymphaea"
## [17] "X..Phragmites"                  "X..Cyperus"
## [19] "X..Salvinia"                    "X..Pistia"
## [21] "X..Azolla"                      "Photos...y...both.before.after."
## [23] "dayssince"                      "beforeemergent"
## [25] "beforefloating"                 "afteremergent"
## [27] "afterfloating"                  "difference.emergent"
## [29] "difference.floating"            "total.difference"
## [31] "log.total.diff."
```

```
par(mar=c(5,6,4,1)+.1)


ggplot(bacikg, aes(x=log.kg., y=log.total.diff.)) + geom_point(size=2)+
  geom_smooth(method=lm,
            color="black", fill="gray")+
  labs(title="",
       x="Log (kg vegetation removed)", y = "Log (Sq. m vegetation lost)")+
  theme_classic(base_size = 16)  + scale_y_continuous(
    labels = scales::number_format(accuracy = 1,
                                 decimal.mark = '.'))+ xlim(2, 5)+theme(axis.text=element_text(size=1
                                                                         axis.title=element_text(size=
```

```
## `geom_smooth()` using formula 'y ~ x'

## Warning: Removed 74 rows containing non-finite values (stat_smooth).

## Warning: Removed 74 rows containing missing values (geom_point).
```

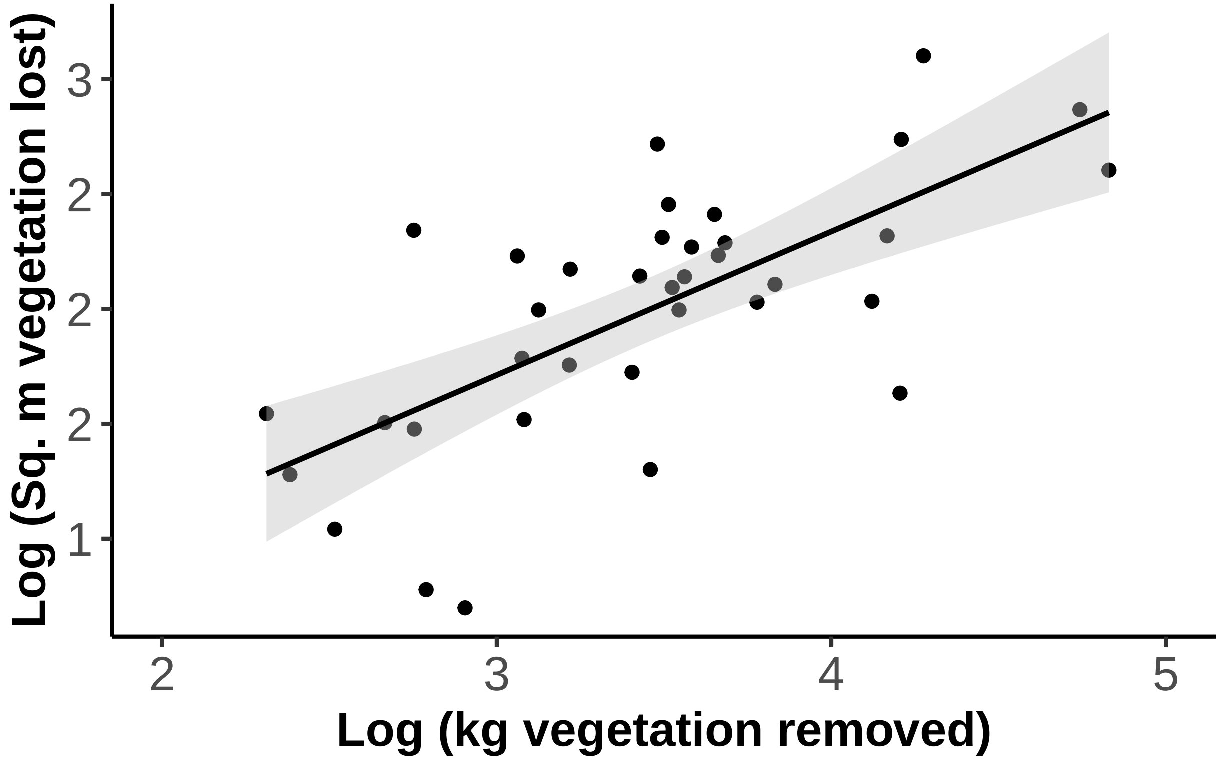


```
#above is...
#Sq.m vegetation cover lost vs. kg removed, note : log-transformed vegetation
#lost is only available for sites where vegetation cover was lost after removal
#thus not include where vegetation increased after removal (n=7 site visits)
```

```r
#Here is kg removed vs. number of snails lost after the removal event per sweep:
# load VegSnails - this is a list of each point by kg removed and change in number of snails, organized
#Number of snails removed by species per aquatic veg removed per sweep
VegSnails<-read.csv("VegSnails.csv")
VegSnails$lnDiff2<-(VegSnails$lnDiff*-1) # Change in number of snails is negative for fewer snails
cor.test(VegSnails$lnkmremoved,VegSnails$lnDiff2,method="spearman")
```

```
## Warning in cor.test.default(VegSnails$lnkmremoved, VegSnails$lnDiff2, method =
## "spearman"): Cannot compute exact p-value with ties
```

```
##
##  Spearman's rank correlation rho
##
## data:  VegSnails$lnkmremoved and VegSnails$lnDiff2
## S = 98634, p-value = 3.621e-05
## alternative hypothesis: true rho is not equal to 0
## sample estimates:
##        rho
## -0.4606985
```

```r
g<-ggplot(data = VegSnails, aes(x =lnkmremoved, y = lnDiff2)) +
  geom_point(aes(shape=Species, linetype=Species, color=Species), size=2)+
  scale_shape_manual(values=c(0, 2))+
  geom_smooth(method = "lm",
              aes(linetype = Species,color=Species,fill=Species)) +


  labs(x = expression(Natural~log~of~kg~of~aquatic~vegetation~removed/sweep),y = expression(Change~of~Na
```

```
## Warning: Ignoring unknown aesthetics: linetype
```

```r
g1<-g+ theme(panel.grid.major = element_blank(), panel.grid.minor = element_blank(),
        panel.background = element_blank(), panel.border = element_blank(), axis.line = element_line(c
g1
```

```
## `geom_smooth()` using formula 'y ~ x'
```

```
## Warning: Removed 4 rows containing non-finite values (stat_smooth).
```

```
## Warning: Removed 4 rows containing missing values (geom_point).
```

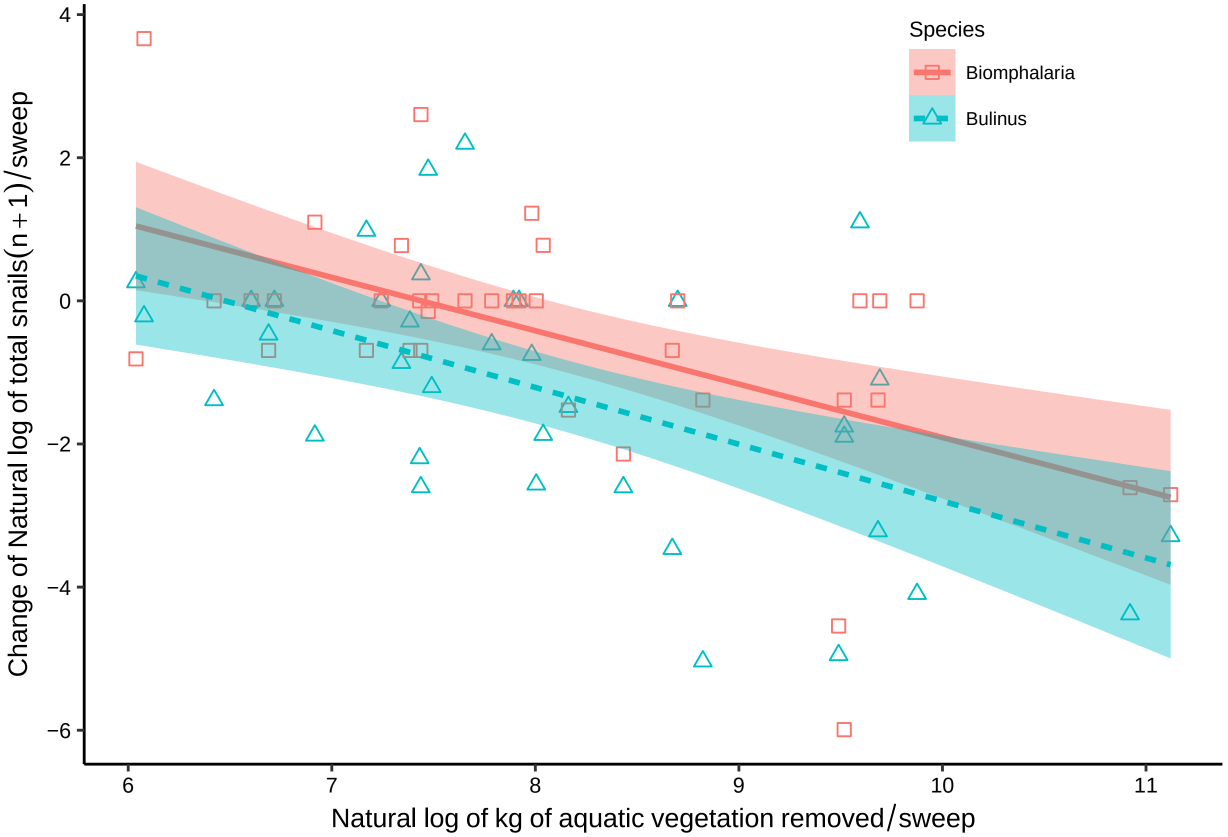


```r
#Figure 2B
```

```r
################################################
# repeated measures anova for kg veg removed :
################################################
bacikg$log.sitearea1=log(bacikg$sitearea1+1)
modelkg = glmmTMB(log.kg.~ manhr+Removal.round*logsitearea1+(1|site1/Removal.round),data=bacikg,na.acti
Anova(modelkg)
```

```
## Analysis of Deviance Table (Type II Wald chisquare tests)
##
## Response: log.kg.
##                              Chisq Df Pr(>Chisq)
## manhr                       2.1130  1  0.1460530
## Removal.round              11.4174  1  0.0007276 ***
## logsitearea1                0.5906  1  0.4421955
## Removal.round:logsitearea1  0.0380  1  0.8455114
## ---
## Signif. codes:  0 '***' 0.001 '**' 0.01 '*' 0.05 '.' 0.1 ' ' 1
```

```r
AIC(modelkg)
```

```
## [1] 130.1022
```

```r
#drop interaction:
modelkg2 = glmmTMB(log.kg.~ manhr+logsitearea1+Removal.round+(1|site1/Removal.round),data=bacikg,na.act
Anova(modelkg2)
```

```
## Analysis of Deviance Table (Type II Wald chisquare tests)
##
## Response: log.kg.
##                 Chisq Df Pr(>Chisq)
## manhr          2.0629  1  0.1509241
## logsitearea1   0.5924  1  0.4414744
## Removal.round 11.4031  1  0.0007332 ***
## ---
## Signif. codes:  0 '***' 0.001 '**' 0.01 '*' 0.05 '.' 0.1 ' ' 1
```

```r
AIC(modelkg2)
```

```
## [1] 128.1402
```

```r
#drop mnhr:
modelkg2.5 = glmmTMB(log.kg.~ logsitearea1+Removal.round+(1|site1/Removal.round),data=bacikg,na.action=
Anova(modelkg2.5)
```

```
## Analysis of Deviance Table (Type II Wald chisquare tests)
##
## Response: log.kg.
##                Chisq Df Pr(>Chisq)
## logsitearea1  3.2730  1    0.07043 .
## Removal.round 3.2959  1    0.06945 .
## ---
## Signif. codes:  0 '***' 0.001 '**' 0.01 '*' 0.05 '.' 0.1 ' ' 1
```

```r
AIC(modelkg2.5)
```

```
## [1] 162.6774
```

```r
visreg(modelkg2.5, "Removal.round", par.strip.text = list(col = "black"), points=list(col="black", cex=
```

```
## Warning in plot.window(...): "par.strip.text" is not a graphical parameter

## Warning in plot.xy(xy, type, ...): "par.strip.text" is not a graphical parameter

## Warning in axis(side = side, at = at, labels = labels, ...): "par.strip.text" is
## not a graphical parameter

## Warning in axis(side = side, at = at, labels = labels, ...): "par.strip.text" is
## not a graphical parameter

## Warning in box(...): "par.strip.text" is not a graphical parameter
```

```
## Warning in title(...): "par.strip.text" is not a graphical parameter
```

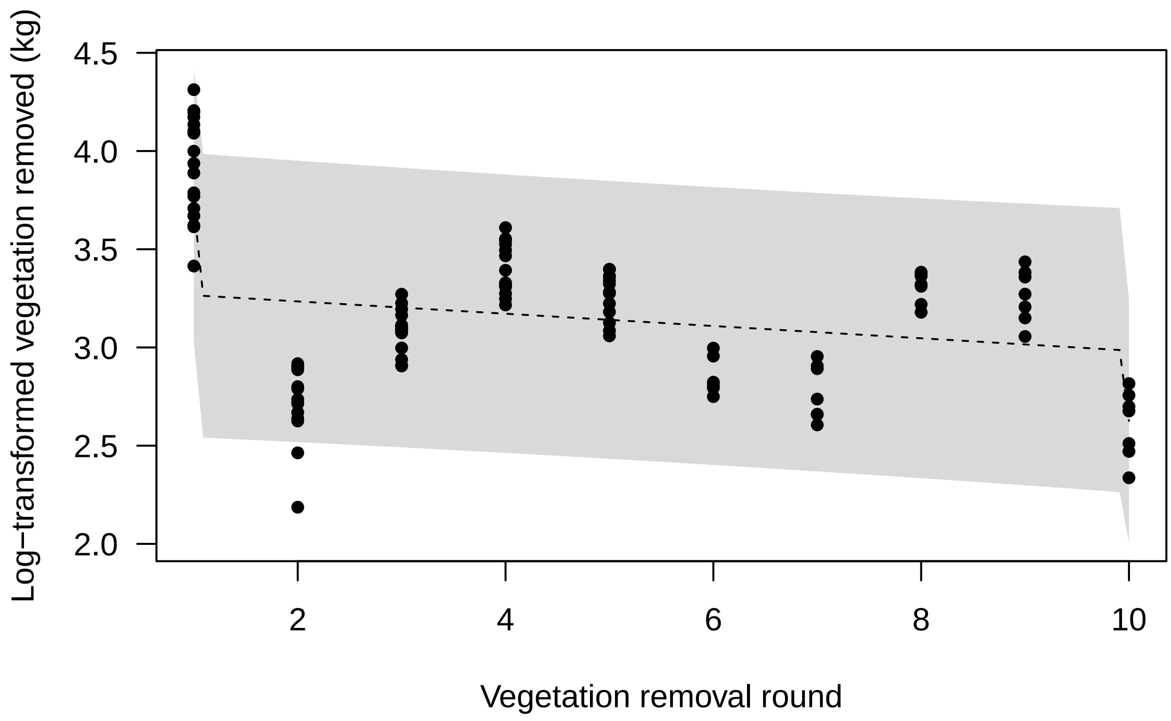


```r
#the above shows no variation in kg removed based on any round after the 1st...
#this means days since removal is unlikely to have much impact on kg removed or likely area of veg befo

visreg(modelkg2.5, "logsitearea1", par.strip.text = list(col = "black"),points=list(col="black", cex=.75
```

```
## Warning in plot.window(...): "par.strip.text" is not a graphical parameter

## Warning in plot.xy(xy, type, ...): "par.strip.text" is not a graphical parameter

## Warning in axis(side = side, at = at, labels = labels, ...): "par.strip.text" is
## not a graphical parameter

## Warning in axis(side = side, at = at, labels = labels, ...): "par.strip.text" is
## not a graphical parameter

## Warning in box(...): "par.strip.text" is not a graphical parameter

## Warning in title(...): "par.strip.text" is not a graphical parameter
```

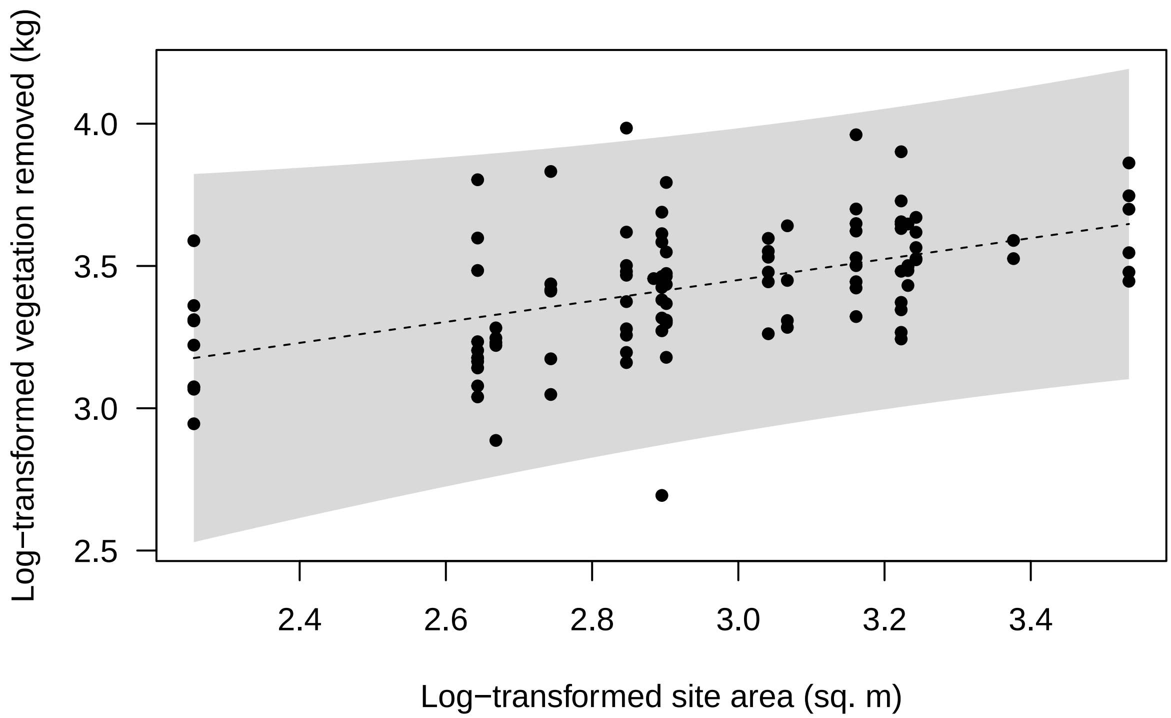


```
##########################
###### SNAIL PLOTS #######
##########################
#This file has all the malacology sampling from 2017-2019:
snails_data<-read.csv("snails_data.csv")
bacisnailall<-snails_data
bacisnailall$Year<-as.factor(bacisnailall$Year)
bacisnailall$nomass<-as.factor(bacisnailall$hadvegmassbeforeafter)
selected1718=c("2017","2018")
baci1718=bacisnailall[bacisnailall$Year %in% selected1718,]
baci1718$year <- baci1718$year[ , drop=TRUE]
baci1718$bul_and_biom<-(baci1718$Bulinus+baci1718$Biomphalaria)
baci1718$Location<-as.factor(baci1718$Location)
nall1718nb = glmer.nb(round(bul_and_biom) ~ Treatment*Time +(1|Site.ID/AccessID/Location), data=baci171
nall1718z = glmmTMB(round(bul_and_biom) ~ Treatment*Time +(1|Site.ID/AccessID/Location), data=baci1718,
nall1718P <- glmer(round(bul_and_biom) ~ Treatment*Time +(1|Site.ID/AccessID/Location), data=baci1718, 
AIC(nall1718P,nall1718nb,nall1718z)
```

```
##             df      AIC
## nall1718P    7 5504.076
## nall1718nb   8 3923.614
## nall1718z   10 3841.891
```

```
Anova(nall1718z)
```

```
## Analysis of Deviance Table (Type II Wald chisquare tests)
```

```
##
## Response: round(bul_and_biom)
##                  Chisq Df Pr(>Chisq)
## Treatment       1.4775  1     0.2242
## Time           55.3756  1  9.957e-14 ***
## Treatment:Time 40.9688  1  1.547e-10 ***
## ---
## Signif. codes:  0 '***' 0.001 '**' 0.01 '*' 0.05 '.' 0.1 ' ' 1
```

```
summary(nall1718z)
```

```
##  Family: nbinom2  ( log )
## Formula:
## round(bul_and_biom) ~ Treatment * Time + (1 | Site.ID/AccessID/Location)
## Zero inflation:                        ~nomass
## Data: baci1718
##
##      AIC      BIC   logLik deviance df.resid
##   3841.9   3893.6  -1910.9   3821.9     1287
##
## Random effects:
##
## Conditional model:
##  Groups                      Name        Variance Std.Dev.
##  Location:AccessID:Site.ID (Intercept) 0.09986  0.3160
##  AccessID:Site.ID          (Intercept) 2.10637  1.4513
##  Site.ID                   (Intercept) 0.33329  0.5773
## Number of obs: 1297, groups:
## Location:AccessID:Site.ID, 676; AccessID:Site.ID, 39; Site.ID, 19
##
## Dispersion parameter for nbinom2 family (): 0.419
##
## Conditional model:
##                             Estimate Std. Error z value Pr(>|z|)
## (Intercept)                   0.7956     0.4063   1.958  0.05020 .
## TreatmentRemoval             -1.8494     0.6170  -2.998  0.00272 **
## TimeBefore                    0.2199     0.2106   1.044  0.29638
## TreatmentRemoval:TimeBefore   1.9558     0.3056   6.401 1.55e-10 ***
## ---
## Signif. codes:  0 '***' 0.001 '**' 0.01 '*' 0.05 '.' 0.1 ' ' 1
##
## Zero-inflation model:
##             Estimate Std. Error z value Pr(>|z|)
## (Intercept)   1.1339     0.2640   4.295 1.75e-05 ***
## nomass1      -2.7519     0.5622  -4.895 9.84e-07 ***
## ---
## Signif. codes:  0 '***' 0.001 '**' 0.01 '*' 0.05 '.' 0.1 ' ' 1
```

```
lsmeans(nall1718z,pairwise~Treatment|Time)
```

```
## $lsmeans
## Time = After:
##  Treatment lsmean    SE   df lower.CL upper.CL
```

```
##  Control    0.796 0.406 1287 -0.00144    1.593
##  Removal   -1.054 0.491 1287 -2.01675   -0.091
##
## Time = Before:
##  Treatment lsmean    SE   df lower.CL upper.CL
##  Control    1.015 0.408 1287  0.21434    1.817
##  Removal    1.122 0.480 1287  0.17999    2.064
##
## Results are given on the log (not the response) scale.
## Confidence level used: 0.95
##
## $contrasts
## Time = After:
##  contrast          estimate    SE   df t.ratio p.value
##  Control - Removal    1.849 0.617 1287   2.998  0.0028
##
## Time = Before:
##  contrast          estimate    SE   df t.ratio p.value
##  Control - Removal   -0.106 0.605 1287  -0.176  0.8605
##
## Results are given on the log (not the response) scale.
```

```
e=allEffects(nall1718z)
```

```
## Warning in Effect.glmmTMB(predictors, mod, vcov. = vcov., ...): overriding
## variance function for effects: computed variances may be incorrect
```

```
e1=e[[1]]
e.df=as.data.frame(e1)
e.df$Time=factor(e.df$Time,levels=c("Before","After"))
e1718 <- ggplot(data=e.df, aes(x=Time, y=fit, group=Treatment))+ geom_line(aes(linetype=Treatment),size=
  ggtitle("")+ ylab("Snails per sweep")+ xlab("")+
  geom_point(position=position_dodge(w=0.1), size=2)+
  geom_errorbar(aes(ymax=upper, ymin=lower), width=0.4, position=position_dodge(w=0.1),size=0.45)+
  theme_bw() +
  theme(axis.line = element_line(colour = "black"),
        text=element_text(size=10),
        panel.grid.major = element_blank(),
        panel.grid.minor = element_blank(),
        panel.border = element_blank(),
        panel.background = element_blank(),axis.text=element_text(size=10,colour = "black"), legend.posi
e1718
```

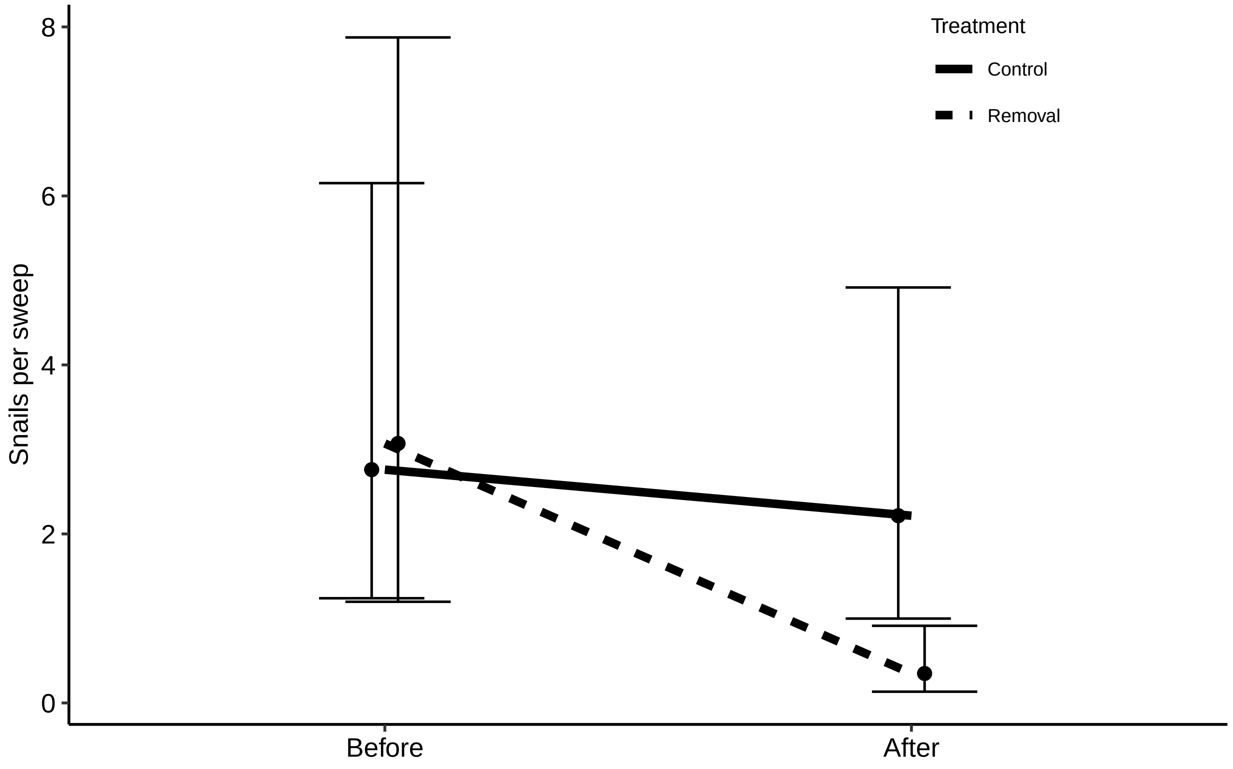


```
#Figure 2C
```

```
##################################
#########Human CHANGE ###########
##################################

#de-identified human infection data:
#updated data with 2019 human results:
#Load human_data_all_years
datanew<-read.csv("human_data_all_years.csv")
datanew$Intervention.type<-as.factor(datanew$Intervention.type)
datanew$Intervention.time<-as.factor(datanew$Intervention.time)
datanew$Intervention.time<- factor(datanew$Intervention.time,levels = c("before", "after"))
datanew$Sampling.bout<- factor(datanew$Sampling.bout)
#start with before and after visit for each village (see metadata and "Veg Removal Prelim Data 4_28")
datanew2=subset(datanew, Intervention.time=="before" | Intervention.time=="after")
datanew2$Intervention.time<- factor(datanew2$Intervention.time,levels = c("before", "after"))

#need to check that sd are about equal among groups before running BACI stats:
reports =ddply(datanew2,c("Intervention.time","Intervention.type"),summarize,
                    sdSh=sd(Sh,na.rm=TRUE),
                    sdSm=sd(Sm,na.rm=TRUE))
range(reports$sdSh) # SD for S. h
```

```
## [1] 0.4464321 0.4734385
```

```r
range(reports$sdSm)# SD for S. m
```

```
## [1] 0.3972223 0.4647883
```

```r
#acceptable
```

```r
          ###########################################
         ## BACI  (using the year before and after)##
          ##########################################

#Use data only from year before and after

datanew2=subset(datanew, Intervention.time=="before" | Intervention.time=="after")
datanew2$Intervention.time<- factor(datanew2$Intervention.time,levels = c("before", "after"))
dataFull=subset(datanew2, is.na(ShW)==FALSE & is.na(SmW)==FALSE)
dataFull=subset(datanew2, is.na(ShW)==FALSE & is.na(SmW)==FALSE)
dataFull=subset(dataFull, is.na(sex)==FALSE & is.na(lake)==FALSE & is.na(ClassNum)==FALSE)
dataFull=subset(dataFull, sex=="F" | sex=="M")
# Must have tested child for both Sm and Sh
#Class Num 4 includes grades 4-6
```

```r
# GLMM and NB GLMM Models for SAC infections (Before and after, 1 year each)
dataFull$lnSmW=log(dataFull$SmW+1)
dataFull$lnShW=log(dataFull$ShW+1)

# For Dredge comparison
veg.Sm.intensity_1b = lmer(lnSmW ~ Intervention.type*Intervention.time + sex+lake+ClassNum+(1|School/In
summary(veg.Sm.intensity_1b)
```

```
## Linear mixed model fit by maximum likelihood  ['lmerMod']
## Formula: lnSmW ~ Intervention.type * Intervention.time + sex + lake +
##     ClassNum + (1 | School/Intervention.time)
##    Data: dataFull
##
##      AIC      BIC   logLik deviance df.resid
##   7991.4   8048.6  -3985.7   7971.4     2237
##
## Scaled residuals:
##     Min      1Q  Median      3Q     Max
## -3.6137 -0.3482 -0.1473 -0.0280  4.7040
##
## Random effects:
##  Groups                   Name        Variance Std.Dev.
##  Intervention.time:School (Intercept) 0.1376   0.371
##  School                   (Intercept) 1.3610   1.167
##  Residual                             1.9442   1.394
## Number of obs: 2247, groups:  Intervention.time:School, 32; School, 16
##
## Fixed effects:
##                                              Estimate Std. Error t value
## (Intercept)                                   1.02280    0.50235   2.036
## Intervention.typeremoval                      0.34355    0.61772   0.556
```

```
## Intervention.timeafter                             0.47331    0.20708   2.286
## sexM                                                0.14924    0.05932   2.516
## lakeriver                                          -0.85029    0.62106  -1.369
## ClassNum                                           -0.01007    0.03368  -0.299
## Intervention.typeremoval:Intervention.timeafter -0.46079    0.29132  -1.582
##
## Correlation of Fixed Effects:
##             (Intr) Intrvntn.ty Intrvntn.tm sexM   lakrvr ClssNm
## Intrvntn.ty -0.615
## Intrvntn.tm -0.205  0.162
## sexM        -0.046 -0.005       0.003
## lakeriver   -0.469  0.000       0.001      -0.007
## ClassNum    -0.157 -0.001       0.033      -0.041  0.031
## Intrvnt.:I.  0.145 -0.229      -0.711      -0.003  0.000 -0.022
```

```
Anova(veg.Sm.intensity_1b)
```

```
## Analysis of Deviance Table (Type II Wald chisquare tests)
##
## Response: lnSmW
##                                      Chisq Df Pr(>Chisq)
## Intervention.type                   0.0397  1    0.84215
## Intervention.time                   2.7262  1    0.09872 .
## sex                                 6.3295  1    0.01187 *
## lake                                1.8745  1    0.17097
## ClassNum                            0.0894  1    0.76496
## Intervention.type:Intervention.time 2.5019  1    0.11371
## ---
## Signif. codes:  0 '***' 0.001 '**' 0.01 '*' 0.05 '.' 0.1 ' ' 1
```

```
veg.Sh.intensityb = lmer(lnShW ~ Intervention.type*Intervention.time + sex+lake+ClassNum+(1|School/Inte
summary(veg.Sh.intensityb)
```

```
## Linear mixed model fit by maximum likelihood  ['lmerMod']
## Formula: lnShW ~ Intervention.type * Intervention.time + sex + lake +
##     ClassNum + (1 | School/Intervention.time)
##    Data: dataFull
##
##      AIC      BIC   logLik deviance df.resid
##   8541.8   8598.9  -4260.9   8521.8     2237
##
## Scaled residuals:
##     Min      1Q  Median      3Q     Max
## -2.9594 -0.6456 -0.1900  0.6983  3.2583
##
## Random effects:
##  Groups                   Name        Variance Std.Dev.
##  Intervention.time:School (Intercept) 0.4578   0.6766
##  School                   (Intercept) 0.2696   0.5192
##  Residual                             2.4916   1.5785
## Number of obs: 2247, groups:  Intervention.time:School, 32; School, 16
##
## Fixed effects:
```

```
##                                                   Estimate Std. Error t value
## (Intercept)                                       3.108275   0.352460   8.819
## Intervention.typeremoval                         -0.055242   0.436787  -0.126
## Intervention.timeafter                           -0.002514   0.354287  -0.007
## sexM                                              0.295908   0.067158   4.406
## lakeriver                                        -1.642971   0.372305  -4.413
## ClassNum                                         -0.274225   0.038132  -7.191
## Intervention.typeremoval:Intervention.timeafter   0.208186   0.499774   0.417
##
## Correlation of Fixed Effects:
##             (Intr) Intrvntn.ty Intrvntn.tm sexM   lakrvr ClssNm
## Intrvntn.ty -0.620
## Intrvntn.tm -0.502  0.400
## sexM        -0.075 -0.008       0.002
## lakeriver   -0.412  0.000       0.002      -0.013
## ClassNum    -0.253 -0.002       0.021      -0.040  0.057
## Intrvnt.:I.  0.355 -0.565      -0.709      -0.002  0.000 -0.015
```

```
Anova(veg.Sh.intensityb)
```

```
## Analysis of Deviance Table (Type II Wald chisquare tests)
##
## Response: lnShW
##                                     Chisq Df Pr(>Chisq)
## Intervention.type                  0.0174  1     0.8950
## Intervention.time                  0.1670  1     0.6828
## sex                               19.4140  1  1.052e-05 ***
## lake                              19.4744  1  1.020e-05 ***
## ClassNum                          51.7173  1  6.410e-13 ***
## Intervention.type:Intervention.time  0.1735  1     0.6770
## ---
## Signif. codes:  0 '***' 0.001 '**' 0.01 '*' 0.05 '.' 0.1 ' ' 1
```

```
# For Final Model
veg.Sm.intensity_1 = lmer(lnSmW ~ Intervention.type*Intervention.time + sex+lake+ClassNum+(1|School/Inte
summary(veg.Sm.intensity_1b)
```

```
## Linear mixed model fit by maximum likelihood  ['lmerMod']
## Formula: lnSmW ~ Intervention.type * Intervention.time + sex + lake +
##     ClassNum + (1 | School/Intervention.time)
##    Data: dataFull
##
##      AIC      BIC   logLik deviance df.resid
##   7991.4   8048.6  -3985.7   7971.4     2237
##
## Scaled residuals:
##     Min      1Q  Median      3Q     Max
## -3.6137 -0.3482 -0.1473 -0.0280  4.7040
##
## Random effects:
##  Groups                   Name        Variance Std.Dev.
##  Intervention.time:School (Intercept) 0.1376   0.371
##  School                   (Intercept) 1.3610   1.167
```

```
##  Residual                             1.9442   1.394
## Number of obs: 2247, groups:  Intervention.time:School, 32; School, 16
##
## Fixed effects:
##                                                  Estimate Std. Error t value
## (Intercept)                                       1.02280    0.50235   2.036
## Intervention.typeremoval                          0.34355    0.61772   0.556
## Intervention.timeafter                            0.47331    0.20708   2.286
## sexM                                              0.14924    0.05932   2.516
## lakeriver                                        -0.85029    0.62106  -1.369
## ClassNum                                         -0.01007    0.03368  -0.299
## Intervention.typeremoval:Intervention.timeafter -0.46079    0.29132  -1.582
##
## Correlation of Fixed Effects:
##             (Intr) Intrvntn.ty Intrvntn.tm sexM   lakrvr ClssNm
## Intrvntn.ty -0.615
## Intrvntn.tm -0.205  0.162
## sexM        -0.046 -0.005       0.003
## lakeriver   -0.469  0.000       0.001      -0.007
## ClassNum    -0.157 -0.001       0.033      -0.041  0.031
## Intrvnt.:I.  0.145 -0.229      -0.711      -0.003  0.000 -0.022
```

```
Anova(veg.Sm.intensity_1b)
```

```
## Analysis of Deviance Table (Type II Wald chisquare tests)
##
## Response: lnSmW
##                                    Chisq Df Pr(>Chisq)
## Intervention.type                 0.0397  1    0.84215
## Intervention.time                 2.7262  1    0.09872 .
## sex                               6.3295  1    0.01187 *
## lake                              1.8745  1    0.17097
## ClassNum                          0.0894  1    0.76496
## Intervention.type:Intervention.time 2.5019  1    0.11371
## ---
## Signif. codes:  0 '***' 0.001 '**' 0.01 '*' 0.05 '.' 0.1 ' ' 1
```

```
veg.Sh.intensity = lmer(lnShW ~ Intervention.type*Intervention.time + sex+lake+ClassNum+(1|School/Inter
summary(veg.Sh.intensityb)
```

```
## Linear mixed model fit by maximum likelihood  ['lmerMod']
## Formula: lnShW ~ Intervention.type * Intervention.time + sex + lake +
##     ClassNum + (1 | School/Intervention.time)
##    Data: dataFull
##
##      AIC      BIC   logLik deviance df.resid
##   8541.8   8598.9  -4260.9   8521.8     2237
##
## Scaled residuals:
##     Min      1Q  Median      3Q     Max
## -2.9594 -0.6456 -0.1900  0.6983  3.2583
##
## Random effects:
```

```
##  Groups                     Name        Variance Std.Dev.
##  Intervention.time:School (Intercept) 0.4578   0.6766
##  School                   (Intercept) 0.2696   0.5192
##  Residual                             2.4916   1.5785
## Number of obs: 2247, groups:  Intervention.time:School, 32; School, 16
##
## Fixed effects:
##                                             Estimate Std. Error t value
## (Intercept)                                 3.108275   0.352460   8.819
## Intervention.typeremoval                   -0.055242   0.436787  -0.126
## Intervention.timeafter                     -0.002514   0.354287  -0.007
## sexM                                        0.295908   0.067158   4.406
## lakeriver                                  -1.642971   0.372305  -4.413
## ClassNum                                   -0.274225   0.038132  -7.191
## Intervention.typeremoval:Intervention.timeafter  0.208186   0.499774   0.417
##
## Correlation of Fixed Effects:
##             (Intr) Intrvntn.ty Intrvntn.tm sexM   lakrvr ClssNm
## Intrvntn.ty -0.620
## Intrvntn.tm -0.502  0.400
## sexM        -0.075 -0.008       0.002
## lakeriver   -0.412  0.000       0.002      -0.013
## ClassNum    -0.253 -0.002       0.021      -0.040  0.057
## Intrvnt.:I.  0.355 -0.565      -0.709      -0.002  0.000 -0.015
```

```
Anova(veg.Sh.intensityb)
```

```
## Analysis of Deviance Table (Type II Wald chisquare tests)
##
## Response: lnShW
##                                      Chisq Df Pr(>Chisq)
## Intervention.type                   0.0174  1     0.8950
## Intervention.time                   0.1670  1     0.6828
## sex                                19.4140  1  1.052e-05 ***
## lake                               19.4744  1  1.020e-05 ***
## ClassNum                           51.7173  1  6.410e-13 ***
## Intervention.type:Intervention.time  0.1735  1     0.6770
## ---
## Signif. codes:  0 '***' 0.001 '**' 0.01 '*' 0.05 '.' 0.1 ' ' 1
```

```
#For Prev
veg.Sh.prevx <- glmer(Sh ~ Intervention.time*Intervention.type +sex+lake+ClassNum+ (1|School/Interventi
summary(veg.Sh.prevx)
```

```
## Generalized linear mixed model fit by maximum likelihood (Laplace
##   Approximation) [glmerMod]
##  Family: binomial  ( logit )
## Formula: Sh ~ Intervention.time * Intervention.type + sex + lake + ClassNum +
##     (1 | School/Intervention.time)
##    Data: dataFull
## Control: glmerControl(optimizer = "bobyqa", optCtrl = list(maxfun = 1e+05))
##
##      AIC      BIC   logLik deviance df.resid
```

```
##    2118.8   2170.2  -1050.4   2100.8     2238
##
## Scaled residuals:
##     Min      1Q  Median      3Q     Max
## -9.3819 -0.6289  0.3280  0.4880  3.2221
##
## Random effects:
##  Groups                   Name        Variance Std.Dev.
##  Intervention.time:School (Intercept) 0.6436   0.8023
##  School                   (Intercept) 0.8233   0.9074
## Number of obs: 2247, groups:  Intervention.time:School, 32; School, 16
##
## Fixed effects:
##                                                Estimate Std. Error z value
## (Intercept)                                     2.45948    0.53803   4.571
## Intervention.timeafter                         -0.26693    0.46410  -0.575
## Intervention.typeremoval                       -0.43718    0.63633  -0.687
## sexM                                            0.44161    0.11104   3.977
## lakeriver                                      -2.49783    0.57312  -4.358
## ClassNum                                       -0.12673    0.06441  -1.967
## Intervention.timeafter:Intervention.typeremoval  0.30600    0.63738   0.480
##                                                Pr(>|z|)
## (Intercept)                                    4.85e-06 ***
## Intervention.timeafter                           0.5652
## Intervention.typeremoval                         0.4921
## sexM                                           6.97e-05 ***
## lakeriver                                      1.31e-05 ***
## ClassNum                                         0.0491 *
## Intervention.timeafter:Intervention.typeremoval   0.6312
## ---
## Signif. codes:  0 '***' 0.001 '**' 0.01 '*' 0.05 '.' 0.1 ' ' 1
##
## Correlation of Fixed Effects:
##             (Intr) Intrvntn.tm Intrvntn.ty sexM   lakrvr ClssNm
## Intrvntn.tm -0.426
## Intrvntn.ty -0.620  0.352
## sexM        -0.068  0.001      -0.009
## lakeriver   -0.447  0.004       0.021      -0.031
## ClassNum    -0.293  0.035       0.000      -0.029  0.070
## Intrvnt.:I.  0.309 -0.728      -0.483       0.000  0.000 -0.024
```

```
Anova(veg.Sh.prevx)
```

```
## Analysis of Deviance Table (Type II Wald chisquare tests)
##
## Response: Sh
##                                     Chisq Df Pr(>Chisq)
## Intervention.time                  0.1084  1    0.74202
## Intervention.type                  0.2702  1    0.60319
## sex                               15.8177  1  6.975e-05 ***
## lake                              18.9947  1  1.311e-05 ***
## ClassNum                           3.8707  1    0.04913 *
## Intervention.time:Intervention.type  0.2305  1    0.63116
## ---
```

```
## Signif. codes:  0 '***' 0.001 '**' 0.01 '*' 0.05 '.' 0.1 ' ' 1
```

```
veg.Sm.prevx <- glmer(Sm ~ Intervention.time*Intervention.type +sex+lake+ClassNum+ (1|School/Interventi
summary(veg.Sm.prevx)
```

```
## Generalized linear mixed model fit by maximum likelihood (Laplace
##   Approximation) [glmerMod]
##  Family: binomial  ( logit )
## Formula: Sm ~ Intervention.time * Intervention.type + sex + lake + ClassNum +
##     (1 | School/Intervention.time)
##    Data: dataFull
## Control: glmerControl(optimizer = "bobyqa", optCtrl = list(maxfun = 1e+05))
##
##      AIC      BIC   logLik deviance df.resid
##   1863.9   1915.3   -922.9   1845.9     2238
##
## Scaled residuals:
##     Min      1Q  Median      3Q     Max
## -2.9272 -0.4241 -0.2553 -0.1735  5.2005
##
## Random effects:
##  Groups                   Name        Variance Std.Dev.
##  Intervention.time:School (Intercept) 0.09979  0.3159
##  School                   (Intercept) 2.16129  1.4701
## Number of obs: 2247, groups:  Intervention.time:School, 32; School, 16
##
## Fixed effects:
##                                              Estimate Std. Error z value
## (Intercept)                                 -1.681565   0.649771  -2.588
## Intervention.timeafter                       0.779938   0.250847   3.109
## Intervention.typeremoval                     0.633997   0.778050   0.815
## sexM                                         0.211461   0.120261   1.758
## lakeriver                                   -1.026026   0.784887  -1.307
## ClassNum                                    -0.002861   0.068072  -0.042
## Intervention.timeafter:Intervention.typeremoval -0.807807   0.351946  -2.295
##                                             Pr(>|z|)
## (Intercept)                                  0.00966 **
## Intervention.timeafter                       0.00188 **
## Intervention.typeremoval                     0.41516
## sexM                                         0.07869 .
## lakeriver                                    0.19114
## ClassNum                                     0.96648
## Intervention.timeafter:Intervention.typeremoval  0.02172 *
## ---
## Signif. codes:  0 '***' 0.001 '**' 0.01 '*' 0.05 '.' 0.1 ' ' 1
##
## Correlation of Fixed Effects:
##             (Intr) Intrvntn.tm Intrvntn.ty sexM   lakrvr ClssNm
## Intrvntn.tm -0.209
## Intrvntn.ty -0.604  0.158
## sexM        -0.078  0.016      -0.006
## lakeriver   -0.465  0.005      -0.001      -0.014
## ClassNum    -0.245  0.055      -0.005      -0.045  0.050
## Intrvnt.:I.  0.140 -0.700      -0.218      -0.010  0.008 -0.030
```

```r
Anova(veg.Sm.prevx)
```

```
## Analysis of Deviance Table (Type II Wald chisquare tests)
##
## Response: Sm
##                                      Chisq Df Pr(>Chisq)
## Intervention.time                   4.4265  1    0.03538 *
## Intervention.type                   0.1043  1    0.74679
## sex                                 3.0918  1    0.07869 .
## lake                                1.7088  1    0.19114
## ClassNum                            0.0018  1    0.96648
## Intervention.time:Intervention.type 5.2682  1    0.02172 *
## ---
## Signif. codes:  0 '***' 0.001 '**' 0.01 '*' 0.05 '.' 0.1 ' ' 1
```

```r
#Dredge for each model
# Egg count Sm
results2<-dredge(veg.Sm.intensity_1b)
```

```
## Fixed term is "(Intercept)"
```

```r
results2
```

```
## Global model call: lmer(formula = lnSmW ~ Intervention.type * Intervention.time +
##     sex + lake + ClassNum + (1 | School/Intervention.time), data = dataFull,
##     REML = FALSE, na.action = na.fail)
## ---
## Model selection table
##    (Intrc)       ClN Intrv.tim Intrv.typ lak sex Intrv.tim:Intrv.typ df
## 19  0.8545                   +                +                      6
## 17  0.9732                                    +                      5
## 27  1.1720                   +             +  +                      7
## 25  1.2920                                 +  +                      6
## 55  0.6818                   +         +      +                   +  8
## 63  0.9992                   +         +   +  +                   +  9
## 20  0.8757 -0.009923         +                +                      7
## 23  0.7951                   +         +      +                      7
## 18  0.9963 -0.011040                          +                      6
## 28  1.1990 -0.011350         +             +  +                      8
## 21  0.9138                             +      +                      6
## 31  1.1130                   +         +   +  +                      8
## 26  1.3200 -0.012470                       +  +                      7
## 29  1.2330                             +   +  +                      7
## 56  0.7002 -0.008632         +         +      +                   +  9
## 64  1.0230 -0.010070         +         +   +  +                   + 10
## 24  0.8159 -0.009962         +         +      +                      8
## 22  0.9363 -0.011080                   +      +                      7
## 32  1.1390 -0.011400         +         +   +  +                      9
## 3   0.9279                   +                                       5
## 30  1.2610 -0.012510                   +   +  +                      8
## 1   1.0460                                                           4
## 11  1.2430                   +             +                         6
```

```
## 9   1.3620                               +                             5
## 39  0.7513               +         +                              +  7
## 47  1.0660               +         +    +                         +  8
## 7   0.8639               +         +                                 6
## 4   0.9421 -0.006483     +                                           6
## 2   1.0620 -0.007610                                                 5
## 5   0.9821                         +                                 5
## 12  1.2610 -0.007884     +              +                            7
## 15  1.1790               +         +    +                            7
## 10  1.3830 -0.009012                    +                            6
## 13  1.2980                         +    +                            6
## 40  0.7625 -0.005185     +         +                              +  8
## 48  1.0810 -0.006597     +         +    +                         +  9
## 8   0.8777 -0.006527     +         +                                 7
## 6   0.9979 -0.007654               +                                 6
## 16  1.1970 -0.007937     +         +    +                            8
## 14  1.3180 -0.009064               +    +                            7
##       logLik   AICc delta weight
## 19 -3987.807 7987.7  0.00  0.113
## 17 -3988.924 7987.9  0.22  0.101
## 27 -3986.933 7987.9  0.27  0.099
## 25 -3988.046 7988.1  0.48  0.089
## 55 -3986.617 7989.3  1.65  0.050
## 63 -3985.742 7989.6  1.91  0.043
## 20 -3987.764 7989.6  1.93  0.043
## 23 -3987.790 7989.6  1.98  0.042
## 18 -3988.871 7989.8  2.13  0.039
## 28 -3986.877 7989.8  2.17  0.038
## 21 -3988.907 7989.9  2.20  0.038
## 31 -3986.914 7989.9  2.24  0.037
## 26 -3987.977 7990.0  2.35  0.035
## 29 -3988.026 7990.1  2.45  0.033
## 56 -3986.584 7991.2  3.60  0.019
## 64 -3985.698 7991.5  3.84  0.017
## 24 -3987.746 7991.6  3.90  0.016
## 22 -3988.853 7991.8  4.10  0.015
## 32 -3986.857 7991.8  4.14  0.014
## 3  -3990.922 7991.9  4.22  0.014
## 30 -3987.957 7992.0  4.33  0.013
## 1  -3992.040 7992.1  4.45  0.012
## 11 -3990.067 7992.2  4.52  0.012
## 9  -3991.179 7992.4  4.73  0.011
## 39 -3989.733 7993.5  5.86  0.006
## 47 -3988.877 7993.8  6.17  0.005
## 7  -3990.902 7993.8  6.19  0.005
## 4  -3990.904 7993.8  6.19  0.005
## 2  -3992.014 7994.1  6.40  0.005
## 5  -3992.019 7994.1  6.41  0.005
## 12 -3990.039 7994.1  6.48  0.004
## 15 -3990.044 7994.1  6.49  0.004
## 10 -3991.144 7994.3  6.67  0.004
## 13 -3991.157 7994.4  6.70  0.004
## 40 -3989.721 7995.5  7.86  0.002
## 48 -3988.858 7995.8  8.14  0.002
```

```
## 8  -3990.883 7995.8  8.16  0.002
## 6  -3991.994 7996.0  8.37  0.002
## 16 -3990.017 7996.1  8.45  0.002
## 14 -3991.121 7996.3  8.64  0.002
## Models ranked by AICc(x)
## Random terms (all models):
## '1 | School/Intervention.time'
```

```
importance(results2)
```

```
##                         sex  Intervention.time lake Intervention.type ClassNum
## Sum of weights:      0.89 0.59              0.47 0.38              0.28
## N containing models:   20   24                20   24                20
##                         Intervention.time:Intervention.type
## Sum of weights:      0.14
## N containing models:    8
```

```
Smi_2<-model.avg(results2, subset= delta <2, revised.var = TRUE)
Smi_2
```

```
##
## Call:
## model.avg(object = results2, subset = delta < 2, revised.var = TRUE)
##
## Component models:
## '25'    '5'     '245'   '45'    '2356'  '23456' '125'   '235'
##
## Coefficients:
##        (Intercept) Intervention.timeafter     sexM  lakeriver
## full     0.9897642              0.2002312 0.148288 -0.3376231
## subset   0.9897642              0.2977715 0.148288 -0.8466021
##        Intervention.typeremoval Intervention.timeafter:Intervention.typeremoval
## full                 0.06367541                                    -0.07415517
## subset               0.27355007                                    -0.46251365
##             ClassNum
## full   -0.0007377965
## subset -0.0099230710
```

```
# Egg count Sh
results3<-dredge(veg.Sh.intensityb)
```

```
## Fixed term is "(Intercept)"
```

```
results3
```

```
## Global model call: lmer(formula = lnShW ~ Intervention.type * Intervention.time +
##     sex + lake + ClassNum + (1 | School/Intervention.time), data = dataFull,
##     REML = FALSE, na.action = na.fail)
## ---
## Model selection table
##    (Intrc)     ClN Intrv.tim Intrv.typ lak sex Intrv.tim:Intrv.typ df    logLik
```

```
## 26   3.131 -0.2741                       +   +                      7 -4261.054
## 28   3.080 -0.2739         +             +   +                      8 -4260.972
## 30   3.107 -0.2742                   +   +   +                      8 -4261.046
## 32   3.056 -0.2740         +         +   +   +                      9 -4260.963
## 64   3.108 -0.2742         +         +   +   +                   + 10 -4260.877
## 18   2.507 -0.2710                           +                      6 -4267.452
## 20   2.456 -0.2708         +                 +                      7 -4267.369
## 22   2.483 -0.2710                   +       +                      7 -4267.448
## 24   2.432 -0.2708         +         +       +                      8 -4267.365
## 56   2.485 -0.2711         +         +       +                   +  9 -4267.275
## 10   3.255 -0.2674                       +                          6 -4270.728
## 12   3.204 -0.2672         +             +                          7 -4270.648
## 14   3.222 -0.2675                   +   +                          7 -4270.712
## 16   3.172 -0.2672         +         +   +                          8 -4270.631
## 48   3.225 -0.2675         +         +   +                       +  9 -4270.541
## 2    2.638 -0.2642                                                  5 -4277.027
## 4    2.587 -0.2640         +                                        6 -4276.944
## 6    2.605 -0.2642                   +                              6 -4277.019
## 8    2.555 -0.2640         +         +                              7 -4276.937
## 40   2.609 -0.2643         +         +                           +  8 -4276.844
## 25   2.501                               +   +                      6 -4286.593
## 27   2.437                 +             +   +                      7 -4286.469
## 29   2.493                           +   +   +                      7 -4286.592
## 31   2.429                 +         +   +   +                      8 -4286.468
## 63   2.467                 +         +   +   +                   +  9 -4286.424
## 17   1.941                                   +                      5 -4292.298
## 19   1.876                 +                 +                      6 -4292.172
## 21   1.932                           +       +                      6 -4292.297
## 23   1.868                 +         +       +                      7 -4292.172
## 9    2.632                               +                          5 -4294.873
## 55   1.907                 +         +       +                   +  8 -4292.125
## 11   2.568                 +             +                          6 -4294.751
## 13   2.615                           +   +                          6 -4294.868
## 15   2.552                 +         +   +                          7 -4294.747
## 47   2.591                 +         +   +                       +  8 -4294.700
## 1    2.076                                                          4 -4300.488
## 3    2.013                 +                                        5 -4300.365
## 5    2.059                           +                              5 -4300.486
## 7    1.996                 +         +                              6 -4300.363
## 39   2.036                 +         +                           +  7 -4300.314
##      AICc delta weight
## 26 8536.2  0.00  0.505
## 28 8538.0  1.85  0.200
## 30 8538.2  2.00  0.186
## 32 8540.0  3.85  0.074
## 64 8541.9  5.69  0.029
## 18 8546.9 10.78  0.002
## 20 8548.8 12.63  0.001
## 22 8548.9 12.79  0.001
## 24 8550.8 14.63  0.000
## 56 8552.6 16.47  0.000
## 10 8553.5 17.34  0.000
## 12 8555.3 19.19  0.000
## 14 8555.5 19.31  0.000
```

```
## 16 8557.3 21.17  0.000
## 48 8559.2 23.00  0.000
## 2  8564.1 27.92  0.000
## 4  8565.9 29.77  0.000
## 6  8566.1 29.92  0.000
## 8  8567.9 31.76  0.000
## 40 8569.8 33.59  0.000
## 25 8585.2 49.06  0.000
## 27 8587.0 50.83  0.000
## 29 8587.2 51.07  0.000
## 31 8589.0 52.84  0.000
## 63 8590.9 54.77  0.000
## 17 8594.6 58.46  0.000
## 19 8596.4 60.22  0.000
## 21 8596.6 60.47  0.000
## 23 8598.4 62.23  0.000
## 9  8599.8 63.61  0.000
## 55 8600.3 64.16  0.000
## 11 8601.5 65.38  0.000
## 13 8601.8 65.62  0.000
## 15 8603.5 67.39  0.000
## 47 8605.5 69.31  0.000
## 1  8609.0 72.84  0.000
## 3  8610.8 74.60  0.000
## 5  8611.0 74.84  0.000
## 7  8612.8 76.61  0.000
## 39 8614.7 78.52  0.000
## Models ranked by AICc(x)
## Random terms (all models):
## '1 | School/Intervention.time'
```

```
importance(results3)
```

```
##                      ClassNum sex  lake Intervention.time Intervention.type
## Sum of weights:      1.00     1.00 1.00 0.30              0.29
## N containing models:   20       20   20   24                24
##                      Intervention.time:Intervention.type
## Sum of weights:      0.03
## N containing models:    8
```

```
Shi_3<-model.avg(results3, subset= delta <2, revised.var = TRUE)
Shi_3
```

```
##
## Call:
## model.avg(object = results3, subset = delta < 2, revised.var = TRUE)
##
## Component models:
## '145'  '1245' '1345'
##
## Coefficients:
##        (Intercept)   ClassNum lakeriver      sexM Intervention.timeafter
## full      3.114434 -0.2740775 -1.643425 0.2960226             0.02295057
```

```
## subset    3.114434 -0.2740775 -1.643425 0.2960226             0.10215381
##        Intervention.typeremoval
## full                0.009965576
## subset              0.047735891
```

```r
# Sh prev
results4<-dredge(veg.Sh.prevx)
```

```
## Fixed term is "(Intercept)"
```

```r
results4
```

```
## Global model call: glmer(formula = Sh ~ Intervention.time * Intervention.type +
##     sex + lake + ClassNum + (1 | School/Intervention.time), data = dataFull,
##     family = binomial, control = glmerControl(optimizer = "bobyqa",
##         optCtrl = list(maxfun = 1e+05)), na.action = na.fail)
## ---
## Model selection table
##    (Intrc)     ClN Intrv.tim Intrv.typ lak sex Intrv.tim:Intrv.typ df    logLik
## 26  2.1800 -0.1258                       +   +                      6 -1050.683
## 30  2.3290 -0.1254                   +   +   +                      7 -1050.547
## 25  1.8840                               +   +                      5 -1052.568
## 28  2.2310 -0.1263         +             +   +                      7 -1050.632
## 29  2.0430                           +   +   +                      6 -1052.419
## 32  2.3800 -0.1259         +         +   +   +                      8 -1050.496
## 27  1.9270                 +             +   +                      6 -1052.530
## 31  2.0860                 +         +   +   +                      7 -1052.382
## 64  2.4600 -0.1267         +         +   +   +                   +  9 -1050.383
## 63  2.1550                 +         +   +   +                   +  8 -1052.292
## 18  1.2420 -0.1161                           +                      5 -1056.905
## 17  0.9944                                   +                      4 -1058.508
## 22  1.4110 -0.1158                   +       +                      6 -1056.827
## 20  1.2930 -0.1167         +                 +                      6 -1056.853
## 21  1.1690                           +       +                      5 -1058.421
## 19  1.0380                 +                 +                      5 -1058.469
## 10  2.3550 -0.1193                       +                          5 -1058.561
## 24  1.4620 -0.1164         +         +       +                      7 -1056.775
## 9   2.0730                               +                          4 -1060.271
## 23  1.2120                 +         +       +                      6 -1058.383
## 14  2.4930 -0.1189                   +   +                          6 -1058.446
## 12  2.4080 -0.1199         +             +                          6 -1058.505
## 56  1.5440 -0.1172         +         +       +                   +  8 -1056.658
## 13  2.2180                           +   +                          5 -1060.145
## 11  2.1170                 +             +                          5 -1060.230
## 55  1.2850                 +         +       +                   +  7 -1058.288
## 16  2.5450 -0.1195         +         +   +                          7 -1058.390
## 15  2.2620                 +         +   +                          6 -1060.104
## 48  2.6250 -0.1203         +         +   +                       +  8 -1058.272
## 47  2.3330                 +         +   +                       +  7 -1060.008
## 2   1.4350 -0.1096                                                  4 -1064.670
## 1   1.1980                                                          3 -1066.112
## 6   1.5910 -0.1094                   +                              5 -1064.601
## 4   1.4870 -0.1102         +                                        5 -1064.614
```

```
## 5   1.3600                         +                             4 -1066.036
## 3   1.2420               +                                       4 -1066.070
## 8   1.6420 -0.1100       +         +                             6 -1064.546
## 7   1.4040               +         +                             5 -1065.995
## 40  1.7250 -0.1108       +         +                          +  7 -1064.422
## 39  1.4780               +         +                          +  6 -1065.893
##      AICc delta weight
## 26 2113.4  0.00  0.346
## 30 2115.1  1.74  0.145
## 25 2115.2  1.76  0.143
## 28 2115.3  1.91  0.133
## 29 2116.9  3.47  0.061
## 32 2117.1  3.65  0.056
## 27 2117.1  3.69  0.055
## 31 2118.8  5.41  0.023
## 64 2118.8  5.44  0.023
## 63 2120.6  7.24  0.009
## 18 2123.8 10.43  0.002
## 17 2125.0 11.63  0.001
## 22 2125.7 12.29  0.001
## 20 2125.7 12.34  0.001
## 21 2126.9 13.47  0.000
## 19 2127.0 13.56  0.000
## 10 2127.1 13.74  0.000
## 24 2127.6 14.20  0.000
## 9  2128.6 15.16  0.000
## 23 2128.8 15.40  0.000
## 14 2128.9 15.52  0.000
## 12 2129.0 15.64  0.000
## 56 2129.4 15.98  0.000
## 13 2130.3 16.91  0.000
## 11 2130.5 17.08  0.000
## 55 2130.6 17.22  0.000
## 16 2130.8 17.43  0.000
## 15 2132.2 18.84  0.000
## 48 2132.6 19.20  0.000
## 47 2134.1 20.66  0.000
## 2  2137.4 23.95  0.000
## 1  2138.2 24.83  0.000
## 6  2139.2 25.83  0.000
## 4  2139.3 25.85  0.000
## 5  2140.1 26.69  0.000
## 3  2140.2 26.75  0.000
## 8  2141.1 27.73  0.000
## 7  2142.0 28.61  0.000
## 40 2142.9 29.49  0.000
## 39 2143.8 30.42  0.000
## Models ranked by AICc(x)
## Random terms (all models):
## '1 | School/Intervention.time'
```

```
importance(results4)
```

```
##                       sex  lake ClassNum Intervention.type Intervention.time
```

```
## Sum of weights:      1.00 0.99 0.71     0.32              0.30
## N containing models:   20   20   20       24                24
##                       Intervention.time:Intervention.type
## Sum of weights:      0.03
## N containing models:    8
```

```r
Shp_4<-model.avg(results4, subset= delta <2, revised.var = TRUE)
Shp_4
```

```
##
## Call:
## model.avg(object = results4, subset = delta < 2, revised.var = TRUE)
##
## Component models:
## '145'  '1345' '45'   '1245'
##
## Coefficients:
##        (Intercept)   ClassNum lakeriver      sexM Intervention.typeremoval
## full      2.161458 -0.1022696 -2.480316 0.4404063              -0.05472666
## subset    2.161458 -0.1258076 -2.480316 0.4404063              -0.28997949
##        Intervention.timeafter
## full              -0.01802985
## subset            -0.10394464
```

```r
# Sm prev
results5<-dredge(veg.Sm.prevx)
```

```
## Fixed term is "(Intercept)"
```

```r
results5
```

```
## Global model call: glmer(formula = Sm ~ Intervention.time * Intervention.type +
##     sex + lake + ClassNum + (1 | School/Intervention.time), data = dataFull,
##     family = binomial, control = glmerControl(optimizer = "bobyqa",
##         optCtrl = list(maxfun = 1e+05)), na.action = na.fail)
## ---
## Model selection table
##    (Intrc)       ClN Intrv.tim Intrv.typ lak sex Intrv.tim:Intrv.typ df
## 55  -2.081                   +         +       +                   +  7
## 63  -1.688                   +         +   +   +                   +  8
## 19  -1.738                   +                 +                      5
## 39  -1.977                   +         +                           +  6
## 17  -1.579                                     +                      4
## 27  -1.346                   +             +   +                      6
## 47  -1.589                   +         +   +                       +  7
## 25  -1.179                                 +   +                      5
## 3   -1.631                   +                                        4
## 56  -2.084  0.001634         +         +       +                   +  8
## 1   -1.474                                                            3
## 11  -1.243                   +             +                          5
## 64  -1.682 -0.002861         +         +   +   +                   +  9
## 9   -1.078                                 +                          4
```

```
## 23  -1.865                    +         +       +                   6
## 20  -1.730 -0.004009          +                 +                   6
## 40  -1.992  0.006992          +         +                       + 7
## 21  -1.706                              +       +                   5
## 18  -1.564 -0.007374                            +                   5
## 31  -1.470                    +         +   +   +                   7
## 28  -1.326 -0.008526          +             +   +                   7
## 29  -1.302                              +   +   +                   6
## 26  -1.151 -0.011870                        +   +                   6
## 48  -1.595  0.002593          +         +   +                   + 8
## 7   -1.764                    +         +                           5
## 4   -1.634  0.001330          +                                     5
## 5   -1.607                              +                           4
## 2   -1.470 -0.001907                                                4
## 15  -1.373                    +         +   +                       6
## 12  -1.235 -0.003089          +             +                       6
## 13  -1.208                              +   +                       5
## 10  -1.063 -0.006302                        +                       5
## 24  -1.857 -0.004253          +         +       +                   7
## 22  -1.691 -0.007610                    +       +                   6
## 32  -1.450 -0.008810          +         +   +   +                   8
## 30  -1.275 -0.012140                    +   +   +                   7
## 8   -1.767  0.001064          +         +                           6
## 6   -1.602 -0.002164                    +                           5
## 16  -1.365 -0.003399          +         +   +                       7
## 14  -1.193 -0.006603                    +   +                       6
##       logLik   AICc delta weight
## 55 -923.734 1861.5  0.00  0.093
## 63 -922.929 1861.9  0.40  0.076
## 19 -926.170 1862.4  0.85  0.061
## 39 -925.243 1862.5  1.00  0.056
## 17 -927.318 1862.7  1.13  0.052
## 27 -925.357 1862.8  1.23  0.050
## 47 -924.456 1863.0  1.44  0.045
## 25 -926.479 1863.0  1.47  0.044
## 3  -927.668 1863.4  1.84  0.037
## 56 -923.734 1863.5  2.01  0.034
## 1  -928.807 1863.6  2.11  0.032
## 11 -926.874 1863.8  2.26  0.030
## 64 -922.929 1863.9  2.42  0.028
## 9  -927.987 1864.0  2.47  0.027
## 23 -926.120 1864.3  2.76  0.023
## 20 -926.168 1864.4  2.85  0.022
## 40 -925.238 1864.5  3.01  0.021
## 21 -927.269 1864.6  3.05  0.020
## 18 -927.312 1864.7  3.13  0.019
## 31 -925.305 1864.7  3.14  0.019
## 28 -925.350 1864.7  3.23  0.018
## 29 -926.428 1864.9  3.37  0.017
## 26 -926.464 1865.0  3.45  0.017
## 48 -924.455 1865.0  3.46  0.016
## 7  -927.613 1865.3  3.73  0.014
## 4  -927.668 1865.4  3.84  0.014
## 5  -928.753 1865.5  4.01  0.012
```

```
## 2  -928.806 1865.6  4.11  0.012
## 15 -926.817 1865.7  4.15  0.012
## 12 -926.873 1865.8  4.27  0.011
## 13 -927.930 1865.9  4.37  0.010
## 10 -927.983 1866.0  4.47  0.010
## 24 -926.118 1866.3  4.77  0.009
## 22 -927.263 1866.6  5.04  0.007
## 32 -925.297 1866.7  5.14  0.007
## 30 -926.412 1866.9  5.36  0.006
## 8  -927.613 1867.3  5.75  0.005
## 6  -928.753 1867.5  6.01  0.005
## 16 -926.815 1867.7  6.16  0.004
## 14 -927.925 1867.9  6.37  0.004
## Models ranked by AICc(x)
## Random terms (all models):
## '1 | School/Intervention.time'
```

```
importance(results5)
```

```
##                        Intervention.time sex  Intervention.type lake
## Sum of weights:        0.70              0.62 0.54              0.45
## N containing models:    24                 20   24                20
##                        Intervention.time:Intervention.type ClassNum
## Sum of weights:        0.37                                0.27
## N containing models:    8                                    20
```

```
Smp_5<-model.avg(results5, subset= delta <2, revised.var = TRUE)
Smp_5
```

```
##
## Call:
## model.avg(object = results5, subset = delta < 2, revised.var = TRUE)
##
## Component models:
## '1245'  '12345' '14'    '125'   '4'     '134'   '1235'  '34'    '1'
##
## Coefficients:
##        (Intercept) Intervention.timeafter Intervention.typeremoval      sexM
## full     -1.694958              0.5050754                0.3357967 0.1536181
## subset   -1.694958              0.6226032                0.6408025 0.2100308
##        Intervention.timeafter:Intervention.typeremoval  lakeriver
## full                                        -0.4221684 -0.4313688
## subset                                      -0.8056259 -1.0304271
```

```
# Estimated Marginal Means
# In orginal direction
emSm<-emmeans(veg.Sm.prevx, ~ Intervention.type | Intervention.time, type = "response")
CEmSm<- contrast(emSm, interaction = c(Intervention.time = "poly", Intervention.type = "poly"),by = NUL
CEmSm
```

```
##  Intervention.time_poly Intervention.type_poly odds.ratio    SE  df null
##  linear                 linear                      0.446 0.157 Inf    1
```

```
##  z.ratio p.value
##   -2.295  0.0217
##
## Results are averaged over the levels of: sex, lake
## Tests are performed on the log odds ratio scale
```

```
# Reverse so represents prevention
dataFull$Sm2<-0
dataFull$Sm2[dataFull$Sm=="0"]<-1
table(dataFull$Sm,dataFull$Sm2)
```

```
##
##         0    1
##   0     0 1705
##   1   542    0
```

```
# Sm Prev - In manuscript
veg.Sm.prevx2 <- glmer(Sm2 ~ Intervention.time*Intervention.type +sex+lake+ClassNum+ (1|School/Interven
summary(veg.Sm.prevx2)
```

```
## Generalized linear mixed model fit by maximum likelihood (Laplace
##   Approximation) [glmerMod]
##  Family: binomial  ( logit )
## Formula: Sm2 ~ Intervention.time * Intervention.type + sex + lake + ClassNum +
##     (1 | School/Intervention.time)
##    Data: dataFull
##
##      AIC      BIC   logLik deviance df.resid
##   1863.9   1915.3   -922.9   1845.9     2238
##
## Scaled residuals:
##     Min      1Q  Median      3Q     Max
## -5.2005  0.1735  0.2553  0.4241  2.9272
##
## Random effects:
##  Groups                   Name        Variance Std.Dev.
##  Intervention.time:School (Intercept) 0.09979  0.3159
##  School                   (Intercept) 2.16128  1.4701
## Number of obs: 2247, groups:  Intervention.time:School, 32; School, 16
##
## Fixed effects:
##                                              Estimate Std. Error z value
## (Intercept)                                   1.68157    0.65010   2.587
## Intervention.timeafter                       -0.77994    0.25085  -3.109
## Intervention.typeremoval                     -0.63399    0.77835  -0.815
## sexM                                         -0.21146    0.12026  -1.758
## lakeriver                                     1.02603    0.78522   1.307
## ClassNum                                      0.00286    0.06807   0.042
## Intervention.timeafter:Intervention.typeremoval  0.80781    0.35195   2.295
##                                              Pr(>|z|)
## (Intercept)                                   0.00969 **
## Intervention.timeafter                        0.00188 **
## Intervention.typeremoval                      0.41534
```

```
## sexM                                          0.07869 .
## lakeriver                                     0.19132
## ClassNum                                      0.96649
## Intervention.timeafter:Intervention.typeremoval  0.02172 *
## ---
## Signif. codes:  0 '***' 0.001 '**' 0.01 '*' 0.05 '.' 0.1 ' ' 1
##
## Correlation of Fixed Effects:
##             (Intr) Intrvntn.tm Intrvntn.ty sexM   lakrvr ClssNm
## Intrvntn.tm -0.209
## Intrvntn.ty -0.604  0.158
## sexM        -0.078  0.016      -0.006
## lakeriver   -0.465  0.005       0.000      -0.014
## ClassNum    -0.245  0.056      -0.005      -0.045  0.050
## Intrvnt.:I.  0.141 -0.700      -0.218      -0.010  0.008 -0.030
```

```
emSm2<-emmeans(veg.Sm.prevx2, ~ Intervention.time| Intervention.type, type = "response")
CEmSm2<- contrast(emSm2, interaction = c(Intervention.time = "consec", Intervention.type = "poly"),by =
CEmSm2
```

```
##  Intervention.time_consec Intervention.type_poly odds.ratio    SE  df null
##  after / before           linear                       2.24 0.789 Inf    1
##  z.ratio p.value
##    2.295  0.0217
##
## Results are averaged over the levels of: sex, lake
## Tests are performed on the log odds ratio scale
```

```
# In orginal direction - Sm prev
emSm3<-emmeans(veg.Sm.prevx, ~ Intervention.type| Intervention.time, type = "response")
CEmSm3<- contrast(emSm3, interaction = c(Intervention.time = "consec", Intervention.type = "poly"), typ
plot(CEmSm3)
```

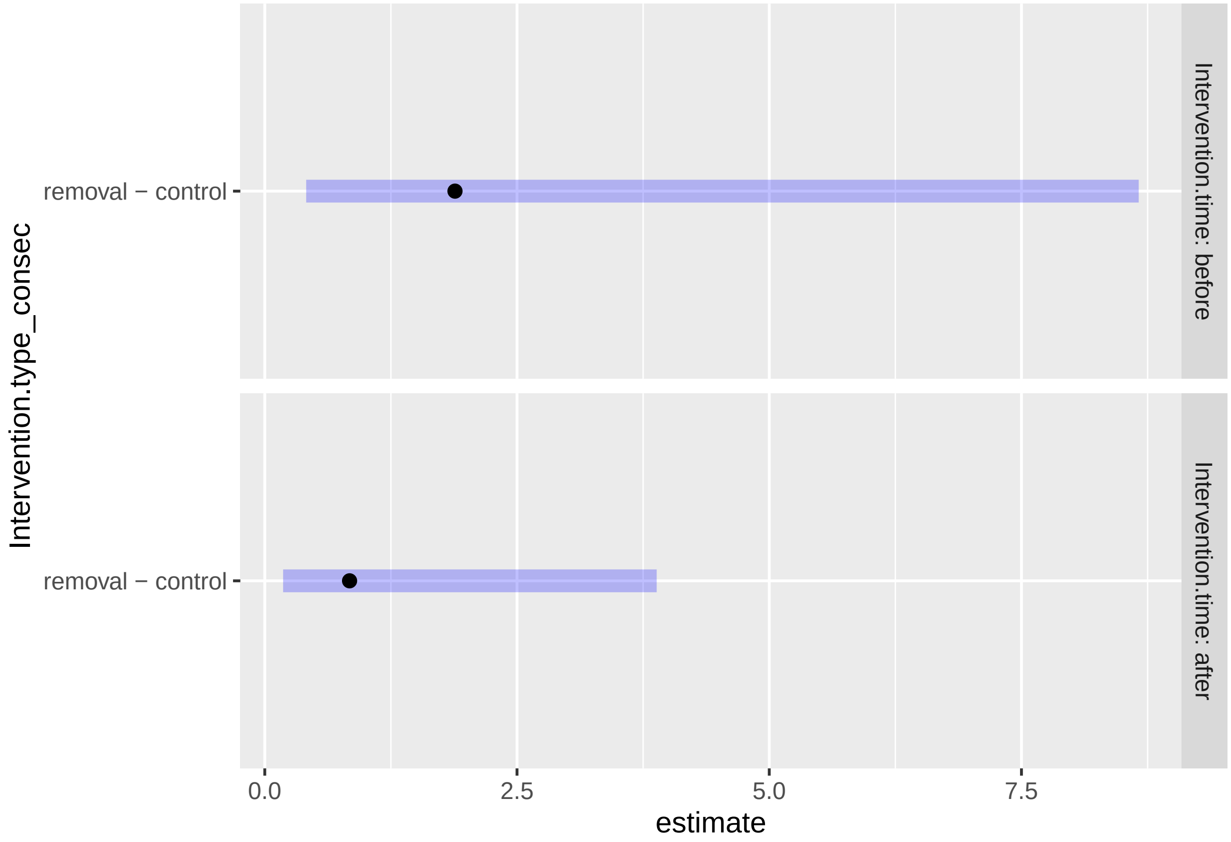


```
CEmSm3
```

```
## Intervention.time = before:
##  Intervention.type_consec odds.ratio    SE  df null z.ratio p.value
##  removal / control              1.89 1.467 Inf    1   0.815  0.4152
##
## Intervention.time = after:
##  Intervention.type_consec odds.ratio    SE  df null z.ratio p.value
##  removal / control              0.84 0.656 Inf    1  -0.223  0.8239
##
## Results are averaged over the levels of: sex, lake
## Tests are performed on the log odds ratio scale
```

```
emSm4<-emmeans(veg.Sm.prevx, ~ Intervention.type* Intervention.time, type = "response")
pwpp(emSm4)
```

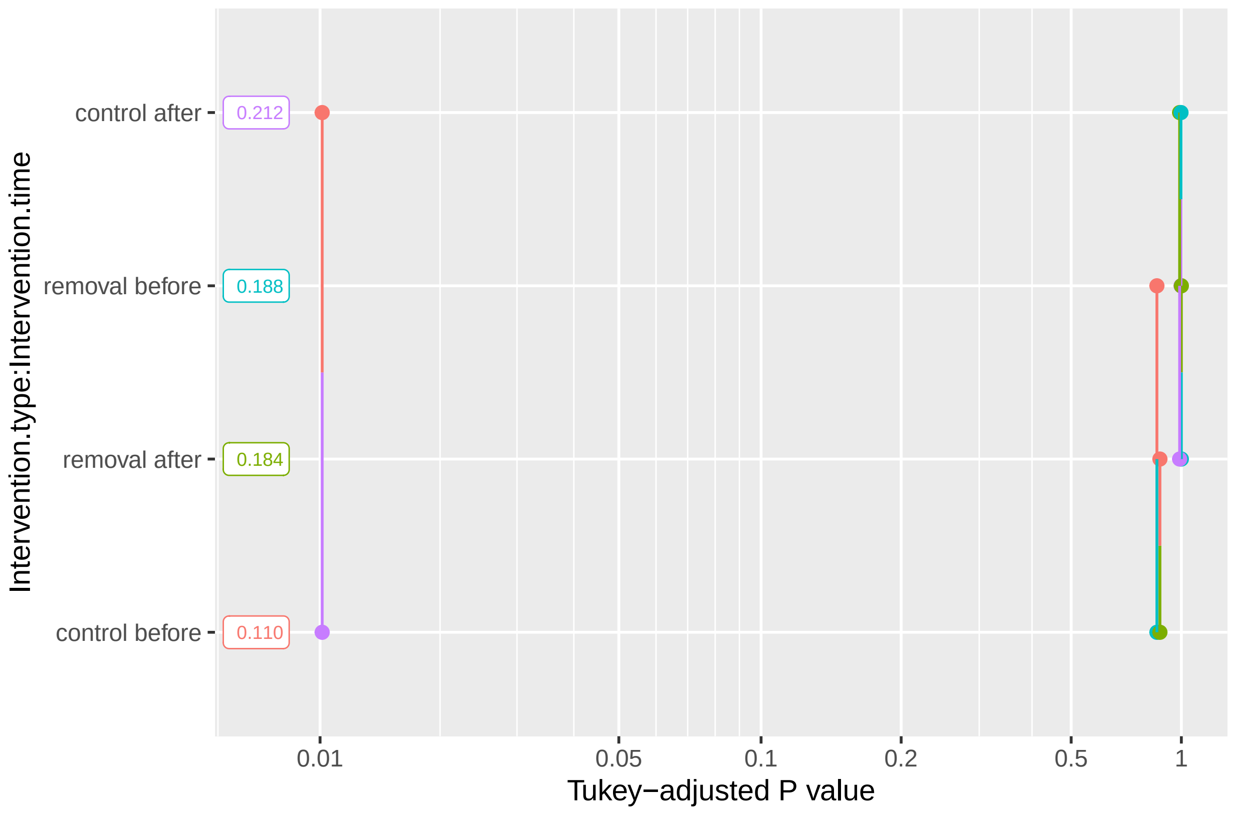


```
# Sh Prev
emmeans(veg.Sh.prevx, ~ Intervention.type | Intervention.time, type = "response")
```

```
## Intervention.time = before:
##  Intervention.type  prob     SE  df asymp.LCL asymp.UCL
##  control           0.763 0.0833 Inf     0.566     0.888
##  removal           0.676 0.0979 Inf     0.465     0.833
##
## Intervention.time = after:
##  Intervention.type  prob     SE  df asymp.LCL asymp.UCL
##  control           0.712 0.0963 Inf     0.496     0.861
##  removal           0.684 0.0983 Inf     0.470     0.841
##
## Results are averaged over the levels of: sex, lake
## Confidence level used: 0.95
## Intervals are back-transformed from the logit scale
```

```
emSh<-emmeans(veg.Sh.prevx, ~ Intervention.type | Intervention.time, type = "response")
CEmSh<- contrast(emSh, interaction = c(Intervention.time = "poly", Intervention.type = "consec"),by = NU
CEmSh
```

```
##  Intervention.time_poly Intervention.type_consec odds.ratio    SE  df null
##  linear                 removal / control              1.36 0.866 Inf    1
##  z.ratio p.value
##    0.480  0.6312
```

```
##
## Results are averaged over the levels of: sex, lake
## Tests are performed on the log odds ratio scale
```

```
# Emeans All Models
emmeans(veg.Sh.intensity, ~ Intervention.type | Intervention.time, type = "response")
```

```
## Intervention.time = before:
##  Intervention.type emmean    SE   df lower.CL upper.CL
##  control             1.87 0.343 22.4     1.16     2.58
##  removal             1.82 0.342 22.3     1.11     2.53
##
## Intervention.time = after:
##  Intervention.type emmean    SE   df lower.CL upper.CL
##  control             1.87 0.345 23.1     1.16     2.58
##  removal             2.02 0.344 22.8     1.31     2.73
##
## Results are averaged over the levels of: sex, lake
## Degrees-of-freedom method: kenward-roger
## Confidence level used: 0.95
```

```
emmeans(veg.Sm.intensity_1, ~ Intervention.type | Intervention.time, type = "response")
```

```
## Intervention.time = before:
##  Intervention.type emmean    SE   df lower.CL upper.CL
##  control            0.651 0.492 14.3  -0.4020     1.70
##  removal            0.995 0.491 14.3  -0.0568     2.05
##
## Intervention.time = after:
##  Intervention.type emmean    SE   df lower.CL upper.CL
##  control            1.124 0.493 14.5   0.0697     2.18
##  removal            1.008 0.493 14.4  -0.0455     2.06
##
## Results are averaged over the levels of: sex, lake
## Degrees-of-freedom method: kenward-roger
## Confidence level used: 0.95
```

```
emmeans(veg.Sh.prevx, ~ Intervention.type | Intervention.time, type = "response")
```

```
## Intervention.time = before:
##  Intervention.type  prob     SE  df asymp.LCL asymp.UCL
##  control           0.763 0.0833 Inf     0.566     0.888
##  removal           0.676 0.0979 Inf     0.465     0.833
##
## Intervention.time = after:
##  Intervention.type  prob     SE  df asymp.LCL asymp.UCL
##  control           0.712 0.0963 Inf     0.496     0.861
##  removal           0.684 0.0983 Inf     0.470     0.841
##
## Results are averaged over the levels of: sex, lake
## Confidence level used: 0.95
## Intervals are back-transformed from the logit scale
```

```r
emmeans(veg.Sm.prevx, ~ Intervention.type | Intervention.time, type = "response")
```

```
## Intervention.time = before:
##  Intervention.type  prob     SE  df asymp.LCL asymp.UCL
##  control           0.110 0.0549 Inf    0.0393     0.270
##  removal           0.188 0.0850 Inf    0.0724     0.408
##
## Intervention.time = after:
##  Intervention.type  prob     SE  df asymp.LCL asymp.UCL
##  control           0.212 0.0938 Inf    0.0819     0.447
##  removal           0.184 0.0843 Inf    0.0699     0.404
##
## Results are averaged over the levels of: sex, lake
## Confidence level used: 0.95
## Intervals are back-transformed from the logit scale
```

```r
# Interaction-style plots for estimated marginal means - all 4 models
emmip(veg.Sm.prevx, Intervention.type ~ Intervention.time, type = "response") + theme_linedraw() +labs(
```

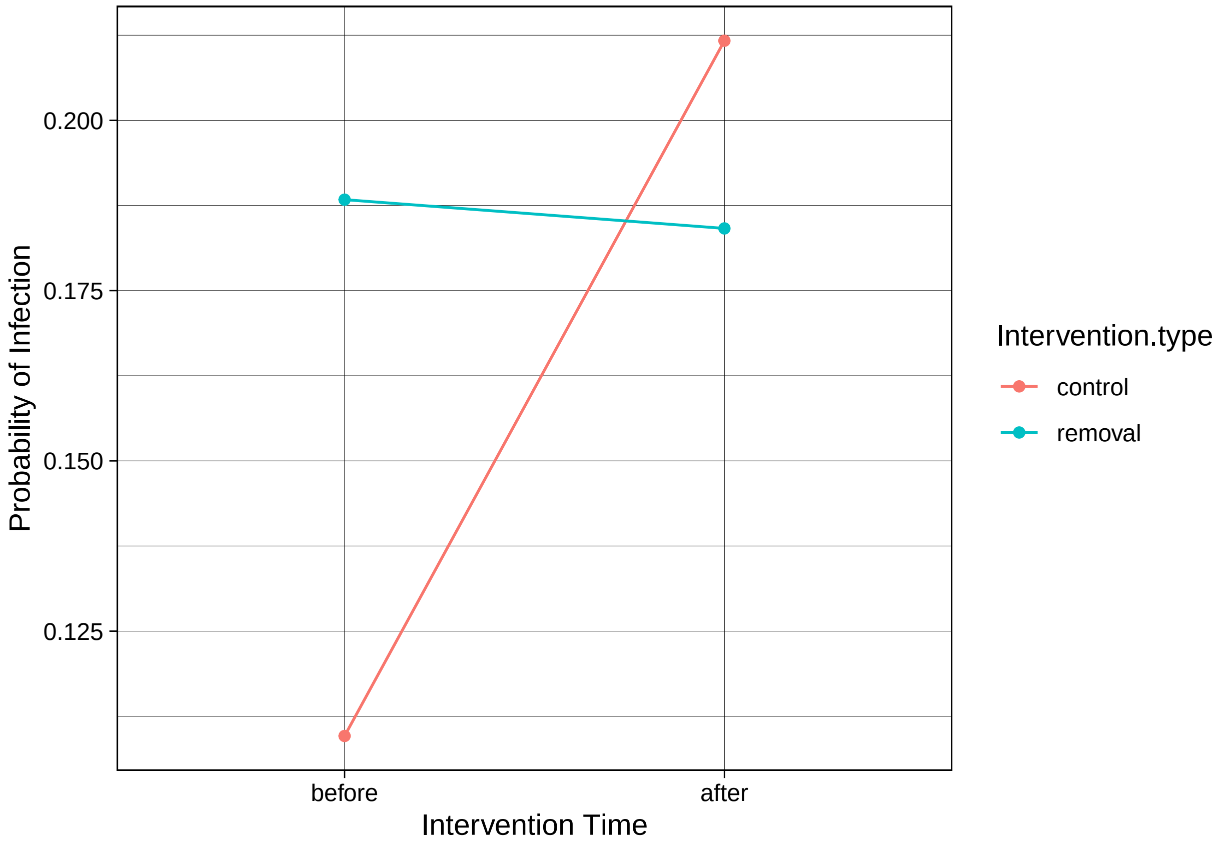


```r
emmip(veg.Sh.prevx, Intervention.type ~ Intervention.time, type = "response") + theme_linedraw() +labs(
```

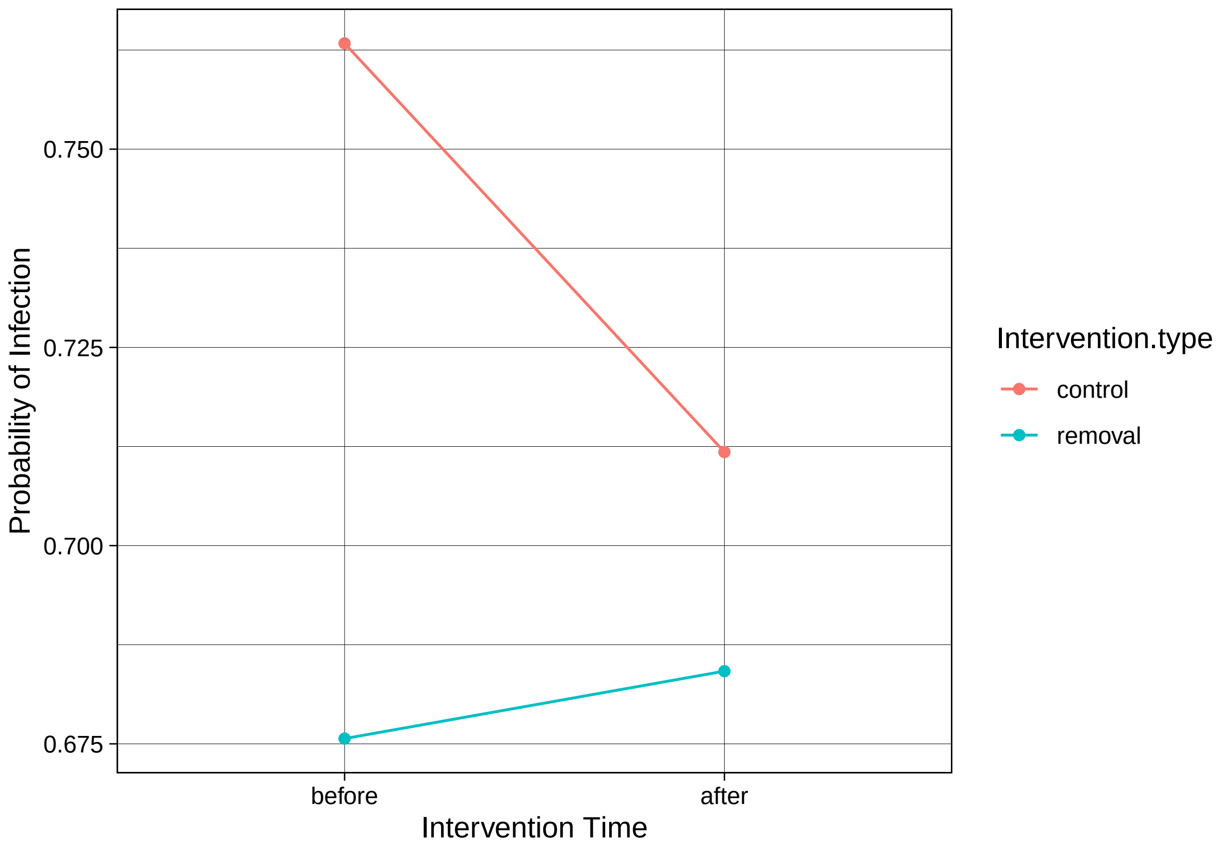


```
emmip(veg.Sm.intensity_1, Intervention.type ~ Intervention.time, type = "response") + theme_linedraw() +
```

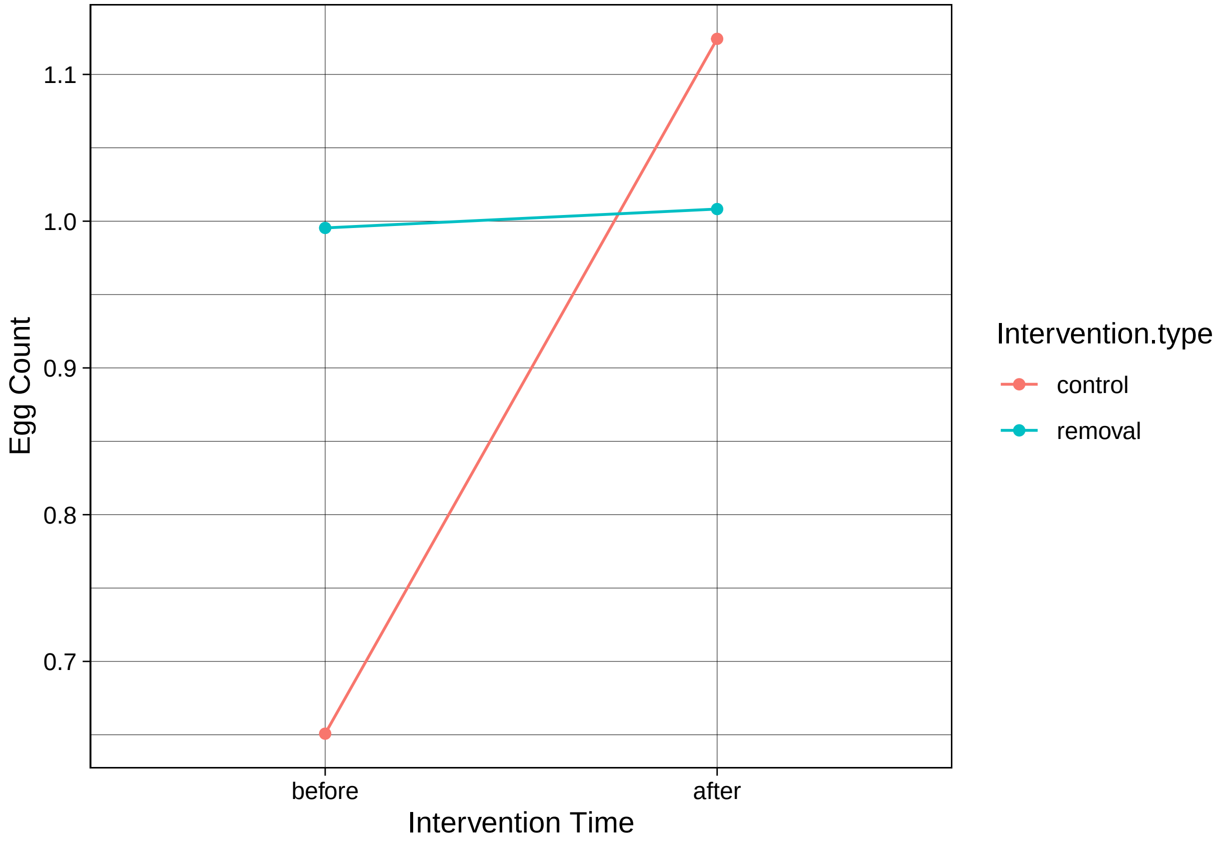


```
emmip(veg.Sh.intensity, Intervention.type ~ Intervention.time, type = "response") + theme_linedraw() +la
```

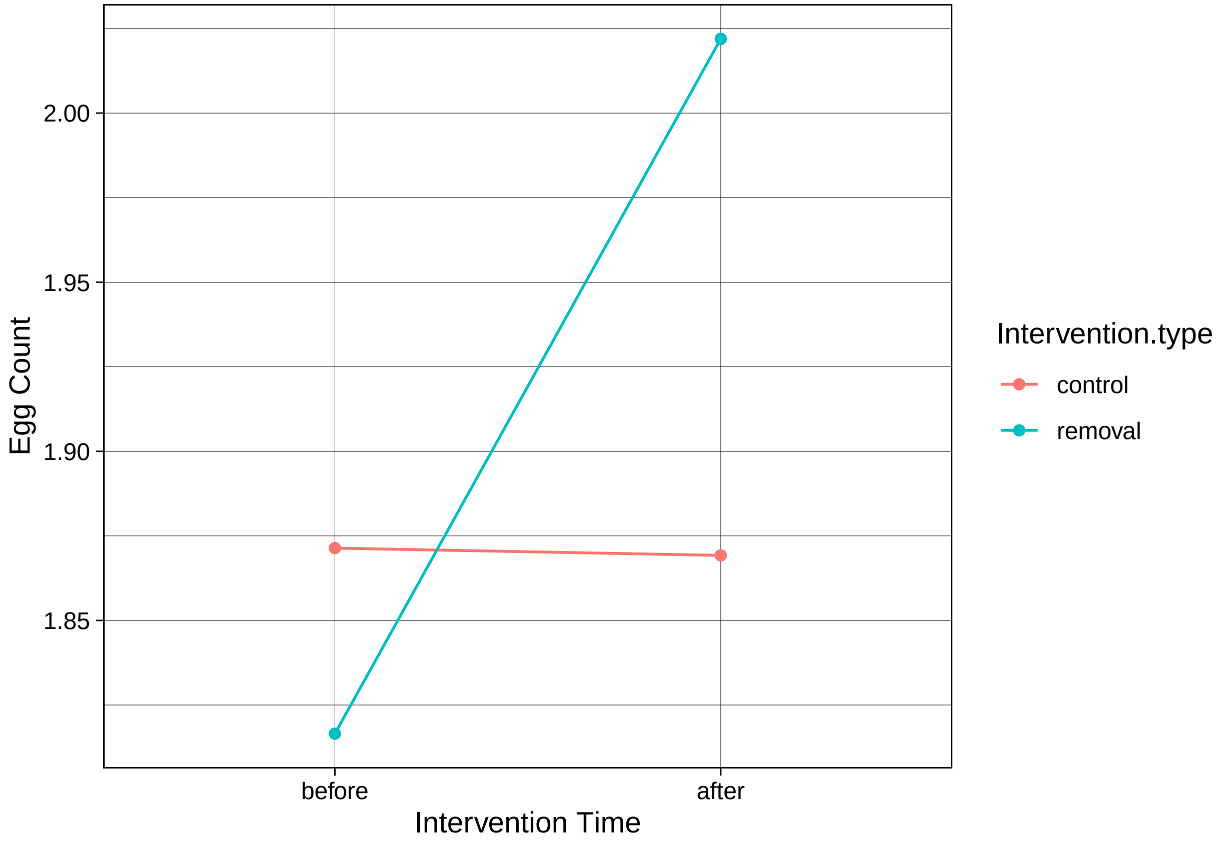


```
# Comparison of Effect
emmip(veg.Sm.prevx, ~ Intervention.time | Intervention.type, CIs = TRUE, type = "response") + theme_line
```

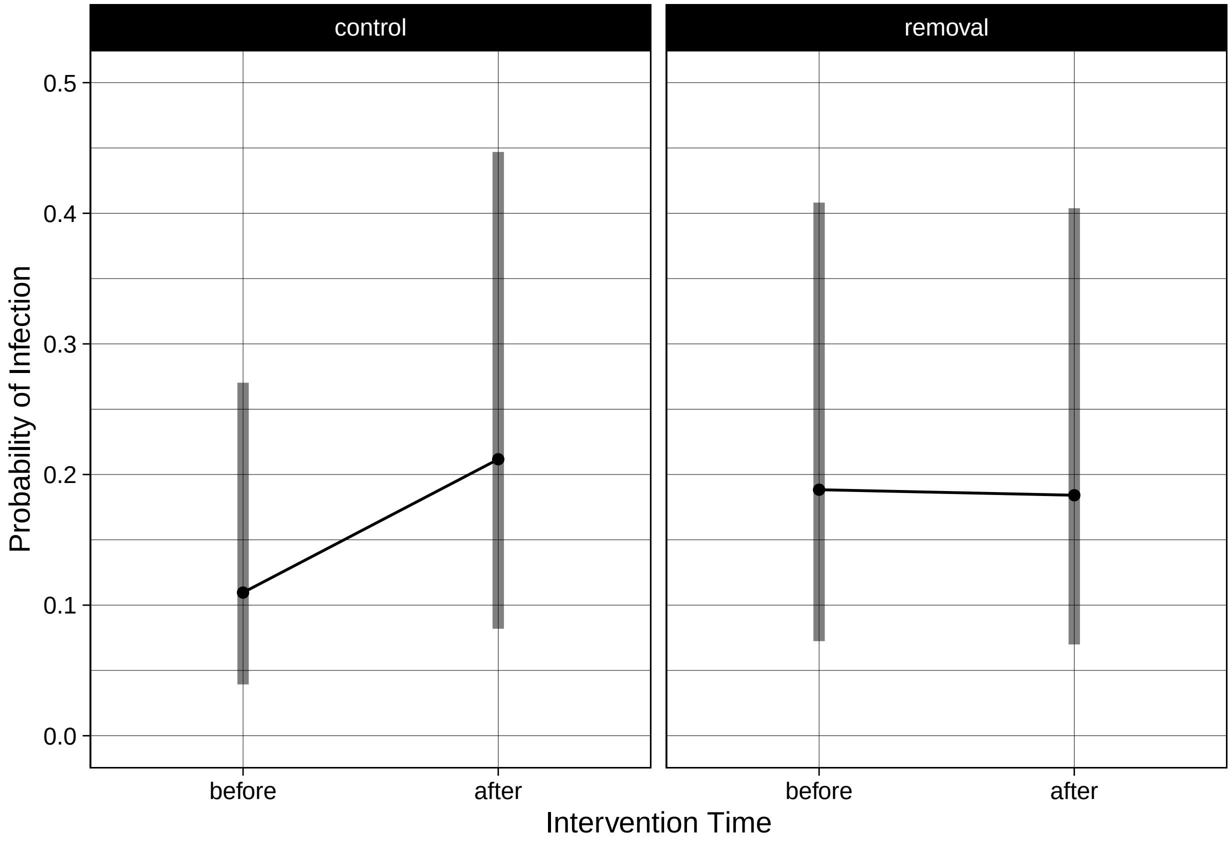


```
emmip(veg.Sh.prevx, ~ Intervention.time | Intervention.type, CIs = TRUE, type = "response") + theme_lin
```

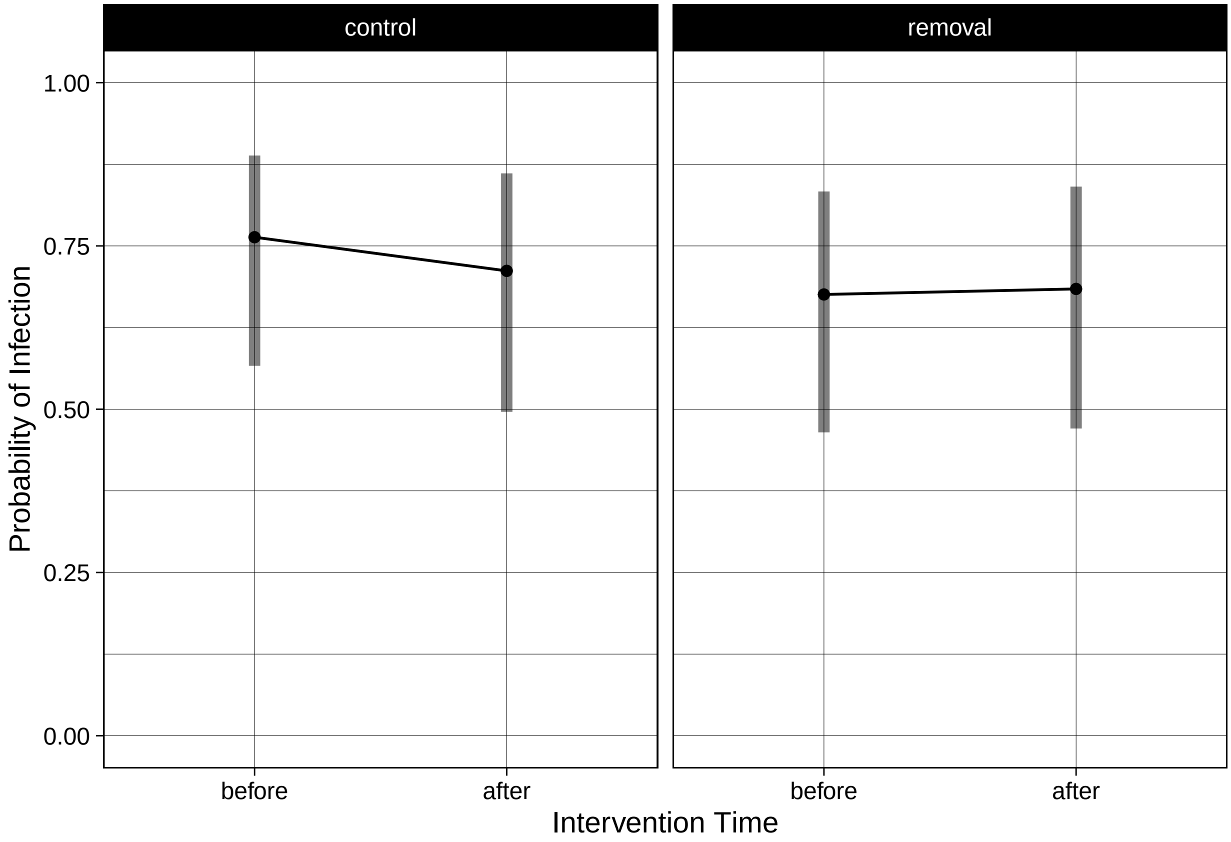


```
emmip(veg.Sh.intensity, ~ Intervention.time | Intervention.type, CIs = TRUE, type = "response") + theme_
```

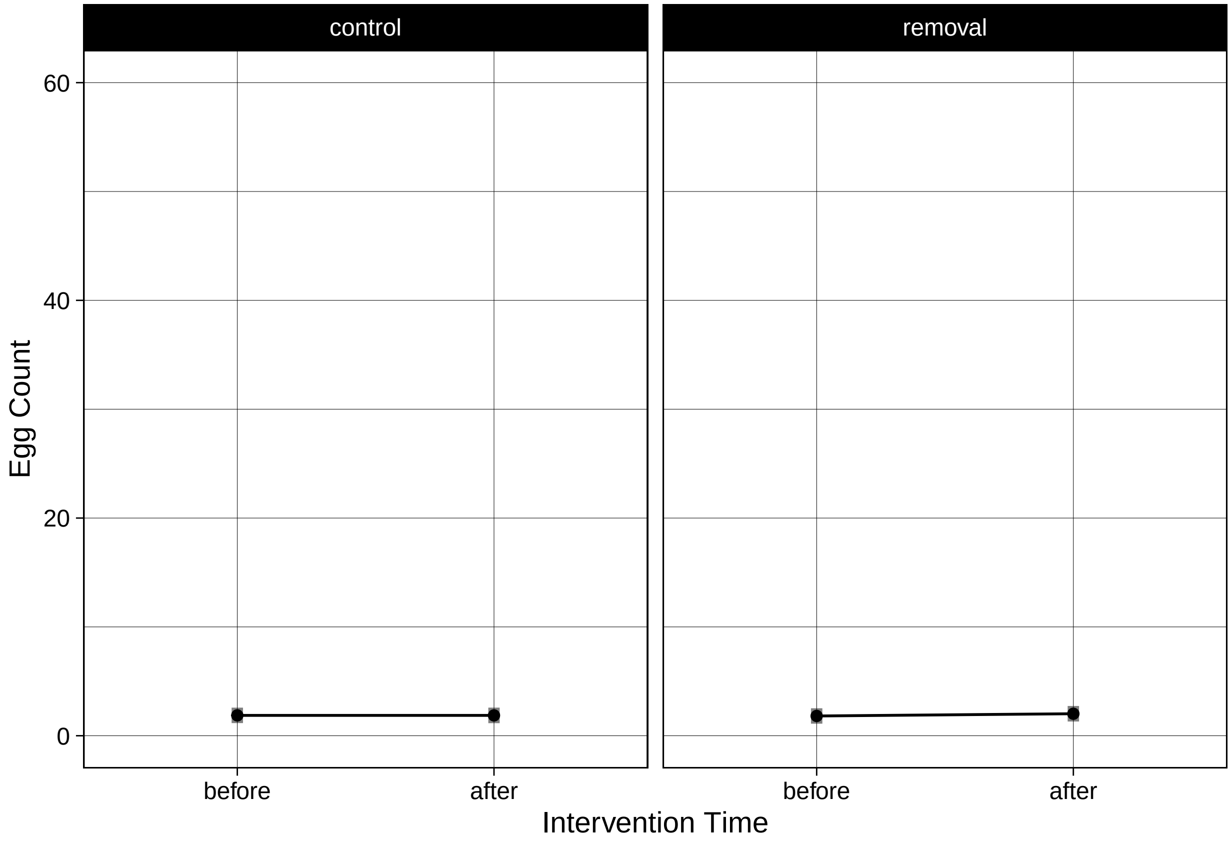


```
emmip(veg.Sm.intensity_1, ~ Intervention.time | Intervention.type, CIs = TRUE, type = "response") + ther
```

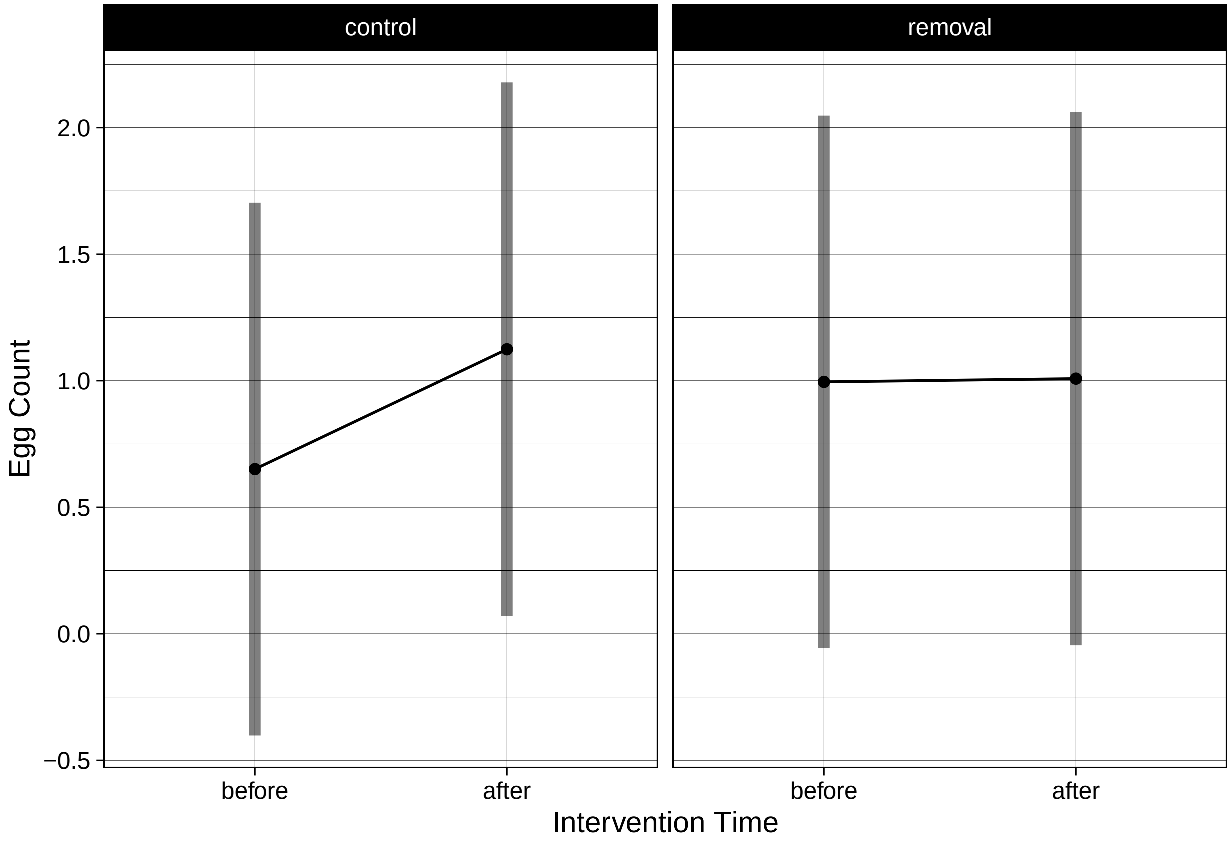


```r
# Total Grids and Graphs for all 4
ref_grid(veg.Sm.prevx) @ grid
```

```
##    Intervention.time Intervention.type sex  lake ClassNum .wgt.
## 1             before           control   F   lac  2.05385   198
## 2              after           control   F   lac  2.05385   152
## 3             before           removal   F   lac  2.05385   179
## 4              after           removal   F   lac  2.05385   149
## 5             before           control   M   lac  2.05385   179
## 6              after           control   M   lac  2.05385   117
## 7             before           removal   M   lac  2.05385   194
## 8              after           removal   M   lac  2.05385   156
## 9             before           control   F river  2.05385   132
## 10             after           control   F river  2.05385    85
## 11            before           removal   F river  2.05385   132
## 12             after           removal   F river  2.05385    85
## 13            before           control   M river  2.05385   127
## 14             after           control   M river  2.05385    94
## 15            before           removal   M river  2.05385   155
## 16             after           removal   M river  2.05385   113
```

```r
emmeans(veg.Sm.prevx, "Intervention.time",type = "response")
```

```
## NOTE: Results may be misleading due to involvement in interactions
```

```
##  Intervention.time  prob     SE  df asymp.LCL asymp.UCL
##  before            0.145 0.0496 Inf    0.0715     0.271
##  after             0.198 0.0640 Inf    0.1004     0.352
##
## Results are averaged over the levels of: Intervention.type, sex, lake
## Confidence level used: 0.95
## Intervals are back-transformed from the logit scale
```

```
emmip(veg.Sm.prevx, Intervention.type ~ Intervention.time, type = "response")
```

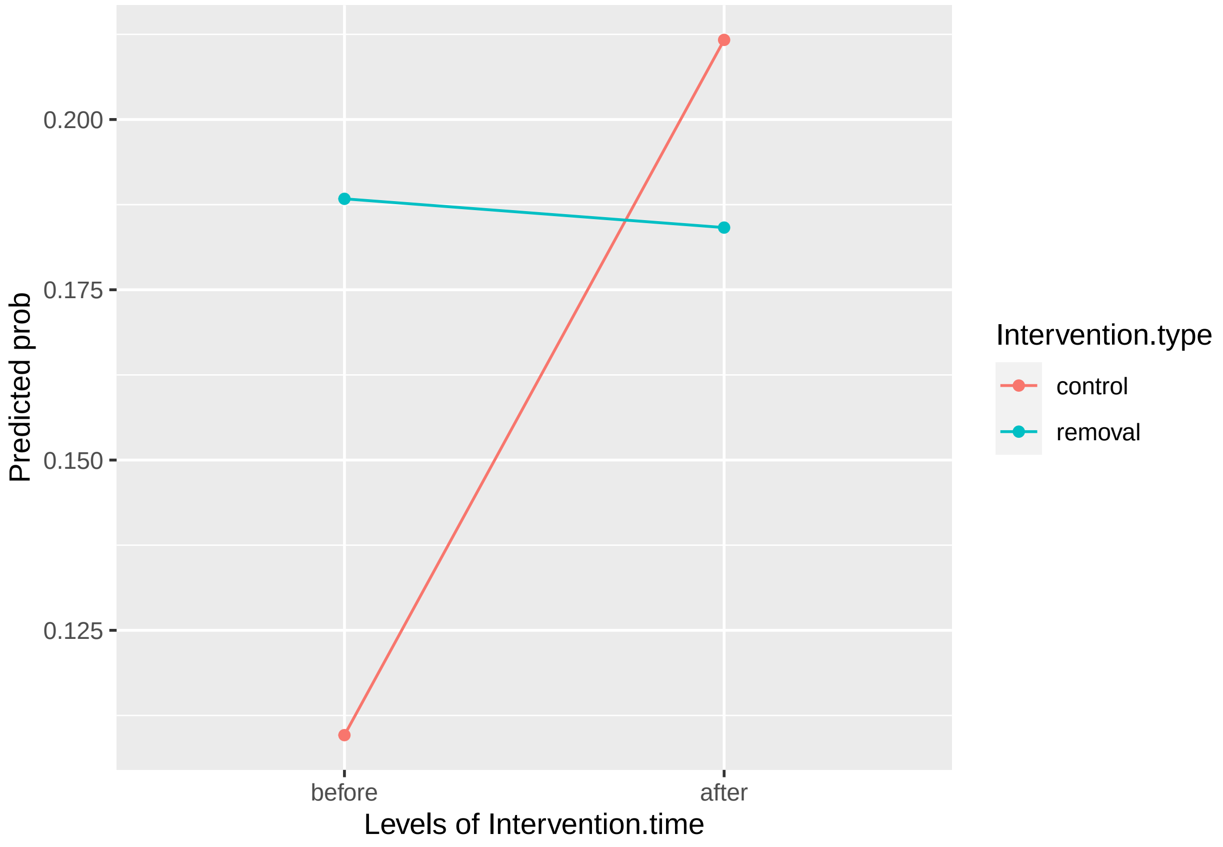


```
ref_grid(veg.Sh.prevx) @ grid
```

```
##    Intervention.time Intervention.type sex  lake ClassNum .wgt.
## 1             before           control   F   lac  2.05385   198
## 2              after           control   F   lac  2.05385   152
## 3             before           removal   F   lac  2.05385   179
## 4              after           removal   F   lac  2.05385   149
## 5             before           control   M   lac  2.05385   179
## 6              after           control   M   lac  2.05385   117
## 7             before           removal   M   lac  2.05385   194
## 8              after           removal   M   lac  2.05385   156
## 9             before           control   F river  2.05385   132
## 10             after           control   F river  2.05385    85
## 11            before           removal   F river  2.05385   132
## 12             after           removal   F river  2.05385    85
```

```
## 13             before           control    M river  2.05385    127
## 14              after           control    M river  2.05385     94
## 15             before           removal    M river  2.05385    155
## 16              after           removal    M river  2.05385    113
```

```r
emmeans(veg.Sh.prevx, "Intervention.time",type = "response")
```

```
## NOTE: Results may be misleading due to involvement in interactions

##  Intervention.time  prob     SE  df asymp.LCL asymp.UCL
##  before            0.722 0.0651 Inf     0.579     0.830
##  after             0.698 0.0695 Inf     0.548     0.815
##
## Results are averaged over the levels of: Intervention.type, sex, lake
## Confidence level used: 0.95
## Intervals are back-transformed from the logit scale
```

```r
emmip(veg.Sh.prevx, Intervention.type ~ Intervention.time, type = "response")
```

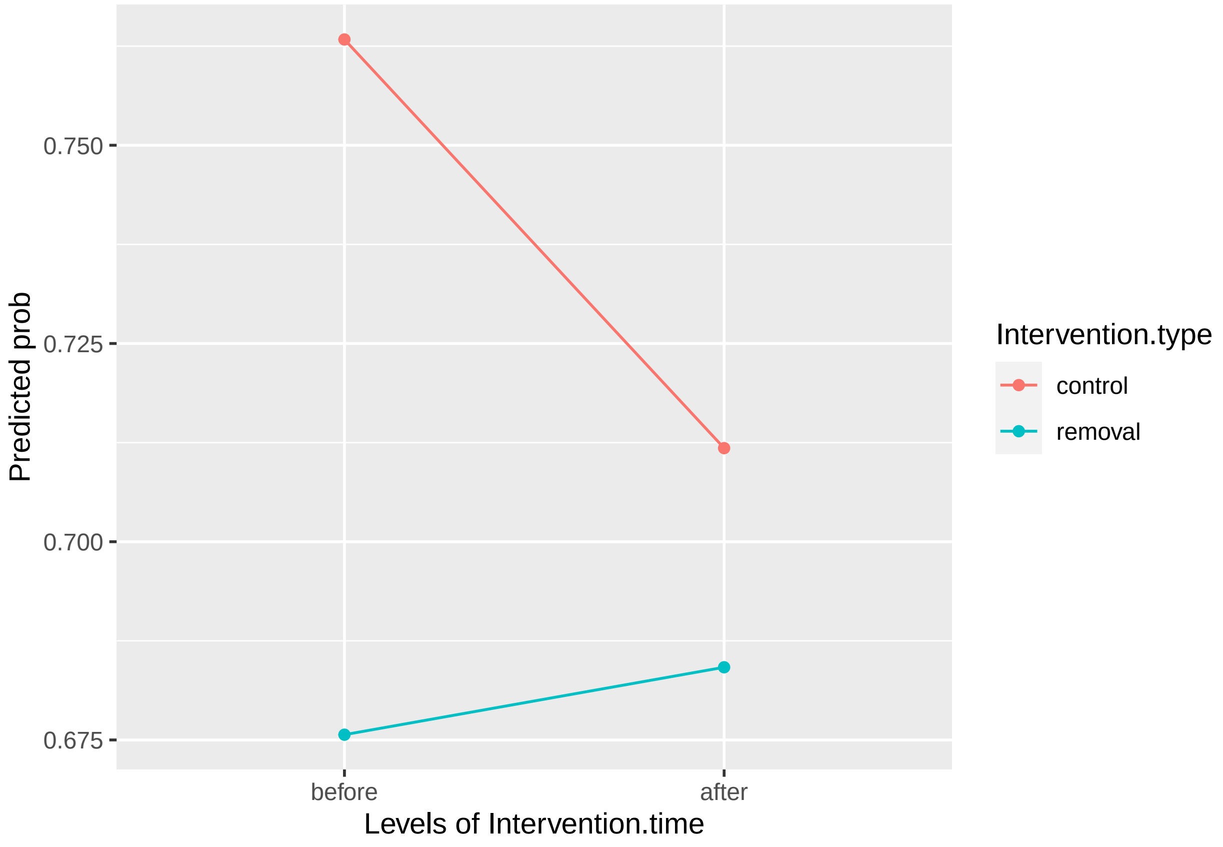


```r
ref_grid(veg.Sm.intensity_1) @ grid
```

```
##    Intervention.type Intervention.time sex  lake ClassNum .wgt.
## 1            control            before   F   lac  2.05385    198
## 2            removal            before   F   lac  2.05385    179
```

```
## 3             control             after   F   lac  2.05385   152
## 4             removal             after   F   lac  2.05385   149
## 5             control            before   M   lac  2.05385   179
## 6             removal            before   M   lac  2.05385   194
## 7             control             after   M   lac  2.05385   117
## 8             removal             after   M   lac  2.05385   156
## 9             control            before   F river  2.05385   132
## 10            removal            before   F river  2.05385   132
## 11            control             after   F river  2.05385    85
## 12            removal             after   F river  2.05385    85
## 13            control            before   M river  2.05385   127
## 14            removal            before   M river  2.05385   155
## 15            control             after   M river  2.05385    94
## 16            removal             after   M river  2.05385   113
```

```
emmeans(veg.Sm.intensity_1, "Intervention.time",type = "response")
```

```
## NOTE: Results may be misleading due to involvement in interactions
```

```
##  Intervention.time emmean    SE   df lower.CL upper.CL
##  before             0.823 0.353 14.2   0.0674     1.58
##  after              1.066 0.354 14.4   0.3094     1.82
##
## Results are averaged over the levels of: Intervention.type, sex, lake
## Degrees-of-freedom method: kenward-roger
## Confidence level used: 0.95
```

```
emmip(veg.Sm.intensity_1, Intervention.type ~ Intervention.time, type = "response")
```

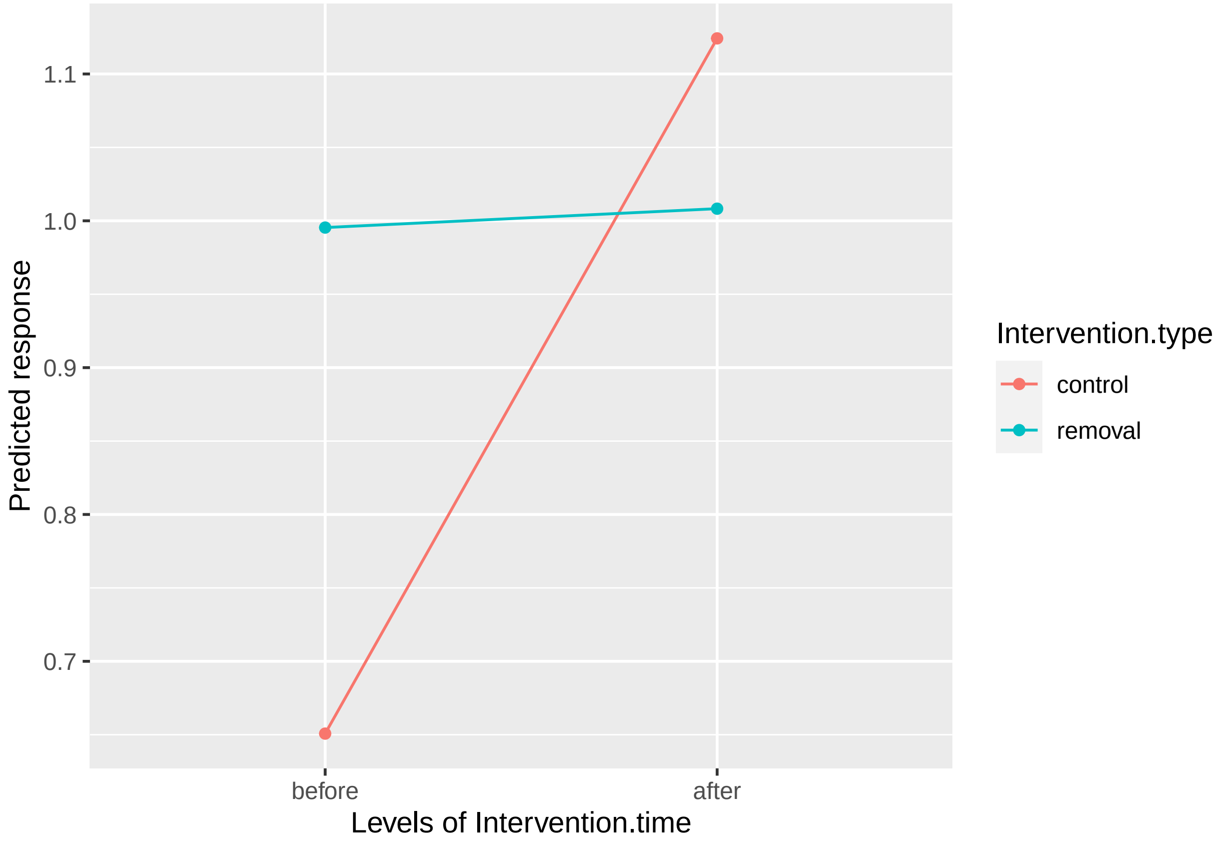


```
ref_grid(veg.Sh.intensity) @ grid
```

```
##    Intervention.type Intervention.time sex  lake ClassNum .wgt.
## 1            control            before   F   lac  2.05385   198
## 2            removal            before   F   lac  2.05385   179
## 3            control             after   F   lac  2.05385   152
## 4            removal             after   F   lac  2.05385   149
## 5            control            before   M   lac  2.05385   179
## 6            removal            before   M   lac  2.05385   194
## 7            control             after   M   lac  2.05385   117
## 8            removal             after   M   lac  2.05385   156
## 9            control            before   F river  2.05385   132
## 10           removal            before   F river  2.05385   132
## 11           control             after   F river  2.05385    85
## 12           removal             after   F river  2.05385    85
## 13           control            before   M river  2.05385   127
## 14           removal            before   M river  2.05385   155
## 15           control             after   M river  2.05385    94
## 16           removal             after   M river  2.05385   113
```

```
emmeans(veg.Sh.intensity, "Intervention.time",type = "response")
```

```
## NOTE: Results may be misleading due to involvement in interactions
```

```
##  Intervention.time emmean    SE   df lower.CL upper.CL
```

```
##   before              1.84 0.245 22.1     1.34     2.35
##   after               1.95 0.246 22.7     1.44     2.46
##
## Results are averaged over the levels of: Intervention.type, sex, lake
## Degrees-of-freedom method: kenward-roger
## Confidence level used: 0.95
```

```
emmip(veg.Sh.intensity, Intervention.type ~ Intervention.time, type = "response")
```

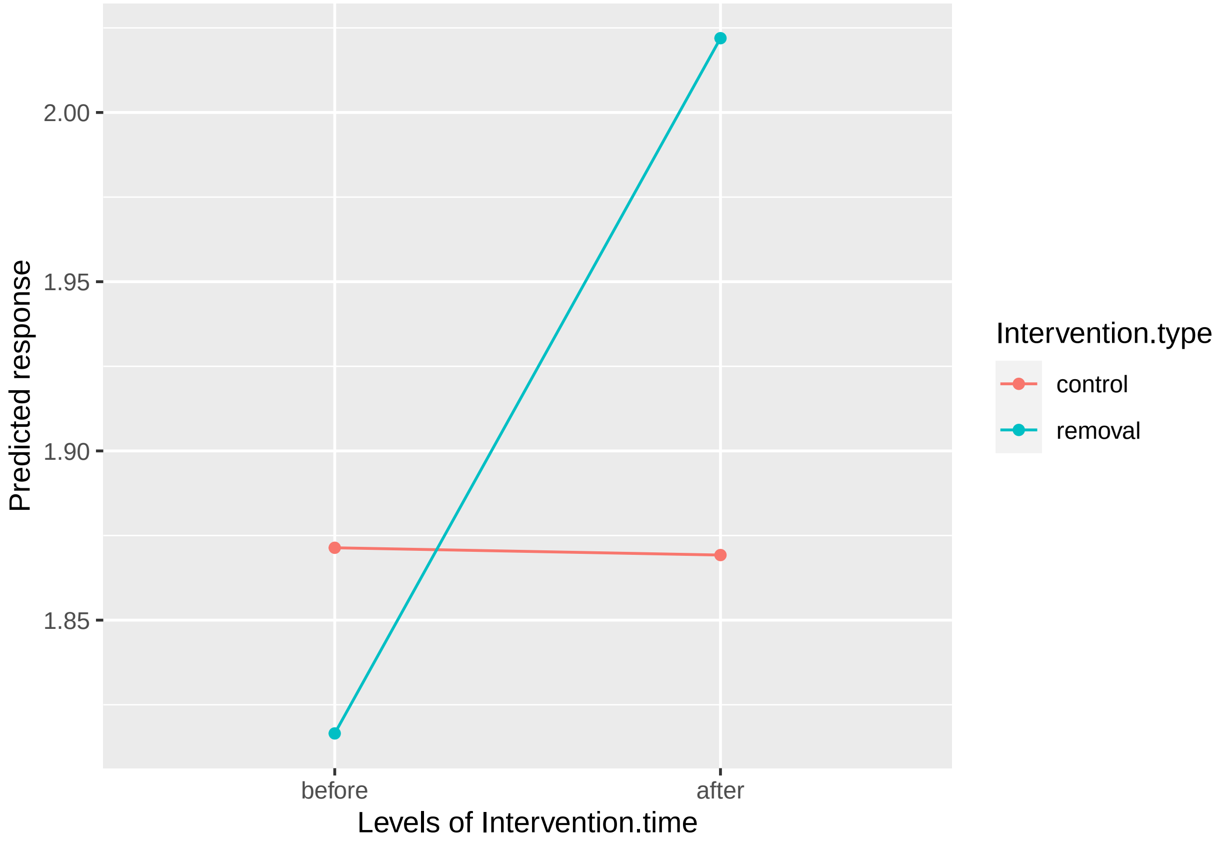


```
# Sm results
r.squaredGLMM(veg.Sm.intensity_1)
```

```
## Warning: 'r.squaredGLMM' now calculates a revised statistic. See the help page.

##             R2m       R2c
## [1,] 0.05238068 0.5138599
```

```
r.squaredGLMM(veg.Sm.prevx)
```

```
## Warning: The null model is correct only if all variables used by the original
## model remain unchanged.

##                    R2m       R2c
## theoretical 0.06014790 0.4429801
## delta       0.04615081 0.3398936
```

```r
# Sh results
r.squaredGLMM(veg.Sh.intensity)
```

```
##            R2m       R2c
## [1,] 0.1591266 0.3786075
```

```r
r.squaredGLMM(veg.Sh.prevx)
```

```
## Warning: The null model is correct only if all variables used by the original
## model remain unchanged.
```

```
##                   R2m       R2c
## theoretical 0.2407245 0.4748768
## delta       0.1979454 0.3904867
```

```r
# Means and confidence intervals for all four
# Egg counts
SHeggC <- dataFull %>%
  group_by(Intervention.type, Intervention.time) %>%
  summarise(
    n=n(),
    mean=mean(ShW),
    sd=sd(ShW),
    median=median(ShW),
  ) %>%
  mutate( se=sd/sqrt(n))  %>%
  mutate( ic=se * qt((1-0.05)/2 + .5, n-1))  %>%
  mutate(Left=mean-qnorm(0.975)*sd/sqrt(n))  %>%
  mutate(Right=mean+qnorm(0.975)*sd/sqrt(n)) %>%
  as.data.frame()
```

```
## `summarise()` has grouped output by 'Intervention.type'. You can override using
## the `.groups` argument.
```

```r
SHeggC
```

```
##   Intervention.type Intervention.time   n     mean        sd median       se
## 1           control            before 636 46.70440 113.80201    3.0 4.512543
## 2           control             after 448 45.16741 108.00301    4.0 5.102663
## 3           removal            before 660 32.59924  71.24371    3.5 2.773158
## 4           removal             after 503 54.05070 135.55539    5.5 6.044116
##          ic     Left    Right
## 1  8.861312 37.85998 55.54882
## 2 10.028188 35.16638 55.16845
## 3  5.445290 27.16395 38.03453
## 4 11.874880 42.20445 65.89695
```

```r
SMeggC <- dataFull %>%
  group_by(Intervention.type, Intervention.time) %>%
  summarise(
```

```
    n=n(),
    mean=mean(SmW),
    sd=sd(SmW),
    median=median(SmW),
  ) %>%
  mutate( se=sd/sqrt(n))  %>%
  mutate( ic=se * qt((1-0.05)/2 + .5, n-1))  %>%
  mutate(Left=mean-qnorm(0.975)*sd/sqrt(n))  %>%
  mutate(Right=mean+qnorm(0.975)*sd/sqrt(n)) %>%
  as.data.frame()
```

```
## `summarise()` has grouped output by 'Intervention.type'. You can override using
## the `.groups` argument.
```

```
SMeggC
```

```
##   Intervention.type Intervention.time   n     mean       sd median        se
## 1           control            before 636 32.50708 224.3736      0  8.896991
## 2           control             after 448 64.77009 350.1838      0 16.544631
## 3           removal            before 660 37.08750 151.1816      0  5.884737
## 4           removal             after 503 55.14911 264.5244      0 11.794560
##         ic     Left    Right
## 1 17.47108 15.06929 49.94486
## 2 32.51492 32.34321 97.19697
## 3 11.55509 25.55363 48.62137
## 4 23.17278 32.03219 78.26602
```

```
# Prev
SMprev <- dataFull %>%
  group_by(Intervention.type, Intervention.time) %>%
  summarise(
    n=n(),
    mean=mean(Sm),
    sd=sd(Sm),
    median=median(Sm),
  ) %>%
  mutate( se=sd/sqrt(n))  %>%
  mutate( ic=se * qt((1-0.05)/2 + .5, n-1))  %>%
  mutate(Left=mean-qnorm(0.975)*sd/sqrt(n))  %>%
  mutate(Right=mean+qnorm(0.975)*sd/sqrt(n)) %>%
  as.data.frame()
```

```
## `summarise()` has grouped output by 'Intervention.type'. You can override using
## the `.groups` argument.
```

```
SMprev
```

```
##   Intervention.type Intervention.time   n      mean        sd median         se
## 1           control            before 636 0.1933962 0.3952718      0 0.01567355
## 2           control             after 448 0.3147321 0.4649283      0 0.02196580
## 3           removal            before 660 0.2424242 0.4288746      0 0.01669392
```

```
## 4           removal             after 503 0.2345924 0.4241657      0 0.01891261
##            ic      Left     Right
## 1 0.03077825 0.1626766 0.2241158
## 2 0.04316906 0.2716800 0.3577843
## 3 0.03277969 0.2097048 0.2751437
## 4 0.03715763 0.1975244 0.2716605
```

```
SHprev <- dataFull %>%
  group_by(Intervention.type, Intervention.time) %>%
  summarise(
    n=n(),
    mean=mean(Sh),
    sd=sd(Sh),
    median=median(Sh),
  ) %>%
  mutate( se=sd/sqrt(n))  %>%
  mutate( ic=se * qt((1-0.05)/2 + .5, n-1))  %>%
  mutate(Left=mean-qnorm(0.975)*sd/sqrt(n))  %>%
  mutate(Right=mean+qnorm(0.975)*sd/sqrt(n)) %>%
  as.data.frame()
```

```
## `summarise()` has grouped output by 'Intervention.type'. You can override using
## the `.groups` argument.
```

```
SHprev
```

```
##   Intervention.type Intervention.time   n      mean        sd median         se
## 1           control            before 636 0.6839623 0.4652938      1 0.01845010
## 2           control             after 448 0.6562500 0.4754899      1 0.02246478
## 3           removal            before 660 0.6803030 0.4667128      1 0.01816677
## 4           removal             after 503 0.7236581 0.4476331      1 0.01995898
##            ic      Left     Right
## 1 0.03623058 0.6478007 0.7201238
## 2 0.04414971 0.6122198 0.7002802
## 3 0.03567173 0.6446968 0.7159092
## 4 0.03921342 0.6845392 0.7627769
```

```
#Compare before and after
# Prevalance
Shtime <- dataFull %>%
  group_by(Intervention.time) %>%
  summarise(
    n=n(),
    mean=mean(Sh),
    sd=sd(Sh),
    median=median(Sh),
  ) %>%
  mutate( se=sd/sqrt(n))  %>%
  mutate( ic=se * qt((1-0.05)/2 + .5, n-1))  %>%
  mutate(Left=mean-qnorm(0.975)*sd/sqrt(n))  %>%
  mutate(Right=mean+qnorm(0.975)*sd/sqrt(n)) %>%
  as.data.frame()
Shtime
```

```
##   Intervention.time    n      mean        sd median         se         ic
## 1            before 1296 0.6820988 0.4658406      1 0.01294002 0.02538569
## 2             after  951 0.6919033 0.4619497      1 0.01497975 0.02939722
##        Left     Right
## 1 0.6567368 0.7074607
## 2 0.6625435 0.7212630
```

```
Smtime <- dataFull %>%
  group_by(Intervention.time) %>%
  summarise(
    n=n(),
    mean=mean(Sm),
    sd=sd(Sm),
    median=median(Sm),
  ) %>%
  mutate( se=sd/sqrt(n))  %>%
  mutate( ic=se * qt((1-0.05)/2 + .5, n-1))  %>%
  mutate(Left=mean-qnorm(0.975)*sd/sqrt(n))  %>%
  mutate(Right=mean+qnorm(0.975)*sd/sqrt(n)) %>%
  as.data.frame()
Smtime
```

```
##   Intervention.time    n      mean        sd median         se         ic
## 1            before 1296 0.2183642 0.4132954      0 0.01148043 0.02252227
## 2             after  951 0.2723449 0.4454007      0 0.01444311 0.02834408
##        Left     Right
## 1 0.1958630 0.2408654
## 2 0.2440369 0.3006529
```

```
#Egg count
SHeggCTime <- dataFull %>%
  group_by(Intervention.time) %>%
  summarise(
    n=n(),
    mean=mean(ShW),
    sd=sd(ShW),
    median=median(ShW),
  ) %>%
  mutate( se=sd/sqrt(n))  %>%
  mutate( ic=se * qt((1-0.05)/2 + .5, n-1))  %>%
  mutate(Left=mean-qnorm(0.975)*sd/sqrt(n))  %>%
  mutate(Right=mean+qnorm(0.975)*sd/sqrt(n)) %>%
  as.data.frame()
SHeggCTime
```

```
##   Intervention.time    n     mean        sd median       se       ic     Left
## 1            before 1296 39.52122  94.77926    3.5 2.632757 5.164936 34.36111
## 2             after  951 49.86593 123.36160    5.0 4.000274 7.850395 42.02554
##      Right
## 1 44.68133
## 2 57.70632
```

```r
SMeggCTime  <- dataFull %>%
  group_by(Intervention.time) %>%
  summarise(
    n=n(),
    mean=mean(SmW),
    sd=sd(SmW),
    median=median(SmW),
  ) %>%
  mutate( se=sd/sqrt(n))  %>%
  mutate( ic=se * qt((1-0.05)/2 + .5, n-1))  %>%
  mutate(Left=mean-qnorm(0.975)*sd/sqrt(n))  %>%
  mutate(Right=mean+qnorm(0.975)*sd/sqrt(n)) %>%
  as.data.frame()
SMeggCTime
```

```
##   Intervention.time    n     mean       sd median       se       ic     Left
## 1            before 1296 34.83970 190.5832      0 5.293978 10.38571 24.46369
## 2             after  951 59.68139 307.7311      0 9.978864 19.58316 40.12317
##      Right
## 1 45.21571
## 2 79.23960
```

```r
# This section looks at the effect of censoring that occurred on one batch and determines that even wit

# Tobit Models for egg count, censored at 200 egg count - Testing the effect of censoring
Th <- vglm(LogShW ~ Intervention.time*Intervention.type +sex+lake+ClassNum, tobit(Upper = 5.298), data =
summary(Th)
```

```
##
## Call:
## vglm(formula = LogShW ~ Intervention.time * Intervention.type +
##     sex + lake + ClassNum, family = tobit(Upper = 5.298), data = dataFull,
##     na.action = na.omit)
##
## Coefficients:
##                                            Estimate Std. Error z value
## (Intercept):1                               1.20391    0.07689  15.657
## (Intercept):2                               0.06236    0.01878   3.321
## Intervention.timeafter                     -0.02896    0.06895  -0.420
## Intervention.typeremoval                   -0.02681    0.06213  -0.432
## sexM                                        0.18593    0.04718   3.941
## lakeriver                                  -1.00017    0.05062 -19.758
## ClassNum                                   -0.12051    0.02458  -4.903
## Intervention.timeafter:Intervention.typeremoval  0.15123    0.09523   1.588
##                                            Pr(>|z|)
## (Intercept):1                               < 2e-16 ***
## (Intercept):2                              0.000898 ***
## Intervention.timeafter                     0.674432
## Intervention.typeremoval                   0.666045
## sexM                                       8.11e-05 ***
## lakeriver                                   < 2e-16 ***
## ClassNum                                   9.46e-07 ***
```

```
## Intervention.timeafter:Intervention.typeremoval 0.112277
## ---
## Signif. codes:  0 '***' 0.001 '**' 0.01 '*' 0.05 '.' 0.1 ' ' 1
##
## Names of linear predictors: mu, loglink(sd)
##
## Log-likelihood: -2864.49 on 4486 degrees of freedom
##
## Number of Fisher scoring iterations: 6
##
## No Hauck-Donner effect found in any of the estimates
```

```
Anova(Th)
```

```
## Analysis of Deviance Table (Type II tests)
##
## Response: LogShW
##                                     Df    Chisq Pr(>Chisq)
## Intervention.time                    1   1.1093     0.2922
## Intervention.type                    1   0.6311     0.4269
## sex                                  1  15.5325  8.110e-05 ***
## lake                                 1 390.3668  < 2.2e-16 ***
## ClassNum                             1  24.0346  9.462e-07 ***
## Intervention.time:Intervention.type  1   2.5219     0.1123
## ---
## Signif. codes:  0 '***' 0.001 '**' 0.01 '*' 0.05 '.' 0.1 ' ' 1
```

```
Tm <- vglm(LogSmW ~ Intervention.time*Intervention.type + sex+lake+ClassNum, tobit(Upper = 5.298), data
summary(Tm)
```

```
##
## Call:
## vglm(formula = LogSmW ~ Intervention.time * Intervention.type +
##     sex + lake + ClassNum, family = tobit(Upper = 5.298), data = dataFull,
##     na.action = na.omit)
##
## Coefficients:
##                                                  Estimate Std. Error z value
## (Intercept):1                                    -1.38086    0.22381  -6.170
## (Intercept):2                                     0.85997    0.03400  25.292
## Intervention.timeafter                            0.87021    0.18900   4.604
## Intervention.typeremoval                          0.38835    0.17596   2.207
## sexM                                              0.24441    0.13052   1.873
## lakeriver                                        -1.18318    0.14505  -8.157
## ClassNum                                         -0.11491    0.06757  -1.701
## Intervention.timeafter:Intervention.typeremoval -0.96581    0.26281  -3.675
##                                                  Pr(>|z|)
## (Intercept):1                                    6.84e-10 ***
## (Intercept):2                                     < 2e-16 ***
## Intervention.timeafter                           4.14e-06 ***
## Intervention.typeremoval                         0.027312 *
## sexM                                             0.061122 .
## lakeriver                                        3.43e-16 ***
```

```
## ClassNum                                         0.089026 .
## Intervention.timeafter:Intervention.typeremoval 0.000238 ***
## ---
## Signif. codes:  0 '***' 0.001 '**' 0.01 '*' 0.05 '.' 0.1 ' ' 1
##
## Names of linear predictors: mu, loglink(sd)
##
## Log-likelihood: -1948.835 on 4486 degrees of freedom
##
## Number of Fisher scoring iterations: 9
##
## No Hauck-Donner effect found in any of the estimates
```

```
Anova(Tm)
```

```
## Analysis of Deviance Table (Type II tests)
##
## Response: LogSmW
##                                    Df   Chisq Pr(>Chisq)
## Intervention.time                   1  7.9654  0.0047681 **
## Intervention.type                   1  0.1261  0.7224821
## sex                                 1  3.5067  0.0611216 .
## lake                                1 66.5399  3.429e-16 ***
## ClassNum                            1  2.8919  0.0890255 .
## Intervention.time:Intervention.type  1 13.5056  0.0002379 ***
## ---
## Signif. codes:  0 '***' 0.001 '**' 0.01 '*' 0.05 '.' 0.1 ' ' 1
```

```
# Sensitivity analysis - 300, 400 and 800
data300<-datanew2
data300$ShW[data300$ShW=="200"]<-300
data400<-datanew2
data400$ShW[data400$ShW=="200"]<-400
data800<-datanew2
data800$ShW[data800$ShW=="200"]<-800
data300c<-datanew2
data300c$ShW[data300c$ShW=="200"& data300$Intervention.type=="control" ]<-300
data400c<-datanew2
data400c$ShW[data400c$ShW=="200"& data400$Intervention.type=="control" ]<-400
data800c<-datanew2
data800c$ShW[data800c$ShW=="200"& data800$Intervention.type=="control" ]<-800


data300t<-datanew2
data300t$ShW[data300t$ShW=="200"& data300t$Intervention.type=="removal" ]<-300
data400t<-datanew2
data400t$ShW[data400t$ShW=="200"& data400t$Intervention.type=="removal" ]<-400
data800t<-datanew2
data800t$ShW[data800t$ShW=="200"& data800t$Intervention.type=="removal" ]<-800

#ALL
veg.Sh.300 = glmer.nb(round(ShW) ~ Intervention.type*Intervention.time + sex+lake+ClassNum+(1|School/In
summary(veg.Sh.300)
```

```
## Generalized linear mixed model fit by maximum likelihood (Laplace
##   Approximation) [glmerMod]
##  Family: Negative Binomial(0.2327)  ( log )
## Formula: round(ShW) ~ Intervention.type * Intervention.time + sex + lake +
##     ClassNum + (1 | School/Intervention.time)
##    Data: data300
##
##      AIC      BIC   logLik deviance df.resid
##  16838.8  16901.8  -8408.4  16816.8     2259
##
## Scaled residuals:
##     Min      1Q  Median      3Q     Max
## -0.4821 -0.4754 -0.4145 -0.0097 19.2005
##
## Random effects:
##  Groups                   Name        Variance Std.Dev.
##  Intervention.time:School (Intercept) 0.6598   0.8123
##  School                   (Intercept) 0.4025   0.6344
## Number of obs: 2270, groups:  Intervention.time:School, 32; School, 16
##
## Fixed effects:
##                                             Estimate Std. Error z value
## (Intercept)                                  4.90317    0.67368   7.278
## Intervention.typeremoval                    -0.04348    0.53031  -0.082
## Intervention.timeafter                      -0.21727    0.42935  -0.506
## sexF                                        -0.53396    0.52596  -1.015
## sexM                                        -0.26066    0.52959  -0.492
## lakeriver                                   -1.51163    0.45340  -3.334
## ClassNum                                    -0.32067    0.05213  -6.152
## Intervention.typeremoval:Intervention.timeafter  0.58286    0.60469   0.964
##                                             Pr(>|z|)
## (Intercept)                                 3.38e-13 ***
## Intervention.typeremoval                    0.934655
## Intervention.timeafter                      0.612829
## sexF                                        0.310001
## sexM                                        0.622584
## lakeriver                                   0.000856 ***
## ClassNum                                    7.67e-10 ***
## Intervention.typeremoval:Intervention.timeafter 0.335102
## ---
## Signif. codes:  0 '***' 0.001 '**' 0.01 '*' 0.05 '.' 0.1 ' ' 1
##
## Correlation of Fixed Effects:
##             (Intr) Intrvntn.ty Intrvntn.tm sexF   sexM   lakrvr ClssNm
## Intrvntn.ty -0.392
## Intrvntn.tm -0.342  0.397
## sexF        -0.768 -0.004       0.030
## sexM        -0.762 -0.009       0.030       0.984
## lakeriver   -0.248  0.002       0.006      -0.018 -0.025
## ClassNum    -0.177  0.014       0.026      -0.006 -0.020  0.055
## Intrvnt.:I.  0.240 -0.562      -0.709      -0.016 -0.016 -0.003 -0.030
```

```
Anova(veg.Sh.300)
```

```
## Analysis of Deviance Table (Type II Wald chisquare tests)
##
## Response: round(ShW)
##                                        Chisq Df Pr(>Chisq)
## Intervention.type                     0.3084  1  0.5786509
## Intervention.time                     0.0635  1  0.8010084
## sex                                   9.2646  2  0.0097323 **
## lake                                 11.1155  1  0.0008561 ***
## ClassNum                             37.8420  1  7.671e-10 ***
## Intervention.type:Intervention.time   0.9291  1  0.3351017
## ---
## Signif. codes:  0 '***' 0.001 '**' 0.01 '*' 0.05 '.' 0.1 ' ' 1
```

```
veg.Sh.400 = glmer.nb(round(ShW) ~ Intervention.type*Intervention.time + sex+lake+ClassNum+(1|School/In
summary(veg.Sh.400)
```

```
## Generalized linear mixed model fit by maximum likelihood (Laplace
##   Approximation) [glmerMod]
##  Family: Negative Binomial(0.2311)  ( log )
## Formula: round(ShW) ~ Intervention.type * Intervention.time + sex + lake +
##     ClassNum + (1 | School/Intervention.time)
##    Data: data400
##
##      AIC      BIC   logLik deviance df.resid
##  16870.6  16933.7  -8424.3  16848.6     2259
##
## Scaled residuals:
##     Min      1Q  Median      3Q     Max
## -0.4805 -0.4738 -0.4133 -0.0228 19.1518
##
## Random effects:
##  Groups                   Name        Variance Std.Dev.
##  Intervention.time:School (Intercept) 0.7025   0.8381
##  School                   (Intercept) 0.3803   0.6167
## Number of obs: 2270, groups:  Intervention.time:School, 32; School, 16
##
## Fixed effects:
##                                              Estimate Std. Error z value
## (Intercept)                                   4.95850    0.67751   7.319
## Intervention.typeremoval                     -0.04202    0.53471  -0.079
## Intervention.timeafter                       -0.16702    0.44176  -0.378
## sexF                                         -0.57399    0.52871  -1.086
## sexM                                         -0.30387    0.53256  -0.571
## lakeriver                                    -1.54929    0.45317  -3.419
## ClassNum                                     -0.32095    0.05233  -6.133
## Intervention.typeremoval:Intervention.timeafter  0.57733    0.62243   0.928
##                                              Pr(>|z|)
## (Intercept)                                  2.50e-13 ***
## Intervention.typeremoval                     0.937369
## Intervention.timeafter                       0.705375
## sexF                                         0.277641
## sexM                                         0.568275
## lakeriver                                    0.000629 ***
## ClassNum                                     8.62e-10 ***
```

```
## Intervention.typeremoval:Intervention.timeafter 0.353651
## ---
## Signif. codes:  0 '***' 0.001 '**' 0.01 '*' 0.05 '.' 0.1 ' ' 1
##
## Correlation of Fixed Effects:
##             (Intr) Intrvntn.ty Intrvntn.tm sexF   sexM   lakrvr ClssNm
## Intrvntn.ty -0.392
## Intrvntn.tm -0.348  0.405
## sexF        -0.768 -0.006       0.029
## sexM        -0.761 -0.011       0.029       0.984
## lakeriver   -0.246  0.001       0.006      -0.018 -0.024
## ClassNum    -0.180  0.014       0.026       0.000 -0.014  0.055
## Intrvnt.:I.  0.245 -0.573      -0.709      -0.016 -0.016 -0.002 -0.030
```

```
Anova(veg.Sh.400)
```

```
## Analysis of Deviance Table (Type II Wald chisquare tests)
##
## Response: round(ShW)
##                                       Chisq Df Pr(>Chisq)
## Intervention.type                    0.3053  1   0.580560
## Intervention.time                    0.1576  1   0.691372
## sex                                  9.1526  2   0.010293 *
## lake                                11.6882  1   0.000629 ***
## ClassNum                            37.6143  1  8.621e-10 ***
## Intervention.type:Intervention.time  0.8603  1   0.353651
## ---
## Signif. codes:  0 '***' 0.001 '**' 0.01 '*' 0.05 '.' 0.1 ' ' 1
```

```
veg.Sh.800 = glmer.nb(round(ShW) ~ Intervention.type*Intervention.time + sex+lake+ClassNum+(1|School/In
summary(veg.Sh.800)
```

```
## Generalized linear mixed model fit by maximum likelihood (Laplace
##   Approximation) [glmerMod]
##  Family: Negative Binomial(0.2262)  ( log )
## Formula: round(ShW) ~ Intervention.type * Intervention.time + sex + lake +
##     ClassNum + (1 | School/Intervention.time)
##    Data: data800
##
##      AIC      BIC   logLik deviance df.resid
##  16957.9  17020.9  -8467.9  16935.9     2259
##
## Scaled residuals:
##     Min      1Q  Median      3Q     Max
## -0.4754 -0.4690 -0.4093 -0.0440 18.9843
##
## Random effects:
##  Groups                   Name        Variance Std.Dev.
##  Intervention.time:School (Intercept) 0.8559   0.9252
##  School                   (Intercept) 0.3091   0.5560
## Number of obs: 2270, groups:  Intervention.time:School, 32; School, 16
##
## Fixed effects:
```

```
##                                                 Estimate Std. Error z value
## (Intercept)                                      5.07692    0.70127   7.240
## Intervention.typeremoval                        -0.03880    0.55454  -0.070
## Intervention.timeafter                          -0.01664    0.48424  -0.034
## sexF                                            -0.64466    0.54581  -1.181
## sexM                                            -0.38307    0.55005  -0.696
## lakeriver                                       -1.66452    0.45524  -3.656
## ClassNum                                        -0.32211    0.05299  -6.079
## Intervention.typeremoval:Intervention.timeafter  0.56456    0.68224   0.828
##                                                 Pr(>|z|)
## (Intercept)                                     4.50e-13 ***
## Intervention.typeremoval                        0.944223
## Intervention.timeafter                          0.972580
## sexF                                            0.237558
## sexM                                            0.486160
## lakeriver                                       0.000256 ***
## ClassNum                                        1.21e-09 ***
## Intervention.typeremoval:Intervention.timeafter 0.407947
## ---
## Signif. codes:  0 '***' 0.001 '**' 0.01 '*' 0.05 '.' 0.1 ' ' 1
##
## Correlation of Fixed Effects:
##             (Intr) Intrvntn.ty Intrvntn.tm sexF   sexM   lakrvr ClssNm
## Intrvntn.ty -0.395
## Intrvntn.tm -0.370  0.430
## sexF        -0.770 -0.003       0.032
## sexM        -0.764 -0.008       0.031       0.985
## lakeriver   -0.239  0.002       0.006      -0.018 -0.024
## ClassNum    -0.187  0.013       0.026       0.013 -0.001  0.055
## Intrvnt.:I.  0.263 -0.608      -0.710      -0.020 -0.020 -0.002 -0.030
```

```
Anova(veg.Sh.800)
```

```
## Analysis of Deviance Table (Type II Wald chisquare tests)
##
## Response: round(ShW)
##                                       Chisq Df Pr(>Chisq)
## Intervention.type                    0.2970  1  0.5857837
## Intervention.time                    0.6154  1  0.4327594
## sex                                  8.6914  2  0.0129622 *
## lake                                13.3690  1  0.0002558 ***
## ClassNum                            36.9539  1   1.21e-09 ***
## Intervention.type:Intervention.time  0.6848  1  0.4079472
## ---
## Signif. codes:  0 '***' 0.001 '**' 0.01 '*' 0.05 '.' 0.1 ' ' 1
```

```
# Control
veg.Sh.300c = glmer.nb(round(ShW) ~ Intervention.type*Intervention.time + sex+lake+ClassNum+(1|School/I
summary(veg.Sh.300c)
```

```
## Generalized linear mixed model fit by maximum likelihood (Laplace
##   Approximation) [glmerMod]
##  Family: Negative Binomial(0.2335)  ( log )
```

```
## Formula: round(ShW) ~ Intervention.type * Intervention.time + sex + lake +
##     ClassNum + (1 | School/Intervention.time)
##    Data: data300c
##
##      AIC      BIC   logLik deviance df.resid
##  16821.2  16884.2  -8399.6  16799.2     2259
##
## Scaled residuals:
##     Min      1Q  Median      3Q     Max
## -0.4830 -0.4762 -0.4152 -0.0024 19.2242
##
## Random effects:
##  Groups                   Name        Variance Std.Dev.
##  Intervention.time:School (Intercept) 0.6304   0.7940
##  School                   (Intercept) 0.4273   0.6537
## Number of obs: 2270, groups:  Intervention.time:School, 32; School, 16
##
## Fixed effects:
##                                               Estimate Std. Error z value
## (Intercept)                                    4.92078    0.66623   7.386
## Intervention.typeremoval                      -0.04463    0.52836  -0.084
## Intervention.timeafter                        -0.21819    0.41991  -0.520
## sexF                                          -0.55613    0.51906  -1.071
## sexM                                          -0.28046    0.52262  -0.537
## lakeriver                                     -1.49112    0.45568  -3.272
## ClassNum                                      -0.32244    0.05201  -6.200
## Intervention.typeremoval:Intervention.timeafter  0.53213    0.59179   0.899
##                                               Pr(>|z|)
## (Intercept)                                   1.51e-13 ***
## Intervention.typeremoval                       0.93269
## Intervention.timeafter                         0.60332
## sexF                                           0.28399
## sexM                                           0.59152
## lakeriver                                      0.00107 **
## ClassNum                                      5.65e-10 ***
## Intervention.typeremoval:Intervention.timeafter  0.36855
## ---
## Signif. codes:  0 '***' 0.001 '**' 0.01 '*' 0.05 '.' 0.1 ' ' 1
##
## Correlation of Fixed Effects:
##             (Intr) Intrvntn.ty Intrvntn.tm sexF   sexM   lakrvr ClssNm
## Intrvntn.ty -0.393
## Intrvntn.tm -0.336  0.389
## sexF        -0.764 -0.006       0.027
## sexM        -0.757 -0.012       0.027       0.984
## lakeriver   -0.251  0.001       0.006      -0.019 -0.025
## ClassNum    -0.176  0.013       0.026      -0.008 -0.022  0.054
## Intrvnt.:I.  0.235 -0.550      -0.709      -0.014 -0.014 -0.002 -0.030
```

```
Anova(veg.Sh.300c)
```

```
## Analysis of Deviance Table (Type II Wald chisquare tests)
##
## Response: round(ShW)
```

```
##                                     Chisq Df Pr(>Chisq)
## Intervention.type                  0.2416  1   0.623078
## Intervention.time                  0.0279  1   0.867338
## sex                                9.5294  2   0.008525 **
## lake                              10.7079  1   0.001067 **
## ClassNum                          38.4372  1  5.655e-10 ***
## Intervention.type:Intervention.time  0.8085  1   0.368555
## ---
## Signif. codes:  0 '***' 0.001 '**' 0.01 '*' 0.05 '.' 0.1 ' ' 1
```

```
veg.Sh.400c = glmer.nb(round(ShW) ~ Intervention.type*Intervention.time + sex+lake+ClassNum+(1|School/In
summary(veg.Sh.400c)
```

```
## Generalized linear mixed model fit by maximum likelihood (Laplace
##   Approximation) [glmerMod]
##  Family: Negative Binomial(0.2326)  ( log )
## Formula: round(ShW) ~ Intervention.type * Intervention.time + sex + lake +
##     ClassNum + (1 | School/Intervention.time)
##    Data: data400c
##
##      AIC      BIC   logLik deviance df.resid
##  16839.0  16902.0  -8408.5  16817.0     2259
##
## Scaled residuals:
##     Min      1Q  Median      3Q     Max
## -0.4821 -0.4753 -0.4149 -0.0100 19.1978
##
## Random effects:
##  Groups                   Name        Variance Std.Dev.
##  Intervention.time:School (Intercept) 0.6461   0.8038
##  School                   (Intercept) 0.4267   0.6532
## Number of obs: 2270, groups:  Intervention.time:School, 32; School, 16
##
## Fixed effects:
##                                              Estimate Std. Error z value
## (Intercept)                                   4.98756    0.67618   7.376
## Intervention.typeremoval                     -0.04401    0.53328  -0.083
## Intervention.timeafter                       -0.16862    0.42539  -0.396
## sexF                                         -0.61207    0.52649  -1.163
## sexM                                         -0.33764    0.53020  -0.637
## lakeriver                                    -1.51093    0.45820  -3.298
## ClassNum                                     -0.32402    0.05212  -6.216
## Intervention.typeremoval:Intervention.timeafter  0.48239    0.59975   0.804
##                                              Pr(>|z|)
## (Intercept)                                  1.63e-13 ***
## Intervention.typeremoval                     0.934222
## Intervention.timeafter                       0.691827
## sexF                                         0.245014
## sexM                                         0.524249
## lakeriver                                    0.000975 ***
## ClassNum                                     5.09e-10 ***
## Intervention.typeremoval:Intervention.timeafter 0.421211
## ---
## Signif. codes:  0 '***' 0.001 '**' 0.01 '*' 0.05 '.' 0.1 ' ' 1
```

```
##
## Correlation of Fixed Effects:
##              (Intr) Intrvntn.ty Intrvntn.tm sexF   sexM   lakrvr ClssNm
## Intrvntn.ty -0.393
## Intrvntn.tm -0.339  0.392
## sexF        -0.767 -0.004       0.031
## sexM        -0.761 -0.009       0.031       0.984
## lakeriver   -0.250  0.002       0.006      -0.018 -0.024
## ClassNum    -0.179  0.014       0.027      -0.001 -0.016  0.054
## Intrvnt.:I.  0.238 -0.554      -0.710      -0.016 -0.017 -0.003 -0.031
```

```
Anova(veg.Sh.400c)
```

```
## Analysis of Deviance Table (Type II Wald chisquare tests)
##
## Response: round(ShW)
##                                      Chisq Df Pr(>Chisq)
## Intervention.type                   0.1906  1  0.6624340
## Intervention.time                   0.0613  1  0.8044795
## sex                                 9.6115  2  0.0081827 **
## lake                               10.8736  1  0.0009755 ***
## ClassNum                           38.6443  1  5.085e-10 ***
## Intervention.type:Intervention.time  0.6469  1  0.4212107
## ---
## Signif. codes:  0 '***' 0.001 '**' 0.01 '*' 0.05 '.' 0.1 ' ' 1
```

```
veg.Sh.800c = glmer.nb(round(ShW) ~ Intervention.type*Intervention.time + sex+lake+ClassNum+(1|School/In
summary(veg.Sh.800c)
```

```
## Generalized linear mixed model fit by maximum likelihood (Laplace
##   Approximation) [glmerMod]
##  Family: Negative Binomial(0.23)  ( log )
## Formula: round(ShW) ~ Intervention.type * Intervention.time + sex + lake +
##     ClassNum + (1 | School/Intervention.time)
##    Data: data800c
##
##      AIC      BIC   logLik deviance df.resid
##  16887.8  16950.8  -8432.9  16865.8     2259
##
## Scaled residuals:
##     Min      1Q  Median      3Q     Max
## -0.4793 -0.4727 -0.4130 -0.0234 19.1024
##
## Random effects:
##  Groups                   Name        Variance Std.Dev.
##  Intervention.time:School (Intercept) 0.7087   0.8418
##  School                   (Intercept) 0.4217   0.6494
## Number of obs: 2270, groups:  Intervention.time:School, 32; School, 16
##
## Fixed effects:
##                                                   Estimate Std. Error z value
## (Intercept)                                        5.12749    0.69111   7.419
## Intervention.typeremoval                          -0.04271    0.54676  -0.078
```

```
## Intervention.timeafter                          -0.01969    0.44433  -0.044
## sexF                                            -0.72002    0.53601  -1.343
## sexM                                            -0.44871    0.53999  -0.831
## lakeriver                                       -1.57133    0.46654  -3.368
## ClassNum                                        -0.32825    0.05247  -6.256
## Intervention.typeremoval:Intervention.timeafter  0.33338    0.62636   0.532
##                                                 Pr(>|z|)
## (Intercept)                                     1.18e-13 ***
## Intervention.typeremoval                        0.937730
## Intervention.timeafter                          0.964650
## sexF                                            0.179173
## sexM                                            0.405996
## lakeriver                                       0.000757 ***
## ClassNum                                        3.96e-10 ***
## Intervention.typeremoval:Intervention.timeafter 0.594554
## ---
## Signif. codes:  0 '***' 0.001 '**' 0.01 '*' 0.05 '.' 0.1 ' ' 1
##
## Correlation of Fixed Effects:
##              (Intr) Intrvntn.ty Intrvntn.tm sexF   sexM   lakrvr ClssNm
## Intrvntn.ty -0.395
## Intrvntn.tm -0.347  0.400
## sexF        -0.766 -0.003       0.032
## sexM        -0.760 -0.009       0.031       0.985
## lakeriver   -0.249  0.002       0.006      -0.018 -0.024
## ClassNum    -0.186  0.014       0.028       0.010 -0.004  0.053
## Intrvnt.:I.  0.244 -0.565      -0.710      -0.018 -0.018 -0.003 -0.031
```

```
Anova(veg.Sh.800c)
```

```
## Analysis of Deviance Table (Type II Wald chisquare tests)
##
## Response: round(ShW)
##                                       Chisq Df Pr(>Chisq)
## Intervention.type                    0.0727  1  0.7875031
## Intervention.time                    0.2247  1  0.6354635
## sex                                  9.7486  2  0.0076404 **
## lake                                11.3436  1  0.0007571 ***
## ClassNum                            39.1315  1  3.962e-10 ***
## Intervention.type:Intervention.time  0.2833  1  0.5945535
## ---
## Signif. codes:  0 '***' 0.001 '**' 0.01 '*' 0.05 '.' 0.1 ' ' 1
```

```
#Intervention

veg.Sh.300t = glmer.nb(round(ShW) ~ Intervention.type*Intervention.time + sex+lake+ClassNum+(1|School/I
summary(veg.Sh.300t)
```

```
## Generalized linear mixed model fit by maximum likelihood (Laplace
##   Approximation) [glmerMod]
##  Family: Negative Binomial(0.2336)  ( log )
## Formula: round(ShW) ~ Intervention.type * Intervention.time + sex + lake +
##     ClassNum + (1 | School/Intervention.time)
```

```
##    Data: data300t
##
##      AIC      BIC   logLik deviance df.resid
##  16816.1  16879.1  -8397.0  16794.1     2259
##
## Scaled residuals:
##     Min      1Q  Median      3Q     Max
## -0.4831 -0.4763 -0.4147 -0.0072 19.2258
##
## Random effects:
##  Groups                    Name        Variance Std.Dev.
##  Intervention.time:School (Intercept) 0.6451   0.8032
##  School                    (Intercept) 0.4025   0.6344
## Number of obs: 2270, groups:  Intervention.time:School, 32; School, 16
##
## Fixed effects:
##                                                Estimate Std. Error z value
## (Intercept)                                     4.80180    0.66491   7.222
## Intervention.typeremoval                       -0.04437    0.52623  -0.084
## Intervention.timeafter                         -0.27532    0.42441  -0.649
## sexF                                           -0.44521    0.51878  -0.858
## sexM                                           -0.17039    0.52225  -0.326
## lakeriver                                      -1.48873    0.45122  -3.299
## ClassNum                                       -0.31875    0.05201  -6.129
## Intervention.typeremoval:Intervention.timeafter  0.64118    0.59777   1.073
##                                                Pr(>|z|)
## (Intercept)                                    5.13e-13 ***
## Intervention.typeremoval                       0.932808
## Intervention.timeafter                         0.516520
## sexF                                           0.390786
## sexM                                           0.744218
## lakeriver                                      0.000969 ***
## ClassNum                                       8.87e-10 ***
## Intervention.typeremoval:Intervention.timeafter 0.283435
## ---
## Signif. codes:  0 '***' 0.001 '**' 0.01 '*' 0.05 '.' 0.1 ' ' 1
##
## Correlation of Fixed Effects:
##             (Intr) Intrvntn.ty Intrvntn.tm sexF   sexM   lakrvr ClssNm
## Intrvntn.ty -0.394
## Intrvntn.tm -0.340  0.395
## sexF        -0.765 -0.005       0.027
## sexM        -0.758 -0.010       0.026       0.984
## lakeriver   -0.250  0.001       0.006      -0.019 -0.025
## ClassNum    -0.172  0.014       0.025      -0.014 -0.028  0.055
## Intrvnt.:I.  0.239 -0.559      -0.709      -0.014 -0.014 -0.002 -0.030
```

```
Anova(veg.Sh.300t)
```

```
## Analysis of Deviance Table (Type II Wald chisquare tests)
##
## Response: round(ShW)
##                                    Chisq Df Pr(>Chisq)
## Intervention.type                 0.3860  1  0.5344222
```

```
## Intervention.time                        0.0251  1  0.8740216
## sex                                      9.1340  2  0.0103891 *
## lake                                    10.8854  1  0.0009693 ***
## ClassNum                                37.5598  1  8.865e-10 ***
## Intervention.type:Intervention.time      1.1505  1  0.2834351
## ---
## Signif. codes:  0 '***' 0.001 '**' 0.01 '*' 0.05 '.' 0.1 ' ' 1
```

```
veg.Sh.400t = glmer.nb(round(ShW) ~ Intervention.type*Intervention.time + sex+lake+ClassNum+(1|School/I
summary(veg.Sh.400t)
```

```
## Generalized linear mixed model fit by maximum likelihood (Laplace
##   Approximation) [glmerMod]
##  Family: Negative Binomial(0.2328)  ( log )
## Formula: round(ShW) ~ Intervention.type * Intervention.time + sex + lake +
##     ClassNum + (1 | School/Intervention.time)
##    Data: data400t
##
##      AIC      BIC   logLik deviance df.resid
##  16830.1  16893.1  -8404.1  16808.1     2259
##
## Scaled residuals:
##     Min      1Q  Median      3Q     Max
## -0.4823 -0.4756 -0.4142 -0.0122 19.2014
##
## Random effects:
##  Groups                   Name        Variance Std.Dev.
##  Intervention.time:School (Intercept) 0.6719   0.8197
##  School                   (Intercept) 0.3815   0.6177
## Number of obs: 2270, groups:  Intervention.time:School, 32; School, 16
##
## Fixed effects:
##                                                Estimate Std. Error z value
## (Intercept)                                     4.78779    0.66801   7.167
## Intervention.typeremoval                       -0.04347    0.52924  -0.082
## Intervention.timeafter                         -0.27460    0.43357  -0.633
## sexF                                           -0.42679    0.52122  -0.819
## sexM                                           -0.15392    0.52474  -0.293
## lakeriver                                      -1.50654    0.44908  -3.355
## ClassNum                                       -0.31744    0.05211  -6.091
## Intervention.typeremoval:Intervention.timeafter  0.68517    0.61119   1.121
##                                                Pr(>|z|)
## (Intercept)                                    7.65e-13 ***
## Intervention.typeremoval                       0.934542
## Intervention.timeafter                         0.526501
## sexF                                           0.412883
## sexM                                           0.769272
## lakeriver                                      0.000794 ***
## ClassNum                                       1.12e-09 ***
## Intervention.typeremoval:Intervention.timeafter 0.262269
## ---
## Signif. codes:  0 '***' 0.001 '**' 0.01 '*' 0.05 '.' 0.1 ' ' 1
##
## Correlation of Fixed Effects:
```

```
##              (Intr) Intrvntn.ty Intrvntn.tm sexF   sexM   lakrvr ClssNm
## Intrvntn.ty -0.395
## Intrvntn.tm -0.347  0.404
## sexF        -0.765 -0.005       0.028
## sexM        -0.759 -0.010       0.028       0.984
## lakeriver   -0.248  0.002       0.006      -0.018 -0.025
## ClassNum    -0.172  0.014       0.024      -0.013 -0.027  0.056
## Intrvnt.:I.  0.245 -0.570      -0.710      -0.016 -0.016 -0.002 -0.030
```

```
Anova(veg.Sh.400t)
```

```
## Analysis of Deviance Table (Type II Wald chisquare tests)
##
## Response: round(ShW)
##                                      Chisq Df Pr(>Chisq)
## Intervention.type                   0.4597  1  0.4977654
## Intervention.time                   0.0536  1  0.8168792
## sex                                 8.9348  2  0.0114772 *
## lake                               11.2543  1  0.0007944 ***
## ClassNum                           37.1023  1  1.121e-09 ***
## Intervention.type:Intervention.time  1.2567  1  0.2622686
## ---
## Signif. codes:  0 '***' 0.001 '**' 0.01 '*' 0.05 '.' 0.1 ' ' 1
```

```
veg.Sh.800t = glmer.nb(round(ShW) ~ Intervention.type*Intervention.time + sex+lake+ClassNum+(1|School/In
summary(veg.Sh.800t)
```

```
## Generalized linear mixed model fit by maximum likelihood (Laplace
##   Approximation) [glmerMod]
##  Family: Negative Binomial(0.2305)  ( log )
## Formula: round(ShW) ~ Intervention.type * Intervention.time + sex + lake +
##     ClassNum + (1 | School/Intervention.time)
##    Data: data800t
##
##      AIC      BIC   logLik deviance df.resid
##  16869.3  16932.3  -8423.7  16847.3     2259
##
## Scaled residuals:
##     Min      1Q  Median      3Q     Max
## -0.4799 -0.4732 -0.4117 -0.0250 19.1137
##
## Random effects:
##  Groups                   Name        Variance Std.Dev.
##  Intervention.time:School (Intercept) 0.7620   0.8729
##  School                   (Intercept) 0.3189   0.5647
## Number of obs: 2270, groups:  Intervention.time:School, 32; School, 16
##
## Fixed effects:
##                                              Estimate Std. Error z value
## (Intercept)                                   4.75651    0.67413   7.056
## Intervention.typeremoval                     -0.04124    0.53503  -0.077
## Intervention.timeafter                       -0.27283    0.45822  -0.595
## sexF                                         -0.38019    0.52660  -0.722
```

```
## sexM                                            -0.11260    0.53029  -0.212
## lakeriver                                       -1.56137    0.44409  -3.516
## ClassNum                                        -0.31430    0.05244  -5.994
## Intervention.typeremoval:Intervention.timeafter  0.82038    0.64594   1.270
##                                                 Pr(>|z|)
## (Intercept)                                     1.72e-12 ***
## Intervention.typeremoval                        0.938557
## Intervention.timeafter                          0.551569
## sexF                                            0.470316
## sexM                                            0.831839
## lakeriver                                       0.000438 ***
## ClassNum                                        2.05e-09 ***
## Intervention.typeremoval:Intervention.timeafter 0.204060
## ---
## Signif. codes:  0 '***' 0.001 '**' 0.01 '*' 0.05 '.' 0.1 ' ' 1
##
## Correlation of Fixed Effects:
##             (Intr) Intrvntn.ty Intrvntn.tm sexF   sexM   lakrvr ClssNm
## Intrvntn.ty -0.394
## Intrvntn.tm -0.361  0.421
## sexF        -0.766 -0.006       0.027
## sexM        -0.760 -0.011       0.027       0.984
## lakeriver   -0.243  0.002       0.006      -0.018 -0.025
## ClassNum    -0.173  0.014       0.023      -0.012 -0.026  0.057
## Intrvnt.:I.  0.255 -0.596      -0.709      -0.015 -0.015 -0.002 -0.029
```

```
Anova(veg.Sh.800t)
```

```
## Analysis of Deviance Table (Type II Wald chisquare tests)
##
## Response: round(ShW)
##                                     Chisq Df Pr(>Chisq)
## Intervention.type                  0.7173  1  0.3970195
## Intervention.time                  0.1871  1  0.6653182
## sex                                8.4017  2  0.0149831 *
## lake                              12.3613  1  0.0004383 ***
## ClassNum                          35.9287  1  2.047e-09 ***
## Intervention.type:Intervention.time  1.6131  1  0.2040605
## ---
## Signif. codes:  0 '***' 0.001 '**' 0.01 '*' 0.05 '.' 0.1 ' ' 1
```

```
####################################
################### HUMAN EGG BURDEN
####################################

#For BACI plots for eggs see the Means_difference_scores.xls (difference scores)

#############   BACI comparisons for eggs and intensity###################

#SH Intensity... Use subset to select values that were > 0- not included in paper

dataf1=subset(datanew, ShW!=0)
dataf2=subset(dataf1, Intervention.time=="before" | Intervention.time=="after")
```

```
dataf2$Intervention.time<- factor(dataf2$Intervention.time,levels = c("before", "after"))
veg.Sh.intensity = glmer.nb(round(ShW) ~ Intervention.type*Intervention.time +sex+ lake+(1|School/Inter
Anova(veg.Sh.intensity)
```

```
## Analysis of Deviance Table (Type II Wald chisquare tests)
##
## Response: round(ShW)
##                                     Chisq Df Pr(>Chisq)
## Intervention.type                  0.0946  1     0.7585
## Intervention.time                  1.3194  1     0.2507
## sex                                0.7381  2     0.6914
## lake                               1.3957  1     0.2374
## Intervention.type:Intervention.time 0.3737  1     0.5410
```

```
#SM Intensity for those Infected (egg count>0) ##########################
dataf1=subset(datanew, SmW!=0)
dataf2=subset(dataf1, Intervention.time=="before" | Intervention.time=="after")
dataf2$Intervention.time<- factor(dataf2$Intervention.time,levels = c("before", "after"))
veg.Sm.intensity = lmer(round(LogSmW) ~ Intervention.type*Intervention.time +sex+ lake+(1|School/Interv
Anova(veg.Sm.intensity)
```

```
## Analysis of Deviance Table (Type II Wald chisquare tests)
##
## Response: round(LogSmW)
##                                     Chisq Df Pr(>Chisq)
## Intervention.type                  0.1963  1     0.6577
## Intervention.time                  0.6969  1     0.4038
## sex                                2.5979  2     0.2728
## lake                               1.7257  1     0.1890
## Intervention.type:Intervention.time 0.1382  1     0.7101
```

```
#######################
#Human water contact:
#######################

#Age range of the person (<15 years = 1, 15-35 = 2, > 35 = 3
waterdat<-read.csv("water_contact.csv")
names(waterdat)
```

```
##  [1] "Observation.ID"   "village"          "Sampling.bout"    "time"
##  [5] "time2"            "intervention"     "sex"              "age.range"
##  [9] "Activity"         "level.of.contact" "time.of.day"      "starting.time"
## [13] "ending.time"      "Duration.minutes"
```

```
waterdat$Sampling.bout=factor(waterdat$Sampling.bout)
#Can model ordinal data with mixed models (1,2,3...) in the ordinal package:

waterdat$level=as.factor(waterdat$level.of.contact)
waterdat$age=as.factor(waterdat$age.range)
waterdat$vil_time<-paste(waterdat$village,waterdat$time,sep="_")
fmm1 <- clmm(level ~ intervention*time+time.of.day+sex+age+(1|village)+(1|vil_time),data=waterdat, na.a
```

```
## Warning: Using formula(x) is deprecated when x is a character vector of length > 1.
##   Consider formula(paste(x, collapse = " ")) instead.
```

```r
summary(fmm1)
```

```
## Cumulative Link Mixed Model fitted with the Laplace approximation
##
## formula: level ~ intervention * time + time.of.day + sex + age + (1 |
##     village) + (1 | vil_time)
## data:    waterdat
##
##  link  threshold nobs logLik  AIC     niter      max.grad cond.H
##  logit flexible  524  -628.13 1282.26 1118(2449) 1.48e-03 3.0e+04
##
## Random effects:
##  Groups   Name        Variance  Std.Dev.
##  vil_time (Intercept) 1.350e-02 1.162e-01
##  village  (Intercept) 1.651e-13 4.063e-07
## Number of groups:  vil_time 8,  village 4
##
## Coefficients:
##                                Estimate Std. Error z value Pr(>|z|)
## interventionremoval             1.20379    0.26215   4.592 4.39e-06 ***
## timebefore                     -0.10644    0.28127  -0.378    0.705
## time.of.day                     0.03595    0.03573   1.006    0.314
## sexM                            0.73932    0.18946   3.902 9.53e-05 ***
## age2                           -1.12362    0.21239  -5.290 1.22e-07 ***
## age3                           -1.11701    0.23019  -4.853 1.22e-06 ***
## interventionremoval:timebefore  0.01859    0.38307   0.049    0.961
## ---
## Signif. codes:  0 '***' 0.001 '**' 0.01 '*' 0.05 '.' 0.1 ' ' 1
##
## Threshold coefficients:
##     Estimate Std. Error z value
## 1|2  -2.0721     0.5835  -3.551
## 2|3  -0.3173     0.5703  -0.556
## 3|4   0.9477     0.5704   1.662
## 4|5   1.1373     0.5705   1.994
## (2 observations deleted due to missingness)
```

```r
#duration of water contact:

hist(log(waterdat$Duration.minutes))
```

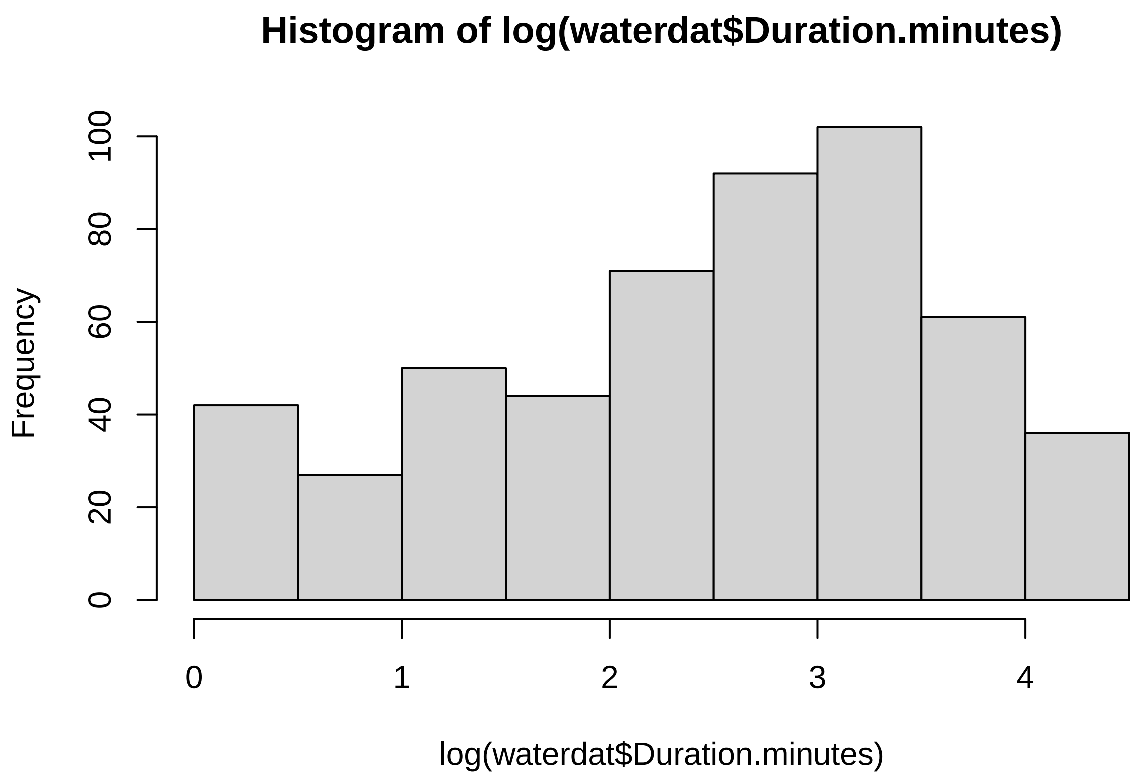


```
waterdat$logmin=log(waterdat$Duration.minutes+1)
water2= lmer(logmin ~ intervention*time +time.of.day+sex+age+(1|village/time),data=waterdat, na.action=
```

```
## boundary (singular) fit: see ?isSingular
```

```
Anova(water2)
```

```
## Analysis of Deviance Table (Type II Wald chisquare tests)
##
## Response: logmin
##                      Chisq Df Pr(>Chisq)
## intervention        0.0030  1     0.9561
## time                0.0564  1     0.8123
## time.of.day        16.2043  1  5.686e-05 ***
## sex                22.5400  1  2.058e-06 ***
## age                27.1761  2  1.255e-06 ***
## intervention:time   0.1358  1     0.7125
## ---
## Signif. codes:  0 '***' 0.001 '**' 0.01 '*' 0.05 '.' 0.1 ' ' 1
```

```
summary(water2)
```

```
## Linear mixed model fit by REML ['lmerMod']
## Formula: logmin ~ intervention * time + time.of.day + sex + age + (1 |
```

```
##     village/time)
##    Data: waterdat
##
## REML criterion at convergence: 1430.6
##
## Scaled residuals:
##     Min      1Q  Median      3Q     Max
## -2.9195 -0.6701  0.1927  0.7212  2.0535
##
## Random effects:
##  Groups       Name        Variance Std.Dev.
##  time:village (Intercept) 0.0000   0.0000
##  village      (Intercept) 4.8728   2.2075
##  Residual                 0.8415   0.9173
## Number of obs: 526, groups:  time:village, 8; village, 4
##
## Fixed effects:
##                                Estimate Std. Error t value
## (Intercept)                    10.21418    2.40316   4.250
## interventionremoval             0.14912    2.21034   0.067
## timebefore                      0.01867    0.13045   0.143
## time.of.day                    -0.53129    0.13198  -4.025
## sexM                           -0.41398    0.08720  -4.748
## age2                           -0.46536    0.10290  -4.523
## age3                           -0.43924    0.11032  -3.982
## interventionremoval:timebefore -0.06128    0.16627  -0.369
##
## Correlation of Fixed Effects:
##             (Intr) intrvn timbfr tm.f.d sexM   age2   age3
## intrvntnrmv -0.463
## timebefore   0.043  0.026
## time.of.day -0.759  0.004 -0.088
## sexM        -0.018  0.003  0.028 -0.007
## age2        -0.024  0.002 -0.036  0.007  0.278
## age3         0.000  0.001  0.004 -0.025  0.271  0.342
## intrvntnrm: -0.049 -0.034 -0.786  0.089 -0.018  0.032 -0.003
## optimizer (nloptwrap) convergence code: 0 (OK)
## boundary (singular) fit: see ?isSingular
```

```
#no trends for intervention. I think time of day is consuming the effect because Raph did observations
#in the afternoon at the removal villages and morning at the control
```

```
########################################################
#Crops trials using compost made from removed vegetation
########################################################



#In each of 6 rows (A1,B1,C1  or A2,B2,C2) the whole-plot is UreaorNot and the sub-plot is treatment TC
#thus, the block in the experimental is row because it's repeated for both sub and whole plot factors



#### PEPPER ############
#crops at all villages in one, with blocking by village (because for pepper there was a field at two vi
crop=read.csv("crops_all.csv")
```

```r
crop$row=as.factor(crop$row)
crop$harvestID=as.factor(crop$harvestID)
crop$Urea.Treatment=as.factor(crop$UorN)
crop$row=as.factor(crop$row)
crop$harvestID=as.factor(crop$harvestID)
crop$treatment<-as.factor(crop$treatment)
crop$treatment<-relevel(crop$treatment, ref="NC")
#this matches the example here:https://people.bath.ac.uk/jjf23/mixchange/split.html
#I think we should select pepper and run with effect of village but combined:
selectedp=c("pepper")
crop2=crop[crop$crop %in% selectedp,]

# total number of peppers
data.pep1 <- glmer.nb(fruits ~  treatment*Urea.Treatment+village +(1 | harvestID), data = crop2, na.acti
Anova(data.pep1)
```

```
## Analysis of Deviance Table (Type II Wald chisquare tests)
##
## Response: fruits
##                             Chisq Df Pr(>Chisq)
## treatment                101.8036  2    < 2e-16 ***
## Urea.Treatment             0.5513  1    0.45779
## village                  151.7430  1    < 2e-16 ***
## treatment:Urea.Treatment   8.4798  2    0.01441 *
## ---
## Signif. codes:  0 '***' 0.001 '**' 0.01 '*' 0.05 '.' 0.1 ' ' 1
```

```r
summary(data.pep1)
```

```
## Generalized linear mixed model fit by maximum likelihood (Laplace
##   Approximation) [glmerMod]
##  Family: Negative Binomial(1.244)  ( log )
## Formula: fruits ~ treatment * Urea.Treatment + village + (1 | harvestID)
##    Data: crop2
##
##      AIC      BIC   logLik deviance df.resid
##   2849.6   2884.6  -1415.8   2831.6      351
##
## Scaled residuals:
##     Min      1Q  Median      3Q     Max
## -1.0692 -0.7057 -0.3463  0.3428  6.7590
##
## Random effects:
##  Groups    Name        Variance Std.Dev.
##  harvestID (Intercept) 0.3371   0.5806
## Number of obs: 360, groups:  harvestID, 11
##
## Fixed effects:
##                              Estimate Std. Error z value Pr(>|z|)
## (Intercept)                    1.5509     0.2321   6.682 2.36e-11 ***
## treatmentC                     0.9081     0.1780   5.103 3.34e-07 ***
## treatmentTC                    0.9281     0.1800   5.156 2.52e-07 ***
```

```
## Urea.TreatmentU               -0.3187     0.1745  -1.826  0.06779 .
## villagembane                   1.4612     0.1186  12.318  < 2e-16 ***
## treatmentC:Urea.TreatmentU     0.4671     0.2444   1.911  0.05601 .
## treatmentTC:Urea.TreatmentU    0.7010     0.2446   2.866  0.00416 **
## ---
## Signif. codes:  0 '***' 0.001 '**' 0.01 '*' 0.05 '.' 0.1 ' ' 1
##
## Correlation of Fixed Effects:
##             (Intr) trtmnC trtmTC Ur.TrU vllgmb tC:U.T
## treatmentC  -0.462
## treatmentTC -0.469  0.540
## Ure.TrtmntU -0.373  0.483  0.479
## villagemban -0.373  0.238  0.261  0.008
## trtmnC:U.TU  0.245 -0.666 -0.326 -0.713  0.042
## trtmTC:U.TU  0.244 -0.331 -0.661 -0.713  0.055  0.512
```

```
data.pep1p <- glmer(fruits ~  treatment*Urea.Treatment+village+(1 | harvestID), data = crop2, na.action=
AIC(data.pep1,data.pep1p) # NB lower AIC
```

```
##            df      AIC
## data.pep1   9 2849.626
## data.pep1p  8 5913.026
```

```
# total weight grown
crop2$lnweight=log(crop2$weight+1)
data.pep2 <- glmmTMB(lnweight ~  treatment*Urea.Treatment+village +(1 |harvestID),data = crop2, na.acti
Anova(data.pep2)
```

```
## Analysis of Deviance Table (Type II Wald chisquare tests)
##
## Response: lnweight
##                             Chisq Df Pr(>Chisq)
## treatment                141.7235  2     <2e-16 ***
## Urea.Treatment             1.5184  1     0.2179
## village                  155.2452  1     <2e-16 ***
## treatment:Urea.Treatment   2.2010  2     0.3327
## ---
## Signif. codes:  0 '***' 0.001 '**' 0.01 '*' 0.05 '.' 0.1 ' ' 1
```

```
summary(data.pep2)
```

```
##  Family: gaussian  ( identity )
## Formula:
## lnweight ~ treatment * Urea.Treatment + village + (1 | harvestID)
## Data: crop2
##
##      AIC      BIC   logLik deviance df.resid
##   1416.6   1451.6   -699.3   1398.6      351
##
## Random effects:
##
## Conditional model:
```

```
##  Groups    Name        Variance Std.Dev.
##  harvestID (Intercept) 0.5464   0.7392
##  Residual              2.6794   1.6369
## Number of obs: 360, groups:  harvestID, 11
##
## Dispersion estimate for gaussian family (sigma^2): 2.68
##
## Conditional model:
##                              Estimate Std. Error z value Pr(>|z|)
## (Intercept)                  3.006398   0.316894   9.487  < 2e-16 ***
## treatmentC                   1.955667   0.298855   6.544 6.00e-11 ***
## treatmentTC                  2.048962   0.298855   6.856 7.08e-12 ***
## Urea.TreatmentU              0.005779   0.298855   0.019    0.985
## villagembane                 2.244821   0.180166  12.460  < 2e-16 ***
## treatmentC:Urea.TreatmentU   0.052952   0.422645   0.125    0.900
## treatmentTC:Urea.TreatmentU  0.567554   0.422645   1.343    0.179
## ---
## Signif. codes:  0 '***' 0.001 '**' 0.01 '*' 0.05 '.' 0.1 ' ' 1
```

```
##########Onion: (only at village Lampsar)################
#For onion, there can be no village effect or harvestid
selectedo=c("onion")
onion=crop[crop$crop %in% selectedo,]
onion$row=as.factor(onion$row)

OnionNB<- glm.nb(fruits ~  treatment*Urea.Treatment,data = onion)
OnionP <- glm(fruits ~  treatment*Urea.Treatment,data = onion, family = "poisson")
AIC(OnionNB, OnionP)#better fit
```

```
##         df      AIC
## OnionNB  7 192.0753
## OnionP   6 220.6876
```

```
summary(OnionNB)
```

```
##
## Call:
## glm.nb(formula = fruits ~ treatment * Urea.Treatment, data = onion,
##     init.theta = 72.95324983, link = log)
##
## Deviance Residuals:
##     Min       1Q   Median       3Q      Max
## -1.6322  -0.9128  -0.1045   0.6544   2.2750
##
## Coefficients:
##                              Estimate Std. Error z value Pr(>|z|)
## (Intercept)                   5.24702    0.07952  65.983  < 2e-16 ***
## treatmentC                    0.93575    0.10761   8.695  < 2e-16 ***
## treatmentTC                   0.96291    0.10753   8.955  < 2e-16 ***
## Urea.TreatmentU              -0.82418    0.12204  -6.754 1.44e-11 ***
## treatmentC:Urea.TreatmentU   -0.01786    0.16222  -0.110   0.9123
## treatmentTC:Urea.TreatmentU   0.27024    0.16083   1.680   0.0929 .
## ---
```

```
## Signif. codes:  0 '***' 0.001 '**' 0.01 '*' 0.05 '.' 0.1 ' ' 1
##
## (Dispersion parameter for Negative Binomial(72.9532) family taken to be 1)
##
##     Null deviance: 348.647  on 17  degrees of freedom
## Residual deviance:  18.809  on 12  degrees of freedom
## AIC: 192.08
##
## Number of Fisher Scoring iterations: 1
##
##
##               Theta:  73.0
##           Std. Err.:  33.5
##
##  2 x log-likelihood:  -178.075
```

```
Anova(OnionNB)
```

```
## Analysis of Deviance Table (Type II tests)
##
## Response: fruits
##                          LR Chisq Df Pr(>Chisq)
## treatment                 198.242  2     <2e-16 ***
## Urea.Treatment            130.593  1     <2e-16 ***
## treatment:Urea.Treatment    4.517  2     0.1045
## ---
## Signif. codes:  0 '***' 0.001 '**' 0.01 '*' 0.05 '.' 0.1 ' ' 1
```

```
onion$lnweight=log(onion$weight+1)
OnionW <- lm(lnweight ~ treatment * Urea.Treatment,data = onion, na.action=na.omit)
Anova(OnionW)
```

```
## Anova Table (Type II tests)
##
## Response: lnweight
##                           Sum Sq Df F value   Pr(>F)
## treatment                12.3611  2 60.3466 5.47e-07 ***
## Urea.Treatment            0.0401  1  0.3916   0.5432
## treatment:Urea.Treatment  0.0481  2  0.2349   0.7942
## Residuals                 1.2290 12
## ---
## Signif. codes:  0 '***' 0.001 '**' 0.01 '*' 0.05 '.' 0.1 ' ' 1
```

```
summary(OnionW)
```

```
##
## Call:
## lm(formula = lnweight ~ treatment * Urea.Treatment, data = onion,
##     na.action = na.omit)
##
## Residuals:
##     Min      1Q  Median      3Q     Max
```

```
## -0.3853 -0.2036 -0.0199  0.1482  0.5135
##
## Coefficients:
##                              Estimate Std. Error t value Pr(>|t|)
## (Intercept)                    1.2048     0.1848   6.520 2.85e-05 ***
## treatmentC                     1.7401     0.2613   6.659 2.33e-05 ***
## treatmentTC                    1.9469     0.2613   7.451 7.73e-06 ***
## Urea.TreatmentU                0.2277     0.2613   0.871    0.401
## treatmentC:Urea.TreatmentU    -0.2520     0.3695  -0.682    0.508
## treatmentTC:Urea.TreatmentU   -0.1478     0.3695  -0.400    0.696
## ---
## Signif. codes:  0 '***' 0.001 '**' 0.01 '*' 0.05 '.' 0.1 ' ' 1
##
## Residual standard error: 0.32 on 12 degrees of freedom
## Multiple R-squared:  0.9101, Adjusted R-squared:  0.8727
## F-statistic: 24.31 on 5 and 12 DF,  p-value: 6.842e-06
```

```
# Onion rot
onion$sick<-onion$kgsick.onion.

OnionRot <- lm(sick~ UorN*treatment, data=onion, na.action=na.omit)
summary(OnionRot)
```

```
##
## Call:
## lm(formula = sick ~ UorN * treatment, data = onion, na.action = na.omit)
##
## Residuals:
##    Min     1Q Median     3Q    Max
##   -3.5    0.0    0.0    0.0    2.5
##
## Coefficients:
##                      Estimate Std. Error t value Pr(>|t|)
## (Intercept)         9.315e-16  7.728e-01   0.000 1.000000
## UorNU              -1.675e-15  1.093e+00   0.000 1.000000
## treatmentC         -5.128e-16  1.093e+00   0.000 1.000000
## treatmentTC        -1.026e-15  1.093e+00   0.000 1.000000
## UorNU:treatmentC    8.500e+00  1.546e+00   5.499 0.000136 ***
## UorNU:treatmentTC   7.000e+00  1.546e+00   4.529 0.000691 ***
## ---
## Signif. codes:  0 '***' 0.001 '**' 0.01 '*' 0.05 '.' 0.1 ' ' 1
##
## Residual standard error: 1.339 on 12 degrees of freedom
## Multiple R-squared:  0.9189, Adjusted R-squared:  0.8851
## F-statistic:  27.2 on 5 and 12 DF,  p-value: 3.743e-06
```

```
Anova(OnionRot)
```

```
## Anova Table (Type II tests)
##
## Response: sick
##                 Sum Sq Df F value    Pr(>F)
## UorN            120.12  1  67.046 2.961e-06 ***
```

```
## treatment       61.75  2  17.233 0.0002967 ***
## UorN:treatment  61.75  2  17.233 0.0002967 ***
## Residuals       21.50 12
## ---
## Signif. codes:  0 '***' 0.001 '**' 0.01 '*' 0.05 '.' 0.1 ' ' 1
```

```
 #Probability of change in human infection (upload Fig file - formatted for figure)
Fig<-read.csv("Fig.csv")
```

```
## Warning in read.table(file = file, header = header, sep = sep, quote = quote, :
## incomplete final line found by readTableHeader on 'Fig.csv'
```

```
profileplotv4 <- ggplot(Fig, aes(x=Type, y=Point))+
  ggtitle("")+
  labs( x = "", y = expression(paste("Change in odds of ", italic("S. mansoni"), " infection")))+
  geom_point(position=position_dodge(w=0.1), size=2)+ geom_line(position=position_dodge(w=1), size=1)+
  geom_errorbar(aes(ymin=Point-SE, ymax=Point+SE), width=0.4, position=position_dodge(w=0.1))+
  theme_bw() + theme(axis.line = element_line(colour = "black"),
        panel.grid.major = element_blank(),
        panel.grid.minor = element_blank(),
        panel.border = element_blank(),
        panel.background = element_blank()) +theme(axis.text=element_text(size=10,colour = "black"),
                                                   axis.title=element_text(size=10, colour = "black"),)
profileplotv4
```

```
## geom_path: Each group consists of only one observation. Do you need to adjust
## the group aesthetic?
## geom_path: Each group consists of only one observation. Do you need to adjust
## the group aesthetic?
```

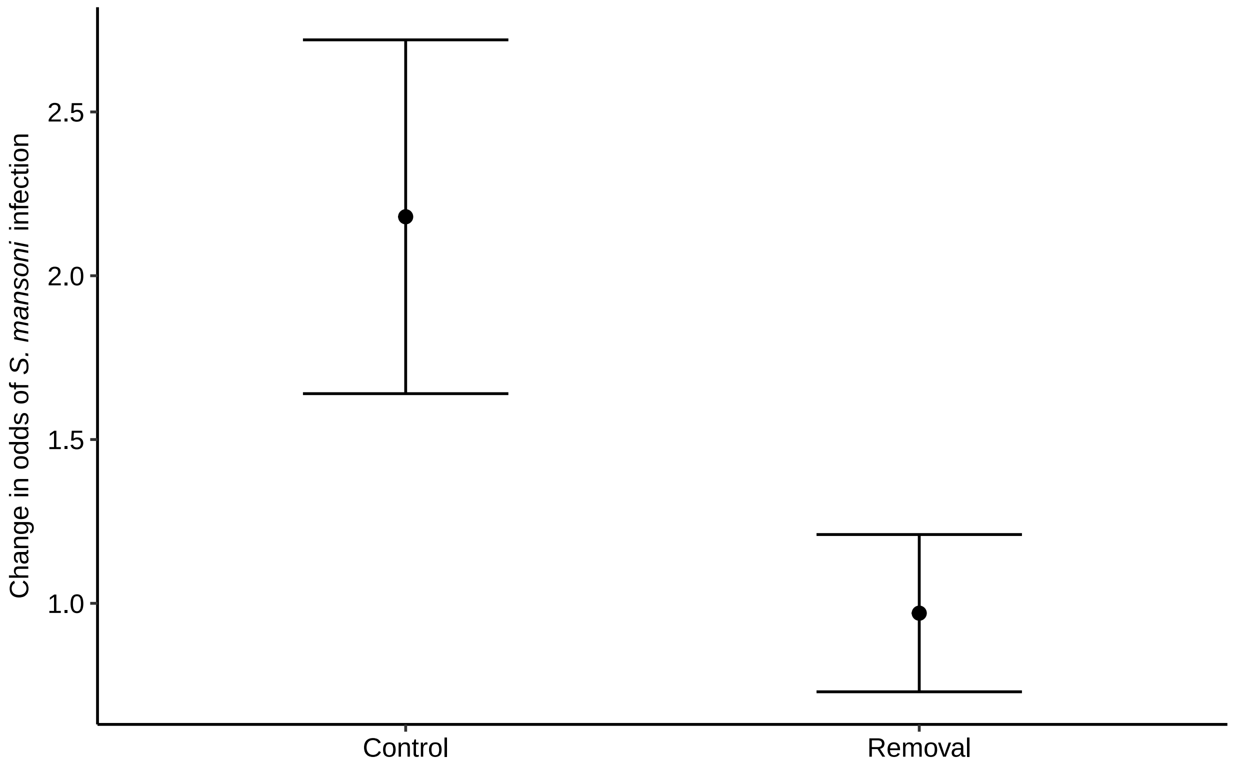


```
p <- ggplot(baciveg2, aes(x=Time, y=totalveg,color=Time)) +
  geom_violin(show.legend = FALSE)
p1 <-p + geom_boxplot(width=0.04, show.legend = FALSE, color="black")+ ylab("Aquatic vegetation mass/swe




p1 # Fig 2A
```

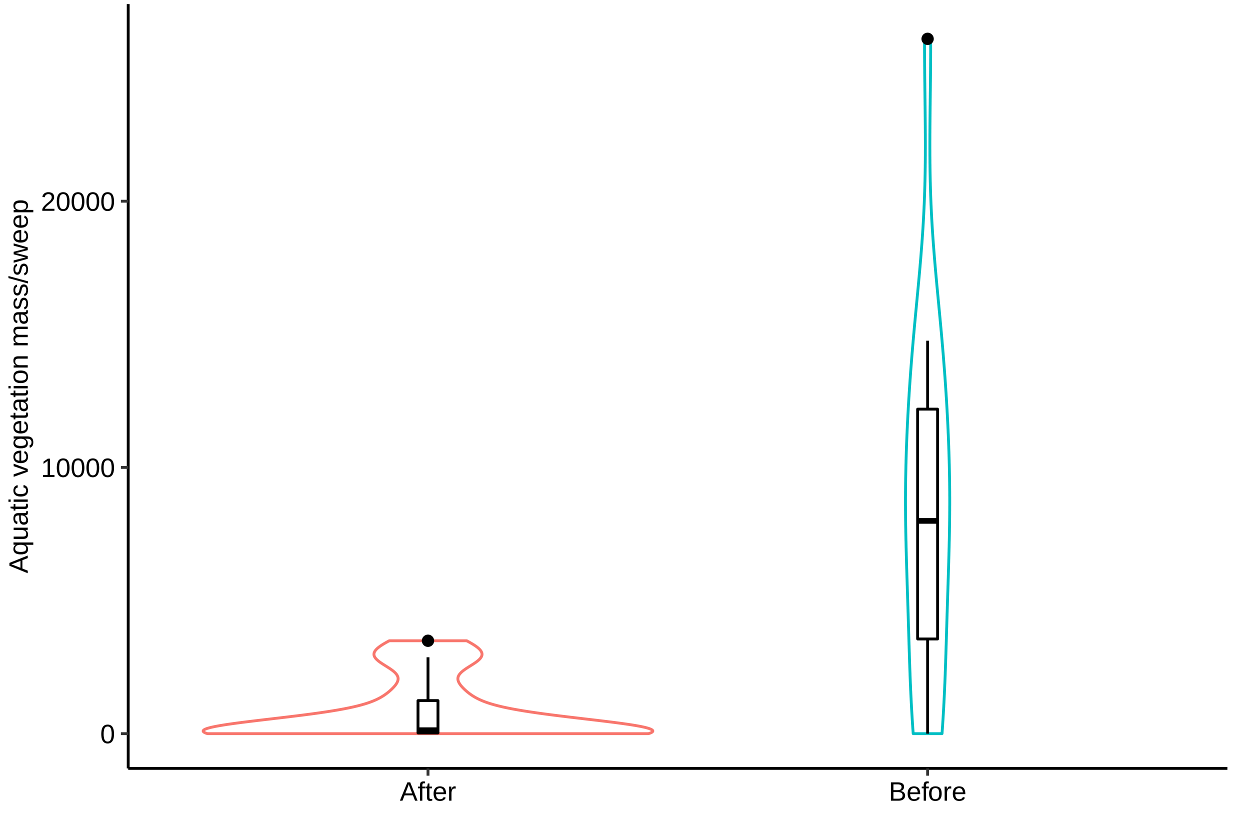


```
g1# Fig 2B
```

```
## `geom_smooth()` using formula 'y ~ x'
```

```
## Warning: Removed 4 rows containing non-finite values (stat_smooth).
```

```
## Warning: Removed 4 rows containing missing values (geom_point).
```

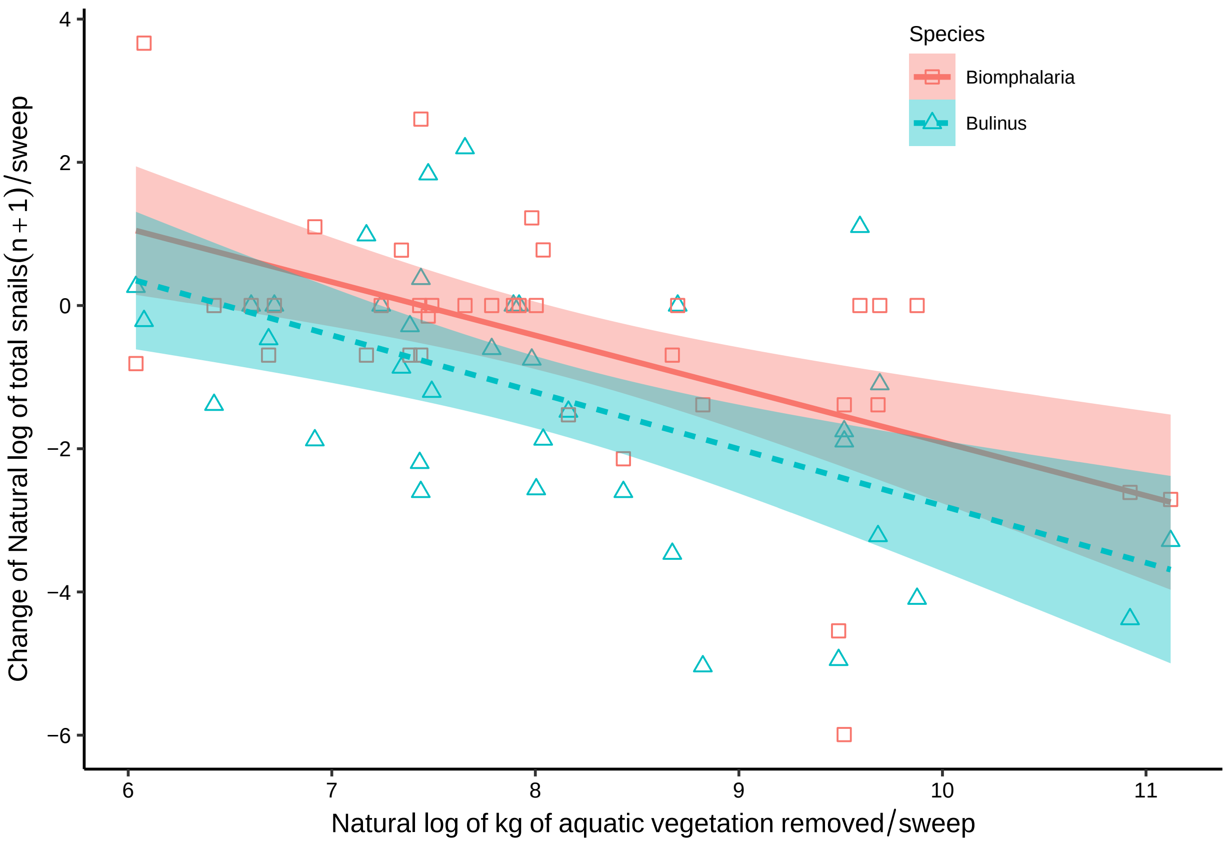
Species
Biomphalaria
Bulinus
Change of Natural log of total snails(n + 1)/sweep
Natural log of kg of aquatic vegetation removed/sweep
4
2
0
−2
−4
−6
6
7
8
9
10
11

```
e1718# Fig 2C
```

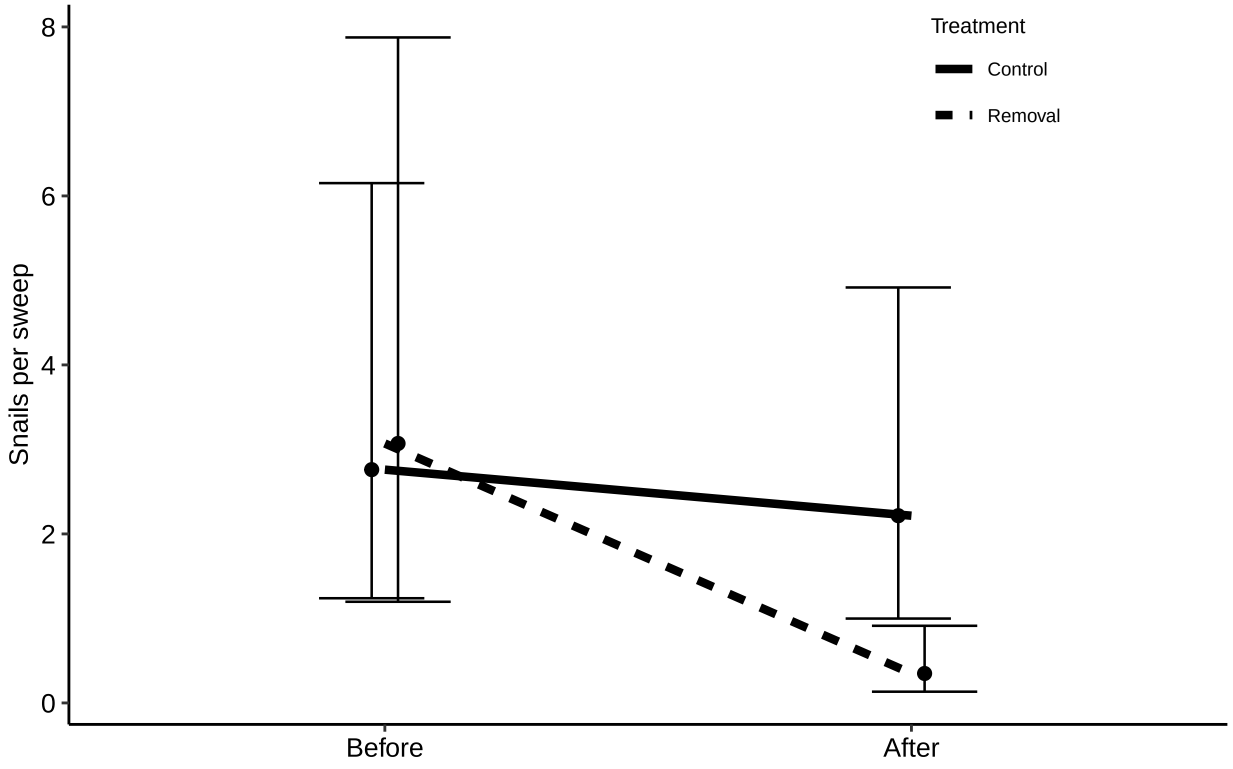


```
profileplotv4# Fig 2D
```

```
## geom_path: Each group consists of only one observation. Do you need to adjust
## the group aesthetic?

## geom_path: Each group consists of only one observation. Do you need to adjust
## the group aesthetic?
```

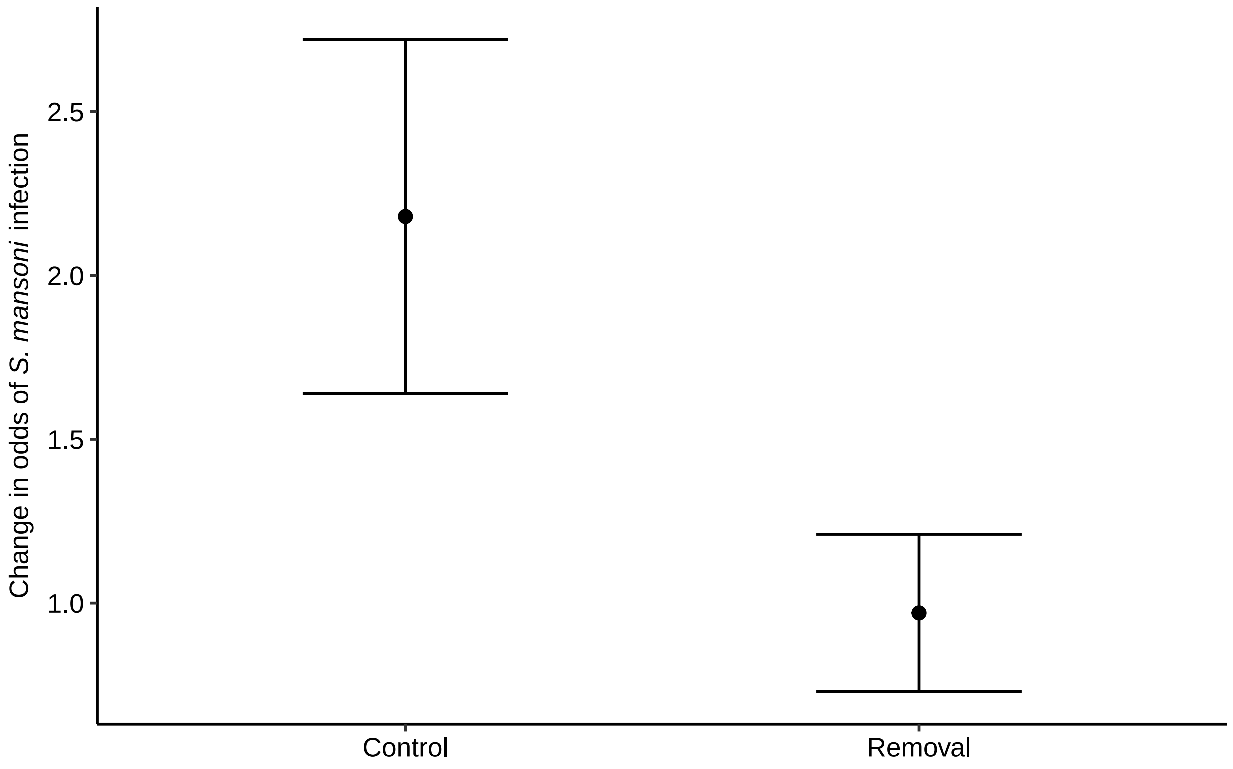


```
plot_grid(p1, g1, e1718, profileplotv4, labels = "AUTO",label_size = 11,font=11,align="vh",axis="rlbt",
```

```
## `geom_smooth()` using formula 'y ~ x'

## Warning: Removed 4 rows containing non-finite values (stat_smooth).
## Removed 4 rows containing missing values (geom_point).

## geom_path: Each group consists of only one observation. Do you need to adjust
## the group aesthetic?
## geom_path: Each group consists of only one observation. Do you need to adjust
## the group aesthetic?

## Warning in as_grob.default(plot): Cannot convert object of class numeric into a
## grob.
```

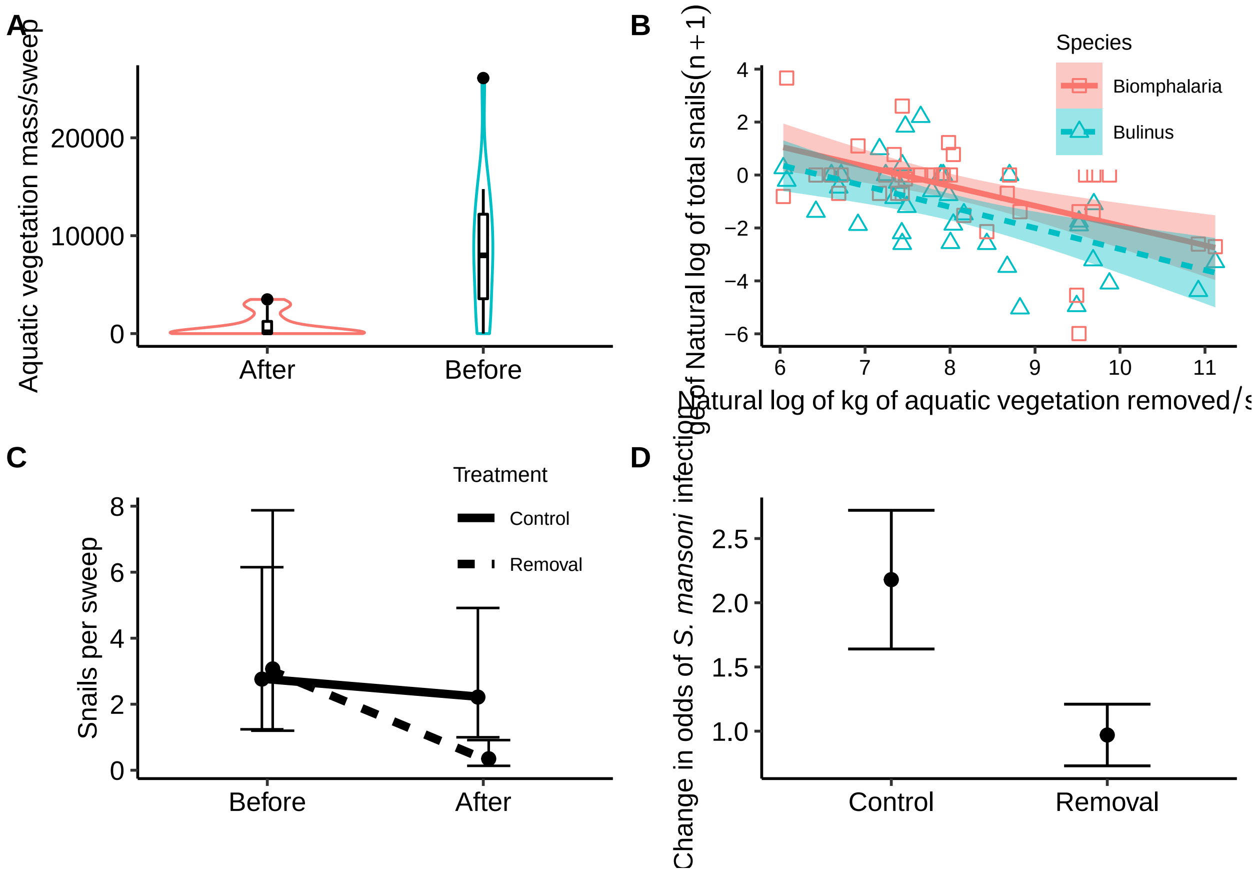


```
# Water quality check from before and after vegitation removel
water<-read.csv("Water.csv")
WaterSum <- water %>%
  group_by(Time,Treatment) %>%
  summarise(
    n=n(),
    meanT=mean(Temperature),
    sdT=sd(Temperature),
    meanC=mean(Conductivity),
    sdC=sd(Conductivity),
    meanS=mean(salinity),
    sdS=sd(salinity),
    meanO=mean(DOppt),
    sdO=sd(DOppt),
    meanpH=mean(pH),
    sdpH=sd(pH),
    meanN=mean(Nitrate),
    sdN=sd(Nitrate),
    meanPeri=mean(PeriFt),
    sdPeri=sd(PeriFt),
    meanPhy=mean(PhytoFt),
    sdPhy=sd(PhytoFt),
    ) %>%
  as.data.frame()
```

```
## `summarise()` has grouped output by 'Time'. You can override using the `.groups`
```

```
## argument.
```

```r
WaterSum
```

```
##     Time Treatment  n    meanT       sdT    meanC       sdC      meanS
## 1  After   Control 23 28.80000 1.9336729 179.8087  75.70108 0.12739130
## 2  After   Removal 16 27.65000 3.3891985 180.1375 110.30205 0.08312500
## 3 Before   Control 23 29.25217 0.9990312 194.0478  77.49791 0.08956522
## 4 Before   Removal 16 29.46875 1.5317610 206.9375 142.93878 0.09625000
##          sdS    meanO      sdO   meanpH      sdpH     meanN       sdN meanPeri
## 1 0.20885439 2.364348 2.334395       NA        NA        NA        NA       NA
## 2 0.05449388 5.647500 2.248734       NA        NA        NA        NA       NA
## 3 0.03636660 3.433478 2.467941 7.133913 0.4255557 0.4247826 0.5784295 3085.261
## 4 0.06839834 4.826250 3.477673 7.544375 1.1231501        NA        NA       NA
##     sdPeri  meanPhy    sdPhy
## 1       NA 372.4783 420.4149
## 2       NA 358.8125 283.7128
## 3 3289.714 260.5652 275.9196
## 4       NA 351.0625 259.9632
```

```r
Temp <- lmer(Temperature ~ Time*Treatment+ (1|SiteID), data=water, na.action=na.omit)
summary(Temp)
```

```
## Linear mixed model fit by REML ['lmerMod']
## Formula: Temperature ~ Time * Treatment + (1 | SiteID)
##    Data: water
##
## REML criterion at convergence: 315.5
##
## Scaled residuals:
##     Min      1Q  Median      3Q     Max
## -4.6797 -0.2844 -0.0092  0.4571  2.0380
##
## Random effects:
##  Groups   Name        Variance Std.Dev.
##  SiteID   (Intercept) 2.527    1.590
##  Residual             2.514    1.586
## Number of obs: 78, groups:  SiteID, 19
##
## Fixed effects:
##                             Estimate Std. Error t value
## (Intercept)                  28.6348     0.6023  47.541
## TimeBefore                    0.3040     0.4728   0.643
## TreatmentRemoval             -1.1624     0.9308  -1.249
## TimeBefore:TreatmentRemoval   1.2659     0.7385   1.714
##
## Correlation of Fixed Effects:
##             (Intr) TimBfr TrtmnR
## TimeBefore  -0.392
## TretmntRmvl -0.647  0.254
## TmBfr:TrtmR  0.251 -0.640 -0.406
```

```r
Anova(Temp)
```

```
## Analysis of Deviance Table (Type II Wald chisquare tests)
##
## Response: Temperature
##                 Chisq Df Pr(>Chisq)
## Time           5.1316  1    0.02349 *
## Treatment      0.3653  1    0.54559
## Time:Treatment 2.9381  1    0.08651 .
## ---
## Signif. codes:  0 '***' 0.001 '**' 0.01 '*' 0.05 '.' 0.1 ' ' 1
```

```r
Con <- lmer(Conductivity ~ Time*Treatment+ (1|SiteID), data=water, na.action=na.omit)
summary(Con)
```

```
## Linear mixed model fit by REML ['lmerMod']
## Formula: Conductivity ~ Time * Treatment + (1 | SiteID)
##    Data: water
##
## REML criterion at convergence: 800
##
## Scaled residuals:
##      Min       1Q   Median       3Q      Max
## -1.84306 -0.38618 -0.05227  0.21582  2.86216
##
## Random effects:
##  Groups   Name        Variance Std.Dev.
##  SiteID   (Intercept) 13216    114.96
##  Residual              1043     32.29
## Number of obs: 78, groups:  SiteID, 19
##
## Fixed effects:
##                              Estimate Std. Error t value
## (Intercept)                   173.976     35.521   4.898
## TimeBefore                     17.222      9.719   1.772
## TreatmentRemoval               32.876     54.766   0.600
## TimeBefore:TreatmentRemoval     9.394     15.169   0.619
##
## Correlation of Fixed Effects:
##             (Intr) TimBfr TrtmnR
## TimeBefore  -0.137
## TretmntRmvl -0.649  0.089
## TmBfr:TrtmR  0.088 -0.641 -0.145
```

```r
Anova(Con)
```

```
## Analysis of Deviance Table (Type II Wald chisquare tests)
##
## Response: Conductivity
##                 Chisq Df Pr(>Chisq)
## Time           7.9792  1   0.004732 **
## Treatment      0.4863  1   0.485602
```

```
## Time:Treatment 0.3835  1   0.535723
## ---
## Signif. codes:  0 '***' 0.001 '**' 0.01 '*' 0.05 '.' 0.1 ' ' 1
```

```
Sal <- lmer(salinity ~ Time*Treatment+ (1|SiteID), data=water, na.action=na.omit)
summary(Sal)
```

```
## Linear mixed model fit by REML ['lmerMod']
## Formula: salinity ~ Time * Treatment + (1 | SiteID)
##    Data: water
##
## REML criterion at convergence: -89.4
##
## Scaled residuals:
##     Min      1Q  Median      3Q     Max
## -0.7121 -0.4215 -0.1080  0.0803  7.7156
##
## Random effects:
##  Groups   Name        Variance  Std.Dev.
##  SiteID   (Intercept) 4.846e-05 0.006961
##  Residual             1.487e-02 0.121932
## Number of obs: 78, groups:  SiteID, 19
##
## Fixed effects:
##                               Estimate Std. Error t value
## (Intercept)                    0.12717    0.02554   4.979
## TimeBefore                    -0.03783    0.03596  -1.052
## TreatmentRemoval              -0.04394    0.03987  -1.102
## TimeBefore:TreatmentRemoval    0.05095    0.05614   0.908
##
## Correlation of Fixed Effects:
##             (Intr) TimBfr TrtmnR
## TimeBefore  -0.704
## TretmntRmvl -0.641  0.451
## TmBfr:TrtmR  0.451 -0.641 -0.704
```

```
Anova(Sal)
```

```
## Analysis of Deviance Table (Type II Wald chisquare tests)
##
## Response: salinity
##                 Chisq Df Pr(>Chisq)
## Time           0.3757  1     0.5399
## Treatment      0.4254  1     0.5143
## Time:Treatment 0.8238  1     0.3641
```

```
Ox <- lmer(DOppt ~ Time*Treatment+ (1|SiteID), data=water, na.action=na.omit)
summary(Ox)
```

```
## Linear mixed model fit by REML ['lmerMod']
## Formula: DOppt ~ Time * Treatment + (1 | SiteID)
##    Data: water
```

```
##
## REML criterion at convergence: 357.9
##
## Scaled residuals:
##     Min      1Q  Median      3Q     Max
## -2.4946 -0.5886 -0.1561  0.6406  2.0467
##
## Random effects:
##  Groups   Name        Variance Std.Dev.
##  SiteID   (Intercept) 1.717    1.310
##  Residual             5.256    2.293
## Number of obs: 78, groups:  SiteID, 19
##
## Fixed effects:
##                             Estimate Std. Error t value
## (Intercept)                   2.4205     0.6434   3.762
## TimeBefore                    1.0321     0.6797   1.518
## TreatmentRemoval              2.9168     0.9968   2.926
## TimeBefore:TreatmentRemoval  -1.8344     1.0619  -1.727
##
## Correlation of Fixed Effects:
##             (Intr) TimBfr TrtmnR
## TimeBefore  -0.528
## TretmntRmvl -0.645  0.341
## TmBfr:TrtmR  0.338 -0.640 -0.539
```

```
Anova(Ox)
```

```
## Analysis of Deviance Table (Type II Wald chisquare tests)
##
## Response: DOppt
##                 Chisq Df Pr(>Chisq)
## Time           0.2886  1    0.59109
## Treatment      5.6108  1    0.01785 *
## Time:Treatment 2.9841  1    0.08408 .
## ---
## Signif. codes:  0 '***' 0.001 '**' 0.01 '*' 0.05 '.' 0.1 ' ' 1
```

```
pH <- lmer(pH ~ Time*Treatment+ (1|SiteID), data=water, na.action=na.omit)
summary(pH)
```

```
## Linear mixed model fit by REML ['lmerMod']
## Formula: pH ~ Time * Treatment + (1 | SiteID)
##    Data: water
##
## REML criterion at convergence: 147.3
##
## Scaled residuals:
##      Min       1Q   Median       3Q      Max
## -2.36730 -0.59950 -0.06688  0.50840  2.48252
##
## Random effects:
##  Groups   Name        Variance Std.Dev.
```

```
##  SiteID   (Intercept) 0.1438   0.3792
##  Residual             0.3076   0.5547
## Number of obs: 76, groups:  SiteID, 19
##
## Fixed effects:
##                              Estimate Std. Error t value
## (Intercept)                   6.83095    0.17323  39.434
## TimeBefore                    0.28091    0.16722   1.680
## TreatmentRemoval              0.27354    0.26988   1.014
## TimeBefore:TreatmentRemoval  0.01612    0.26244   0.061
##
## Correlation of Fixed Effects:
##             (Intr) TimBfr TrtmnR
## TimeBefore  -0.507
## TretmntRmvl -0.642  0.325
## TmBfr:TrtmR  0.323 -0.637 -0.523
```

```
Anova(pH)
```

```
## Analysis of Deviance Table (Type II Wald chisquare tests)
##
## Response: pH
##                 Chisq Df Pr(>Chisq)
## Time           4.9748  1    0.02572 *
## Treatment      1.5054  1    0.21984
## Time:Treatment 0.0038  1    0.95104
## ---
## Signif. codes:  0 '***' 0.001 '**' 0.01 '*' 0.05 '.' 0.1 ' ' 1
```

```
water2=subset(water, is.na(water$PeriFt)==FALSE)
WaterSum2 <- water2 %>%
  group_by(Time,Treatment) %>%
  summarise(
    n=n(),
    meanT=mean(Temperature),
    sdT=sd(Temperature),
    meanC=mean(Conductivity),
    sdC=sd(Conductivity),
    meanS=mean(salinity),
    sdS=sd(salinity),
    meanO=mean(DOppt),
    sdO=sd(DOppt),
    meanpH=mean(pH),
    sdpH=sd(pH),
    meanN=mean(Nitrate),
    sdN=sd(Nitrate),
    meanPeri=mean(PeriFt),
    sdPeri=sd(PeriFt),
    meanPhy=mean(PhytoFt),
    sdPhy=sd(PhytoFt),
  ) %>%
  as.data.frame()
```

```
## `summarise()` has grouped output by 'Time'. You can override using the `.groups`
```

```
## argument.
```

```r
WaterSum2
```

```
##     Time Treatment  n    meanT       sdT    meanC       sdC      meanS
## 1  After   Control 16 28.60000 2.2603834 194.4313  87.44121 0.09125000
## 2  After   Removal 15 27.55333 3.4852478 187.2067 110.35804 0.08666667
## 3 Before   Control 23 29.25217 0.9990312 194.0478  77.49791 0.08956522
## 4 Before   Removal 12 29.05000 1.3338257 222.1583 160.44247 0.10416667
##          sdS    meanO      sdO   meanpH      sdpH     meanN       sdN meanPeri
## 1 0.04318565 2.903125 2.567919       NA        NA        NA        NA 3862.438
## 2 0.05446712 5.748667 2.289663       NA        NA        NA        NA 2476.067
## 3 0.03636660 3.433478 2.467941 7.133913 0.4255557 0.4247826 0.5784295 3085.261
## 4 0.07668807 4.554167 3.202435 7.384167 1.0334449        NA        NA 1869.167
##     sdPeri  meanPhy    sdPhy
## 1 3153.113 484.4375 463.5370
## 2 3790.612 372.6000 288.0689
## 3 3289.714 260.5652 275.9196
## 4 2381.423 385.2500 294.2785
```

```r
Nit <- lmer(Nitrate ~ Time*Treatment+ (1|SiteID), data=water, na.action=na.omit)
summary(Nit)
```

```
## Linear mixed model fit by REML ['lmerMod']
## Formula: Nitrate ~ Time * Treatment + (1 | SiteID)
##    Data: water
##
## REML criterion at convergence: 84.2
##
## Scaled residuals:
##     Min      1Q  Median      3Q     Max
## -1.2442 -0.5002 -0.2086  0.0760  3.2777
##
## Random effects:
##  Groups   Name        Variance Std.Dev.
##  SiteID   (Intercept) 0.1100   0.3316
##  Residual             0.2436   0.4936
## Number of obs: 49, groups:  SiteID, 17
##
## Fixed effects:
##                             Estimate Std. Error t value
## (Intercept)                  0.81092    0.23316   3.478
## TimeBefore                  -0.36030    0.22777  -1.582
## TreatmentRemoval            -0.18190    0.38875  -0.468
## TimeBefore:TreatmentRemoval  0.03395    0.38311   0.089
##
## Correlation of Fixed Effects:
##             (Intr) TimBfr TrtmnR
## TimeBefore  -0.785
## TretmntRmvl -0.600  0.471
## TmBfr:TrtmR  0.467 -0.595 -0.804
```

```r
Anova(Nit)
```

```
## Analysis of Deviance Table (Type II Wald chisquare tests)
##
## Response: Nitrate
##                 Chisq Df Pr(>Chisq)
## Time           3.6169  1     0.0572 .
## Treatment      0.4457  1     0.5044
## Time:Treatment 0.0079  1     0.9294
## ---
## Signif. codes:  0 '***' 0.001 '**' 0.01 '*' 0.05 '.' 0.1 ' ' 1
```

```r
Phyto <- lmer(PhytoFt ~ Time*Treatment+ (1|SiteID), data=water, na.action=na.omit)
summary(Phyto)
```

```
## Linear mixed model fit by REML ['lmerMod']
## Formula: PhytoFt ~ Time * Treatment + (1 | SiteID)
##    Data: water
##
## REML criterion at convergence: 1065.9
##
## Scaled residuals:
##     Min      1Q  Median      3Q     Max
## -2.4347 -0.3685 -0.1336  0.1000  5.0688
##
## Random effects:
##  Groups   Name        Variance Std.Dev.
##  SiteID   (Intercept) 39723    199.3
##  Residual             69767    264.1
## Number of obs: 78, groups:  SiteID, 19
##
## Fixed effects:
##                              Estimate Std. Error t value
## (Intercept)                    362.30      84.65   4.280
## TimeBefore                    -113.05      78.51  -1.440
## TreatmentRemoval                38.37     130.96   0.293
## TimeBefore:TreatmentRemoval     97.29     122.66   0.793
##
## Correlation of Fixed Effects:
##             (Intr) TimBfr TrtmnR
## TimeBefore  -0.464
## TretmntRmvl -0.646  0.300
## TmBfr:TrtmR  0.297 -0.640 -0.476
```

```r
Anova(Phyto)
```

```
## Analysis of Deviance Table (Type II Wald chisquare tests)
##
## Response: PhytoFt
##                 Chisq Df Pr(>Chisq)
## Time           1.4723  1     0.2250
## Treatment      0.5822  1     0.4455
## Time:Treatment 0.6291  1     0.4277
```

```r
Peri <- lmer(PeriFt ~ Time*Treatment+ (1|SiteID), data=water, na.action=na.omit)
summary(Peri)
```

```
## Linear mixed model fit by REML ['lmerMod']
## Formula: PeriFt ~ Time * Treatment + (1 | SiteID)
##    Data: water
##
## REML criterion at convergence: 1187.1
##
## Scaled residuals:
##     Min      1Q  Median      3Q     Max
## -1.1593 -0.6336 -0.2488  0.3641  3.6589
##
## Random effects:
##  Groups   Name        Variance Std.Dev.
##  SiteID   (Intercept) 1925831  1388
##  Residual             8772784  2962
## Number of obs: 66, groups:  SiteID, 19
##
## Fixed effects:
##                              Estimate Std. Error t value
## (Intercept)                   3813.79     895.88   4.257
## TimeBefore                    -587.57     998.80  -0.588
## TreatmentRemoval             -1217.12    1298.93  -0.937
## TimeBefore:TreatmentRemoval     89.64    1531.61   0.059
##
## Correlation of Fixed Effects:
##             (Intr) TimBfr TrtmnR
## TimeBefore  -0.673
## TretmntRmvl -0.690  0.464
## TmBfr:TrtmR  0.439 -0.652 -0.612
```

```r
Anova(Peri)
```

```
## Analysis of Deviance Table (Type II Wald chisquare tests)
##
## Response: PeriFt
##                 Chisq Df Pr(>Chisq)
## Time           0.5265  1     0.4681
## Treatment      1.2995  1     0.2543
## Time:Treatment 0.0034  1     0.9533
```

```r
#SHEEP Preliminary Results
All_Sheep<-read.csv("All_Sheep.csv")

#linear regression for final weight after accounting for percent cerato as a continuous variable by Age
AS1 <- lm(FW~Group*Age, data= All_Sheep)
summary(AS1)
```

```
##
## Call:
## lm(formula = FW ~ Group * Age, data = All_Sheep)
```

```
##
## Residuals:
##     Min      1Q  Median      3Q     Max
## -6.7116 -3.6437  0.2884  2.5608  8.2884
##
## Coefficients:
##              Estimate Std. Error t value Pr(>|t|)
## (Intercept) 22.958777   2.139162  10.733 4.77e-11 ***
## Group        0.008865   5.577180   0.002    0.999
## AgeJV        0.032691   2.923773   0.011    0.991
## Group:AgeJV -1.874622   7.563334  -0.248    0.806
## ---
## Signif. codes:  0 '***' 0.001 '**' 0.01 '*' 0.05 '.' 0.1 ' ' 1
##
## Residual standard error: 4.499 on 26 degrees of freedom
## Multiple R-squared:  0.00973,    Adjusted R-squared:  -0.1045
## F-statistic: 0.08515 on 3 and 26 DF,  p-value: 0.9675
```

```
Anova(AS1)
```

```
## Anova Table (Type II tests)
##
## Response: FW
##           Sum Sq Df F value Pr(>F)
## Group       1.46  1  0.0719 0.7906
## Age         2.34  1  0.1157 0.7365
## Group:Age   1.24  1  0.0614 0.8062
## Residuals 526.30 26
```

```
# Checking normal errors
par(mfrow=c(2,2))
hist(AS1$resid)
plot(AS1$resid~AS1$fitted)
lines(lowess(AS1$resid~AS1$fitted))
plot(All_Sheep$FW~All_Sheep$Group)
lines(lowess(All_Sheep$FW~All_Sheep$Group))

All_Sheep$SW<-All_Sheep$`1`

#Summary of Starting Weight
StartSheep <- All_Sheep %>%
  group_by(Group,Age) %>%
  summarise(
    n=n(),
    mean=mean(FW),
    sd=sd(FW),
    min=min(FW),
    max=max(FW)
    )
```

```
## `summarise()` has grouped output by 'Group'. You can override using the
## `.groups` argument.
```

```r
#Summary of All Weights
All_SheepL <- gather(All_Sheep, date, weight, "X1":"X14", factor_key=TRUE)
AllWeeks <- All_SheepL %>%
  group_by(Group,Age) %>%
  summarise(
    n=n(),
    mean=mean(weight),
    sd=sd(weight),
    min=min(weight),
    max=max(weight)
  )
```

```
## `summarise()` has grouped output by 'Group'. You can override using the
## `.groups` argument.
```

```r
#Ending Results
End <- All_Sheep %>%
  group_by(Group,Age) %>%
  summarise(
    n=n(),
    mean=mean(FW),
    sd=sd(FW)
  ) %>%
  mutate( se=sd/sqrt(n))  %>%
  mutate( ic=se * qt((1-0.05)/2 + .5, n-1))
```

```
## `summarise()` has grouped output by 'Group'. You can override using the
## `.groups` argument.
```

```r
# End Percent Diff
AS2 <- lm(Diff~Group*Age, data= All_Sheep)
summary(AS2)
```

```
##
## Call:
## lm(formula = Diff ~ Group * Age, data = All_Sheep)
##
## Residuals:
##    Min     1Q Median     3Q    Max
## -4.457 -1.373 -0.032  1.546  5.705
##
## Coefficients:
##             Estimate Std. Error t value Pr(>|t|)
## (Intercept)    5.471      1.052   5.201 1.98e-05 ***
## Group         -5.585      2.742  -2.037    0.052 .
## AgeJV         -1.641      1.438  -1.141    0.264
## Group:AgeJV    4.123      3.719   1.109    0.278
## ---
## Signif. codes:  0 '***' 0.001 '**' 0.01 '*' 0.05 '.' 0.1 ' ' 1
##
## Residual standard error: 2.212 on 26 degrees of freedom
## Multiple R-squared:  0.1531, Adjusted R-squared:  0.05543
## F-statistic: 1.567 on 3 and 26 DF,  p-value: 0.2211
```

```r
Anova(AS2)
```

```
## Anova Table (Type II tests)
##
## Response: Diff
##            Sum Sq Df F value  Pr(>F)
## Group      15.941  1  3.2576 0.08269 .
## Age         0.791  1  0.1617 0.69090
## Group:Age   6.016  1  1.2294 0.27768
## Residuals 127.233 26
## ---
## Signif. codes:  0 '***' 0.001 '**' 0.01 '*' 0.05 '.' 0.1 ' ' 1
```

```r
# Start Weight
AS1 <- lm(FW~Group*Age, data= All_Sheep)
summary(AS1)
```

```
##
## Call:
## lm(formula = FW ~ Group * Age, data = All_Sheep)
##
## Residuals:
##     Min      1Q  Median      3Q     Max
## -6.7116 -3.6437  0.2884  2.5608  8.2884
##
## Coefficients:
##                Estimate Std. Error t value Pr(>|t|)
## (Intercept) 22.958777   2.139162  10.733 4.77e-11 ***
## Group        0.008865   5.577180   0.002    0.999
## AgeJV        0.032691   2.923773   0.011    0.991
## Group:AgeJV -1.874622   7.563334  -0.248    0.806
## ---
## Signif. codes:  0 '***' 0.001 '**' 0.01 '*' 0.05 '.' 0.1 ' ' 1
##
## Residual standard error: 4.499 on 26 degrees of freedom
## Multiple R-squared:  0.00973,    Adjusted R-squared:  -0.1045
## F-statistic: 0.08515 on 3 and 26 DF,  p-value: 0.9675
```

```r
Anova(AS1)
```

```
## Anova Table (Type II tests)
##
## Response: FW
##           Sum Sq Df F value Pr(>F)
## Group       1.46  1  0.0719 0.7906
## Age         2.34  1  0.1157 0.7365
## Group:Age   1.24  1  0.0614 0.8062
## Residuals 526.30 26
```

```r
#  All Weights
All_SheepL$date<-as.numeric(All_SheepL$date)
ASM1 <- lmer(weight ~ Group*Age + (1 | ID), data = All_SheepL, na.action=na.omit)
summary(ASM1)
```

```
## Linear mixed model fit by REML ['lmerMod']
## Formula: weight ~ Group * Age + (1 | ID)
##    Data: All_SheepL
##
## REML criterion at convergence: 1747.6
##
## Scaled residuals:
##     Min      1Q  Median      3Q     Max
## -3.9252 -0.4931  0.0418  0.5637  2.7543
##
## Random effects:
##  Groups   Name        Variance Std.Dev.
##  ID       (Intercept) 16.56    4.069
##  Residual              2.86    1.691
## Number of obs: 420, groups:  ID, 30
##
## Fixed effects:
##             Estimate Std. Error t value
## (Intercept)  21.9861     1.9466  11.295
## Group         1.0322     5.0751   0.203
## AgeJV        -0.7954     2.6606  -0.299
## Group:AgeJV  -2.5944     6.8824  -0.377
##
## Correlation of Fixed Effects:
##             (Intr) Group  AgeJV
## Group       -0.812
## AgeJV       -0.732  0.594
## Group:AgeJV  0.599 -0.737 -0.824
```

```
Anova(ASM1)
```

```
## Analysis of Deviance Table (Type II Wald chisquare tests)
##
## Response: weight
##            Chisq Df Pr(>Chisq)
## Group     0.0122  1     0.9121
## Age       1.1540  1     0.2827
## Group:Age 0.1421  1     0.7062
```

```
library(jtools)
```

```
## Warning: package 'jtools' was built under R version 4.1.2
```

```
effect_plot(ASM1, pred = Group, interval = TRUE, plot.points = TRUE)
```

```
## Confidence intervals for merMod models is an experimental feature. The
## intervals reflect only the variance of the fixed effects, not the random
## effects.
```

```
#examine interaction
ASM2 <- lmer(weight ~ as.factor(Group)*Age*date + (1 | ID), data = All_SheepL, na.action=na.omit)
summary(ASM2)
```

```
## Linear mixed model fit by REML ['lmerMod']
## Formula: weight ~ as.factor(Group) * Age * date + (1 | ID)
##    Data: All_SheepL
##
## REML criterion at convergence: 1655.7
##
## Scaled residuals:
##     Min      1Q  Median      3Q     Max
## -5.2001 -0.4452  0.0305  0.4799  3.4343
##
## Random effects:
##  Groups   Name        Variance Std.Dev.
##  ID       (Intercept) 19.438   4.409
##  Residual              2.238   1.496
## Number of obs: 420, groups:  ID, 30
##
## Fixed effects:
##                                Estimate Std. Error t value
## (Intercept)                    20.76740    2.59176   8.013
## as.factor(Group)0.15            0.15842    4.09794   0.039
## as.factor(Group)0.3            -0.08059    4.09794  -0.020
## as.factor(Group)0.45            0.35348    3.66531   0.096
## as.factor(Group)0.6             2.32967    3.66531   0.636
## AgeJV                          -2.45971    3.66531  -0.671
## date                            0.18974    0.05726   3.313
## as.factor(Group)0.15:AgeJV     -0.05037    5.49796  -0.009
## as.factor(Group)0.3:AgeJV       2.45604    5.49796   0.447
## as.factor(Group)0.45:AgeJV      1.87454    5.01893   0.373
## as.factor(Group)0.6:AgeJV      -3.24588    5.01893  -0.647
## as.factor(Group)0.15:date      -0.04414    0.09054  -0.487
## as.factor(Group)0.3:date        0.09487    0.09054   1.048
## as.factor(Group)0.45:date      -0.18205    0.08098  -2.248
## as.factor(Group)0.6:date       -0.14872    0.08098  -1.836
## AgeJV:date                      0.12637    0.08098   1.560
## as.factor(Group)0.15:AgeJV:date -0.08773    0.12148  -0.722
## as.factor(Group)0.3:AgeJV:date  -0.19414    0.12148  -1.598
## as.factor(Group)0.45:AgeJV:date  0.09451    0.11089   0.852
## as.factor(Group)0.6:AgeJV:date  -0.01245    0.11089  -0.112

##
## Correlation matrix not shown by default, as p = 20 > 12.
## Use print(x, correlation=TRUE)  or
##     vcov(x)        if you need it
```

```
Anova(ASM2)
```

```
## Analysis of Deviance Table (Type II Wald chisquare tests)
##
## Response: weight
##                            Chisq Df Pr(>Chisq)
## as.factor(Group)          0.7238  4    0.94837
## Age                       1.0069  1    0.31564
## date                     93.8037  1    < 2e-16 ***
```

```
## as.factor(Group):Age       1.6448  4    0.80071
## as.factor(Group):date     11.2372  4    0.02402 *
## Age:date                   7.3624  1    0.00666 **
## as.factor(Group):Age:date  6.5782  4    0.15993
## ---
## Signif. codes:  0 '***' 0.001 '**' 0.01 '*' 0.05 '.' 0.1 ' ' 1
```

```
ASM2b <- lmer(weight ~ Group*Age + (1 | ID), data = All_SheepL, na.action=na.omit)
summary(ASM2b)
```

```
## Linear mixed model fit by REML ['lmerMod']
## Formula: weight ~ Group * Age + (1 | ID)
##    Data: All_SheepL
##
## REML criterion at convergence: 1747.6
##
## Scaled residuals:
##     Min      1Q  Median      3Q     Max
## -3.9252 -0.4931  0.0418  0.5637  2.7543
##
## Random effects:
##  Groups   Name        Variance Std.Dev.
##  ID       (Intercept) 16.56    4.069
##  Residual              2.86    1.691
## Number of obs: 420, groups:  ID, 30
##
## Fixed effects:
##             Estimate Std. Error t value
## (Intercept)  21.9861     1.9466  11.295
## Group         1.0322     5.0751   0.203
## AgeJV        -0.7954     2.6606  -0.299
## Group:AgeJV  -2.5944     6.8824  -0.377
##
## Correlation of Fixed Effects:
##             (Intr) Group  AgeJV
## Group       -0.812
## AgeJV       -0.732  0.594
## Group:AgeJV  0.599 -0.737 -0.824
```

```
Anova(ASM2b)
```

```
## Analysis of Deviance Table (Type II Wald chisquare tests)
##
## Response: weight
##            Chisq Df Pr(>Chisq)
## Group     0.0122  1     0.9121
## Age       1.1540  1     0.2827
## Group:Age 0.1421  1     0.7062
```

```
#Figure 3 E
SheepAD <- subset(All_Sheep, Age=="A")
SheepJV<- subset(All_Sheep, Age=="JV")
```

```r
Ad1 <- lm(FW ~ Group, data = SheepAD, na.action=na.omit)
summary(Ad1)
```

```
##
## Call:
## lm(formula = FW ~ Group, data = SheepAD, na.action = na.omit)
##
## Residuals:
##    Min     1Q Median     3Q    Max
## -5.964 -3.963  1.039  2.539  7.541
##
## Coefficients:
##              Estimate Std. Error t value Pr(>|t|)
## (Intercept) 22.958777   2.352239   9.760 9.42e-07 ***
## Group        0.008865   6.132710   0.001    0.999
## ---
## Signif. codes:  0 '***' 0.001 '**' 0.01 '*' 0.05 '.' 0.1 ' ' 1
##
## Residual standard error: 4.947 on 11 degrees of freedom
## Multiple R-squared:  1.9e-07,    Adjusted R-squared:  -0.09091
## F-statistic: 2.09e-06 on 1 and 11 DF,  p-value: 0.9989
```

```r
Anova(Ad1)
```

```
## Anova Table (Type II tests)
##
## Response: FW
##           Sum Sq Df F value Pr(>F)
## Group       0.00  1       0 0.9989
## Residuals 269.23 11
```

```r
SA <- effect_plot(Ad1, pred = Group, interval = TRUE, plot.points = TRUE,jitter = c(0.03, 0)) +
 ylab("Adult Mean Final Weight (kg)")+ xlab("Proportion Cerato in Feed (Treatment Group)") +theme(axis.



SA

JV1 <- lm(FW ~ Group, data = SheepJV, na.action=na.omit)
summary(JV1)
```

```
##
## Call:
## lm(formula = FW ~ Group, data = SheepJV, na.action = na.omit)
##
## Residuals:
##     Min      1Q  Median      3Q     Max
## -6.7116 -1.9915  0.0085  2.5683  8.2884
##
```

```
## Coefficients:
##             Estimate Std. Error t value Pr(>|t|)    
## (Intercept)   22.991      1.834  12.537 2.37e-09 ***
## Group         -1.866      4.701  -0.397    0.697    
## ---
## Signif. codes:  0 '***' 0.001 '**' 0.01 '*' 0.05 '.' 0.1 ' ' 1
##
## Residual standard error: 4.14 on 15 degrees of freedom
## Multiple R-squared:  0.01039,    Adjusted R-squared:  -0.05558 
## F-statistic: 0.1575 on 1 and 15 DF,  p-value: 0.697
```

```
Anova(JV1)
```

```
## Anova Table (Type II tests)
##
## Response: FW
##           Sum Sq Df F value Pr(>F)
## Group       2.70  1  0.1575  0.697
## Residuals 257.06 15
```

```
SJ <- effect_plot(JV1, pred = Group, interval = TRUE, plot.points = TRUE, jitter = c(0.03, 0)) +
 ylab("Juvenile Mean Final Weight (kg)")+ xlab("Proportion Cerato in Feed (Treatment Group)") +theme(ax





SJ
library(cowplot)
plot_grid(SA,SJ,align="vh",axis="rlbt",nrow=2,ncol=1,label_x = -.005)
```

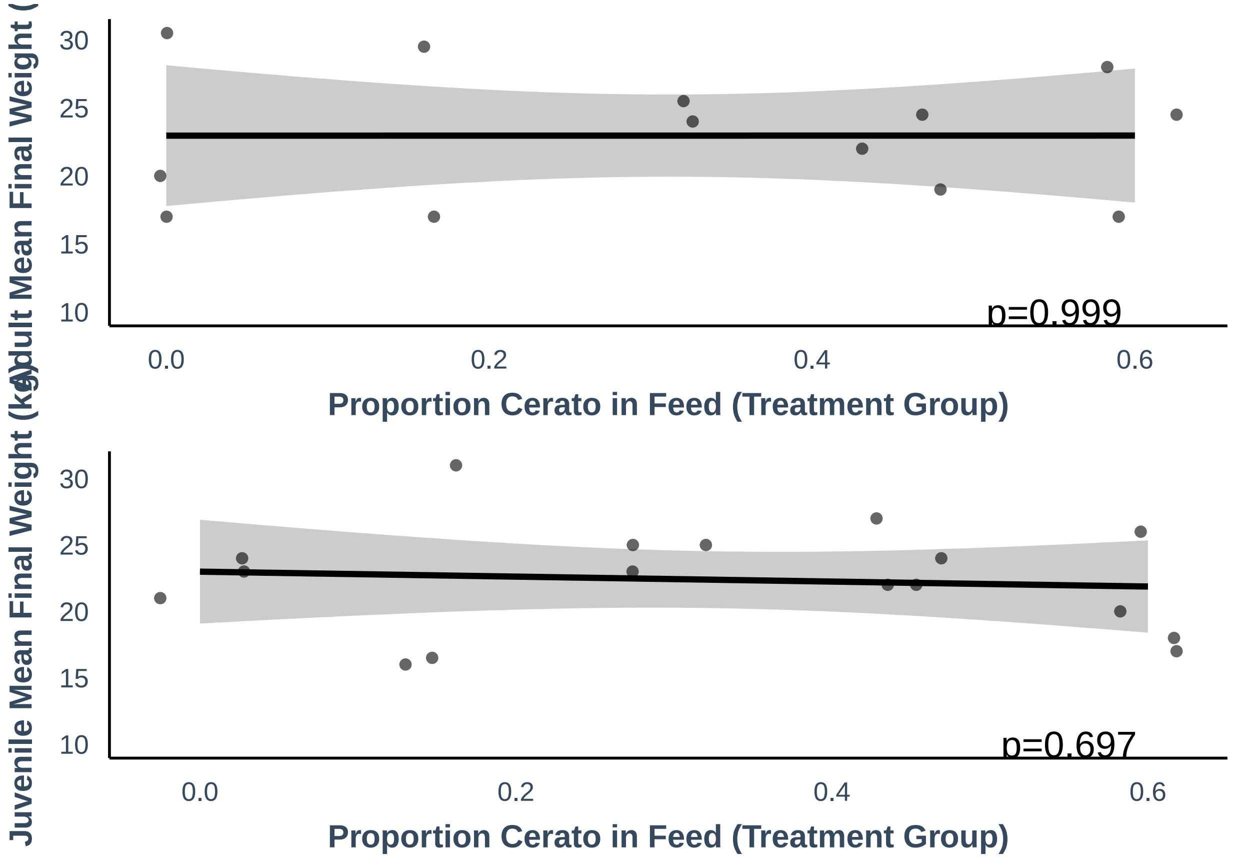
Adult Mean Final Weight (
30
25
20
15
10
p=0.999
0.0
0.2
0.4
0.6
Proportion Cerato in Feed (Treatment Group)
Juvenile Mean Final Weight (kg)
30
25
20
15
10
p=0.697
0.0
0.2
0.4
0.6
Proportion Cerato in Feed (Treatment Group)
