## Supplementary material for "A planetary health solution for disease, sustainability, food, water, and poverty challenges": Ag. Correlation and Path Model RMarkdown

### AGRICULTURE - CORRELATION

Chris and Lexi

7/26/2022

```r
#Packages:
library(plyr)
library(car)
```

```
## Loading required package: carData
```

```r
library(lme4)
```

```
## Loading required package: Matrix
```

```r
library(visreg)
library(MASS)
library(lattice)
library(rmarkdown)
```

```
## Warning: package 'rmarkdown' was built under R version 4.1.2
```

```r
library(fmsb)
```

```
## Warning: package 'fmsb' was built under R version 4.1.2
```

```r
library(ggtext)
library(ggplot2)
library(PerformanceAnalytics)
```

```
## Loading required package: xts

## Loading required package: zoo

##
## Attaching package: 'zoo'

## The following objects are masked from 'package:base':
##
##     as.Date, as.Date.numeric

##
## Attaching package: 'PerformanceAnalytics'
```

```
## The following object is masked from 'package:graphics':
##
##     legend
```

```
library(AICcmodavg)
```

```
##
## Attaching package: 'AICcmodavg'

## The following object is masked from 'package:lme4':
##
##     checkConv
```

```
library(ciTools)
```

```
## ciTools version 0.6.1 (C) Institute for Defense Analyses
```

```
library(piecewiseSEM)
```

```
##
##   This is piecewiseSEM version 2.1.0.
##
##
##   Questions or bugs can be addressed to <>.
```

```
library(nlme)
```

```
##
## Attaching package: 'nlme'

## The following object is masked from 'package:lme4':
##
##     lmList
```

```
library(gridExtra)
library(glmmTMB)
```

```
## Warning in checkDepPackageVersion(dep_pkg = "TMB"): Package version inconsistency detected.
## glmmTMB was built with TMB version 1.7.21
## Current TMB version is 1.7.22
## Please re-install glmmTMB from source or restore original 'TMB' package (see '?reinstalling' for more
```

```
library(jtools)
```

```
## Warning: package 'jtools' was built under R version 4.1.2
```

```
library (cowplot)
library(olsrr)
```

```
##
## Attaching package: 'olsrr'

## The following object is masked from 'package:MASS':
##
##     cement

## The following object is masked from 'package:datasets':
##
##     rivers
```

```
library(fmsb)
library(ggtext)
```

```
#===============================================
  #### AGRICULTURE -CORRELATION ANALYSES #####
#================================================

# DATA PREPARATION:


landp1 <- read.csv("landuse_stanford_and_our7village_weightedmeans_1_13.csv")
landp1$prevbase=landp1$prevalence_baseline
landp1$prevre1=landp1$prevalence_reinfection_2017/100
landp1$prevre2=landp1$prevalence_reinfection2018/100


###PLOT REINFECTION 1 to BASELINE:###  Fig. S1A
ggplot(landp1, aes(x=prevbase, y=prevre1)) + geom_point(size=2)+
  geom_smooth(method=lm,
              color="black", fill="gray")+
  labs(title="",
       x="Baseline prevalence 2016", y = "Re-infection prevalence 2017")+
  theme_classic(base_size = 16)  + scale_y_continuous(
    labels = scales::number_format(accuracy = 0.1,
                                   decimal.mark = '.'))
```

```
## `geom_smooth()` using formula 'y ~ x'

## Warning: Removed 7 rows containing non-finite values (stat_smooth).

## Warning: Removed 7 rows containing missing values (geom_point).
```

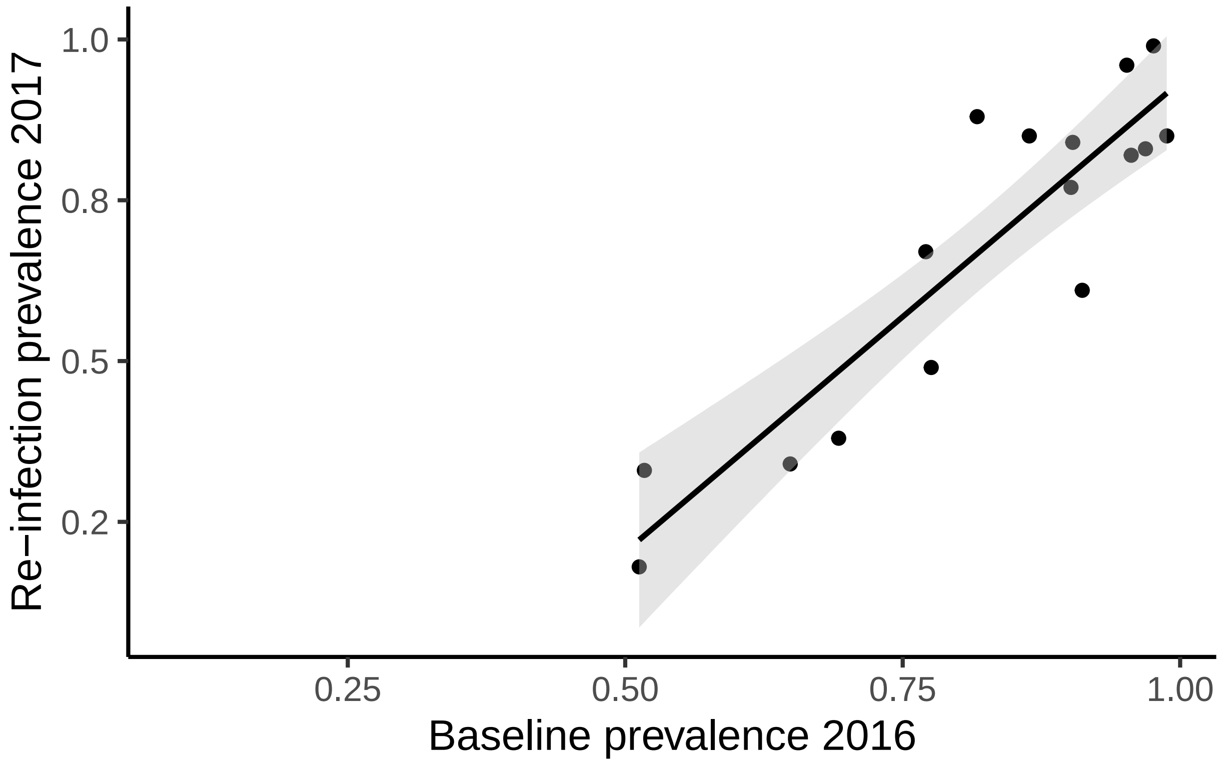


```
Checka<-lm(prevbase~prevre1, data=landp1)
summary(Checka)
```

```
##
## Call:
## lm(formula = prevbase ~ prevre1, data = landp1)
##
## Residuals:
##       Min        1Q    Median        3Q       Max
## -0.120361 -0.036711  0.000984  0.053243  0.125606
##
## Coefficients:
##             Estimate Std. Error t value Pr(>|t|)
## (Intercept)  0.44439    0.05055   8.791 4.50e-07 ***
## prevre1      0.56028    0.07043   7.955 1.46e-06 ***
## ---
## Signif. codes:  0 '***' 0.001 '**' 0.01 '*' 0.05 '.' 0.1 ' ' 1
##
## Residual standard error: 0.06925 on 14 degrees of freedom
##   (7 observations deleted due to missingness)
## Multiple R-squared:  0.8189, Adjusted R-squared:  0.8059
## F-statistic: 63.29 on 1 and 14 DF,  p-value: 1.462e-06
```

```
### PLOT REINFECTION 2 to BASELINE:###
```

```r
ggplot(landp1, aes(x=prevbase, y=prevre2)) + geom_point(size=2)+
  geom_smooth(method=lm,
              color="black", fill="gray")+
  labs(title="",
       x="Baseline prevalence 2016", y = "Re-infection prevalence 2018")+
  theme_classic(base_size = 16)  + scale_y_continuous(
    labels = scales::number_format(accuracy = 0.1,
                                   decimal.mark = '.'))
```

```
## `geom_smooth()` using formula 'y ~ x'

## Warning: Removed 7 rows containing non-finite values (stat_smooth).
## Removed 7 rows containing missing values (geom_point).
```

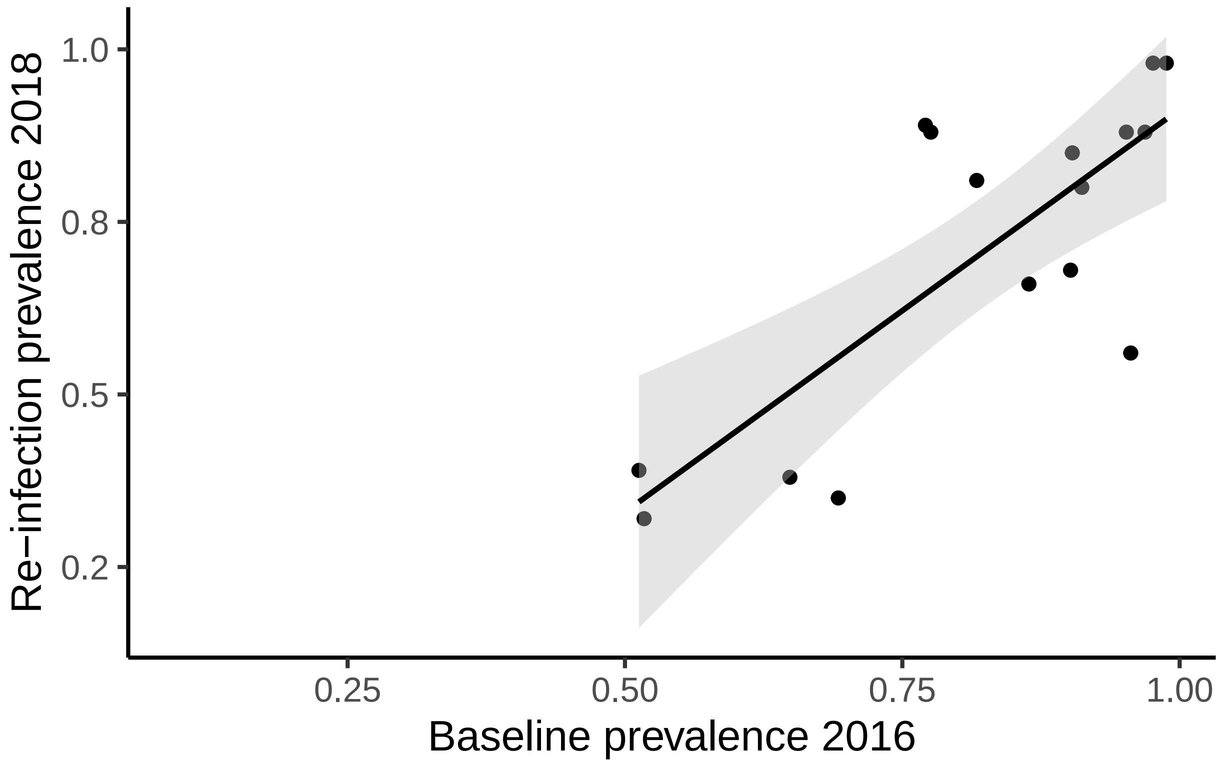


```r
Checkb<-lm(prevbase~prevre2, data=landp1)
summary(Checkb)
```

```
##
## Call:
## lm(formula = prevbase ~ prevre2, data = landp1)
##
## Residuals:
##      Min       1Q   Median       3Q      Max
## -0.14850 -0.07057  0.01524  0.05536  0.21035
```

```
##
## Coefficients:
##             Estimate Std. Error t value Pr(>|t|)
## (Intercept)  0.45060    0.08261   5.455 8.48e-05 ***
## prevre2      0.52667    0.11147   4.725 0.000325 ***
## ---
## Signif. codes:  0 '***' 0.001 '**' 0.01 '*' 0.05 '.' 0.1 ' ' 1
##
## Residual standard error: 0.101 on 14 degrees of freedom
##   (7 observations deleted due to missingness)
## Multiple R-squared:  0.6146, Adjusted R-squared:  0.5871
## F-statistic: 22.33 on 1 and 14 DF,  p-value: 0.0003255
```

```
###########################################################
##########################HUMAN INFECTION #################
###########################################################


#Regression of human infection using all 23 villages:
landp1$baseprev=landp1$prevalence_baseline
landp1$accessArea=landp1$Sum.of.area_FM3_17


#NEGATIVE BINOMIAL REGRESSIONS OF VILLAGE ATTRIBUTES
landp1$negkids_baseline=landp1$kids_baseline-landp1$infectedkids_baseline
landp1$negkids_baselineM=landp1$kids_baseline-landp1$infectedkids_baseline_mansoni

#Preliminary examination of which scales best:
m0.5km=glm(cbind(infectedkids_baseline,negkids_baseline)~ sqkmtotalcrop0.5km+ sqkmwater0.5km+population
m1km=glm(cbind(infectedkids_baseline,negkids_baseline)~ sqkmtotalcrop1km+ sqkmwater1km+population, na.a
m2km=glm(cbind(infectedkids_baseline,negkids_baseline)~ sqkmtotalcrop2km+sqkmwater2km+population, na.ac

models_1<-list(m0.5km,m1km,m2km)
Modnames_1 <- c('0.5km','1km','2km')
aictab(cand.set = models_1, modnames = Modnames_1, sort = TRUE)
```

```
##
## Model selection based on AICc:
##
##       K   AICc Delta_AICc AICcWt Cum.Wt      LL
## 0.5km 4 372.91       0.00      1      1 -181.35
## 2km   4 438.14      65.22      0      1 -213.96
## 1km   4 469.95      97.04      0      1 -229.86
```

```
# Figure 2 C-E and regression analysis
L1<-glm(cbind(infectedkids_baseline,negkids_baseline)~sqkmtotalcrop0.5km, data=landp1,family=binomial)
summary(L1)
```

```
##
## Call:
## glm(formula = cbind(infectedkids_baseline, negkids_baseline) ~
##     sqkmtotalcrop0.5km, family = binomial, data = landp1)
##
```

```
## Deviance Residuals:
##     Min       1Q   Median       3Q      Max
## -7.6113  -2.3268   0.1892   2.7746   5.7606
##
## Coefficients:
##                     Estimate Std. Error z value Pr(>|z|)
## (Intercept)          0.06731    0.15177   0.444    0.657
## sqkmtotalcrop0.5km  3.12673    0.37707   8.292   <2e-16 ***
## ---
## Signif. codes:  0 '***' 0.001 '**' 0.01 '*' 0.05 '.' 0.1 ' ' 1
##
## (Dispersion parameter for binomial family taken to be 1)
##
##     Null deviance: 390.94  on 22  degrees of freedom
## Residual deviance: 312.67  on 21  degrees of freedom
## AIC: 402.74
##
## Number of Fisher Scoring iterations: 5
```

```
Anova(L1)
```

```
## Analysis of Deviance Table (Type II tests)
##
## Response: cbind(infectedkids_baseline, negkids_baseline)
##                     LR Chisq Df Pr(>Chisq)
## sqkmtotalcrop0.5km   78.273  1  < 2.2e-16 ***
## ---
## Signif. codes:  0 '***' 0.001 '**' 0.01 '*' 0.05 '.' 0.1 ' ' 1
```

```
L1b<-glm(baseprev~sqkmtotalcrop0.5km, data=landp1,family=binomial,weights=kids_baseline)
summary(L1b)
```

```
##
## Call:
## glm(formula = baseprev ~ sqkmtotalcrop0.5km, family = binomial,
##     data = landp1, weights = kids_baseline)
##
## Deviance Residuals:
##     Min       1Q   Median       3Q      Max
## -7.6113  -2.3268   0.1892   2.7746   5.7606
##
## Coefficients:
##                     Estimate Std. Error z value Pr(>|z|)
## (Intercept)          0.06731    0.15177   0.444    0.657
## sqkmtotalcrop0.5km  3.12673    0.37707   8.292   <2e-16 ***
## ---
## Signif. codes:  0 '***' 0.001 '**' 0.01 '*' 0.05 '.' 0.1 ' ' 1
##
## (Dispersion parameter for binomial family taken to be 1)
##
##     Null deviance: 390.94  on 22  degrees of freedom
## Residual deviance: 312.67  on 21  degrees of freedom
## AIC: 402.74
```

```
##
## Number of Fisher Scoring iterations: 5
```

```
add_ci(landp1, L1b, names = c("lcb", "ucb"))
```

```
##       villages waterway sqkmtotalcrop0.5km sqkmwater1km sqkmtotalcrop2km
## 1      Diokhor      lac           0.222936     0.948440         3.629160
## 2     Diokhoul      lac           0.788500     0.756350         2.207280
## 3         Foss      lac           0.407990     0.478890         2.970460
## 4     Gankette      lac           0.483480     0.103240         6.943366
## 5      Guidick      lac           0.474450     1.133790         5.071914
## 6      Lampsar    river           0.275712     0.464620         5.734602
## 7    MakaDiama    river           0.369406     1.235390         4.113670
## 8        Malla      lac           0.491930     0.871230         5.098390
## 9   Malla Tack      lac           0.552050     0.400590         5.322650
## 10    Mbakhana    river           0.167428     0.737780         5.567160
## 11       Mbane      lac           0.326500     0.000000         6.802490
## 12    Mbarigot    river           0.323032     0.490510         3.112438
## 13 MerinaGewel      lac           0.704450     0.000000         4.886382
## 14   Ndiawdoune    river           0.262928     0.709190         1.901738
## 15  NdiolMaure    river           0.557480     0.140310         5.853860
## 16        Syer      lac           0.763150     0.105024         4.875906
## 17        assy    river           0.295000     0.449110         4.722270
## 18    diaminar      lac           0.582590     0.445770         4.967710
## 19    minguene    river           0.491556     0.000000         0.205056
## 20      ndelle    river           0.320152     0.000000         0.365020
## 21   ndiakhaye      lac           0.419310     0.083660         3.801654
## 22     salguir    river           0.120500     0.095184         5.055170
## 23     thilla1    river           0.322340     0.652248         3.818372
##    sqkmwater2km sqkmtotalcrop1km sqkmwater0.5km Year.baseline kids_baseline
## 1      4.891430         0.893824       0.056200          2016           122
## 2      5.379250         2.088930       0.032890          2016            48
## 3      3.629500         0.478890       0.000800          2016            31
## 4      0.934820         0.103240       0.000000          2016            68
## 5      5.845900         1.133790       0.144990          2016            82
## 6      3.533780         0.209150       0.057054          2016           107
## 7      5.322240         0.709190       0.200770          2016            96
## 8      5.191700         0.464620       0.015360          2016            83
## 9      2.196150         1.235390       0.269990          2016           104
## 10     4.795540         1.290938       0.298140          2016           111
## 11     0.000000         1.579750       0.084260          2016           103
## 12     0.581034         1.351856       0.029946          2016           119
## 13     0.135702         1.657150       0.081500          2016           102
## 14     1.491240         1.988846       0.085850          2016            91
## 15     0.497988         0.842542       0.006762          2016            87
## 16     0.224136         1.961830       0.009800          2016           125
## 17     1.785180         1.385320       0.000000          2017            30
## 18     1.450450         1.684590       0.005330          2017            30
## 19     0.414610         1.386876       0.052068          2017            30
## 20     0.000000         1.327750       0.000000          2017            30
## 21     0.255246         1.170220       0.108730          2017            30
## 22     0.246150         0.193104       0.000000          2017            31
## 23     1.139672         0.365020       0.000000          2017            30
##    infectedkids_baseline_H infectedkids_baseline_mansoni baseline_CI
```

```
## 1                       110                             17          17
## 2                        34                              7           4
## 3                        28                              1           1
## 4                        57                             63          55
## 5                        67                              0           0
## 6                        81                             42          40
## 7                        85                             31          23
## 8                        82                             68          68
## 9                        99                              9           9
## 10                       57                             42          27
## 11                       88                             18          17
## 12                       52                             30          21
## 13                       87                             76          70
## 14                       57                             32          26
## 15                       37                             12           4
## 16                      121                             28          27
## 17                       20                              2           2
## 18                       27                              1           1
## 19                       21                              1           1
## 20                       15                              0           0
## 21                       28                              0           0
## 22                        3                              0           0
## 23                        7                              0           0
##    infectedkids_baseline population rural prevalence_baseline
## 1                    110       1500 23818          0.90163934
## 2                     37       1300  3369          0.77083333
## 3                     28        107  7901          0.90322581
## 4                     65        459  6473          0.95588235
## 5                     67        400  5623          0.81707317
## 6                     83       1623 24486          0.77570094
## 7                     93        609 12516          0.96875000
## 8                     82        648  6273          0.98795181
## 9                     99        833 10923          0.95192308
## 10                    72       1120 35465          0.64864865
## 11                    89       1852 66884          0.86407767
## 12                    61       1316 11575          0.51260504
## 13                    93        761 10419          0.91176471
## 14                    63       1542 32881          0.69230769
## 15                    45        771  5975          0.51724138
## 16                   122        866  9463          0.97600000
## 17                    20        165  1502          0.66666667
## 18                    27         91  4278          0.90000000
## 19                    21       1146 18862          0.70000000
## 20                    15       2200 27035          0.50000000
## 21                    28        517 13564          0.93333333
## 22                     3        237  2656          0.09677419
## 23                     7        521  7860          0.23333333
##    prevalence_baseline_H prevalence_baseline_M prevalence_reinfection_2017
## 1             0.90163934            0.13934426                          77
## 2             0.70833333            0.14583333                          67
## 3             0.90322581            0.03225807                          84
## 4             0.83823529            0.92647059                          82
## 5             0.81707317            0.00000000                          88
## 6             0.75700935            0.39252336                          49
```

```
## 7       0.88541667       0.32291667                          83
## 8       0.98795181       0.81927711                          85
## 9       0.95192308       0.08653846                          96
## 10      0.51351351       0.37837838                          34
## 11      0.85436893       0.17475728                          85
## 12      0.43697479       0.25210084                          18
## 13      0.85294118       0.74509804                          61
## 14      0.62637363       0.35164835                          38
## 15      0.42528736       0.13793103                          33
## 16      0.96800000       0.22400000                          99
## 17      0.66666667       0.06666667                          NA
## 18      0.90000000       0.03333333                          NA
## 19      0.70000000       0.03333333                          NA
## 20      0.50000000       0.00000000                          NA
## 21      0.93333333       0.00000000                          NA
## 22      0.09677419       0.00000000                          NA
## 23      0.23333333       0.00000000                          NA
##    prevalence_reinfection2018 kids.tested.reinfection.Sh.2017
## 1                          68                              82
## 2                          89                              39
## 3                          85                              25
## 4                          56                              57
## 5                          81                              69
## 6                          88                             102
## 7                          88                              91
## 8                          98                              76
## 9                          88                             101
## 10                         38                             103
## 11                         66                             100
## 12                         39                             106
## 13                         80                              90
## 14                         35                              82
## 15                         32                              84
## 16                         98                             109
## 17                         NA                              NA
## 18                         NA                              NA
## 19                         NA                              NA
## 20                         NA                              NA
## 21                         NA                              NA
## 22                         NA                              NA
## 23                         NA                              NA
##    kids.reinfected.Sm.2017 kids.reinfected.Sh.2017 kids.tested.Sh.2018
## 1                       17                      63                 111
## 2                        7                      26                  38
## 3                        1                      21                  26
## 4                       63                      47                  55
## 5                        0                      61                  77
## 6                       42                      47                  99
## 7                       31                      64                  90
## 8                       68                      53                  57
## 9                        9                      97                  98
## 10                      42                      56                  94
## 11                      18                      85                 100
## 12                      30                      17                 104
```

```
## 13                     76                      55                    87
## 14                     32                      31                    79
## 15                     12                      23                    73
## 16                     28                     108                   105
## 17                     NA                      NA                    NA
## 18                     NA                      NA                    NA
## 19                     NA                      NA                    NA
## 20                     NA                      NA                    NA
## 21                     NA                      NA                    NA
## 22                     NA                      NA                    NA
## 23                     NA                      NA                    NA
##    infected.kids.Sh.2018 Sum.of.area_FM3_17 AccessArea_FM4_17
## 1                     76           3369.060              4085
## 2                     34           5496.101              3976
## 3                     22           3031.277              3225
## 4                     31           1677.355              1750
## 5                     62           2536.556              3433
## 6                     87           3081.368              1143
## 7                     79           1608.964              1633
## 8                     56            519.440               488
## 9                     86           6416.058              4774
## 10                    36           1498.642              1099
## 11                    66           2090.476              2215
## 12                    41            848.107               817
## 13                    70           2820.774              2675
## 14                    28           1386.967              1510
## 15                    23           2691.446              1300
## 16                   103           6807.490              4722
## 17                    NA           1921.000              1921
## 18                    NA           4920.000              4920
## 19                    NA           3775.000              3775
## 20                    NA           1905.000              1905
## 21                    NA           2135.000              2135
## 22                    NA           2464.000              2464
## 23                    NA           1565.000              1565
##    EmergentArea_FM4_17 FloatingArea_FM4_17 Mud_Area_FM4_17 avgFloatFM1FM4
## 1                 2645               160.9         1279.10        1255.49
## 2                  545              1077.0         2354.00        2584.98
## 3                  591               342.0         2292.00         520.99
## 4                 1351                46.0          353.00         623.63
## 5                  722               678.8         2032.20         944.26
## 6                  645               136.0          362.00        1553.70
## 7                  429               189.0         1015.00         583.36
## 8                  158                22.0          308.00          81.46
## 9                 1080               975.9         2718.10        2481.98
## 10                 122                90.0          887.00         351.00
## 11                 969               257.0          989.00         882.07
## 12                 209                29.2          578.80         168.54
## 13                 695                25.2         1954.80         278.57
## 14                 722               157.0          631.00         692.73
## 15                 697                58.0         1050.38        1050.38
## 16                 834              1355.0         3347.20        3347.20
## 17                 213                73.0         1635.00             NA
## 18                1174               506.0         3240.00             NA
```

```
## 19             1307            1200.0         1268.00             NA
## 20              554             558.0          793.00             NA
## 21             1206             308.0          621.00             NA
## 22             1192             100.0         1172.00             NA
## 23              681             131.0          753.00             NA
##    DateofAgricovariates average.field.area number.of.referenced.fields
## 1                  2015               1.77                          90
## 2                  2015               2.47                          47
## 3                  2015               1.50                          42
## 4                  2015               1.47                          60
## 5                  2015               1.60                          51
## 6                  2015               0.88                          68
## 7                  2015               3.93                          49
## 8                  2015               0.74                          69
## 9                  2015               1.79                          83
## 10                 2015               0.82                          63
## 11                 2015               1.34                         122
## 12                 2015               0.55                          44
## 13                 2015               1.53                          46
## 14                 2015               0.83                          36
## 15                 2015               0.85                          58
## 16                 2015               3.26                          51
## 17                   NA                 NA                          NA
## 18                   NA                 NA                          NA
## 19                   NA                 NA                          NA
## 20                   NA                 NA                          NA
## 21                   NA                 NA                          NA
## 22                   NA                 NA                          NA
## 23                   NA                 NA                          NA
##    ha.total.of.referenced.fields fallow wmelon rice groundnut bean onion
## 1                         159.60     12      9    0        17    1     7
## 2                         116.30     34     29    0        17   21     1
## 3                          63.00      9      9    0        15    8     2
## 4                          88.00      2     27    0        23   18    15
## 5                          81.55      5     30    0        27    7     4
## 6                          59.79      2      0   40         1    0     5
## 7                         192.80     24      0  141         0    0     8
## 8                          50.76      2      4    2         1    1     9
## 9                         148.35     20     31    0        39   45     2
## 10                         51.90     23      1    7         1    1     0
## 11                        163.15     23     33    3        26    8    51
## 12                         24.15      0      4    3         1    7     4
## 13                         70.25      7      8    0        17   28     0
## 14                         29.90      2      0    2         1    2     7
## 15                         49.19      6      2   33         1    0     3
## 16                        166.05     72     58    0        14   10     3
## 17                            NA     NA     NA   NA        NA   NA    NA
## 18                            NA     NA     NA   NA        NA   NA    NA
## 19                            NA     NA     NA   NA        NA   NA    NA
## 20                            NA     NA     NA   NA        NA   NA    NA
## 21                            NA     NA     NA   NA        NA   NA    NA
## 22                            NA     NA     NA   NA        NA   NA    NA
## 23                            NA     NA     NA   NA        NA   NA    NA
##    sweetpotatos tomato othercrop nofertilizer  Urea  NPK DAP Manure Other
```

```
## 1            65      2         47         16.5 124.6  8.5 4.5    1.0   4.5
## 2            NA      1         14         59.3  32.0 17.0 3.0    5.0   0.0
## 3            12      1          9         24.8  24.6  4.0 1.6    8.0   0.0
## 4             0      0          4         61.5   5.8 17.8 0.0    0.5   2.5
## 5             0      0          9         27.1  31.0 14.0 7.3    0.0   2.3
## 6             0      7          5          4.0  33.1 10.9 9.0    0.0   2.8
## 7             0      1         19         38.8 134.0 18.0 0.0    2.0   0.0
## 8             5     15         13          4.0  24.6 15.2 2.0    1.5   3.5
## 9             9      0          2         61.3  61.5 17.3 4.3    0.0   4.0
## 10            0      2         18         36.9   9.0  1.8 0.0    4.3   0.0
## 11            7      2         10         49.8  76.0 16.9 5.5    7.5   7.5
## 12            0      0          6          7.0   3.8  4.0 0.5    8.9   0.0
## 13            0      0         11         52.5   4.3  5.0 5.0    3.5   0.0
## 14            0      1         17         14.5  14.2  1.3 0.0    0.0   0.0
## 15            0      0          4          6.3  32.1  7.1 2.7    0.0   1.0
## 16            1      2          1        128.5  28.5  5.8 1.5    1.0   0.8
## 17           NA     NA         NA           NA    NA   NA  NA     NA    NA
## 18           NA     NA         NA           NA    NA   NA  NA     NA    NA
## 19           NA     NA         NA           NA    NA   NA  NA     NA    NA
## 20           NA     NA         NA           NA    NA   NA  NA     NA    NA
## 21           NA     NA         NA           NA    NA   NA  NA     NA    NA
## 22           NA     NA         NA           NA    NA   NA  NA     NA    NA
## 23           NA     NA         NA           NA    NA   NA  NA     NA    NA
##    TotalFert no.insecticide spithoate.dimethoate DECIS15.deltamethrine Bomec
## 1      143.1           74.0                 80.1                   0.0   0.0
## 2       57.0           50.8                 53.5                   7.0   0.0
## 3       38.2           23.6                 31.5                   3.7   4.2
## 4       26.6           49.5                 31.0                   2.5   0.0
## 5       54.6           23.5                 56.1                   0.5   1.0
## 6       55.8           37.4                  8.9                   7.3   0.0
## 7      154.0          118.8                  5.5                  22.0   0.0
## 8       46.8           21.3                 11.1                   1.5   6.7
## 9       87.1           48.1                 73.0                   2.0  15.0
## 10      15.1           42.6                  3.3                   4.6   0.0
## 11     113.4           62.4                 79.4                   5.3  11.5
## 12      17.2           14.8                  2.8                   3.7   1.0
## 13      17.8           56.0                  8.8                   5.5   0.0
## 14      15.5           13.3                  6.2                   7.5   0.0
## 15      42.9           24.2                  2.8                   9.6   1.5
## 16      37.6           90.5                 62.3                   0.3   1.0
## 17        NA             NA                   NA                    NA    NA
## 18        NA             NA                   NA                    NA    NA
## 19        NA             NA                   NA                    NA    NA
## 20        NA             NA                   NA                    NA    NA
## 21        NA             NA                   NA                    NA    NA
## 22        NA             NA                   NA                    NA    NA
## 23        NA             NA                   NA                    NA    NA
##    Viden Metaforce Vertimec.abamectine Arsenal Optimal Furadan Laprine
## 1    0.0       0.0                 0.0     1.0     3.0     1.5       0
## 2    0.0       0.0                 0.0     0.0     0.0     0.0       0
## 3    0.0       0.0                 0.0     0.0     0.0     0.0       0
## 4    0.0       3.0                 2.0     0.0     0.0     0.0       0
## 5    0.0       0.0                 0.0     0.0     0.0     0.0       0
## 6    1.5       0.0                 2.0     0.0     0.0     1.6       0
```

```
## 7   39.0       0.0                0.0     6.0     0.0     1.0       0
## 8    0.0       0.0                0.0     3.0     1.2     0.0       6
## 9    0.0       0.3                6.0     0.0     4.0     0.0       0
## 10   0.0       0.0                0.5     0.0     0.0     0.0       0
## 11   0.0       1.4                0.0     0.0     1.5     0.0       0
## 12   0.0       0.0                0.5     0.0     1.0     0.0       0
## 13   0.0       0.0                0.0     0.0     0.0     0.0       0
## 14   0.0       0.0                1.5     0.0     0.0     0.0       0
## 15   0.0       0.0                2.3     0.7     0.0     2.5       0
## 16   0.0      12.0                0.0     0.0     0.0     0.0       0
## 17    NA        NA                 NA      NA      NA      NA      NA
## 18    NA        NA                 NA      NA      NA      NA      NA
## 19    NA        NA                 NA      NA      NA      NA      NA
## 20    NA        NA                 NA      NA      NA      NA      NA
## 21    NA        NA                 NA      NA      NA      NA      NA
## 22    NA        NA                 NA      NA      NA      NA      NA
## 23    NA        NA                 NA      NA      NA      NA      NA
##    Other.insecticide Total.insecticide No.herbicide Spiriz.360.EC.Propanil
## 1                0.0              85.6        150.1                    0.0
## 2                5.0              65.5        116.3                    0.0
## 3                0.0              39.4         59.0                    1.0
## 4                0.0              38.5         88.0                    0.0
## 5                0.5              58.1         81.6                    0.0
## 6                1.3              22.6         30.5                   15.7
## 7                0.5              74.0         51.3                   83.0
## 8                0.0              29.5         44.0                    0.0
## 9                0.0             100.3        120.1                    0.5
## 10               1.0               9.4         45.4                    0.5
## 11               1.7             100.8        117.1                   11.0
## 12               0.5               9.5         17.8                    2.0
## 13               0.0              14.3         66.3                    0.0
## 14               1.5              16.7         21.7                    3.0
## 15               5.7              25.1         10.2                   14.5
## 16               0.0              75.6        161.1                    0.0
## 17                NA                NA           NA                     NA
## 18                NA                NA           NA                     NA
## 19                NA                NA           NA                     NA
## 20                NA                NA           NA                     NA
## 21                NA                NA           NA                     NA
## 22                NA                NA           NA                     NA
## 23                NA                NA           NA                     NA
##    Londax.Bensulfuron.methyl Calliherbe.2.4.D Other.herbicides Total.Herbicide
## 1                        2.0              1.0              7.5            10.5
## 2                        0.0              0.0              0.0             0.0
## 3                        3.0              0.0              0.0             4.0
## 4                        0.0              0.0              0.0             0.0
## 5                        0.0              0.0              0.0             0.0
## 6                        8.2              1.0              5.4            30.3
## 7                       52.5              0.0              6.0           141.5
## 8                        6.0              0.5              0.8             7.3
## 9                        0.3              0.0             27.5            28.3
## 10                       4.0              0.0              2.0             6.5
## 11                       3.6              4.0             31.5            50.1
## 12                       2.7              0.0              1.8             6.5
```

```
## 13                     1.0             3.0             3.0            7.0
## 14                     1.0             0.0             4.3            8.3
## 15                    16.0             2.3             8.5           41.3
## 16                     3.0             0.0             2.0            5.0
## 17                      NA              NA              NA             NA
## 18                      NA              NA              NA             NA
## 19                      NA              NA              NA             NA
## 20                      NA              NA              NA             NA
## 21                      NA              NA              NA             NA
## 22                      NA              NA              NA             NA
## 23                      NA              NA              NA             NA
##      prevbase prevre1 prevre2   baseprev accessArea negkids_baseline
## 1  0.90163934    0.77    0.68 0.90163934   3369.060               12
## 2  0.77083333    0.67    0.89 0.77083333   5496.101               11
## 3  0.90322581    0.84    0.85 0.90322581   3031.277                3
## 4  0.95588235    0.82    0.56 0.95588235   1677.355                3
## 5  0.81707317    0.88    0.81 0.81707317   2536.556               15
## 6  0.77570094    0.49    0.88 0.77570094   3081.368               24
## 7  0.96875000    0.83    0.88 0.96875000   1608.964                3
## 8  0.98795181    0.85    0.98 0.98795181    519.440                1
## 9  0.95192308    0.96    0.88 0.95192308   6416.058                5
## 10 0.64864865    0.34    0.38 0.64864865   1498.642               39
## 11 0.86407767    0.85    0.66 0.86407767   2090.476               14
## 12 0.51260504    0.18    0.39 0.51260504    848.107               58
## 13 0.91176471    0.61    0.80 0.91176471   2820.774                9
## 14 0.69230769    0.38    0.35 0.69230769   1386.967               28
## 15 0.51724138    0.33    0.32 0.51724138   2691.446               42
## 16 0.97600000    0.99    0.98 0.97600000   6807.490                3
## 17 0.66666667      NA      NA 0.66666667   1921.000               10
## 18 0.90000000      NA      NA 0.90000000   4920.000                3
## 19 0.70000000      NA      NA 0.70000000   3775.000                9
## 20 0.50000000      NA      NA 0.50000000   1905.000               15
## 21 0.93333333      NA      NA 0.93333333   2135.000                2
## 22 0.09677419      NA      NA 0.09677419   2464.000               28
## 23 0.23333333      NA      NA 0.23333333   1565.000               23
##    negkids_baselineM      pred       lcb       ucb
## 1                105 0.6823023 0.6465056 0.7160660
## 2                 41 0.9264051 0.9002933 0.9460881
## 3                 30 0.7929805 0.7721354 0.8123820
## 4                  5 0.8290660 0.8073441 0.8487974
## 5                 82 0.8250275 0.8035094 0.8446446
## 6                 65 0.7169512 0.6882244 0.7440156
## 7                 65 0.7724758 0.7508655 0.7927290
## 8                 15 0.8327777 0.8108524 0.8526235
## 9                 95 0.8573463 0.8338379 0.8780153
## 10                69 0.6435519 0.5987320 0.6859911
## 11                85 0.7480405 0.7241300 0.7705352
## 12                89 0.7459912 0.7218230 0.7687325
## 13                26 0.9063562 0.8802470 0.9272435
## 14                59 0.7087697 0.6785025 0.7372908
## 15                75 0.8594102 0.8357611 0.8801423
## 16                97 0.9208151 0.8946027 0.9409388
## 17                28 0.7290280 0.7023864 0.7541196
## 18                29 0.8686314 0.8443629 0.8896104
```

```
## 19                 29 0.8326148 0.8106987 0.8524554
## 20                 30 0.7442811 0.7198905 0.7672346
## 21                 30 0.7987307 0.7779159 0.8180510
## 22                 31 0.6092315 0.5560877 0.6599041
## 23                 30 0.7455810 0.7213600 0.7683727
```

```
#this gives us the predicted values of the model , along with the confidence bands

(est <- cbind(Estimate = coef(L1b), confint(L1b)))
```

```
## Waiting for profiling to be done...
```

```
##                      Estimate      2.5 %    97.5 %
## (Intercept)         0.0673125 -0.2307795 0.3644918
## sqkmtotalcrop0.5km 3.1267262  2.3998260 3.8789738
```

```
#To get incidence rate  (we exponentiate the coefficients.. and confidence intervals!):
exp(est)
```

```
##                     Estimate      2.5 %    97.5 %
## (Intercept)          1.06963  0.7939145  1.439782
## sqkmtotalcrop0.5km 22.79922 11.0212588 48.374547
```

```
#R2 for model
with(summary(L1b), 1 - deviance/null.deviance)
```

```
## [1] 0.2002148
```

```
r2<-"0.200"
p<-"0.001"
lb1 <- paste("'p <'~",p,"~R^2==~", r2)

PL1<-effect_plot(L1b, pred = sqkmtotalcrop0.5km, interval = TRUE, plot.points = TRUE, data=landp1)+labs
       x="Total crop cover in 0.5km", y = "Baseline human prevalence")+
  theme_classic(base_size = 16) + scale_y_continuous(limits = c(0, 1),
    labels = scales::number_format(accuracy = 0.1,
                                   decimal.mark = '.'))+ theme(legend.position="right")+ annotate("text

PL1
```

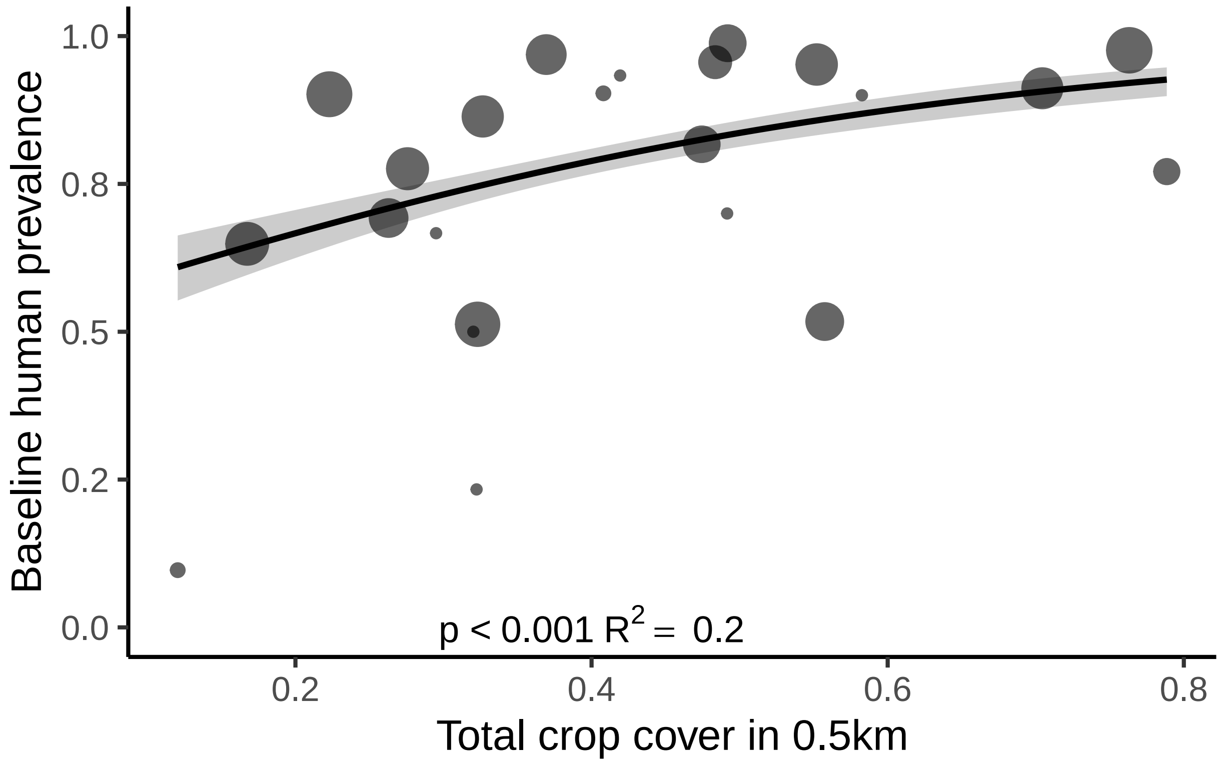


```
L2<-lm(TotalFert~ha.total.of.referenced.fields, data=landp1)
summary(L2)
```

```
##
## Call:
## lm(formula = TotalFert ~ ha.total.of.referenced.fields, data = landp1)
##
## Residuals:
##     Min      1Q  Median      3Q     Max
## -66.708 -14.667   3.365  15.667  43.007
##
## Coefficients:
##                               Estimate Std. Error t value Pr(>|t|)
## (Intercept)                    -4.1896    13.8405  -0.303 0.766568
## ha.total.of.referenced.fields   0.6534     0.1275   5.123 0.000155 ***
## ---
## Signif. codes:  0 '***' 0.001 '**' 0.01 '*' 0.05 '.' 0.1 ' ' 1
##
## Residual standard error: 27.06 on 14 degrees of freedom
##   (7 observations deleted due to missingness)
## Multiple R-squared:  0.6521, Adjusted R-squared:  0.6273
## F-statistic: 26.24 on 1 and 14 DF,  p-value: 0.000155
```

```
library(pscl)
```

```
## Warning: package 'pscl' was built under R version 4.1.2

## Classes and Methods for R developed in the
## Political Science Computational Laboratory
## Department of Political Science
## Stanford University
## Simon Jackman
## hurdle and zeroinfl functions by Achim Zeileis
```

```
r2<-"0.627"
p<-"0.001"
lb1 <- paste("'p <'~",p,"~R^2==~", r2)

PL2<-effect_plot(L2, pred = ha.total.of.referenced.fields, interval = TRUE, plot.points = TRUE)+ labs(t
        x="Total field area (ha)", y = "Total fertilizer use (ha)")+
  theme_classic(base_size = 16) + annotate("text", x=110, y=-15, label=lb1, parse=TRUE, size = 5)

PL2
```

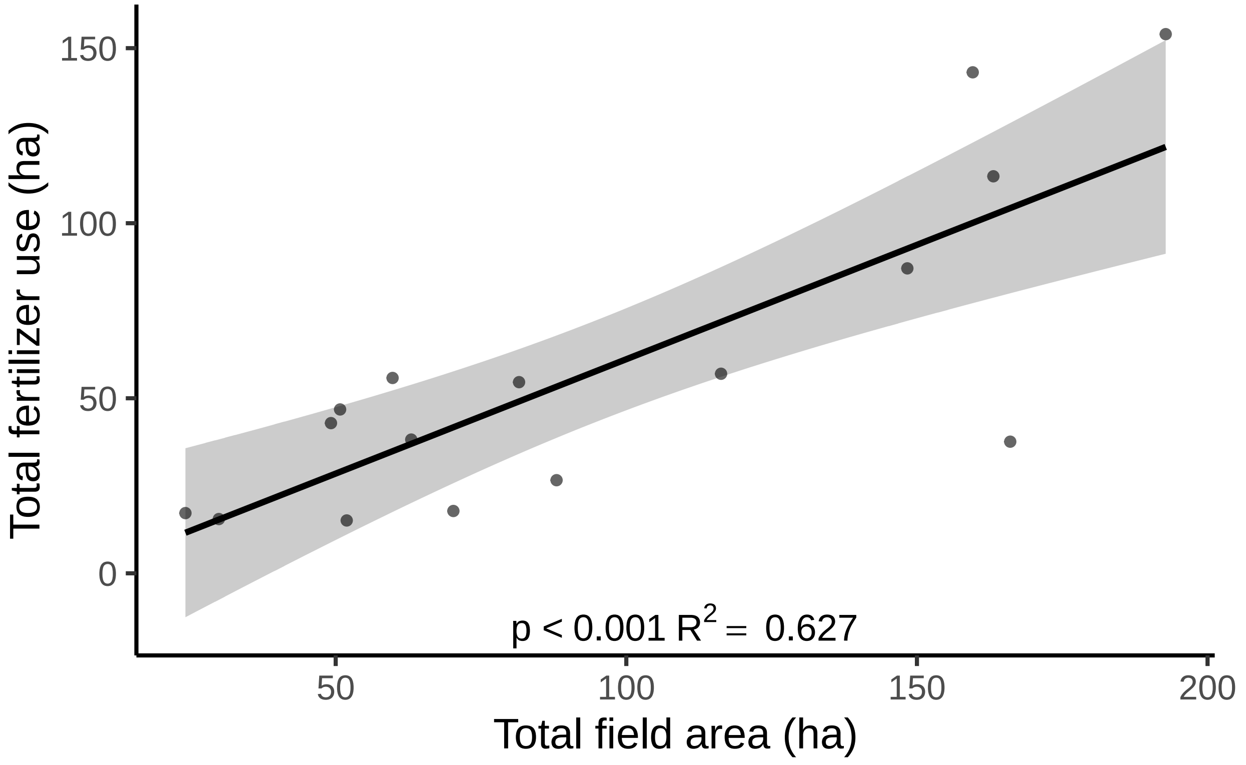


```
landp1$logtotalfert=log(landp1$TotalFert+1)
landp1$logavgFloat=log(landp1$avgFloatFM1FM4+1)
L3<-lm(logavgFloat~logtotalfert, data=landp1)
summary(L3)
```

```
##
```

```
## Call:
## lm(formula = logavgFloat ~ logtotalfert, data = landp1)
##
## Residuals:
##      Min       1Q   Median       3Q      Max
## -2.21098 -0.36177  0.00914  0.54035  1.61181
##
## Coefficients:
##              Estimate Std. Error t value Pr(>|t|)
## (Intercept)    4.4719     1.2729   3.513  0.00344 **
## logtotalfert   0.5563     0.3278   1.697  0.11175
## ---
## Signif. codes:  0 '***' 0.001 '**' 0.01 '*' 0.05 '.' 0.1 ' ' 1
##
## Residual standard error: 0.9442 on 14 degrees of freedom
##   (7 observations deleted due to missingness)
## Multiple R-squared:  0.1707, Adjusted R-squared:  0.1114
## F-statistic: 2.881 on 1 and 14 DF,  p-value: 0.1117
```

```
ols_plot_resid_qq(L3)
```

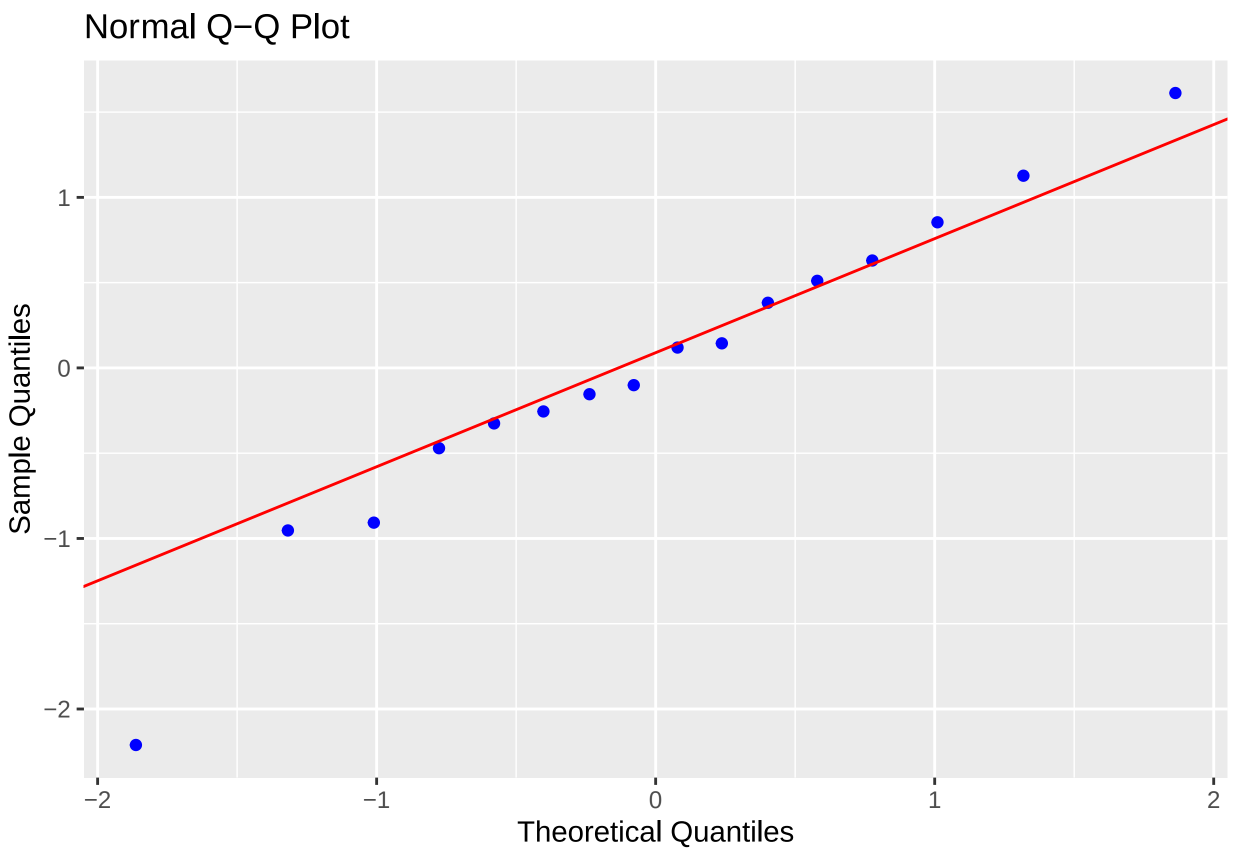


```
ols_plot_resid_fit(L3)# CHECK OUTLIER
```

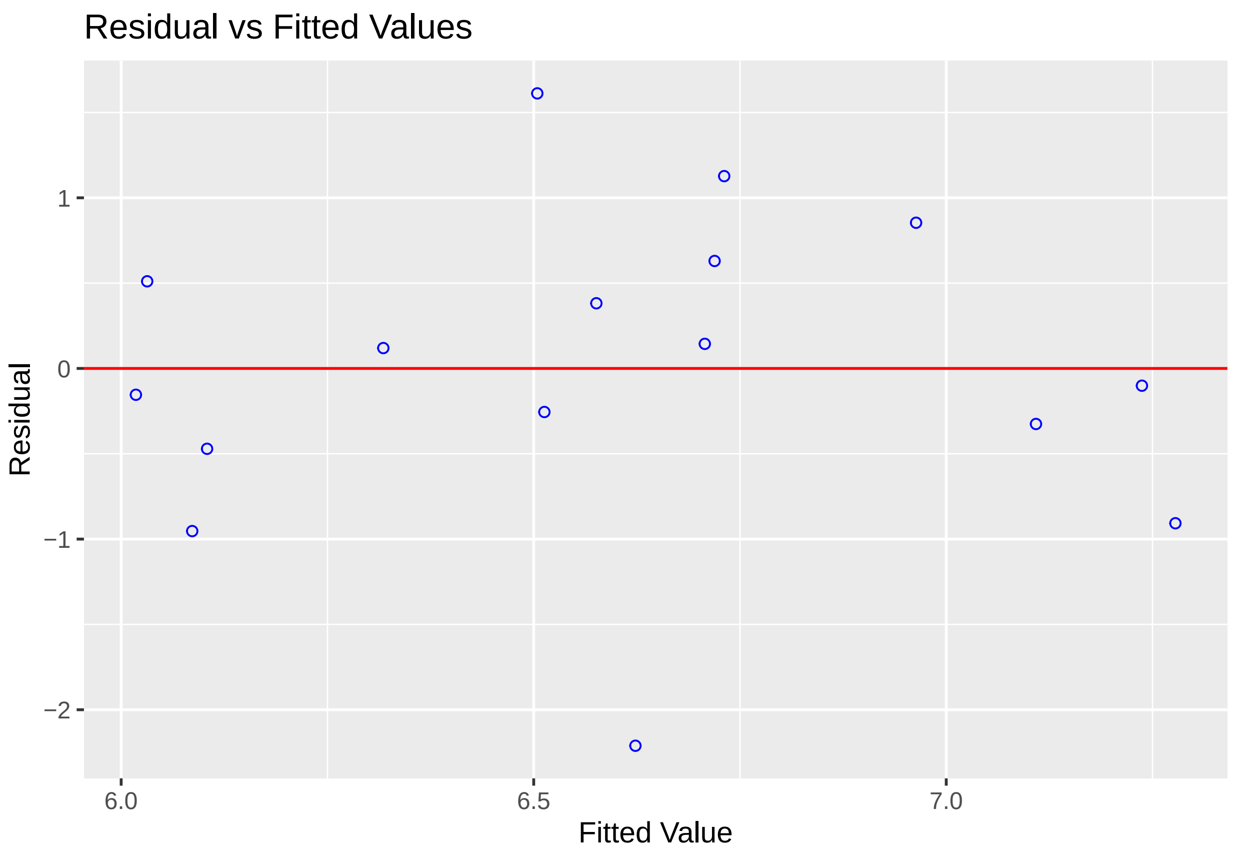


```
#Remove outlier (see q-q plot and residual over -2)
L3b<-lm(logavgFloat~logtotalfert, data=subset(landp1,landp1$logavgFloat>5))
summary(L3b)
```

```
##
## Call:
## lm(formula = logavgFloat ~ logtotalfert, data = subset(landp1,
##     landp1$logavgFloat > 5))
##
## Residuals:
##      Min       1Q   Median       3Q      Max
## -1.08719 -0.44342 -0.02082  0.42843  1.46672
##
## Coefficients:
##              Estimate Std. Error t value Pr(>|t|)
## (Intercept)    4.5640     1.0083   4.527 0.000569 ***
## logtotalfert   0.5708     0.2596   2.199 0.046564 *
## ---
## Signif. codes:  0 '***' 0.001 '**' 0.01 '*' 0.05 '.' 0.1 ' ' 1
##
## Residual standard error: 0.7476 on 13 degrees of freedom
## Multiple R-squared:  0.2712, Adjusted R-squared:  0.2151
## F-statistic: 4.837 on 1 and 13 DF,  p-value: 0.04656
```

```r
ols_plot_resid_qq(L3b)
```

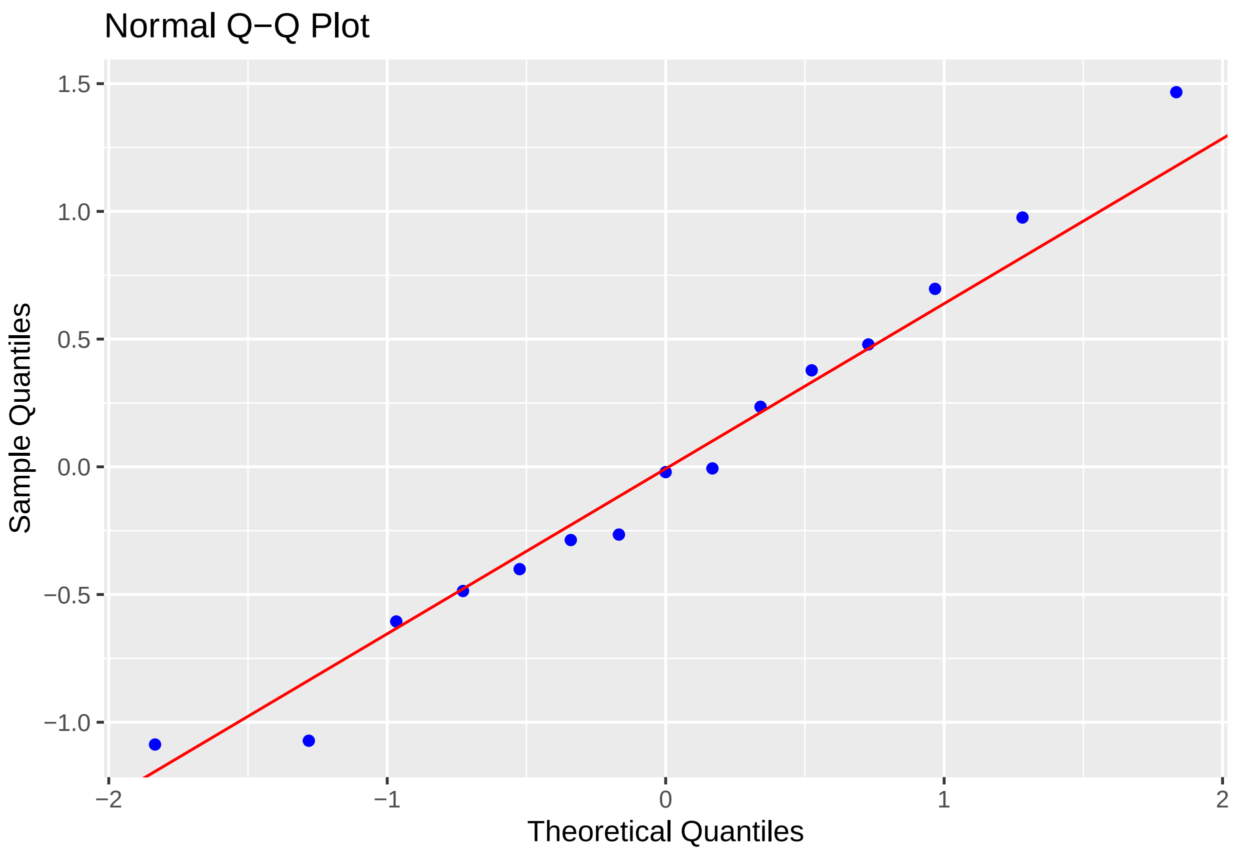


```r
ols_plot_resid_fit(L3b)
```

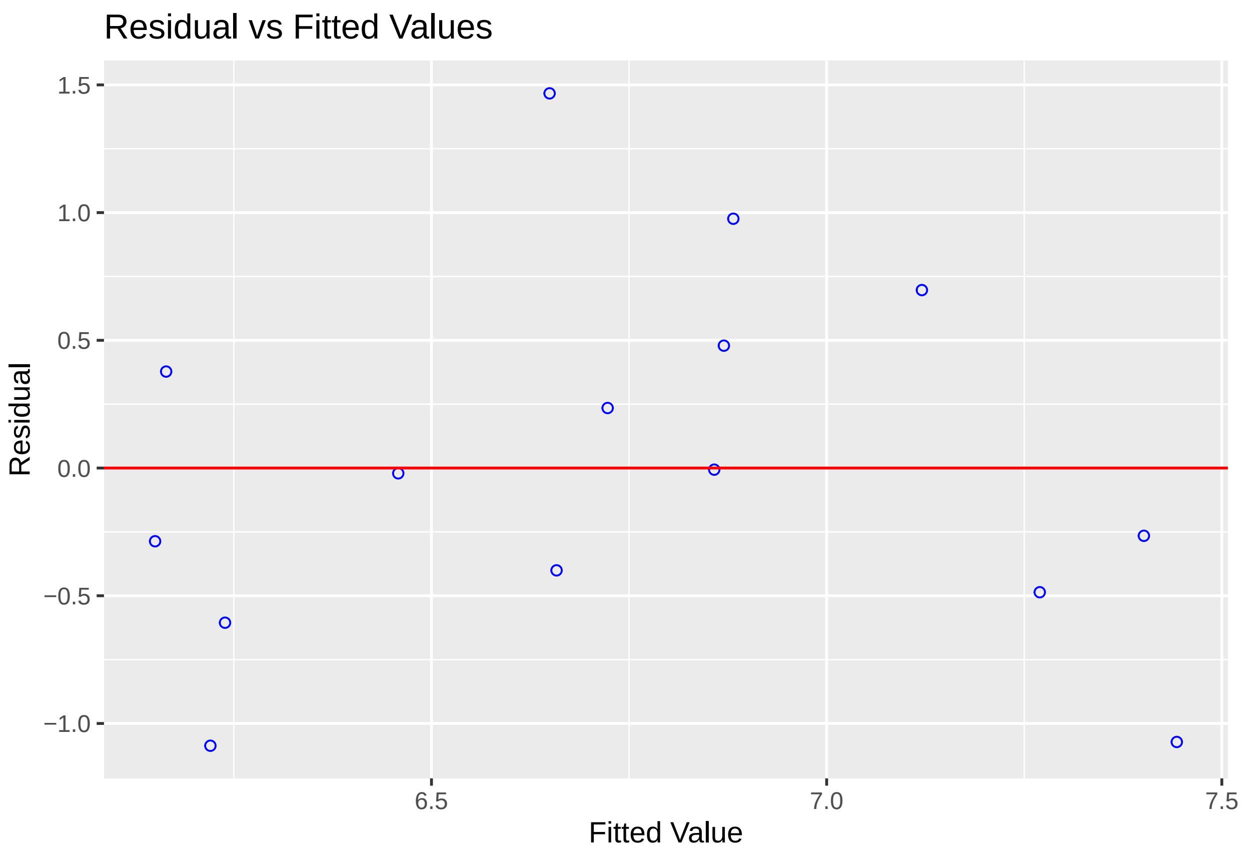


```
r2<-"0.215"
p<-"0.046"
lb1 <- paste("'p ='~",p, "~R^2==~", r2)

PL3<-effect_plot(L3b, pred = logtotalfert, interval = TRUE, plot.points = TRUE)+
  labs(title="",
       x="Ln-transformed total fertilizer", y = "Ln-mean floating vegetation area (m2)")+
  theme_classic(base_size = 16) + annotate("text", x=4, y=5, label=lb1, parse=TRUE, size = 5)
PL3
```

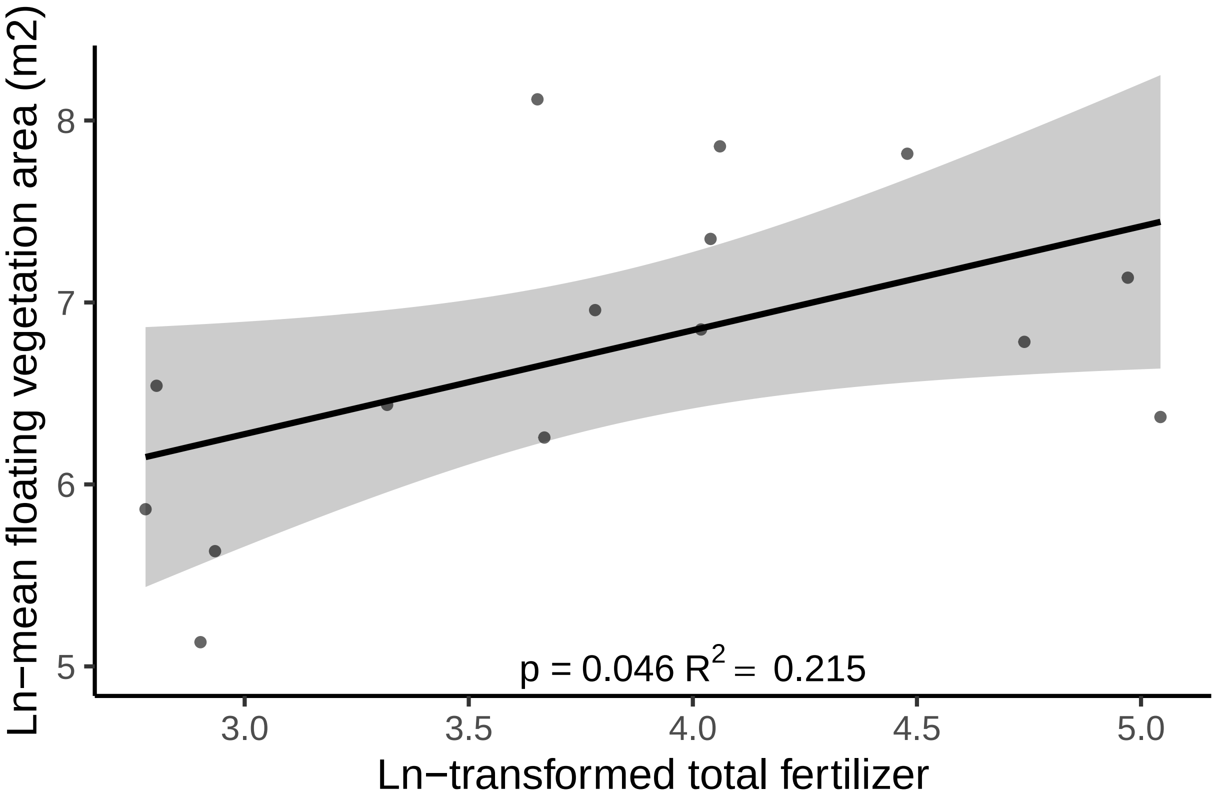


```
plot_grid(PL1, PL2, PL3, align="vh",axis="rlbt",nrow=1,ncol=3)
```

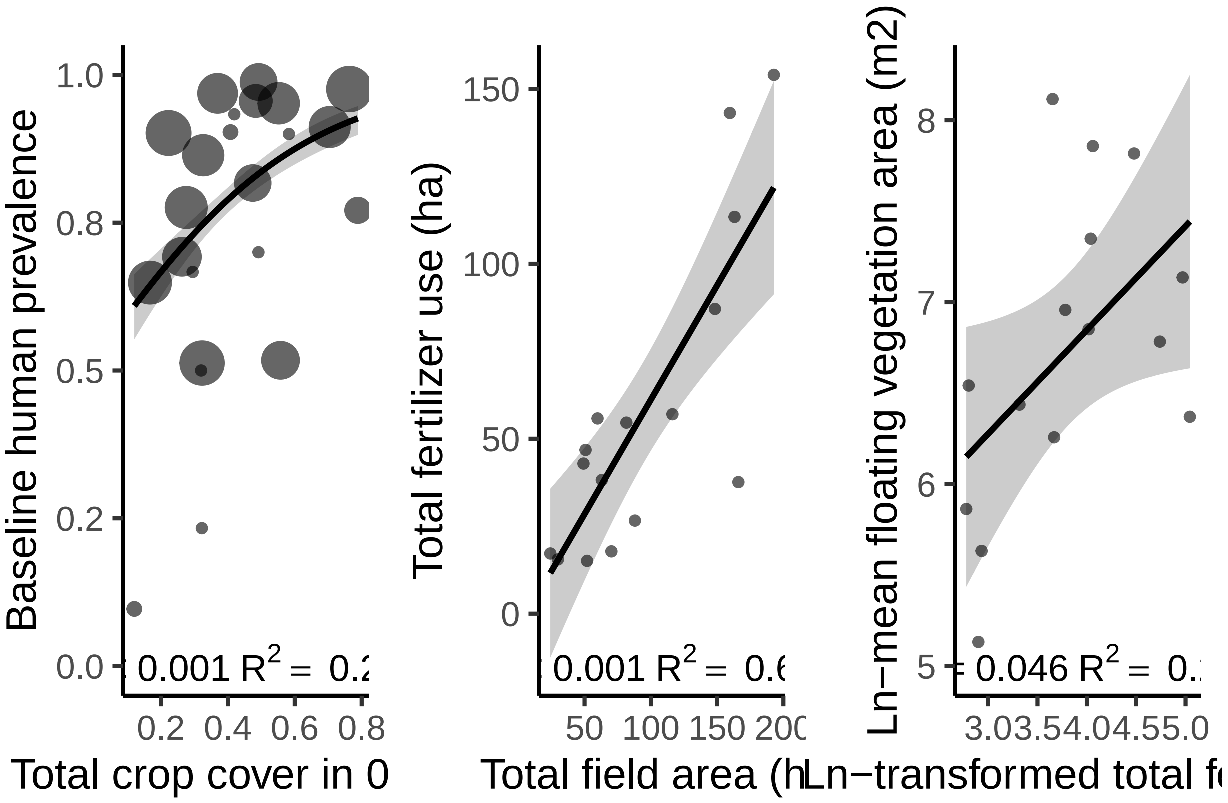


```
#0.5km is best when examinded alone

mm1=glm(cbind(infectedkids_baseline,negkids_baseline)~ sqkmtotalcrop0.5km+ sqkmwater0.5km+population, na
mm2=glm(cbind(infectedkids_baseline,negkids_baseline)~ sqkmtotalcrop0.5km+population, na.action=na.omit
mm3=glm(cbind(infectedkids_baseline,negkids_baseline)~ sqkmtotalcrop0.5km+sqkmwater0.5km, na.action=na.
mm4=glm(cbind(infectedkids_baseline,negkids_baseline)~ sqkmtotalcrop0.5km, na.action=na.omit,data = lan
mm5=glm(cbind(infectedkids_baseline,negkids_baseline)~ sqkmwater0.5km, na.action=na.omit,data = landp1,
mm6=glm(cbind(infectedkids_baseline,negkids_baseline)~ sqkmwater0.5km+population, na.action=na.omit,data
mm7=glm(cbind(infectedkids_baseline,negkids_baseline)~ sqkmwater0.5km+Sum.of.area_FM3_17, na.action=na.
mm8=glm(cbind(infectedkids_baseline,negkids_baseline)~ sqkmwater0.5km+Sum.of.area_FM3_17+population, na
mm9=glm(cbind(infectedkids_baseline,negkids_baseline)~ population , na.action=na.omit,data = landp1, fa
mm10=glm(cbind(infectedkids_baseline,negkids_baseline)~ population +Sum.of.area_FM3_17, na.action=na.om
mm11=glm(cbind(infectedkids_baseline,negkids_baseline)~ population +Sum.of.area_FM3_17+sqkmwater0.5km, n
mm12=glm(cbind(infectedkids_baseline,negkids_baseline)~ Sum.of.area_FM3_17, na.action=na.omit,data = la

mm13=glm(cbind(infectedkids_baseline,negkids_baseline)~ sqkmtotalcrop0.5km+Sum.of.area_FM3_17, na.actio
mm14=glm(cbind(infectedkids_baseline,negkids_baseline)~ sqkmtotalcrop0.5km+ sqkmwater0.5km+Sum.of.area_
mm15=glm(cbind(infectedkids_baseline,negkids_baseline)~ sqkmtotalcrop0.5km+ sqkmwater0.5km+population+S
mm16=glm(cbind(infectedkids_baseline,negkids_baseline)~ sqkmtotalcrop0.5km+population+Sum.of.area_FM3_1

mm1.1=glm(cbind(infectedkids_baseline,negkids_baseline)~ sqkmtotalcrop1km+ sqkmwater1km+population, na.
mm2.1=glm(cbind(infectedkids_baseline,negkids_baseline)~ sqkmtotalcrop1km+population, na.action=na.omit
mm3.1=glm(cbind(infectedkids_baseline,negkids_baseline)~ sqkmtotalcrop1km+sqkmwater1km, na.action=na.om
mm4.1=glm(cbind(infectedkids_baseline,negkids_baseline)~ sqkmtotalcrop1km, na.action=na.omit,data = lan
mm5.1=glm(cbind(infectedkids_baseline,negkids_baseline)~ sqkmwater1km, na.action=na.omit,data = landp1,
mm6.1=glm(cbind(infectedkids_baseline,negkids_baseline)~ sqkmwater1km+population, na.action=na.omit,dat
```

```
mm7.1=glm(cbind(infectedkids_baseline,negkids_baseline)~ sqkmwater1km+Sum.of.area_FM3_17, na.action=na.o
mm8.1=glm(cbind(infectedkids_baseline,negkids_baseline)~ sqkmwater1km+Sum.of.area_FM3_17+population, na

mm13.1=glm(cbind(infectedkids_baseline,negkids_baseline)~ sqkmtotalcrop1km+Sum.of.area_FM3_17, na.action
mm14.1=glm(cbind(infectedkids_baseline,negkids_baseline)~ sqkmtotalcrop1km+ sqkmwater1km+Sum.of.area_FM3
mm15.1=glm(cbind(infectedkids_baseline,negkids_baseline)~ sqkmtotalcrop1km+ sqkmwater1km+population+Sum
mm16.1=glm(cbind(infectedkids_baseline,negkids_baseline)~ sqkmtotalcrop1km+population+Sum.of.area_FM3_17

mm1.2=glm(cbind(infectedkids_baseline,negkids_baseline)~ sqkmtotalcrop2km+ sqkmwater2km+population, na.a
mm2.2=glm(cbind(infectedkids_baseline,negkids_baseline)~ sqkmtotalcrop2km+population, na.action=na.omit
mm3.2=glm(cbind(infectedkids_baseline,negkids_baseline)~ sqkmtotalcrop2km+sqkmwater2km, na.action=na.omi
mm4.2=glm(cbind(infectedkids_baseline,negkids_baseline)~ sqkmtotalcrop2km, na.action=na.omit,data = land
mm5.2=glm(cbind(infectedkids_baseline,negkids_baseline)~ sqkmwater2km, na.action=na.omit,data = landp1,
mm6.2=glm(cbind(infectedkids_baseline,negkids_baseline)~ sqkmwater2km+population, na.action=na.omit,data
mm7.2=glm(cbind(infectedkids_baseline,negkids_baseline)~ sqkmwater2km+Sum.of.area_FM3_17, na.action=na.o
mm8.2=glm(cbind(infectedkids_baseline,negkids_baseline)~ sqkmwater2km+Sum.of.area_FM3_17+population, na

mm13.2=glm(cbind(infectedkids_baseline,negkids_baseline)~ sqkmtotalcrop2km+Sum.of.area_FM3_17, na.action
mm14.2=glm(cbind(infectedkids_baseline,negkids_baseline)~ sqkmtotalcrop2km+ sqkmwater2km+Sum.of.area_FM3
mm15.2=glm(cbind(infectedkids_baseline,negkids_baseline)~ sqkmtotalcrop2km+ sqkmwater2km+population+Sum
mm16.2=glm(cbind(infectedkids_baseline,negkids_baseline)~ sqkmtotalcrop2km+population+Sum.of.area_FM3_17


models_big<-list(mm1,mm2,mm3,mm4,mm5,mm6,mm7,mm8,mm9,mm10,mm11,mm12,mm13,mm14,mm15,mm16,mm1.1,mm2.1,mm3
Modnames_big <- c('sqkmtotalcrop0.5km+ sqkmwater0.5km+population',
              'sqkmtotalcrop0.5km+population',
              'sqkmtotalcrop0.5km+sqkmwater0.5km',
              'sqkmtotalcrop0.5km',
              'sqkmwater0.5km',
              'sqkmwater0.5km+population',
              'sqkmwater0.5km+Sum.of.area_FM3_17',
              'sqkmwater0.5km+Sum.of.area_FM3_17+population',
              'population',
              'population +Sum.of.area_FM3_17',
              'population +Sum.of.area_FM3_17+sqkmwater0.5km',
              'Sum.of.area_FM3_17',
              'sqkmtotalcrop0.5km+Sum.of.area_FM3_17',
              'sqkmtotalcrop0.5km+ sqkmwater0.5km+Sum.of.area_FM3_17',
              'sqkmtotalcrop0.5km+ sqkmwater0.5km+population+Sum.of.area_FM3_17',
              'sqkmtotalcrop0.5km+population+Sum.of.area_FM3_17',
              'sqkmtotalcrop1km+ sqkmwater1km+population',
              'sqkmtotalcrop1km+population',
              'sqkmtotalcrop1km+sqkmwater1km',
              'sqkmtotalcrop1km',
              'sqkmwater1km',
              'sqkmwater1km+population',
              'sqkmwater1km+Sum.of.area_FM3_17',
              'sqkmwater1km+Sum.of.area_FM3_17+population',
              'sqkmtotalcrop1km+Sum.of.area_FM3_17',
              'sqkmtotalcrop1km+ sqkmwater1km+Sum.of.area_FM3_17',
              'sqkmtotalcrop1km+ sqkmwater1km+population+Sum.of.area_FM3_17',
              'sqkmtotalcrop1km+population+Sum.of.area_FM3_17',
              'sqkmtotalcrop2km+ sqkmwater2km+population',
```

```r
             'sqkmtotalcrop2km+population',
             'sqkmtotalcrop2km+sqkmwater2km',
             'sqkmtotalcrop2km',
             'sqkmwater2km',
             'sqkmwater2km+population',
             'sqkmwater2km+Sum.of.area_FM3_17',
             'sqkmwater2km+Sum.of.area_FM3_17+population',
             'sqkmtotalcrop2km+Sum.of.area_FM3_17',
             'sqkmtotalcrop2km+ sqkmwater2km+Sum.of.area_FM3_17',
             'sqkmtotalcrop2km+ sqkmwater2km+population+Sum.of.area_FM3_17',
             'sqkmtotalcrop2km+population+Sum.of.area_FM3_17')
aictab(cand.set = models_big, modnames = Modnames_big, sort = TRUE)
```

```
## Warning in aictab.AICglm.lm(cand.set = models_big, modnames = Modnames_big, :
## Check model structure carefully as some models may be redundant
```

```
##
## Model selection based on AICc:
##
##                                                                  K   AICc
## sqkmtotalcrop0.5km+ sqkmwater0.5km+Sum.of.area_FM3_17            4 367.05
## sqkmtotalcrop0.5km+ sqkmwater0.5km+population+Sum.of.area_FM3_17 5 369.11
## sqkmtotalcrop0.5km+sqkmwater0.5km                                3 372.46
## sqkmtotalcrop0.5km+ sqkmwater0.5km+population                    4 372.91
## sqkmtotalcrop2km+ sqkmwater2km+Sum.of.area_FM3_17                4 374.33
## sqkmtotalcrop2km+ sqkmwater2km+population+Sum.of.area_FM3_17     5 377.55
## sqkmwater2km+Sum.of.area_FM3_17                                  3 386.96
## sqkmwater2km+Sum.of.area_FM3_17+population                       4 389.45
## sqkmtotalcrop0.5km+Sum.of.area_FM3_17                            3 392.75
## sqkmtotalcrop0.5km+population+Sum.of.area_FM3_17                 4 395.35
## sqkmtotalcrop0.5km                                               2 403.34
## sqkmwater1km+Sum.of.area_FM3_17                                  3 404.16
## sqkmtotalcrop0.5km+population                                    3 404.61
## sqkmtotalcrop2km+Sum.of.area_FM3_17                              3 404.87
## sqkmwater1km+Sum.of.area_FM3_17+population                       4 405.92
## sqkmtotalcrop1km+ sqkmwater1km+Sum.of.area_FM3_17                4 407.11
## sqkmtotalcrop2km+population+Sum.of.area_FM3_17                   4 407.28
## sqkmtotalcrop1km+ sqkmwater1km+population+Sum.of.area_FM3_17     5 409.08
## sqkmwater0.5km+Sum.of.area_FM3_17+population                     4 411.04
## population +Sum.of.area_FM3_17+sqkmwater0.5km                    4 411.04
## sqkmwater0.5km+Sum.of.area_FM3_17                                3 411.77
## population +Sum.of.area_FM3_17                                   3 420.04
## Sum.of.area_FM3_17                                               2 420.73
## sqkmtotalcrop1km+population+Sum.of.area_FM3_17                   4 423.00
## sqkmtotalcrop1km+Sum.of.area_FM3_17                              3 423.01
## sqkmtotalcrop2km+sqkmwater2km                                    3 435.60
## sqkmtotalcrop2km+ sqkmwater2km+population                        4 438.14
## sqkmwater2km+population                                          3 455.24
## sqkmwater2km                                                     2 456.06
## sqkmtotalcrop2km                                                 2 458.78
## sqkmtotalcrop2km+population                                      3 460.09
## sqkmwater0.5km+population                                        3 466.30
## sqkmwater0.5km                                                   2 469.31
```

```
## sqkmtotalcrop1km+ sqkmwater1km+population                         4 469.95
## sqkmtotalcrop1km+population                                        3 472.92
## sqkmtotalcrop1km+sqkmwater1km                                      3 474.42
## sqkmwater1km+population                                            3 475.05
## population                                                         2 476.06
## sqkmwater1km                                                       2 477.03
## sqkmtotalcrop1km                                                   2 478.40
##                                                                   Delta_AICc
## sqkmtotalcrop0.5km+ sqkmwater0.5km+Sum.of.area_FM3_17                   0.00
## sqkmtotalcrop0.5km+ sqkmwater0.5km+population+Sum.of.area_FM3_17        2.06
## sqkmtotalcrop0.5km+sqkmwater0.5km                                       5.42
## sqkmtotalcrop0.5km+ sqkmwater0.5km+population                           5.87
## sqkmtotalcrop2km+ sqkmwater2km+Sum.of.area_FM3_17                       7.28
## sqkmtotalcrop2km+ sqkmwater2km+population+Sum.of.area_FM3_17           10.51
## sqkmwater2km+Sum.of.area_FM3_17                                        19.91
## sqkmwater2km+Sum.of.area_FM3_17+population                             22.40
## sqkmtotalcrop0.5km+Sum.of.area_FM3_17                                  25.70
## sqkmtotalcrop0.5km+population+Sum.of.area_FM3_17                       28.31
## sqkmtotalcrop0.5km                                                     36.29
## sqkmwater1km+Sum.of.area_FM3_17                                        37.11
## sqkmtotalcrop0.5km+population                                          37.56
## sqkmtotalcrop2km+Sum.of.area_FM3_17                                    37.83
## sqkmwater1km+Sum.of.area_FM3_17+population                             38.88
## sqkmtotalcrop1km+ sqkmwater1km+Sum.of.area_FM3_17                      40.07
## sqkmtotalcrop2km+population+Sum.of.area_FM3_17                         40.24
## sqkmtotalcrop1km+ sqkmwater1km+population+Sum.of.area_FM3_17           42.03
## sqkmwater0.5km+Sum.of.area_FM3_17+population                           44.00
## population +Sum.of.area_FM3_17+sqkmwater0.5km                          44.00
## sqkmwater0.5km+Sum.of.area_FM3_17                                      44.72
## population +Sum.of.area_FM3_17                                         53.00
## Sum.of.area_FM3_17                                                     53.68
## sqkmtotalcrop1km+population+Sum.of.area_FM3_17                         55.96
## sqkmtotalcrop1km+Sum.of.area_FM3_17                                    55.97
## sqkmtotalcrop2km+sqkmwater2km                                          68.56
## sqkmtotalcrop2km+ sqkmwater2km+population                              71.09
## sqkmwater2km+population                                                88.20
## sqkmwater2km                                                           89.01
## sqkmtotalcrop2km                                                       91.74
## sqkmtotalcrop2km+population                                            93.04
## sqkmwater0.5km+population                                              99.25
## sqkmwater0.5km                                                        102.27
## sqkmtotalcrop1km+ sqkmwater1km+population                             102.91
## sqkmtotalcrop1km+population                                           105.87
## sqkmtotalcrop1km+sqkmwater1km                                         107.37
## sqkmwater1km+population                                               108.01
## population                                                            109.02
## sqkmwater1km                                                          109.98
## sqkmtotalcrop1km                                                      111.35
##                                                                   AICcWt Cum.Wt
## sqkmtotalcrop0.5km+ sqkmwater0.5km+Sum.of.area_FM3_17               0.66   0.66
## sqkmtotalcrop0.5km+ sqkmwater0.5km+population+Sum.of.area_FM3_17    0.24   0.90
## sqkmtotalcrop0.5km+sqkmwater0.5km                                   0.04   0.94
## sqkmtotalcrop0.5km+ sqkmwater0.5km+population                       0.04   0.98
## sqkmtotalcrop2km+ sqkmwater2km+Sum.of.area_FM3_17                   0.02   1.00
```

```
## sqkmtotalcrop2km+ sqkmwater2km+population+Sum.of.area_FM3_17       0.00   1.00
## sqkmwater2km+Sum.of.area_FM3_17                                     0.00   1.00
## sqkmwater2km+Sum.of.area_FM3_17+population                          0.00   1.00
## sqkmtotalcrop0.5km+Sum.of.area_FM3_17                               0.00   1.00
## sqkmtotalcrop0.5km+population+Sum.of.area_FM3_17                    0.00   1.00
## sqkmtotalcrop0.5km                                                  0.00   1.00
## sqkmwater1km+Sum.of.area_FM3_17                                     0.00   1.00
## sqkmtotalcrop0.5km+population                                       0.00   1.00
## sqkmtotalcrop2km+Sum.of.area_FM3_17                                 0.00   1.00
## sqkmwater1km+Sum.of.area_FM3_17+population                          0.00   1.00
## sqkmtotalcrop1km+ sqkmwater1km+Sum.of.area_FM3_17                   0.00   1.00
## sqkmtotalcrop2km+population+Sum.of.area_FM3_17                      0.00   1.00
## sqkmtotalcrop1km+ sqkmwater1km+population+Sum.of.area_FM3_17       0.00   1.00
## sqkmwater0.5km+Sum.of.area_FM3_17+population                        0.00   1.00
## population +Sum.of.area_FM3_17+sqkmwater0.5km                       0.00   1.00
## sqkmwater0.5km+Sum.of.area_FM3_17                                   0.00   1.00
## population +Sum.of.area_FM3_17                                      0.00   1.00
## Sum.of.area_FM3_17                                                  0.00   1.00
## sqkmtotalcrop1km+population+Sum.of.area_FM3_17                      0.00   1.00
## sqkmtotalcrop1km+Sum.of.area_FM3_17                                 0.00   1.00
## sqkmtotalcrop2km+sqkmwater2km                                       0.00   1.00
## sqkmtotalcrop2km+ sqkmwater2km+population                           0.00   1.00
## sqkmwater2km+population                                             0.00   1.00
## sqkmwater2km                                                        0.00   1.00
## sqkmtotalcrop2km                                                    0.00   1.00
## sqkmtotalcrop2km+population                                         0.00   1.00
## sqkmwater0.5km+population                                           0.00   1.00
## sqkmwater0.5km                                                      0.00   1.00
## sqkmtotalcrop1km+ sqkmwater1km+population                           0.00   1.00
## sqkmtotalcrop1km+population                                         0.00   1.00
## sqkmtotalcrop1km+sqkmwater1km                                       0.00   1.00
## sqkmwater1km+population                                             0.00   1.00
## population                                                          0.00   1.00
## sqkmwater1km                                                        0.00   1.00
## sqkmtotalcrop1km                                                    0.00   1.00
##                                                                        LL
## sqkmtotalcrop0.5km+ sqkmwater0.5km+Sum.of.area_FM3_17            -178.41
## sqkmtotalcrop0.5km+ sqkmwater0.5km+population+Sum.of.area_FM3_17 -177.79
## sqkmtotalcrop0.5km+sqkmwater0.5km                                -182.60
## sqkmtotalcrop0.5km+ sqkmwater0.5km+population                    -181.35
## sqkmtotalcrop2km+ sqkmwater2km+Sum.of.area_FM3_17                -182.05
## sqkmtotalcrop2km+ sqkmwater2km+population+Sum.of.area_FM3_17     -182.01
## sqkmwater2km+Sum.of.area_FM3_17                                  -189.85
## sqkmwater2km+Sum.of.area_FM3_17+population                       -189.61
## sqkmtotalcrop0.5km+Sum.of.area_FM3_17                            -192.74
## sqkmtotalcrop0.5km+population+Sum.of.area_FM3_17                 -192.57
## sqkmtotalcrop0.5km                                               -199.37
## sqkmwater1km+Sum.of.area_FM3_17                                  -198.45
## sqkmtotalcrop0.5km+population                                    -198.67
## sqkmtotalcrop2km+Sum.of.area_FM3_17                              -198.80
## sqkmwater1km+Sum.of.area_FM3_17+population                       -197.85
## sqkmtotalcrop1km+ sqkmwater1km+Sum.of.area_FM3_17                -198.45
## sqkmtotalcrop2km+population+Sum.of.area_FM3_17                   -198.53
## sqkmtotalcrop1km+ sqkmwater1km+population+Sum.of.area_FM3_17     -197.78
```

```
## sqkmwater0.5km+Sum.of.area_FM3_17+population                          -200.41
## population +Sum.of.area_FM3_17+sqkmwater0.5km                         -200.41
## sqkmwater0.5km+Sum.of.area_FM3_17                                     -202.25
## population +Sum.of.area_FM3_17                                        -206.39
## Sum.of.area_FM3_17                                                    -208.06
## sqkmtotalcrop1km+population+Sum.of.area_FM3_17                        -206.39
## sqkmtotalcrop1km+Sum.of.area_FM3_17                                   -207.88
## sqkmtotalcrop2km+sqkmwater2km                                         -214.17
## sqkmtotalcrop2km+ sqkmwater2km+population                             -213.96
## sqkmwater2km+population                                               -223.99
## sqkmwater2km                                                          -225.73
## sqkmtotalcrop2km                                                      -227.09
## sqkmtotalcrop2km+population                                           -226.41
## sqkmwater0.5km+population                                             -229.52
## sqkmwater0.5km                                                        -232.36
## sqkmtotalcrop1km+ sqkmwater1km+population                             -229.86
## sqkmtotalcrop1km+population                                           -232.83
## sqkmtotalcrop1km+sqkmwater1km                                         -233.58
## sqkmwater1km+population                                               -233.89
## population                                                            -235.73
## sqkmwater1km                                                          -236.21
## sqkmtotalcrop1km                                                      -236.90
```

```
#Table for paper
#MMAIC<- aictab(cand.set = models_big, modnames = Modnames_big, sort = TRUE)
#MMAIC<-round(MMAIC[2:8], digits=2)
#write.table(MMAIC, file = "MMAIC.txt", sep = ",", quote = FALSE, row.names = F)

summary(mm14.2)
```

```
##
## Call:
## glm(formula = cbind(infectedkids_baseline, negkids_baseline) ~
##     sqkmtotalcrop2km + sqkmwater2km + Sum.of.area_FM3_17, family = binomial,
##     data = landp1, na.action = na.omit)
##
## Deviance Residuals:
##     Min       1Q   Median       3Q      Max
## -7.5146  -2.0680   0.4107   2.9068   5.7356
##
## Coefficients:
##                      Estimate Std. Error z value Pr(>|z|)
## (Intercept)        -0.5032798  0.1994250  -2.524   0.0116 *
## sqkmtotalcrop2km    0.1449362  0.0366072   3.959 7.52e-05 ***
## sqkmwater2km        0.1683035  0.0297972   5.648 1.62e-08 ***
## Sum.of.area_FM3_17  0.0003037  0.0000417   7.284 3.25e-13 ***
## ---
## Signif. codes:  0 '***' 0.001 '**' 0.01 '*' 0.05 '.' 0.1 ' ' 1
##
## (Dispersion parameter for binomial family taken to be 1)
##
##     Null deviance: 390.94  on 22  degrees of freedom
## Residual deviance: 278.04  on 19  degrees of freedom
## AIC: 372.1
```

```
##
## Number of Fisher Scoring iterations: 5
```

```
add_ci(landp1, mm14.2, names = c("lcb", "ucb"))
```

```
##       villages waterway sqkmtotalcrop0.5km sqkmwater1km sqkmtotalcrop2km
## 1      Diokhor      lac           0.222936     0.948440         3.629160
## 2     Diokhoul      lac           0.788500     0.756350         2.207280
## 3         Foss      lac           0.407990     0.478890         2.970460
## 4     Gankette      lac           0.483480     0.103240         6.943366
## 5      Guidick      lac           0.474450     1.133790         5.071914
## 6      Lampsar    river           0.275712     0.464620         5.734602
## 7    MakaDiama    river           0.369406     1.235390         4.113670
## 8        Malla      lac           0.491930     0.871230         5.098390
## 9   Malla Tack      lac           0.552050     0.400590         5.322650
## 10    Mbakhana    river           0.167428     0.737780         5.567160
## 11       Mbane      lac           0.326500     0.000000         6.802490
## 12    Mbarigot    river           0.323032     0.490510         3.112438
## 13 MerinaGewel      lac           0.704450     0.000000         4.886382
## 14   Ndiawdoune    river           0.262928     0.709190         1.901738
## 15   NdiolMaure    river           0.557480     0.140310         5.853860
## 16        Syer      lac           0.763150     0.105024         4.875906
## 17        assy    river           0.295000     0.449110         4.722270
## 18    diaminar      lac           0.582590     0.445770         4.967710
## 19    minguene    river           0.491556     0.000000         0.205056
## 20      ndelle    river           0.320152     0.000000         0.365020
## 21   ndiakhaye      lac           0.419310     0.083660         3.801654
## 22     salguir    river           0.120500     0.095184         5.055170
## 23     thilla1    river           0.322340     0.652248         3.818372
##    sqkmwater2km sqkmtotalcrop1km sqkmwater0.5km Year.baseline kids_baseline
## 1      4.891430         0.893824       0.056200          2016           122
## 2      5.379250         2.088930       0.032890          2016            48
## 3      3.629500         0.478890       0.000800          2016            31
## 4      0.934820         0.103240       0.000000          2016            68
## 5      5.845900         1.133790       0.144990          2016            82
## 6      3.533780         0.209150       0.057054          2016           107
## 7      5.322240         0.709190       0.200770          2016            96
## 8      5.191700         0.464620       0.015360          2016            83
## 9      2.196150         1.235390       0.269990          2016           104
## 10     4.795540         1.290938       0.298140          2016           111
## 11     0.000000         1.579750       0.084260          2016           103
## 12     0.581034         1.351856       0.029946          2016           119
## 13     0.135702         1.657150       0.081500          2016           102
## 14     1.491240         1.988846       0.085850          2016            91
## 15     0.497988         0.842542       0.006762          2016            87
## 16     0.224136         1.961830       0.009800          2016           125
## 17     1.785180         1.385320       0.000000          2017            30
## 18     1.450450         1.684590       0.005330          2017            30
## 19     0.414610         1.386876       0.052068          2017            30
## 20     0.000000         1.327750       0.000000          2017            30
## 21     0.255246         1.170220       0.108730          2017            30
## 22     0.246150         0.193104       0.000000          2017            31
## 23     1.139672         0.365020       0.000000          2017            30
##    infectedkids_baseline_H infectedkids_baseline_mansoni baseline_CI
```

```
## 1                    110                           17          17
## 2                     34                            7           4
## 3                     28                            1           1
## 4                     57                           63          55
## 5                     67                            0           0
## 6                     81                           42          40
## 7                     85                           31          23
## 8                     82                           68          68
## 9                     99                            9           9
## 10                    57                           42          27
## 11                    88                           18          17
## 12                    52                           30          21
## 13                    87                           76          70
## 14                    57                           32          26
## 15                    37                           12           4
## 16                   121                           28          27
## 17                    20                            2           2
## 18                    27                            1           1
## 19                    21                            1           1
## 20                    15                            0           0
## 21                    28                            0           0
## 22                     3                            0           0
## 23                     7                            0           0
##    infectedkids_baseline population rural prevalence_baseline
## 1                    110       1500 23818          0.90163934
## 2                     37       1300  3369          0.77083333
## 3                     28        107  7901          0.90322581
## 4                     65        459  6473          0.95588235
## 5                     67        400  5623          0.81707317
## 6                     83       1623 24486          0.77570094
## 7                     93        609 12516          0.96875000
## 8                     82        648  6273          0.98795181
## 9                     99        833 10923          0.95192308
## 10                    72       1120 35465          0.64864865
## 11                    89       1852 66884          0.86407767
## 12                    61       1316 11575          0.51260504
## 13                    93        761 10419          0.91176471
## 14                    63       1542 32881          0.69230769
## 15                    45        771  5975          0.51724138
## 16                   122        866  9463          0.97600000
## 17                    20        165  1502          0.66666667
## 18                    27         91  4278          0.90000000
## 19                    21       1146 18862          0.70000000
## 20                    15       2200 27035          0.50000000
## 21                    28        517 13564          0.93333333
## 22                     3        237  2656          0.09677419
## 23                     7        521  7860          0.23333333
##    prevalence_baseline_H prevalence_baseline_M prevalence_reinfection_2017
## 1             0.90163934            0.13934426                          77
## 2             0.70833333            0.14583333                          67
## 3             0.90322581            0.03225807                          84
## 4             0.83823529            0.92647059                          82
## 5             0.81707317            0.00000000                          88
## 6             0.75700935            0.39252336                          49
```

```
## 7           0.88541667          0.32291667                          83
## 8           0.98795181          0.81927711                          85
## 9           0.95192308          0.08653846                          96
## 10          0.51351351          0.37837838                          34
## 11          0.85436893          0.17475728                          85
## 12          0.43697479          0.25210084                          18
## 13          0.85294118          0.74509804                          61
## 14          0.62637363          0.35164835                          38
## 15          0.42528736          0.13793103                          33
## 16          0.96800000          0.22400000                          99
## 17          0.66666667          0.06666667                          NA
## 18          0.90000000          0.03333333                          NA
## 19          0.70000000          0.03333333                          NA
## 20          0.50000000          0.00000000                          NA
## 21          0.93333333          0.00000000                          NA
## 22          0.09677419          0.00000000                          NA
## 23          0.23333333          0.00000000                          NA
##    prevalence_reinfection2018 kids.tested.reinfection.Sh.2017
## 1                          68                              82
## 2                          89                              39
## 3                          85                              25
## 4                          56                              57
## 5                          81                              69
## 6                          88                             102
## 7                          88                              91
## 8                          98                              76
## 9                          88                             101
## 10                         38                             103
## 11                         66                             100
## 12                         39                             106
## 13                         80                              90
## 14                         35                              82
## 15                         32                              84
## 16                         98                             109
## 17                         NA                              NA
## 18                         NA                              NA
## 19                         NA                              NA
## 20                         NA                              NA
## 21                         NA                              NA
## 22                         NA                              NA
## 23                         NA                              NA
##    kids.reinfected.Sm.2017 kids.reinfected.Sh.2017 kids.tested.Sh.2018
## 1                       17                      63                 111
## 2                        7                      26                  38
## 3                        1                      21                  26
## 4                       63                      47                  55
## 5                        0                      61                  77
## 6                       42                      47                  99
## 7                       31                      64                  90
## 8                       68                      53                  57
## 9                        9                      97                  98
## 10                      42                      56                  94
## 11                      18                      85                 100
## 12                      30                      17                 104
```

```
## 13                      76                       55                   87
## 14                      32                       31                   79
## 15                      12                       23                   73
## 16                      28                      108                  105
## 17                      NA                       NA                   NA
## 18                      NA                       NA                   NA
## 19                      NA                       NA                   NA
## 20                      NA                       NA                   NA
## 21                      NA                       NA                   NA
## 22                      NA                       NA                   NA
## 23                      NA                       NA                   NA
##    infected.kids.Sh.2018 Sum.of.area_FM3_17 AccessArea_FM4_17
## 1                     76           3369.060              4085
## 2                     34           5496.101              3976
## 3                     22           3031.277              3225
## 4                     31           1677.355              1750
## 5                     62           2536.556              3433
## 6                     87           3081.368              1143
## 7                     79           1608.964              1633
## 8                     56            519.440               488
## 9                     86           6416.058              4774
## 10                    36           1498.642              1099
## 11                    66           2090.476              2215
## 12                    41            848.107               817
## 13                    70           2820.774              2675
## 14                    28           1386.967              1510
## 15                    23           2691.446              1300
## 16                   103           6807.490              4722
## 17                    NA           1921.000              1921
## 18                    NA           4920.000              4920
## 19                    NA           3775.000              3775
## 20                    NA           1905.000              1905
## 21                    NA           2135.000              2135
## 22                    NA           2464.000              2464
## 23                    NA           1565.000              1565
##    EmergentArea_FM4_17 FloatingArea_FM4_17 Mud_Area_FM4_17 avgFloatFM1FM4
## 1                 2645               160.9         1279.10        1255.49
## 2                  545              1077.0         2354.00        2584.98
## 3                  591               342.0         2292.00         520.99
## 4                 1351                46.0          353.00         623.63
## 5                  722               678.8         2032.20         944.26
## 6                  645               136.0          362.00        1553.70
## 7                  429               189.0         1015.00         583.36
## 8                  158                22.0          308.00          81.46
## 9                 1080               975.9         2718.10        2481.98
## 10                 122                90.0          887.00         351.00
## 11                 969               257.0          989.00         882.07
## 12                 209                29.2          578.80         168.54
## 13                 695                25.2         1954.80         278.57
## 14                 722               157.0          631.00         692.73
## 15                 697                58.0         1050.38        1050.38
## 16                 834              1355.0         3347.20        3347.20
## 17                 213                73.0         1635.00             NA
## 18                1174               506.0         3240.00             NA
```

```
## 19              1307             1200.0         1268.00              NA
## 20               554              558.0          793.00              NA
## 21              1206              308.0          621.00              NA
## 22              1192              100.0         1172.00              NA
## 23               681              131.0          753.00              NA
##    DateofAgricovariates average.field.area number.of.referenced.fields
## 1                  2015               1.77                          90
## 2                  2015               2.47                          47
## 3                  2015               1.50                          42
## 4                  2015               1.47                          60
## 5                  2015               1.60                          51
## 6                  2015               0.88                          68
## 7                  2015               3.93                          49
## 8                  2015               0.74                          69
## 9                  2015               1.79                          83
## 10                 2015               0.82                          63
## 11                 2015               1.34                         122
## 12                 2015               0.55                          44
## 13                 2015               1.53                          46
## 14                 2015               0.83                          36
## 15                 2015               0.85                          58
## 16                 2015               3.26                          51
## 17                   NA                 NA                          NA
## 18                   NA                 NA                          NA
## 19                   NA                 NA                          NA
## 20                   NA                 NA                          NA
## 21                   NA                 NA                          NA
## 22                   NA                 NA                          NA
## 23                   NA                 NA                          NA
##    ha.total.of.referenced.fields fallow wmelon rice groundnut bean onion
## 1                         159.60     12      9    0        17    1     7
## 2                         116.30     34     29    0        17   21     1
## 3                          63.00      9      9    0        15    8     2
## 4                          88.00      2     27    0        23   18    15
## 5                          81.55      5     30    0        27    7     4
## 6                          59.79      2      0   40         1    0     5
## 7                         192.80     24      0  141         0    0     8
## 8                          50.76      2      4    2         1    1     9
## 9                         148.35     20     31    0        39   45     2
## 10                         51.90     23      1    7         1    1     0
## 11                        163.15     23     33    3        26    8    51
## 12                         24.15      0      4    3         1    7     4
## 13                         70.25      7      8    0        17   28     0
## 14                         29.90      2      0    2         1    2     7
## 15                         49.19      6      2   33         1    0     3
## 16                        166.05     72     58    0        14   10     3
## 17                            NA     NA     NA   NA        NA   NA    NA
## 18                            NA     NA     NA   NA        NA   NA    NA
## 19                            NA     NA     NA   NA        NA   NA    NA
## 20                            NA     NA     NA   NA        NA   NA    NA
## 21                            NA     NA     NA   NA        NA   NA    NA
## 22                            NA     NA     NA   NA        NA   NA    NA
## 23                            NA     NA     NA   NA        NA   NA    NA
##    sweetpotatos tomato othercrop nofertilizer  Urea  NPK DAP Manure Other
```

```
## 1            65      2        47         16.5 124.6  8.5 4.5    1.0   4.5
## 2            NA      1        14         59.3  32.0 17.0 3.0    5.0   0.0
## 3            12      1         9         24.8  24.6  4.0 1.6    8.0   0.0
## 4             0      0         4         61.5   5.8 17.8 0.0    0.5   2.5
## 5             0      0         9         27.1  31.0 14.0 7.3    0.0   2.3
## 6             0      7         5          4.0  33.1 10.9 9.0    0.0   2.8
## 7             0      1        19         38.8 134.0 18.0 0.0    2.0   0.0
## 8             5     15        13          4.0  24.6 15.2 2.0    1.5   3.5
## 9             9      0         2         61.3  61.5 17.3 4.3    0.0   4.0
## 10            0      2        18         36.9   9.0  1.8 0.0    4.3   0.0
## 11            7      2        10         49.8  76.0 16.9 5.5    7.5   7.5
## 12            0      0         6          7.0   3.8  4.0 0.5    8.9   0.0
## 13            0      0        11         52.5   4.3  5.0 5.0    3.5   0.0
## 14            0      1        17         14.5  14.2  1.3 0.0    0.0   0.0
## 15            0      0         4          6.3  32.1  7.1 2.7    0.0   1.0
## 16            1      2         1        128.5  28.5  5.8 1.5    1.0   0.8
## 17           NA     NA        NA           NA    NA   NA  NA     NA    NA
## 18           NA     NA        NA           NA    NA   NA  NA     NA    NA
## 19           NA     NA        NA           NA    NA   NA  NA     NA    NA
## 20           NA     NA        NA           NA    NA   NA  NA     NA    NA
## 21           NA     NA        NA           NA    NA   NA  NA     NA    NA
## 22           NA     NA        NA           NA    NA   NA  NA     NA    NA
## 23           NA     NA        NA           NA    NA   NA  NA     NA    NA
##    TotalFert no.insecticide spithoate.dimethoate DECIS15.deltamethrine Bomec
## 1      143.1           74.0                 80.1                   0.0   0.0
## 2       57.0           50.8                 53.5                   7.0   0.0
## 3       38.2           23.6                 31.5                   3.7   4.2
## 4       26.6           49.5                 31.0                   2.5   0.0
## 5       54.6           23.5                 56.1                   0.5   1.0
## 6       55.8           37.4                  8.9                   7.3   0.0
## 7      154.0          118.8                  5.5                  22.0   0.0
## 8       46.8           21.3                 11.1                   1.5   6.7
## 9       87.1           48.1                 73.0                   2.0  15.0
## 10      15.1           42.6                  3.3                   4.6   0.0
## 11     113.4           62.4                 79.4                   5.3  11.5
## 12      17.2           14.8                  2.8                   3.7   1.0
## 13      17.8           56.0                  8.8                   5.5   0.0
## 14      15.5           13.3                  6.2                   7.5   0.0
## 15      42.9           24.2                  2.8                   9.6   1.5
## 16      37.6           90.5                 62.3                   0.3   1.0
## 17        NA             NA                   NA                    NA    NA
## 18        NA             NA                   NA                    NA    NA
## 19        NA             NA                   NA                    NA    NA
## 20        NA             NA                   NA                    NA    NA
## 21        NA             NA                   NA                    NA    NA
## 22        NA             NA                   NA                    NA    NA
## 23        NA             NA                   NA                    NA    NA
##    Viden Metaforce Vertimec.abamectine Arsenal Optimal Furadan Laprine
## 1    0.0       0.0                 0.0     1.0     3.0     1.5       0
## 2    0.0       0.0                 0.0     0.0     0.0     0.0       0
## 3    0.0       0.0                 0.0     0.0     0.0     0.0       0
## 4    0.0       3.0                 2.0     0.0     0.0     0.0       0
## 5    0.0       0.0                 0.0     0.0     0.0     0.0       0
## 6    1.5       0.0                 2.0     0.0     0.0     1.6       0
```

```
## 7   39.0       0.0               0.0     6.0     0.0     1.0       0
## 8    0.0       0.0               0.0     3.0     1.2     0.0       6
## 9    0.0       0.3               6.0     0.0     4.0     0.0       0
## 10   0.0       0.0               0.5     0.0     0.0     0.0       0
## 11   0.0       1.4               0.0     0.0     1.5     0.0       0
## 12   0.0       0.0               0.5     0.0     1.0     0.0       0
## 13   0.0       0.0               0.0     0.0     0.0     0.0       0
## 14   0.0       0.0               1.5     0.0     0.0     0.0       0
## 15   0.0       0.0               2.3     0.7     0.0     2.5       0
## 16   0.0      12.0               0.0     0.0     0.0     0.0       0
## 17    NA        NA                NA      NA      NA      NA      NA
## 18    NA        NA                NA      NA      NA      NA      NA
## 19    NA        NA                NA      NA      NA      NA      NA
## 20    NA        NA                NA      NA      NA      NA      NA
## 21    NA        NA                NA      NA      NA      NA      NA
## 22    NA        NA                NA      NA      NA      NA      NA
## 23    NA        NA                NA      NA      NA      NA      NA
##    Other.insecticide Total.insecticide No.herbicide Spiriz.360.EC.Propanil
## 1                0.0              85.6        150.1                    0.0
## 2                5.0              65.5        116.3                    0.0
## 3                0.0              39.4         59.0                    1.0
## 4                0.0              38.5         88.0                    0.0
## 5                0.5              58.1         81.6                    0.0
## 6                1.3              22.6         30.5                   15.7
## 7                0.5              74.0         51.3                   83.0
## 8                0.0              29.5         44.0                    0.0
## 9                0.0             100.3        120.1                    0.5
## 10               1.0               9.4         45.4                    0.5
## 11               1.7             100.8        117.1                   11.0
## 12               0.5               9.5         17.8                    2.0
## 13               0.0              14.3         66.3                    0.0
## 14               1.5              16.7         21.7                    3.0
## 15               5.7              25.1         10.2                   14.5
## 16               0.0              75.6        161.1                    0.0
## 17                NA                NA           NA                     NA
## 18                NA                NA           NA                     NA
## 19                NA                NA           NA                     NA
## 20                NA                NA           NA                     NA
## 21                NA                NA           NA                     NA
## 22                NA                NA           NA                     NA
## 23                NA                NA           NA                     NA
##    Londax.Bensulfuron.methyl Calliherbe.2.4.D Other.herbicides Total.Herbicide
## 1                        2.0              1.0              7.5            10.5
## 2                        0.0              0.0              0.0             0.0
## 3                        3.0              0.0              0.0             4.0
## 4                        0.0              0.0              0.0             0.0
## 5                        0.0              0.0              0.0             0.0
## 6                        8.2              1.0              5.4            30.3
## 7                       52.5              0.0              6.0           141.5
## 8                        6.0              0.5              0.8             7.3
## 9                        0.3              0.0             27.5            28.3
## 10                       4.0              0.0              2.0             6.5
## 11                       3.6              4.0             31.5            50.1
## 12                       2.7              0.0              1.8             6.5
```

```
## 13                      1.0              3.0              3.0              7.0
## 14                      1.0              0.0              4.3              8.3
## 15                     16.0              2.3              8.5             41.3
## 16                      3.0              0.0              2.0              5.0
## 17                       NA               NA               NA               NA
## 18                       NA               NA               NA               NA
## 19                       NA               NA               NA               NA
## 20                       NA               NA               NA               NA
## 21                       NA               NA               NA               NA
## 22                       NA               NA               NA               NA
## 23                       NA               NA               NA               NA
##      prevbase prevre1 prevre2   baseprev accessArea negkids_baseline
## 1  0.90163934    0.77    0.68 0.90163934   3369.060               12
## 2  0.77083333    0.67    0.89 0.77083333   5496.101               11
## 3  0.90322581    0.84    0.85 0.90322581   3031.277                3
## 4  0.95588235    0.82    0.56 0.95588235   1677.355                3
## 5  0.81707317    0.88    0.81 0.81707317   2536.556               15
## 6  0.77570094    0.49    0.88 0.77570094   3081.368               24
## 7  0.96875000    0.83    0.88 0.96875000   1608.964                3
## 8  0.98795181    0.85    0.98 0.98795181    519.440                1
## 9  0.95192308    0.96    0.88 0.95192308   6416.058                5
## 10 0.64864865    0.34    0.38 0.64864865   1498.642               39
## 11 0.86407767    0.85    0.66 0.86407767   2090.476               14
## 12 0.51260504    0.18    0.39 0.51260504    848.107               58
## 13 0.91176471    0.61    0.80 0.91176471   2820.774                9
## 14 0.69230769    0.38    0.35 0.69230769   1386.967               28
## 15 0.51724138    0.33    0.32 0.51724138   2691.446               42
## 16 0.97600000    0.99    0.98 0.97600000   6807.490                3
## 17 0.66666667      NA      NA 0.66666667   1921.000               10
## 18 0.90000000      NA      NA 0.90000000   4920.000                3
## 19 0.70000000      NA      NA 0.70000000   3775.000                9
## 20 0.50000000      NA      NA 0.50000000   1905.000               15
## 21 0.93333333      NA      NA 0.93333333   2135.000                2
## 22 0.09677419      NA      NA 0.09677419   2464.000               28
## 23 0.23333333      NA      NA 0.23333333   1565.000               23
##    negkids_baselineM logtotalfert logavgFloat      pred       lcb       ucb
## 1                105     4.970508    7.136077 0.8663751 0.8366574 0.8913881
## 2                 41     4.060443    7.857860 0.9161669 0.8797411 0.9422833
## 3                 30     3.668677    6.257648 0.8113531 0.7792966 0.8397113
## 4                  5     3.317816    6.437159 0.7631232 0.7158852 0.8046504
## 5                 82     4.018183    6.851460 0.8793368 0.8495459 0.9038961
## 6                 65     4.039536    7.349038 0.8651285 0.8422517 0.8851401
## 7                 65     5.043425    6.370517 0.8141794 0.7767492 0.8465736
## 8                 15     3.867026    4.412313 0.7802661 0.7319325 0.8220043
## 9                 95     4.478473    7.817215 0.9299943 0.9032038 0.9497822
## 10                69     2.778819    5.863631 0.8271994 0.7928912 0.8568504
## 11                85     4.739701    6.783404 0.7535450 0.7036318 0.7974716
## 12                89     2.901422    5.133089 0.5752294 0.5230954 0.6257418
## 13                26     2.933857    5.633253 0.7473552 0.7135733 0.7783902
## 14                59     2.803360    6.542083 0.6093569 0.5547553 0.6613501
## 15                75     3.781914    6.957859 0.7766883 0.7424468 0.8075574
## 16                97     3.653252    8.116178 0.9096072 0.8722862 0.9368122
## 17                28           NA          NA 0.7436612 0.7179238 0.7678097
## 18                29           NA          NA 0.8760169 0.8473208 0.8999564
```

```
## 19                29          NA          NA 0.6776168 0.5954378 0.7501084
## 20                30          NA          NA 0.5320187 0.4503163 0.6120396
## 21                30          NA          NA 0.6768126 0.6396245 0.7118917
## 22                31          NA          NA 0.7348249 0.7005077 0.7665205
## 23                30          NA          NA 0.6720133 0.6376048 0.7046681
```

```
#this gives us the predicted values of the model , along with the confidence bands

(est <- cbind(Estimate = coef(mm14.2), confint(mm14.2)))
```

```
## Waiting for profiling to be done...

##                        Estimate        2.5 %        97.5 %
## (Intercept)         -0.5032797862 -0.893302915 -0.1108077207
## sqkmtotalcrop2km     0.1449361877  0.073148503  0.2167627207
## sqkmwater2km         0.1683035246  0.110458859  0.2273472537
## Sum.of.area_FM3_17   0.0003037405  0.000224144  0.0003878176
```

```
#To get incidence rate  (we exponentiate the coefficients.. and confidence intervals!):
exp(est)
```

```
##                     Estimate     2.5 %    97.5 %
## (Intercept)         0.6045446 0.4093016 0.8951108
## sqkmtotalcrop2km    1.1559658 1.0758903 1.2420494
## sqkmwater2km        1.1832957 1.1167904 1.2552657
## Sum.of.area_FM3_17  1.0003038 1.0002242 1.0003879
```

```
###  Repeat the above baseline and infection post treatment regression with S. mansoni:

   ### Baseline S. masoni:

mansoni1=glm(cbind(infectedkids_baseline_mansoni,negkids_baselineM)~ sqkmtotalcrop0.5km+ sqkmwater0.5km
mansoni2=glm(cbind(infectedkids_baseline_mansoni,negkids_baselineM)~ sqkmtotalcrop0.5km+population, na.
mansoni3=glm(cbind(infectedkids_baseline_mansoni,negkids_baselineM)~ sqkmtotalcrop0.5km+sqkmwater0.5km,
mansoni4=glm(cbind(infectedkids_baseline_mansoni,negkids_baselineM)~ sqkmtotalcrop0.5km, na.action=na.o
mansoni5=glm(cbind(infectedkids_baseline_mansoni,negkids_baselineM)~ sqkmwater0.5km, na.action=na.omit,
mansoni6=glm(cbind(infectedkids_baseline_mansoni,negkids_baselineM)~ sqkmwater0.5km+population, na.acti
mansoni7=glm(cbind(infectedkids_baseline_mansoni,negkids_baselineM)~ sqkmwater0.5km+Sum.of.area_FM3_17,
mansoni8=glm(cbind(infectedkids_baseline_mansoni,negkids_baselineM)~ sqkmwater0.5km+Sum.of.area_FM3_17+
mansoni9=glm(cbind(infectedkids_baseline_mansoni,negkids_baselineM)~ population , na.action=na.omit,dat
mansoni10=glm(cbind(infectedkids_baseline_mansoni,negkids_baselineM)~ population +Sum.of.area_FM3_17, n
mansoni11=glm(cbind(infectedkids_baseline_mansoni,negkids_baselineM)~ population +Sum.of.area_FM3_17+sq
mansoni12=glm(cbind(infectedkids_baseline_mansoni,negkids_baselineM)~ Sum.of.area_FM3_17, na.action=na.

mansoni13=glm(cbind(infectedkids_baseline_mansoni,negkids_baselineM)~ sqkmtotalcrop0.5km+Sum.of.area_FM
mansoni14=glm(cbind(infectedkids_baseline_mansoni,negkids_baselineM)~ sqkmtotalcrop0.5km+ sqkmwater0.5k
mansoni15=glm(cbind(infectedkids_baseline_mansoni,negkids_baselineM)~ sqkmtotalcrop0.5km+ sqkmwater0.5k
mansoni16=glm(cbind(infectedkids_baseline_mansoni,negkids_baselineM)~ sqkmtotalcrop0.5km+population+Sum

mansoni1.1=glm(cbind(infectedkids_baseline_mansoni,negkids_baselineM)~ sqkmtotalcrop1km+ sqkmwater1km+p
mansoni2.1=glm(cbind(infectedkids_baseline_mansoni,negkids_baselineM)~ sqkmtotalcrop1km+population, na.
mansoni3.1=glm(cbind(infectedkids_baseline_mansoni,negkids_baselineM)~ sqkmtotalcrop1km+sqkmwater1km, n
```

```
mansoni4.1=glm(cbind(infectedkids_baseline_mansoni,negkids_baselineM)~ sqkmtotalcrop1km, na.action=na.o
mansoni5.1=glm(cbind(infectedkids_baseline_mansoni,negkids_baselineM)~ sqkmwater1km, na.action=na.omit,
mansoni6.1=glm(cbind(infectedkids_baseline_mansoni,negkids_baselineM)~ sqkmwater1km+population, na.acti
mansoni7.1=glm(cbind(infectedkids_baseline_mansoni,negkids_baselineM)~ sqkmwater1km+Sum.of.area_FM3_17,
mansoni8.1=glm(cbind(infectedkids_baseline_mansoni,negkids_baselineM)~ sqkmwater1km+Sum.of.area_FM3_17+

mansoni13.1=glm(cbind(infectedkids_baseline_mansoni,negkids_baselineM)~ sqkmtotalcrop1km+Sum.of.area_FM
mansoni14.1=glm(cbind(infectedkids_baseline_mansoni,negkids_baselineM)~ sqkmtotalcrop1km+ sqkmwater1km+
mansoni15.1=glm(cbind(infectedkids_baseline_mansoni,negkids_baselineM)~ sqkmtotalcrop1km+ sqkmwater1km+
mansoni16.1=glm(cbind(infectedkids_baseline_mansoni,negkids_baselineM)~ sqkmtotalcrop1km+population+Sum

mansoni1.2=glm(cbind(infectedkids_baseline_mansoni,negkids_baselineM)~ sqkmtotalcrop2km+ sqkmwater2km+p
mansoni2.2=glm(cbind(infectedkids_baseline_mansoni,negkids_baselineM)~ sqkmtotalcrop2km+population, na.
mansoni3.2=glm(cbind(infectedkids_baseline_mansoni,negkids_baselineM)~ sqkmtotalcrop2km+sqkmwater2km, n
mansoni4.2=glm(cbind(infectedkids_baseline_mansoni,negkids_baselineM)~ sqkmtotalcrop2km, na.action=na.o
mansoni5.2=glm(cbind(infectedkids_baseline_mansoni,negkids_baselineM)~ sqkmwater2km, na.action=na.omit,
mansoni6.2=glm(cbind(infectedkids_baseline_mansoni,negkids_baselineM)~ sqkmwater2km+population, na.acti
mansoni7.2=glm(cbind(infectedkids_baseline_mansoni,negkids_baselineM)~ sqkmwater2km+Sum.of.area_FM3_17,
mansoni8.2=glm(cbind(infectedkids_baseline_mansoni,negkids_baselineM)~ sqkmwater2km+Sum.of.area_FM3_17+

mansoni13.2=glm(cbind(infectedkids_baseline_mansoni,negkids_baselineM)~ sqkmtotalcrop2km+Sum.of.area_FM
mansoni14.2=glm(cbind(infectedkids_baseline_mansoni,negkids_baselineM)~ sqkmtotalcrop2km+ sqkmwater2km+
mansoni15.2=glm(cbind(infectedkids_baseline_mansoni,negkids_baselineM)~ sqkmtotalcrop2km+ sqkmwater2km+
mansoni16.2=glm(cbind(infectedkids_baseline_mansoni,negkids_baselineM)~ sqkmtotalcrop2km+population+Sum


models_man<-list(mansoni1,mansoni2,mansoni3,mansoni4,mansoni5,mansoni6,mansoni7,mansoni8,mansoni9,manso
Modnames_man <- c('sqkmtotalcrop0.5km+ sqkmwater0.5km+population',
                  'sqkmtotalcrop0.5km+population',
                  'sqkmtotalcrop0.5km+sqkmwater0.5km',
                  'sqkmtotalcrop0.5km',
                  'sqkmwater0.5km',
                  'sqkmwater0.5km+population',
                  'sqkmwater0.5km+Sum.of.area_FM3_17',
                  'sqkmwater0.5km+Sum.of.area_FM3_17+population',
                  'population',
                  'population +Sum.of.area_FM3_17',
                  'population +Sum.of.area_FM3_17+sqkmwater0.5km',
                  'Sum.of.area_FM3_17',
                  'sqkmtotalcrop0.5km+Sum.of.area_FM3_17',
                  'sqkmtotalcrop0.5km+ sqkmwater0.5km+Sum.of.area_FM3_17',
                  'sqkmtotalcrop0.5km+ sqkmwater0.5km+population+Sum.of.area_FM3_17',
                  'sqkmtotalcrop0.5km+population+Sum.of.area_FM3_17',
                  'sqkmtotalcrop1km+ sqkmwater1km+population',
                  'sqkmtotalcrop1km+population',
                  'sqkmtotalcrop1km+sqkmwater1km',
                  'sqkmtotalcrop1km',
                  'sqkmwater1km',
                  'sqkmwater1km+population',
                  'sqkmwater1km+Sum.of.area_FM3_17',
                  'sqkmwater1km+Sum.of.area_FM3_17+population',
                  'sqkmtotalcrop1km+Sum.of.area_FM3_17',
                  'sqkmtotalcrop1km+ sqkmwater1km+Sum.of.area_FM3_17',
```

```r
                  'sqkmtotalcrop1km+ sqkmwater1km+population+Sum.of.area_FM3_17',
                  'sqkmtotalcrop1km+population+Sum.of.area_FM3_17',
                  'sqkmtotalcrop2km+ sqkmwater2km+population',
                  'sqkmtotalcrop2km+population',
                  'sqkmtotalcrop2km+sqkmwater2km',
                  'sqkmtotalcrop2km',
                  'sqkmwater2km',
                  'sqkmwater2km+population',
                  'sqkmwater2km+Sum.of.area_FM3_17',
                  'sqkmwater2km+Sum.of.area_FM3_17+population',
                  'sqkmtotalcrop2km+Sum.of.area_FM3_17',
                  'sqkmtotalcrop2km+ sqkmwater2km+Sum.of.area_FM3_17',
                  'sqkmtotalcrop2km+ sqkmwater2km+population+Sum.of.area_FM3_17',
                  'sqkmtotalcrop2km+population+Sum.of.area_FM3_17')
aictab(cand.set = models_man, modnames = Modnames_man, sort = TRUE)
```

```
## Warning in aictab.AICglm.lm(cand.set = models_man, modnames = Modnames_man, :
## Check model structure carefully as some models may be redundant
```

```
##
## Model selection based on AICc:
##
##                                                                  K   AICc
## sqkmtotalcrop0.5km+ sqkmwater0.5km+population+Sum.of.area_FM3_17 5 466.96
## sqkmtotalcrop0.5km+population+Sum.of.area_FM3_17                 4 471.27
## sqkmtotalcrop0.5km+ sqkmwater0.5km+Sum.of.area_FM3_17            4 472.10
## sqkmtotalcrop0.5km+Sum.of.area_FM3_17                            3 474.01
## sqkmtotalcrop2km+Sum.of.area_FM3_17                              3 528.72
## sqkmtotalcrop2km+population+Sum.of.area_FM3_17                   4 530.18
## sqkmtotalcrop2km+ sqkmwater2km+Sum.of.area_FM3_17                4 531.41
## sqkmtotalcrop2km+ sqkmwater2km+population+Sum.of.area_FM3_17     5 532.95
## sqkmtotalcrop1km+ sqkmwater1km+population+Sum.of.area_FM3_17     5 561.38
## sqkmtotalcrop1km+ sqkmwater1km+Sum.of.area_FM3_17                4 562.60
## sqkmwater1km+Sum.of.area_FM3_17+population                       4 564.61
## sqkmwater1km+Sum.of.area_FM3_17                                  3 571.73
## sqkmtotalcrop1km+Sum.of.area_FM3_17                              3 581.00
## sqkmtotalcrop1km+population+Sum.of.area_FM3_17                   4 581.46
## population +Sum.of.area_FM3_17                                   3 583.37
## sqkmwater0.5km+Sum.of.area_FM3_17+population                     4 583.77
## population +Sum.of.area_FM3_17+sqkmwater0.5km                    4 583.77
## sqkmwater2km+Sum.of.area_FM3_17+population                       4 585.18
## sqkmwater0.5km+Sum.of.area_FM3_17                                3 586.67
## Sum.of.area_FM3_17                                               2 586.81
## sqkmwater2km+Sum.of.area_FM3_17                                  3 589.06
## sqkmtotalcrop2km                                                 2 606.94
## sqkmtotalcrop2km+sqkmwater2km                                    3 608.56
## sqkmtotalcrop2km+population                                      3 609.04
## sqkmtotalcrop2km+ sqkmwater2km+population                        4 611.09
## sqkmtotalcrop1km+sqkmwater1km                                    3 625.10
## sqkmtotalcrop1km+ sqkmwater1km+population                        4 627.49
## sqkmtotalcrop1km                                                 2 631.17
## sqkmtotalcrop1km+population                                      3 633.46
## sqkmtotalcrop0.5km                                               2 649.27
```

```
## sqkmtotalcrop0.5km+sqkmwater0.5km                                  3 650.71
## sqkmtotalcrop0.5km+population                                      3 651.41
## sqkmtotalcrop0.5km+ sqkmwater0.5km+population                      4 653.01
## sqkmwater1km+population                                            3 655.83
## sqkmwater0.5km+population                                          3 656.27
## population                                                         2 656.95
## sqkmwater0.5km                                                     2 657.08
## sqkmwater1km                                                       2 657.42
## sqkmwater2km+population                                            3 659.56
## sqkmwater2km                                                       2 660.35
##                                                                    Delta_AICc
## sqkmtotalcrop0.5km+ sqkmwater0.5km+population+Sum.of.area_FM3_17         0.00
## sqkmtotalcrop0.5km+population+Sum.of.area_FM3_17                         4.31
## sqkmtotalcrop0.5km+ sqkmwater0.5km+Sum.of.area_FM3_17                    5.14
## sqkmtotalcrop0.5km+Sum.of.area_FM3_17                                    7.05
## sqkmtotalcrop2km+Sum.of.area_FM3_17                                     61.76
## sqkmtotalcrop2km+population+Sum.of.area_FM3_17                          63.22
## sqkmtotalcrop2km+ sqkmwater2km+Sum.of.area_FM3_17                       64.45
## sqkmtotalcrop2km+ sqkmwater2km+population+Sum.of.area_FM3_17            65.99
## sqkmtotalcrop1km+ sqkmwater1km+population+Sum.of.area_FM3_17            94.42
## sqkmtotalcrop1km+ sqkmwater1km+Sum.of.area_FM3_17                       95.64
## sqkmwater1km+Sum.of.area_FM3_17+population                              97.65
## sqkmwater1km+Sum.of.area_FM3_17                                        104.77
## sqkmtotalcrop1km+Sum.of.area_FM3_17                                    114.04
## sqkmtotalcrop1km+population+Sum.of.area_FM3_17                         114.50
## population +Sum.of.area_FM3_17                                         116.41
## sqkmwater0.5km+Sum.of.area_FM3_17+population                           116.81
## population +Sum.of.area_FM3_17+sqkmwater0.5km                          116.81
## sqkmwater2km+Sum.of.area_FM3_17+population                             118.22
## sqkmwater0.5km+Sum.of.area_FM3_17                                      119.71
## Sum.of.area_FM3_17                                                     119.85
## sqkmwater2km+Sum.of.area_FM3_17                                        122.10
## sqkmtotalcrop2km                                                       139.98
## sqkmtotalcrop2km+sqkmwater2km                                          141.60
## sqkmtotalcrop2km+population                                            142.08
## sqkmtotalcrop2km+ sqkmwater2km+population                              144.12
## sqkmtotalcrop1km+sqkmwater1km                                          158.14
## sqkmtotalcrop1km+ sqkmwater1km+population                              160.53
## sqkmtotalcrop1km                                                       164.21
## sqkmtotalcrop1km+population                                            166.50
## sqkmtotalcrop0.5km                                                     182.31
## sqkmtotalcrop0.5km+sqkmwater0.5km                                      183.75
## sqkmtotalcrop0.5km+population                                          184.45
## sqkmtotalcrop0.5km+ sqkmwater0.5km+population                          186.04
## sqkmwater1km+population                                                188.87
## sqkmwater0.5km+population                                              189.31
## population                                                             189.99
## sqkmwater0.5km                                                         190.12
## sqkmwater1km                                                           190.46
## sqkmwater2km+population                                                192.60
## sqkmwater2km                                                           193.39
##                                                                    AICcWt Cum.Wt
## sqkmtotalcrop0.5km+ sqkmwater0.5km+population+Sum.of.area_FM3_17     0.82   0.82
## sqkmtotalcrop0.5km+population+Sum.of.area_FM3_17                     0.09   0.91
```

```
## sqkmtotalcrop0.5km+ sqkmwater0.5km+Sum.of.area_FM3_17              0.06   0.98
## sqkmtotalcrop0.5km+Sum.of.area_FM3_17                              0.02   1.00
## sqkmtotalcrop2km+Sum.of.area_FM3_17                                0.00   1.00
## sqkmtotalcrop2km+population+Sum.of.area_FM3_17                     0.00   1.00
## sqkmtotalcrop2km+ sqkmwater2km+Sum.of.area_FM3_17                  0.00   1.00
## sqkmtotalcrop2km+ sqkmwater2km+population+Sum.of.area_FM3_17       0.00   1.00
## sqkmtotalcrop1km+ sqkmwater1km+population+Sum.of.area_FM3_17       0.00   1.00
## sqkmtotalcrop1km+ sqkmwater1km+Sum.of.area_FM3_17                  0.00   1.00
## sqkmwater1km+Sum.of.area_FM3_17+population                         0.00   1.00
## sqkmwater1km+Sum.of.area_FM3_17                                    0.00   1.00
## sqkmtotalcrop1km+Sum.of.area_FM3_17                                0.00   1.00
## sqkmtotalcrop1km+population+Sum.of.area_FM3_17                     0.00   1.00
## population +Sum.of.area_FM3_17                                     0.00   1.00
## sqkmwater0.5km+Sum.of.area_FM3_17+population                       0.00   1.00
## population +Sum.of.area_FM3_17+sqkmwater0.5km                      0.00   1.00
## sqkmwater2km+Sum.of.area_FM3_17+population                         0.00   1.00
## sqkmwater0.5km+Sum.of.area_FM3_17                                  0.00   1.00
## Sum.of.area_FM3_17                                                 0.00   1.00
## sqkmwater2km+Sum.of.area_FM3_17                                    0.00   1.00
## sqkmtotalcrop2km                                                   0.00   1.00
## sqkmtotalcrop2km+sqkmwater2km                                      0.00   1.00
## sqkmtotalcrop2km+population                                        0.00   1.00
## sqkmtotalcrop2km+ sqkmwater2km+population                          0.00   1.00
## sqkmtotalcrop1km+sqkmwater1km                                      0.00   1.00
## sqkmtotalcrop1km+ sqkmwater1km+population                          0.00   1.00
## sqkmtotalcrop1km                                                   0.00   1.00
## sqkmtotalcrop1km+population                                        0.00   1.00
## sqkmtotalcrop0.5km                                                 0.00   1.00
## sqkmtotalcrop0.5km+sqkmwater0.5km                                  0.00   1.00
## sqkmtotalcrop0.5km+population                                      0.00   1.00
## sqkmtotalcrop0.5km+ sqkmwater0.5km+population                      0.00   1.00
## sqkmwater1km+population                                            0.00   1.00
## sqkmwater0.5km+population                                          0.00   1.00
## population                                                         0.00   1.00
## sqkmwater0.5km                                                     0.00   1.00
## sqkmwater1km                                                       0.00   1.00
## sqkmwater2km+population                                            0.00   1.00
## sqkmwater2km                                                       0.00   1.00
##                                                                       LL
## sqkmtotalcrop0.5km+ sqkmwater0.5km+population+Sum.of.area_FM3_17 -226.72
## sqkmtotalcrop0.5km+population+Sum.of.area_FM3_17                 -230.53
## sqkmtotalcrop0.5km+ sqkmwater0.5km+Sum.of.area_FM3_17            -230.94
## sqkmtotalcrop0.5km+Sum.of.area_FM3_17                            -233.37
## sqkmtotalcrop2km+Sum.of.area_FM3_17                              -260.73
## sqkmtotalcrop2km+population+Sum.of.area_FM3_17                   -259.98
## sqkmtotalcrop2km+ sqkmwater2km+Sum.of.area_FM3_17                -260.59
## sqkmtotalcrop2km+ sqkmwater2km+population+Sum.of.area_FM3_17     -259.71
## sqkmtotalcrop1km+ sqkmwater1km+population+Sum.of.area_FM3_17     -273.92
## sqkmtotalcrop1km+ sqkmwater1km+Sum.of.area_FM3_17                -276.19
## sqkmwater1km+Sum.of.area_FM3_17+population                       -277.20
## sqkmwater1km+Sum.of.area_FM3_17                                  -282.24
## sqkmtotalcrop1km+Sum.of.area_FM3_17                              -286.87
## sqkmtotalcrop1km+population+Sum.of.area_FM3_17                   -285.62
## population +Sum.of.area_FM3_17                                   -288.05
```

```
## sqkmwater0.5km+Sum.of.area_FM3_17+population                         -286.77
## population +Sum.of.area_FM3_17+sqkmwater0.5km                        -286.77
## sqkmwater2km+Sum.of.area_FM3_17+population                           -287.48
## sqkmwater0.5km+Sum.of.area_FM3_17                                    -289.71
## Sum.of.area_FM3_17                                                   -291.10
## sqkmwater2km+Sum.of.area_FM3_17                                      -290.90
## sqkmtotalcrop2km                                                     -301.17
## sqkmtotalcrop2km+sqkmwater2km                                        -300.65
## sqkmtotalcrop2km+population                                          -300.89
## sqkmtotalcrop2km+ sqkmwater2km+population                            -300.43
## sqkmtotalcrop1km+sqkmwater1km                                        -308.92
## sqkmtotalcrop1km+ sqkmwater1km+population                            -308.63
## sqkmtotalcrop1km                                                     -313.29
## sqkmtotalcrop1km+population                                          -313.10
## sqkmtotalcrop0.5km                                                   -322.33
## sqkmtotalcrop0.5km+sqkmwater0.5km                                    -321.73
## sqkmtotalcrop0.5km+population                                        -322.07
## sqkmtotalcrop0.5km+ sqkmwater0.5km+population                        -321.39
## sqkmwater1km+population                                              -324.28
## sqkmwater0.5km+population                                            -324.50
## population                                                           -326.18
## sqkmwater0.5km                                                       -326.24
## sqkmwater1km                                                         -326.41
## sqkmwater2km+population                                              -326.15
## sqkmwater2km                                                         -327.88
```

```
summary(mansoni15)
```

```
##
## Call:
## glm(formula = cbind(infectedkids_baseline_mansoni, negkids_baselineM) ~
##     sqkmtotalcrop0.5km + sqkmwater0.5km + population + Sum.of.area_FM3_17,
##     family = binomial, data = landp1, na.action = na.omit)
##
## Deviance Residuals:
##    Min      1Q  Median      3Q     Max
## -8.048  -3.304  -1.669   2.344   9.296
##
## Coefficients:
##                      Estimate Std. Error z value Pr(>|z|)
## (Intercept)        -2.1654062  0.2997345  -7.224 5.03e-13 ***
## sqkmtotalcrop0.5km  5.3483165  0.5322518  10.048  < 2e-16 ***
## sqkmwater0.5km      2.0605424  0.7446924   2.767  0.00566 **
## population          0.0003877  0.0001352   2.869  0.00412 **
## Sum.of.area_FM3_17 -0.0006115  0.0000492 -12.428  < 2e-16 ***
## ---
## Signif. codes:  0 '***' 0.001 '**' 0.01 '*' 0.05 '.' 0.1 ' ' 1
##
## (Dispersion parameter for binomial family taken to be 1)
##
##     Null deviance: 584.20  on 22  degrees of freedom
## Residual deviance: 381.71  on 18  degrees of freedom
## AIC: 463.43
##
```

```
## Number of Fisher Scoring iterations: 5
```

```r
add_ci(landp1, mansoni15, names = c("lcb", "ucb"))
```

```
##       villages waterway sqkmtotalcrop0.5km sqkmwater1km sqkmtotalcrop2km
## 1      Diokhor      lac           0.222936     0.948440         3.629160
## 2     Diokhoul      lac           0.788500     0.756350         2.207280
## 3         Foss      lac           0.407990     0.478890         2.970460
## 4     Gankette      lac           0.483480     0.103240         6.943366
## 5      Guidick      lac           0.474450     1.133790         5.071914
## 6      Lampsar    river           0.275712     0.464620         5.734602
## 7    MakaDiama    river           0.369406     1.235390         4.113670
## 8        Malla      lac           0.491930     0.871230         5.098390
## 9   Malla Tack      lac           0.552050     0.400590         5.322650
## 10    Mbakhana    river           0.167428     0.737780         5.567160
## 11       Mbane      lac           0.326500     0.000000         6.802490
## 12    Mbarigot    river           0.323032     0.490510         3.112438
## 13 MerinaGewel      lac           0.704450     0.000000         4.886382
## 14   Ndiawdoune    river           0.262928     0.709190         1.901738
## 15   NdiolMaure    river           0.557480     0.140310         5.853860
## 16        Syer      lac           0.763150     0.105024         4.875906
## 17        assy    river           0.295000     0.449110         4.722270
## 18    diaminar      lac           0.582590     0.445770         4.967710
## 19    minguene    river           0.491556     0.000000         0.205056
## 20      ndelle    river           0.320152     0.000000         0.365020
## 21   ndiakhaye      lac           0.419310     0.083660         3.801654
## 22     salguir    river           0.120500     0.095184         5.055170
## 23     thilla1    river           0.322340     0.652248         3.818372
##    sqkmwater2km sqkmtotalcrop1km sqkmwater0.5km Year.baseline kids_baseline
## 1      4.891430         0.893824       0.056200          2016           122
## 2      5.379250         2.088930       0.032890          2016            48
## 3      3.629500         0.478890       0.000800          2016            31
## 4      0.934820         0.103240       0.000000          2016            68
## 5      5.845900         1.133790       0.144990          2016            82
## 6      3.533780         0.209150       0.057054          2016           107
## 7      5.322240         0.709190       0.200770          2016            96
## 8      5.191700         0.464620       0.015360          2016            83
## 9      2.196150         1.235390       0.269990          2016           104
## 10     4.795540         1.290938       0.298140          2016           111
## 11     0.000000         1.579750       0.084260          2016           103
## 12     0.581034         1.351856       0.029946          2016           119
## 13     0.135702         1.657150       0.081500          2016           102
## 14     1.491240         1.988846       0.085850          2016            91
## 15     0.497988         0.842542       0.006762          2016            87
## 16     0.224136         1.961830       0.009800          2016           125
## 17     1.785180         1.385320       0.000000          2017            30
## 18     1.450450         1.684590       0.005330          2017            30
## 19     0.414610         1.386876       0.052068          2017            30
## 20     0.000000         1.327750       0.000000          2017            30
## 21     0.255246         1.170220       0.108730          2017            30
## 22     0.246150         0.193104       0.000000          2017            31
## 23     1.139672         0.365020       0.000000          2017            30
##    infectedkids_baseline_H infectedkids_baseline_mansoni baseline_CI
## 1                      110                            17          17
```

```
## 2                      34                              7           4
## 3                      28                              1           1
## 4                      57                             63          55
## 5                      67                              0           0
## 6                      81                             42          40
## 7                      85                             31          23
## 8                      82                             68          68
## 9                      99                              9           9
## 10                     57                             42          27
## 11                     88                             18          17
## 12                     52                             30          21
## 13                     87                             76          70
## 14                     57                             32          26
## 15                     37                             12           4
## 16                    121                             28          27
## 17                     20                              2           2
## 18                     27                              1           1
## 19                     21                              1           1
## 20                     15                              0           0
## 21                     28                              0           0
## 22                      3                              0           0
## 23                      7                              0           0
##    infectedkids_baseline population rural prevalence_baseline
## 1                    110       1500 23818          0.90163934
## 2                     37       1300  3369          0.77083333
## 3                     28        107  7901          0.90322581
## 4                     65        459  6473          0.95588235
## 5                     67        400  5623          0.81707317
## 6                     83       1623 24486          0.77570094
## 7                     93        609 12516          0.96875000
## 8                     82        648  6273          0.98795181
## 9                     99        833 10923          0.95192308
## 10                    72       1120 35465          0.64864865
## 11                    89       1852 66884          0.86407767
## 12                    61       1316 11575          0.51260504
## 13                    93        761 10419          0.91176471
## 14                    63       1542 32881          0.69230769
## 15                    45        771  5975          0.51724138
## 16                   122        866  9463          0.97600000
## 17                    20        165  1502          0.66666667
## 18                    27         91  4278          0.90000000
## 19                    21       1146 18862          0.70000000
## 20                    15       2200 27035          0.50000000
## 21                    28        517 13564          0.93333333
## 22                     3        237  2656          0.09677419
## 23                     7        521  7860          0.23333333
##    prevalence_baseline_H prevalence_baseline_M prevalence_reinfection_2017
## 1             0.90163934            0.13934426                          77
## 2             0.70833333            0.14583333                          67
## 3             0.90322581            0.03225807                          84
## 4             0.83823529            0.92647059                          82
## 5             0.81707317            0.00000000                          88
## 6             0.75700935            0.39252336                          49
## 7             0.88541667            0.32291667                          83
```

```
## 8           0.98795181          0.81927711                          85
## 9           0.95192308          0.08653846                          96
## 10          0.51351351          0.37837838                          34
## 11          0.85436893          0.17475728                          85
## 12          0.43697479          0.25210084                          18
## 13          0.85294118          0.74509804                          61
## 14          0.62637363          0.35164835                          38
## 15          0.42528736          0.13793103                          33
## 16          0.96800000          0.22400000                          99
## 17          0.66666667          0.06666667                          NA
## 18          0.90000000          0.03333333                          NA
## 19          0.70000000          0.03333333                          NA
## 20          0.50000000          0.00000000                          NA
## 21          0.93333333          0.00000000                          NA
## 22          0.09677419          0.00000000                          NA
## 23          0.23333333          0.00000000                          NA
##    prevalence_reinfection2018 kids.tested.reinfection.Sh.2017
## 1                          68                              82
## 2                          89                              39
## 3                          85                              25
## 4                          56                              57
## 5                          81                              69
## 6                          88                             102
## 7                          88                              91
## 8                          98                              76
## 9                          88                             101
## 10                         38                             103
## 11                         66                             100
## 12                         39                             106
## 13                         80                              90
## 14                         35                              82
## 15                         32                              84
## 16                         98                             109
## 17                         NA                              NA
## 18                         NA                              NA
## 19                         NA                              NA
## 20                         NA                              NA
## 21                         NA                              NA
## 22                         NA                              NA
## 23                         NA                              NA
##    kids.reinfected.Sm.2017 kids.reinfected.Sh.2017 kids.tested.Sh.2018
## 1                       17                      63                 111
## 2                        7                      26                  38
## 3                        1                      21                  26
## 4                       63                      47                  55
## 5                        0                      61                  77
## 6                       42                      47                  99
## 7                       31                      64                  90
## 8                       68                      53                  57
## 9                        9                      97                  98
## 10                      42                      56                  94
## 11                      18                      85                 100
## 12                      30                      17                 104
## 13                      76                      55                  87
```

```
## 14                      32                      31                   79
## 15                      12                      23                   73
## 16                      28                     108                  105
## 17                      NA                      NA                   NA
## 18                      NA                      NA                   NA
## 19                      NA                      NA                   NA
## 20                      NA                      NA                   NA
## 21                      NA                      NA                   NA
## 22                      NA                      NA                   NA
## 23                      NA                      NA                   NA
##    infected.kids.Sh.2018 Sum.of.area_FM3_17 AccessArea_FM4_17
## 1                     76           3369.060              4085
## 2                     34           5496.101              3976
## 3                     22           3031.277              3225
## 4                     31           1677.355              1750
## 5                     62           2536.556              3433
## 6                     87           3081.368              1143
## 7                     79           1608.964              1633
## 8                     56            519.440               488
## 9                     86           6416.058              4774
## 10                    36           1498.642              1099
## 11                    66           2090.476              2215
## 12                    41            848.107               817
## 13                    70           2820.774              2675
## 14                    28           1386.967              1510
## 15                    23           2691.446              1300
## 16                   103           6807.490              4722
## 17                    NA           1921.000              1921
## 18                    NA           4920.000              4920
## 19                    NA           3775.000              3775
## 20                    NA           1905.000              1905
## 21                    NA           2135.000              2135
## 22                    NA           2464.000              2464
## 23                    NA           1565.000              1565
##    EmergentArea_FM4_17 FloatingArea_FM4_17 Mud_Area_FM4_17 avgFloatFM1FM4
## 1                 2645               160.9         1279.10        1255.49
## 2                  545              1077.0         2354.00        2584.98
## 3                  591               342.0         2292.00         520.99
## 4                 1351                46.0          353.00         623.63
## 5                  722               678.8         2032.20         944.26
## 6                  645               136.0          362.00        1553.70
## 7                  429               189.0         1015.00         583.36
## 8                  158                22.0          308.00          81.46
## 9                 1080               975.9         2718.10        2481.98
## 10                 122                90.0          887.00         351.00
## 11                 969               257.0          989.00         882.07
## 12                 209                29.2          578.80         168.54
## 13                 695                25.2         1954.80         278.57
## 14                 722               157.0          631.00         692.73
## 15                 697                58.0         1050.38        1050.38
## 16                 834              1355.0         3347.20        3347.20
## 17                 213                73.0         1635.00             NA
## 18                1174               506.0         3240.00             NA
## 19                1307              1200.0         1268.00             NA
```

```
## 20                554               558.0          793.00             NA
## 21               1206               308.0          621.00             NA
## 22               1192               100.0         1172.00             NA
## 23                681               131.0          753.00             NA
##    DateofAgricovariates average.field.area number.of.referenced.fields
## 1                  2015               1.77                          90
## 2                  2015               2.47                          47
## 3                  2015               1.50                          42
## 4                  2015               1.47                          60
## 5                  2015               1.60                          51
## 6                  2015               0.88                          68
## 7                  2015               3.93                          49
## 8                  2015               0.74                          69
## 9                  2015               1.79                          83
## 10                 2015               0.82                          63
## 11                 2015               1.34                         122
## 12                 2015               0.55                          44
## 13                 2015               1.53                          46
## 14                 2015               0.83                          36
## 15                 2015               0.85                          58
## 16                 2015               3.26                          51
## 17                   NA                 NA                          NA
## 18                   NA                 NA                          NA
## 19                   NA                 NA                          NA
## 20                   NA                 NA                          NA
## 21                   NA                 NA                          NA
## 22                   NA                 NA                          NA
## 23                   NA                 NA                          NA
##    ha.total.of.referenced.fields fallow wmelon rice groundnut bean onion
## 1                         159.60     12      9    0        17    1     7
## 2                         116.30     34     29    0        17   21     1
## 3                          63.00      9      9    0        15    8     2
## 4                          88.00      2     27    0        23   18    15
## 5                          81.55      5     30    0        27    7     4
## 6                          59.79      2      0   40         1    0     5
## 7                         192.80     24      0  141         0    0     8
## 8                          50.76      2      4    2         1    1     9
## 9                         148.35     20     31    0        39   45     2
## 10                         51.90     23      1    7         1    1     0
## 11                        163.15     23     33    3        26    8    51
## 12                         24.15      0      4    3         1    7     4
## 13                         70.25      7      8    0        17   28     0
## 14                         29.90      2      0    2         1    2     7
## 15                         49.19      6      2   33         1    0     3
## 16                        166.05     72     58    0        14   10     3
## 17                            NA     NA     NA   NA        NA   NA    NA
## 18                            NA     NA     NA   NA        NA   NA    NA
## 19                            NA     NA     NA   NA        NA   NA    NA
## 20                            NA     NA     NA   NA        NA   NA    NA
## 21                            NA     NA     NA   NA        NA   NA    NA
## 22                            NA     NA     NA   NA        NA   NA    NA
## 23                            NA     NA     NA   NA        NA   NA    NA
##    sweetpotatos tomato othercrop nofertilizer  Urea  NPK DAP Manure Other
## 1            65      2        47         16.5 124.6  8.5 4.5    1.0   4.5
```

```
## 2           NA      1        14         59.3  32.0 17.0 3.0    5.0   0.0
## 3           12      1         9         24.8  24.6  4.0 1.6    8.0   0.0
## 4            0      0         4         61.5   5.8 17.8 0.0    0.5   2.5
## 5            0      0         9         27.1  31.0 14.0 7.3    0.0   2.3
## 6            0      7         5          4.0  33.1 10.9 9.0    0.0   2.8
## 7            0      1        19         38.8 134.0 18.0 0.0    2.0   0.0
## 8            5     15        13          4.0  24.6 15.2 2.0    1.5   3.5
## 9            9      0         2         61.3  61.5 17.3 4.3    0.0   4.0
## 10           0      2        18         36.9   9.0  1.8 0.0    4.3   0.0
## 11           7      2        10         49.8  76.0 16.9 5.5    7.5   7.5
## 12           0      0         6          7.0   3.8  4.0 0.5    8.9   0.0
## 13           0      0        11         52.5   4.3  5.0 5.0    3.5   0.0
## 14           0      1        17         14.5  14.2  1.3 0.0    0.0   0.0
## 15           0      0         4          6.3  32.1  7.1 2.7    0.0   1.0
## 16           1      2         1        128.5  28.5  5.8 1.5    1.0   0.8
## 17          NA     NA        NA           NA    NA   NA  NA     NA    NA
## 18          NA     NA        NA           NA    NA   NA  NA     NA    NA
## 19          NA     NA        NA           NA    NA   NA  NA     NA    NA
## 20          NA     NA        NA           NA    NA   NA  NA     NA    NA
## 21          NA     NA        NA           NA    NA   NA  NA     NA    NA
## 22          NA     NA        NA           NA    NA   NA  NA     NA    NA
## 23          NA     NA        NA           NA    NA   NA  NA     NA    NA
##    TotalFert no.insecticide spithoate.dimethoate DECIS15.deltamethrine Bomec
## 1      143.1           74.0                 80.1                   0.0   0.0
## 2       57.0           50.8                 53.5                   7.0   0.0
## 3       38.2           23.6                 31.5                   3.7   4.2
## 4       26.6           49.5                 31.0                   2.5   0.0
## 5       54.6           23.5                 56.1                   0.5   1.0
## 6       55.8           37.4                  8.9                   7.3   0.0
## 7      154.0          118.8                  5.5                  22.0   0.0
## 8       46.8           21.3                 11.1                   1.5   6.7
## 9       87.1           48.1                 73.0                   2.0  15.0
## 10      15.1           42.6                  3.3                   4.6   0.0
## 11     113.4           62.4                 79.4                   5.3  11.5
## 12      17.2           14.8                  2.8                   3.7   1.0
## 13      17.8           56.0                  8.8                   5.5   0.0
## 14      15.5           13.3                  6.2                   7.5   0.0
## 15      42.9           24.2                  2.8                   9.6   1.5
## 16      37.6           90.5                 62.3                   0.3   1.0
## 17        NA             NA                   NA                    NA    NA
## 18        NA             NA                   NA                    NA    NA
## 19        NA             NA                   NA                    NA    NA
## 20        NA             NA                   NA                    NA    NA
## 21        NA             NA                   NA                    NA    NA
## 22        NA             NA                   NA                    NA    NA
## 23        NA             NA                   NA                    NA    NA
##    Viden Metaforce Vertimec.abamectine Arsenal Optimal Furadan Laprine
## 1    0.0       0.0                 0.0     1.0     3.0     1.5       0
## 2    0.0       0.0                 0.0     0.0     0.0     0.0       0
## 3    0.0       0.0                 0.0     0.0     0.0     0.0       0
## 4    0.0       3.0                 2.0     0.0     0.0     0.0       0
## 5    0.0       0.0                 0.0     0.0     0.0     0.0       0
## 6    1.5       0.0                 2.0     0.0     0.0     1.6       0
## 7   39.0       0.0                 0.0     6.0     0.0     1.0       0
```

```
## 8    0.0       0.0                 0.0     3.0     1.2     0.0       6
## 9    0.0       0.3                 6.0     0.0     4.0     0.0       0
## 10   0.0       0.0                 0.5     0.0     0.0     0.0       0
## 11   0.0       1.4                 0.0     0.0     1.5     0.0       0
## 12   0.0       0.0                 0.5     0.0     1.0     0.0       0
## 13   0.0       0.0                 0.0     0.0     0.0     0.0       0
## 14   0.0       0.0                 1.5     0.0     0.0     0.0       0
## 15   0.0       0.0                 2.3     0.7     0.0     2.5       0
## 16   0.0      12.0                 0.0     0.0     0.0     0.0       0
## 17    NA        NA                  NA      NA      NA      NA      NA
## 18    NA        NA                  NA      NA      NA      NA      NA
## 19    NA        NA                  NA      NA      NA      NA      NA
## 20    NA        NA                  NA      NA      NA      NA      NA
## 21    NA        NA                  NA      NA      NA      NA      NA
## 22    NA        NA                  NA      NA      NA      NA      NA
## 23    NA        NA                  NA      NA      NA      NA      NA
##    Other.insecticide Total.insecticide No.herbicide Spiriz.360.EC.Propanil
## 1                0.0              85.6        150.1                    0.0
## 2                5.0              65.5        116.3                    0.0
## 3                0.0              39.4         59.0                    1.0
## 4                0.0              38.5         88.0                    0.0
## 5                0.5              58.1         81.6                    0.0
## 6                1.3              22.6         30.5                   15.7
## 7                0.5              74.0         51.3                   83.0
## 8                0.0              29.5         44.0                    0.0
## 9                0.0             100.3        120.1                    0.5
## 10               1.0               9.4         45.4                    0.5
## 11               1.7             100.8        117.1                   11.0
## 12               0.5               9.5         17.8                    2.0
## 13               0.0              14.3         66.3                    0.0
## 14               1.5              16.7         21.7                    3.0
## 15               5.7              25.1         10.2                   14.5
## 16               0.0              75.6        161.1                    0.0
## 17                NA                NA           NA                     NA
## 18                NA                NA           NA                     NA
## 19                NA                NA           NA                     NA
## 20                NA                NA           NA                     NA
## 21                NA                NA           NA                     NA
## 22                NA                NA           NA                     NA
## 23                NA                NA           NA                     NA
##    Londax.Bensulfuron.methyl Calliherbe.2.4.D Other.herbicides Total.Herbicide
## 1                        2.0              1.0              7.5            10.5
## 2                        0.0              0.0              0.0             0.0
## 3                        3.0              0.0              0.0             4.0
## 4                        0.0              0.0              0.0             0.0
## 5                        0.0              0.0              0.0             0.0
## 6                        8.2              1.0              5.4            30.3
## 7                       52.5              0.0              6.0           141.5
## 8                        6.0              0.5              0.8             7.3
## 9                        0.3              0.0             27.5            28.3
## 10                       4.0              0.0              2.0             6.5
## 11                       3.6              4.0             31.5            50.1
## 12                       2.7              0.0              1.8             6.5
## 13                       1.0              3.0              3.0             7.0
```

```
## 14                     1.0              0.0              4.3             8.3
## 15                    16.0              2.3              8.5            41.3
## 16                     3.0              0.0              2.0             5.0
## 17                      NA               NA               NA              NA
## 18                      NA               NA               NA              NA
## 19                      NA               NA               NA              NA
## 20                      NA               NA               NA              NA
## 21                      NA               NA               NA              NA
## 22                      NA               NA               NA              NA
## 23                      NA               NA               NA              NA
##       prevbase prevre1 prevre2   baseprev accessArea negkids_baseline
## 1  0.90163934    0.77    0.68 0.90163934   3369.060               12
## 2  0.77083333    0.67    0.89 0.77083333   5496.101               11
## 3  0.90322581    0.84    0.85 0.90322581   3031.277                3
## 4  0.95588235    0.82    0.56 0.95588235   1677.355                3
## 5  0.81707317    0.88    0.81 0.81707317   2536.556               15
## 6  0.77570094    0.49    0.88 0.77570094   3081.368               24
## 7  0.96875000    0.83    0.88 0.96875000   1608.964                3
## 8  0.98795181    0.85    0.98 0.98795181    519.440                1
## 9  0.95192308    0.96    0.88 0.95192308   6416.058                5
## 10 0.64864865    0.34    0.38 0.64864865   1498.642               39
## 11 0.86407767    0.85    0.66 0.86407767   2090.476               14
## 12 0.51260504    0.18    0.39 0.51260504    848.107               58
## 13 0.91176471    0.61    0.80 0.91176471   2820.774                9
## 14 0.69230769    0.38    0.35 0.69230769   1386.967               28
## 15 0.51724138    0.33    0.32 0.51724138   2691.446               42
## 16 0.97600000    0.99    0.98 0.97600000   6807.490                3
## 17 0.66666667      NA      NA 0.66666667   1921.000               10
## 18 0.90000000      NA      NA 0.90000000   4920.000                3
## 19 0.70000000      NA      NA 0.70000000   3775.000                9
## 20 0.50000000      NA      NA 0.50000000   1905.000               15
## 21 0.93333333      NA      NA 0.93333333   2135.000                2
## 22 0.09677419      NA      NA 0.09677419   2464.000               28
## 23 0.23333333      NA      NA 0.23333333   1565.000               23
##    negkids_baselineM logtotalfert logavgFloat       pred        lcb        ucb
## 1                105     4.970508    7.136077 0.08820030 0.06749065 0.11448456
## 2                 41     4.060443    7.857860 0.32361985 0.25884276 0.39594752
## 3                 30     3.668677    6.257648 0.14261369 0.10457000 0.19153749
## 4                  5     3.317816    6.437159 0.39477058 0.34563324 0.44613094
## 5                 82     4.018183    6.851460 0.32627326 0.28552816 0.36982311
## 6                 65     4.039536    7.349038 0.13845447 0.11270938 0.16896044
## 7                 65     5.043425    6.370517 0.37197845 0.32296709 0.42377204
## 8                 15     3.867026    4.412313 0.60607528 0.54824497 0.66107906
## 9                 95     4.478473    7.817215 0.09476289 0.06624315 0.13380180
## 10                69     2.778819    5.863631 0.24272890 0.19031175 0.30415987
## 11                85     4.739701    6.783404 0.30878306 0.26263606 0.35908972
## 12                89     2.901422    5.133089 0.40506334 0.36122669 0.45046822
## 13                26     2.933857    5.633253 0.58427946 0.52032778 0.64551360
## 14                59     2.803360    6.542083 0.30310572 0.26588056 0.34310682
## 15                75     3.781914    6.957859 0.37361960 0.33657409 0.41220911
## 16                97     3.653252    8.116178 0.13119010 0.09727678 0.17463924
## 17                28           NA          NA 0.15468523 0.11160752 0.21045112
## 18                29           NA          NA 0.11798009 0.08585446 0.16002243
## 19                29           NA          NA 0.21532472 0.18993709 0.24308745
```

```
## 20                    30            NA          NA 0.31754934 0.25384417 0.38890907
## 21                    30            NA          NA 0.30921082 0.27467242 0.34602038
## 22                    31            NA          NA 0.05041615 0.02998655 0.08356522
## 23                    30            NA          NA 0.23211770 0.18733708 0.28386358
```

```
#For paper
#ManMAIC<- aictab(cand.set = models_man, modnames = Modnames_man, sort = TRUE)
#ManMAIC<-round(ManMAIC[2:8], digits=2)
#write.table(ManMAIC, file = "ManMAIC2.txt", sep = ",", quote = FALSE, row.names = F)

#this gives us the predicted values of the model , along with the confidence bands

(est <- cbind(Estimate = coef(mansoni15), confint(mansoni15)))
```

```
## Waiting for profiling to be done...
```

```
##                         Estimate         2.5 %        97.5 %
## (Intercept)        -2.1654062056 -2.7654559710 -1.5897845851
## sqkmtotalcrop0.5km  5.3483165500  4.3242345972  6.4121112006
## sqkmwater0.5km      2.0605424053  0.5991081983  3.5213462053
## population          0.0003876931  0.0001251745  0.0006553047
## Sum.of.area_FM3_17 -0.0006114697 -0.0007098181 -0.0005167911
```

```
#To get incidence rate  (we exponentiate the coefficients.. and confidence intervals!):
exp(est)
```

```
##                       Estimate       2.5 %      97.5 %
## (Intercept)          0.1147033  0.06294739   0.2039695
## sqkmtotalcrop0.5km 210.2540476 75.50769694 609.1784223
## sqkmwater0.5km       7.8502267  1.82049456  33.8299399
## population           1.0003878  1.00012518   1.0006555
## Sum.of.area_FM3_17   0.9993887  0.99929043   0.9994833
```

```
##################################################
#PATH MODEL FOR HUMAN INFECTION - with NAs removed

#################################################
#HUMAN INFECTION PSEM:

#Examine for significant paths from ag->veg->snails->prev
#We must subset to single Fm per human infection round because only one round for 7 USF villages...
#Analysis uses only summer data : chose FM1,4,17

# Load dataset that excludes only response variables from site-time points that might be vulnerable to
FM1_4_17_2=read.csv("psem_data_1_13_fixed_exclusion.csv")
#Scale predictors:
FM1_4_17_2$prop=FM1_4_17_2$infected.kids/FM1_4_17_2$kids.tested
FM1_4_17_2$scalefloating=as.numeric(scale(FM1_4_17_2$Sum.of.floating))
FM1_4_17_2$scalesqkmtotalcrop0.5km=as.numeric(scale(FM1_4_17_2$sqkmtotalcrop0.5km))
FM1_4_17_2$scaletotalvegmass=as.numeric(scale(FM1_4_17_2$Sum.of.totalvegsum))
FM1_4_17_2$scaletotalbulinus=as.numeric(scale(FM1_4_17_2$Sum.of.totalbulinus))
FM1_4_17_2$scalelogtotalbulinus=as.numeric(scale(log(FM1_4_17_2$Sum.of.totalbulinus+1)))
```

```r
FM1_4_17_2$scalelogtotalbiom=as.numeric(scale(log(FM1_4_17_2$sum_of_biomphalaria+1)))
FM1_4_17_2$scaleCount.of.Microhabitat.Other=as.numeric(scale(FM1_4_17_2$Count.of.Microhabitat.Other))
FM1_4_17_2$logNpoint=log(FM1_4_17_2$Count.of.Microhabitat.Other+1)
FM1_4_17_2$scalelogCount.of.Microhabitat.Other=as.numeric(scale(FM1_4_17_2$logNpoint))
FM1_4_17_2$scalelogvegmass=as.numeric(scale(log(FM1_4_17_2$Sum.of.totalvegsum+1)))
FM1_4_17_2$scalevillagepop=as.numeric(scale(FM1_4_17_2$population))
FM1_4_17_2$scalesexratio=as.numeric(scale(FM1_4_17_2$sexratio))
FM1_4_17_2$scaleaverageclass=as.numeric(scale(FM1_4_17_2$avgclass))
FM1_4_17_2$scalehafert=as.numeric(scale(FM1_4_17_2$Totalfertilizer))

FM1_4_17_2b<-subset(FM1_4_17_2,is.na(FM1_4_17_2$kids.tested)==FALSE)
FM1_4_17_2b<-subset(FM1_4_17_2b,is.na(FM1_4_17_2b$scalefloating)==FALSE)
FM1_4_17_2b<-subset(FM1_4_17_2b,is.na(FM1_4_17_2b$scalelogCount.of.Microhabitat.Other)==FALSE)
#drop villagepop, total fert ha...

psem_finale = psem(
  lme(scalefloating ~ scalesqkmtotalcrop0.5km, random = ~1|villages,data = FM1_4_17_2b),
  lme(scalelogCount.of.Microhabitat.Other ~ scalefloating, random = ~ 1|villages, data = FM1_4_17_2b),
  lme(scalelogvegmass ~ scalelogCount.of.Microhabitat.Other, random = ~ 1|villages,data = FM1_4_17_2b),
  lme(scalelogtotalbulinus ~ scalelogvegmass, random = ~1|villages,data = FM1_4_17_2b),
  glmer(prop~scalelogtotalbulinus + scalefloating+scalesqkmtotalcrop0.5km+(1|villages),weights=kids.tes
  )

summary(psem_finale)
```

```
##   |                                                                              |

## boundary (singular) fit: see ?isSingular

##
## Structural Equation Model of psem_finale
##
## Call:
##   scalefloating ~ scalesqkmtotalcrop0.5km
##   scalelogCount.of.Microhabitat.Other ~ scalefloating
##   scalelogvegmass ~ scalelogCount.of.Microhabitat.Other
##   scalelogtotalbulinus ~ scalelogvegmass
##   prop ~ scalelogtotalbulinus + scalefloating + scalesqkmtotalcrop0.5km
##
##     AIC      BIC
##  59.308   80.218
##
## ---
## Tests of directed separation:
##
##                                                          Independ.Claim Test.Type
##   scalelogCount.of.Microhabitat.Other ~ scalesqkmtotalcrop0.5km + ...      coef
##                       scalelogvegmass ~ scalesqkmtotalcrop0.5km + ...      coef
##                  scalelogtotalbulinus ~ scalesqkmtotalcrop0.5km + ...      coef
##                                 scalelogvegmass ~ scalefloating + ...      coef
##                            scalelogtotalbulinus ~ scalefloating + ...      coef
##                        prop ~ scalelogCount.of.Microhabitat.Other + ...      coef
##      scalelogtotalbulinus ~ scalelogCount.of.Microhabitat.Other + ...      coef
```

```
##                                              scalelogvegmass ~ prop + ...      coef
##   DF Crit.Value P.Value
##   13    -0.6824  0.5070
##   13    -1.5321  0.1495
##   13    -1.8960  0.0804
##    3     1.7416  0.1799
##    3     0.6488  0.5627
##   20     0.9847  0.3248
##    2    -0.1257  0.9114
##    1     0.0700  0.9555
##
## Global goodness-of-fit:
##
##   Fisher's C = 17.308 with P-value = 0.366 and on 16 degrees of freedom
##
## ---
## Coefficients:
##
##                              Response                           Predictor
##                         scalefloating             scalesqkmtotalcrop0.5km
##   scalelogCount.of.Microhabitat.Other                       scalefloating
##                       scalelogvegmass scalelogCount.of.Microhabitat.Other
##                  scalelogtotalbulinus                     scalelogvegmass
##                                  prop                scalelogtotalbulinus
##                                  prop                       scalefloating
##                                  prop             scalesqkmtotalcrop0.5km
##   Estimate Std.Error DF Crit.Value P.Value Std.Estimate
##     0.5925    0.2196 13     2.6988  0.0182       0.5794 *
##     0.3904    0.1897  4     2.0579  0.1087       0.4336
##     0.4132    0.2000  4     2.0656  0.1078        0.436
##     0.7198    0.1781  4     4.0416  0.0156       0.6648 *
##     0.3775    0.2286 20     1.6511  0.0987            -
##     0.5099    0.2538 20     2.0087  0.0446            - *
##     0.7845    0.3379 20     2.3217  0.0203            - *
##
##   Signif. codes:  0 '***' 0.001 '**' 0.01 '*' 0.05
##
## ---
## Individual R-squared:
##
##                              Response method Marginal Conditional
##                         scalefloating   none     0.34        0.83
##   scalelogCount.of.Microhabitat.Other   none     0.16        0.93
##                       scalelogvegmass   none     0.17        0.91
##                  scalelogtotalbulinus   none     0.47        0.80
##                                  prop  delta     0.21        0.35
```

```
#switch to one-tailed p-value given strog apriori hypo supported by literature
#then  address individual-level traits in child-level
```

```
#Path model for S. mansoni

FM1_4_17_2b$prop_m=FM1_4_17_2b$infected.kids.mansoni/FM1_4_17_2b$kids.tested
```

```r
#try initial global
psem_mansoni <- psem(
  lme(scalefloating ~ scalesqkmtotalcrop0.5km+scalehafert, random = ~ 1|villages,data = FM1_4_17_2b),
  lme(scalelogCount.of.Microhabitat.Other ~ scalefloating, random = ~ 1|villages,data = FM1_4_17_2b),
  lme(scalelogvegmass ~ scalelogCount.of.Microhabitat.Other+scalehafert, random = ~ 1|villages,data = Fl
  lme(scalelogtotalbiom ~ scalelogvegmass, random = ~1|villages,data = FM1_4_17_2b),
  glmer(prop_m~scalevillagepop+scalelogtotalbiom + scalefloating+scalesqkmtotalcrop0.5km+(1|villages),w
  )
```

```
## Warning in glmer(prop_m ~ scalevillagepop + scalelogtotalbiom + scalefloating
## + : calling glmer() with family=gaussian (identity link) as a shortcut to lmer()
## is deprecated; please call lmer() directly
```

```r
summary(psem_mansoni)
```

```
##   |                                                                               |

## Registered S3 methods overwritten by 'broom':
##   method            from
##   tidy.glht         jtools
##   tidy.summary.glht jtools

##   |                                                                               |==================

##
## Structural Equation Model of psem_mansoni
##
## Call:
##   scalefloating ~ scalesqkmtotalcrop0.5km + scalehafert
##   scalelogCount.of.Microhabitat.Other ~ scalefloating
##   scalelogvegmass ~ scalelogCount.of.Microhabitat.Other + scalehafert
##   scalelogtotalbiom ~ scalelogvegmass
##   prop_m ~ scalevillagepop + scalelogtotalbiom + scalefloating + scalesqkmtotalcrop0.5km
##
##     AIC      BIC
##  391.192   416.085
##
## ---
## Tests of directed separation:
##
##                                                        Independ.Claim Test.Type
##   scalelogCount.of.Microhabitat.Other ~ scalesqkmtotalcrop0.5km + ...      coef
##                     scalelogvegmass ~ scalesqkmtotalcrop0.5km + ...      coef
##                   scalelogtotalbiom ~ scalesqkmtotalcrop0.5km + ...      coef
##             scalelogCount.of.Microhabitat.Other ~ scalehafert + ...      coef
##                                          prop_m ~ scalehafert + ...      coef
##                               scalelogtotalbiom ~ scalehafert + ...      coef
##                               scalefloating ~ scalevillagepop + ...      coef
##         scalelogCount.of.Microhabitat.Other ~ scalevillagepop + ...      coef
##                             scalelogvegmass ~ scalevillagepop + ...      coef
##                           scalelogtotalbiom ~ scalevillagepop + ...      coef
##                               scalelogvegmass ~ scalefloating + ...      coef
```

```
##                                scalelogtotalbiom ~ scalefloating + ...      coef
##                      prop_m ~ scalelogCount.of.Microhabitat.Other + ...      coef
##          scalelogtotalbiom ~ scalelogCount.of.Microhabitat.Other + ...      coef
##                                         scalelogvegmass ~ prop_m + ...      coef
##         DF Crit.Value P.Value
##      13.00    -0.6824  0.5070
##      12.00    -1.2894  0.2215
##      13.00    -2.1425  0.0517
##      13.00    -0.2214  0.8282
##   18731.71     0.0093  0.9232
##      13.00    -0.4643  0.6501
##      11.00     1.2600  0.2338
##      13.00     1.4797  0.1628
##      12.00     0.6636  0.5195
##      13.00     0.5583  0.5861
##       3.00     1.1573  0.3309
##       3.00     0.1957  0.8574
##   15526.85   312.0575  0.0000 ***
##       2.00    -0.1682  0.8819
##       1.00    -2.2712  0.2640
##
## Global goodness-of-fit:
##
##   Fisher's C = 341.192 with P-value = 0 and on 30 degrees of freedom
##
## ---
## Coefficients:
##
##                               Response                           Predictor
##                          scalefloating            scalesqkmtotalcrop0.5km
##                          scalefloating                         scalehafert
##   scalelogCount.of.Microhabitat.Other                       scalefloating
##                        scalelogvegmass scalelogCount.of.Microhabitat.Other
##                        scalelogvegmass                         scalehafert
##                      scalelogtotalbiom                     scalelogvegmass
##                                 prop_m                     scalevillagepop
##                                 prop_m                   scalelogtotalbiom
##                                 prop_m                       scalefloating
##                                 prop_m             scalesqkmtotalcrop0.5km
##   Estimate Std.Error       DF Crit.Value P.Value Std.Estimate
##     0.6392    0.2217    12.00     2.8833  0.0137       0.6251 *
##     0.2523    0.2331    12.00     1.0825  0.3003       0.2281
##     0.3904    0.1897     4.00     2.0579  0.1087       0.4336
##     0.4346    0.1872     4.00     2.3215  0.0810       0.4586
##     0.4123    0.2210    13.00     1.8657  0.0848       0.4368
##     0.3311    0.1792     4.00     1.8473  0.1384       0.3396
##     0.0181    0.0729 27331.69     0.0616  0.8040       0.0716
##    -0.1032    0.0652 19796.90     2.4845  0.1150      -0.4746
##    -0.0799    0.0546 31947.11     2.1236  0.1451      -0.4414
##    -0.0464    0.0761 22309.26     0.3690  0.5435      -0.2510
##
##   Signif. codes:  0 '***' 0.001 '**' 0.01 '*' 0.05
##
## ---
```

```
## Individual R-squared:
##
##                               Response method Marginal Conditional
##                          scalefloating   none     0.38        0.84
##   scalelogCount.of.Microhabitat.Other   none     0.16        0.93
##                        scalelogvegmass   none     0.35        0.92
##                      scalelogtotalbiom   none     0.14        0.92
##                                 prop_m   none     0.01        0.02
```

```
#drop nonsig paths from each response:
psem_mansoni2 = psem(
  lme(scalefloating ~ scalesqkmtotalcrop0.5km, random = ~ 1|villages,na.action=na.omit,data = FM1_4_17_2
  lme(scalelogvegmass ~ scalelogCount.of.Microhabitat.Other, random = ~ 1|villages,na.action=na.omit,dat
  lme(scalelogtotalbiom ~ scalelogvegmass, random = ~1|villages,na.action=na.omit,data = FM1_4_17_2b),
  glmer(prop_m~scalevillagepop + scalefloating+scalesqkmtotalcrop0.5km+(1|villages),weights=kids.tested
summary(psem_mansoni2)
```

```
##   |                                                                              |

## boundary (singular) fit: see ?isSingular

##
## Structural Equation Model of psem_mansoni2
##
## Call:
##   scalefloating ~ scalesqkmtotalcrop0.5km
##   scalelogvegmass ~ scalelogCount.of.Microhabitat.Other
##   scalelogtotalbiom ~ scalelogvegmass
##   prop_m ~ scalevillagepop + scalefloating + scalesqkmtotalcrop0.5km
##
##     AIC      BIC
##  75.659   92.586
##
## ---
## Tests of directed separation:
##
##                                                     Independ.Claim Test.Type DF
##                 scalelogvegmass ~ scalesqkmtotalcrop0.5km + ...      coef 13
##               scalelogtotalbiom ~ scalesqkmtotalcrop0.5km + ...      coef 13
##       scalefloating ~ scalelogCount.of.Microhabitat.Other + ...      coef  4
##   scalelogtotalbiom ~ scalelogCount.of.Microhabitat.Other + ...      coef  3
##              prop_m ~ scalelogCount.of.Microhabitat.Other + ...      coef 20
##                           scalefloating ~ scalevillagepop + ...      coef 12
##                         scalelogvegmass ~ scalevillagepop + ...      coef 13
##                       scalelogtotalbiom ~ scalevillagepop + ...      coef 13
##                           scalelogvegmass ~ scalefloating + ...      coef  3
##                         scalelogtotalbiom ~ scalefloating + ...      coef  3
##                                prop_m ~ scalelogvegmass + ...      coef 20
##                              prop_m ~ scalelogtotalbiom + ...      coef 20
##   Crit.Value P.Value
##      -1.5321  0.1495
##      -2.1425  0.0517
##       1.9919  0.1172
```

```
##      -0.1883  0.8627
##       3.0092  0.0026 **
##       1.3509  0.2017
##       0.9029  0.3830
##       0.5583  0.5861
##       1.7416  0.1799
##       0.1675  0.8777
##      -0.7350  0.4623
##      -1.5020  0.1331
##
## Global goodness-of-fit:
##
##   Fisher's C = 41.659 with P-value = 0.014 and on 24 degrees of freedom
##
## ---
## Coefficients:
##
##            Response                           Predictor Estimate Std.Error DF
##       scalefloating             scalesqkmtotalcrop0.5km   0.5925    0.2196 13
##     scalelogvegmass scalelogCount.of.Microhabitat.Other   0.4132    0.2000  4
##   scalelogtotalbiom                     scalelogvegmass   0.3311    0.1792  4
##              prop_m                     scalevillagepop   0.7252    0.4643 20
##              prop_m                       scalefloating  -1.0650    0.2802 20
##              prop_m             scalesqkmtotalcrop0.5km   0.4276    0.4084 20
##   Crit.Value P.Value Std.Estimate
##       2.6988  0.0182       0.5794   *
##       2.0656  0.1078        0.436
##       1.8473  0.1384       0.3396
##       1.5619  0.1183            -
##      -3.8009  0.0001            - ***
##       1.0470  0.2951            -
##
##   Signif. codes:  0 '***' 0.001 '**' 0.01 '*' 0.05
##
## ---
## Individual R-squared:
##
##            Response method Marginal Conditional
##       scalefloating   none     0.34        0.83
##     scalelogvegmass   none     0.17        0.91
##   scalelogtotalbiom   none     0.14        0.92
##              prop_m  delta     0.05        0.12
```

```
#Why is the S. mansoni prop negative to floating? I think the massive spike at Gankette is big part - t
#area of open water that we did not account for (new water access area where we would miss vegetation).

FM1_4_17_2b$villages
```

```
##  [1] "Diokhor"     "Foss"        "Guidick"     "Lampsar"     "MakaDiama"
##  [6] "Malla"       "Malla Tack"  "Mbakhana"    "Mbane"       "Mbarigot"
## [11] "NdiolMaure"  "Syer"        "Diokhor"     "Diokhoul"    "Malla Tack"
## [16] "Mbakhana"    "MerinaGewel" "Ndiawdoune"  "NdiolMaure"  "Syer"
```

```r
#lets drop gankette, row 11:

nogankette=FM1_4_17_2b[-c(11), ]
nogankette$villages
```

```
##  [1] "Diokhor"     "Foss"        "Guidick"     "Lampsar"     "MakaDiama"
##  [6] "Malla"       "Malla Tack"  "Mbakhana"    "Mbane"       "Mbarigot"
## [11] "Syer"        "Diokhor"     "Diokhoul"    "Malla Tack"  "Mbakhana"
## [16] "MerinaGewel" "Ndiawdoune"  "NdiolMaure"  "Syer"
```

```r
#try initial global
psem_mansoni3 = psem(
  lme(scalefloating ~ scalesqkmtotalcrop0.5km+scalehafert, random = ~ 1|villages,na.action=na.omit,data
  lme(scalelogCount.of.Microhabitat.Other ~ scalefloating, random = ~ 1|villages,na.action=na.omit,data
  lme(scalelogvegmass ~ scalelogCount.of.Microhabitat.Other+scalehafert, random = ~ 1|villages,na.actio
  lme(scalelogtotalbiom ~ scalelogvegmass, random = ~1|villages,na.action=na.omit,data = nogankette),
  glmer(prop_m~scalevillagepop+scalelogtotalbiom + scalefloating+scalesqkmtotalcrop0.5km+(1|villages),w
```

```
## Warning in checkConv(attr(opt, "derivs"), opt$par, ctrl = control$checkConv, :
## Model failed to converge with max|grad| = 0.0198947 (tol = 0.002, component 1)
```

```r
summary(psem_mansoni3)
```

```
##   |                                                                              |

## Warning in pt(-abs(tVal), fDF): NaNs produced

##   |                                                                              |==================

## boundary (singular) fit: see ?isSingular

##
## Structural Equation Model of psem_mansoni3
##
## Call:
##   scalefloating ~ scalesqkmtotalcrop0.5km + scalehafert
##   scalelogCount.of.Microhabitat.Other ~ scalefloating
##   scalelogvegmass ~ scalelogCount.of.Microhabitat.Other + scalehafert
##   scalelogtotalbiom ~ scalelogvegmass
##   prop_m ~ scalevillagepop + scalelogtotalbiom + scalefloating + scalesqkmtotalcrop0.5km
##
##     AIC      BIC
##  NaN   NaN
##
## ---
## Tests of directed separation:
##
##                                                          Independ.Claim Test.Type
##   scalelogCount.of.Microhabitat.Other ~ scalesqkmtotalcrop0.5km + ...      coef
##                       scalelogvegmass ~ scalesqkmtotalcrop0.5km + ...      coef
```

```
##                    scalelogtotalbiom ~ scalesqkmtotalcrop0.5km + ...      coef
##              scalelogCount.of.Microhabitat.Other ~ scalehafert + ...      coef
##                                        prop_m ~ scalehafert + ...      coef
##                             scalelogtotalbiom ~ scalehafert + ...      coef
##                             scalefloating ~ scalevillagepop + ...      coef
##          scalelogCount.of.Microhabitat.Other ~ scalevillagepop + ...      coef
##                           scalelogvegmass ~ scalevillagepop + ...      coef
##                         scalelogtotalbiom ~ scalevillagepop + ...      coef
##                             scalelogvegmass ~ scalefloating + ...      coef
##                           scalelogtotalbiom ~ scalefloating + ...      coef
##                  prop_m ~ scalelogCount.of.Microhabitat.Other + ...      coef
##        scalelogtotalbiom ~ scalelogCount.of.Microhabitat.Other + ...      coef
##                                   scalelogvegmass ~ prop_m + ...      coef
##   DF Crit.Value P.Value
##   13    -0.6934  0.5002
##   12    -0.9553  0.3583
##   13    -1.8470  0.0876
##   13    -0.2167  0.8318
##   19    37.7593  0.0000  ***
##   13    -0.7083  0.4913
##   11     1.2351  0.2425
##   13     1.4760  0.1638
##   12     1.2355  0.2403
##   13     0.2871  0.7785
##    2     1.3502  0.3095
##    2    -0.1440  0.8987
##   19   606.6598  0.0000  ***
##    1    -0.1676  0.8943
##    0    -2.4893     NaN <NA>
##
## Global goodness-of-fit:
##
##   Fisher's C = NaN with P-value = NaN and on 30 degrees of freedom
##
## ---
## Coefficients:
##
##                               Response                       Predictor
##                          scalefloating           scalesqkmtotalcrop0.5km
##                          scalefloating                       scalehafert
##   scalelogCount.of.Microhabitat.Other                     scalefloating
##                        scalelogvegmass scalelogCount.of.Microhabitat.Other
##                        scalelogvegmass                       scalehafert
##                      scalelogtotalbiom                   scalelogvegmass
##                                 prop_m                   scalevillagepop
##                                 prop_m                 scalelogtotalbiom
##                                 prop_m                     scalefloating
##                                 prop_m           scalesqkmtotalcrop0.5km
##   Estimate Std.Error DF Crit.Value P.Value Std.Estimate
##     0.6490    0.2217 12     2.9277  0.0127       0.6313   *
##     0.2490    0.2331 12     1.0684  0.3064       0.2247
##     0.3901    0.2016  3     1.9350  0.1484       0.4484
##    -0.0026    0.0818  3    -0.0313  0.9770      -0.0026
##     0.4242    0.2715 13     1.5621  0.1423       0.4489
```

```
##     0.4369    0.1989  3     2.1967  0.1155       0.4579
##     0.7220    0.0036 19   202.8701  0.0000            - ***
##    -0.6815    0.0036 19  -191.5061  0.0000            - ***
##    -0.8399    0.0036 19  -236.1206  0.0000            - ***
##    -0.0718    0.0036 19   -20.1730  0.0000            - ***
##
##   Signif. codes:  0 '***' 0.001 '**' 0.01 '*' 0.05
##
## ---
## Individual R-squared:
##
##                              Response method Marginal Conditional
##                         scalefloating   none     0.40        0.82
##   scalelogCount.of.Microhabitat.Other   none     0.16        0.92
##                       scalelogvegmass   none     0.16        1.00
##                     scalelogtotalbiom   none     0.22        0.96
##                                prop_m  delta     0.05        0.11
```

```
#floating is still a sig - predictor of S. mansoni prev.  ????
#note this sig + path in the d-sep tests:prop_m  ~  scalelogCount.of.Microhabitat.Other
psem_mansoni3.1 = psem(
  lme(scalefloating ~ scalesqkmtotalcrop0.5km+scalehafert, random = ~ 1|villages,na.action=na.omit,data
  lme(scalelogCount.of.Microhabitat.Other ~ scalefloating, random = ~ 1|villages,na.action=na.omit,data
  lme(scalelogvegmass ~ scalelogCount.of.Microhabitat.Other+scalehafert, random = ~ 1|villages,na.actio
  lme(scalelogtotalbiom ~ scalelogvegmass, random = ~1|villages,na.action=na.omit,data = nogankette),
  glmer(prop_m~scalevillagepop+scalelogtotalbiom + scalefloating+scalesqkmtotalcrop0.5km+(1|villages),f
  prop_m  %~~%   scalelogCount.of.Microhabitat.Other)
```

```
## Warning in checkConv(attr(opt, "derivs"), opt$par, ctrl = control$checkConv, :
## Model failed to converge with max|grad| = 0.0198947 (tol = 0.002, component 1)
```

```
summary(psem_mansoni3.1)
```

```
##   |                                                                              |

## Warning in pt(-abs(tVal), fDF): NaNs produced

##   |                                                                              |===================

## boundary (singular) fit: see ?isSingular

##
## Structural Equation Model of psem_mansoni3.1
##
## Call:
##   scalefloating ~ scalesqkmtotalcrop0.5km + scalehafert
##   scalelogCount.of.Microhabitat.Other ~ scalefloating
##   scalelogvegmass ~ scalelogCount.of.Microhabitat.Other + scalehafert
##   scalelogtotalbiom ~ scalelogvegmass
##   prop_m ~ scalevillagepop + scalelogtotalbiom + scalefloating + scalesqkmtotalcrop0.5km
##   prop_m ~~ scalelogCount.of.Microhabitat.Other
```

```
##
##     AIC      BIC
##  NaN   NaN
##
## ---
## Tests of directed separation:
##
##                                                          Independ.Claim Test.Type
##   scalelogCount.of.Microhabitat.Other ~ scalesqkmtotalcrop0.5km + ...      coef
##                       scalelogvegmass ~ scalesqkmtotalcrop0.5km + ...      coef
##                     scalelogtotalbiom ~ scalesqkmtotalcrop0.5km + ...      coef
##               scalelogCount.of.Microhabitat.Other ~ scalehafert + ...      coef
##                                             prop_m ~ scalehafert + ...      coef
##                                 scalelogtotalbiom ~ scalehafert + ...      coef
##                                 scalefloating ~ scalevillagepop + ...      coef
##           scalelogCount.of.Microhabitat.Other ~ scalevillagepop + ...      coef
##                               scalelogvegmass ~ scalevillagepop + ...      coef
##                             scalelogtotalbiom ~ scalevillagepop + ...      coef
##                                 scalelogvegmass ~ scalefloating + ...      coef
##                               scalelogtotalbiom ~ scalefloating + ...      coef
##         scalelogtotalbiom ~ scalelogCount.of.Microhabitat.Other + ...      coef
##                                        scalelogvegmass ~ prop_m + ...      coef
##   DF Crit.Value P.Value
##   13    -0.6934  0.5002
##   12    -0.9553  0.3583
##   13    -1.8470  0.0876
##   13    -0.2167  0.8318
##   19    37.7593  0.0000  ***
##   13    -0.7083  0.4913
##   11     1.2351  0.2425
##   13     1.4760  0.1638
##   12     1.2355  0.2403
##   13     0.2871  0.7785
##    2     1.3502  0.3095
##    2    -0.1440  0.8987
##    1    -0.1676  0.8943
##    0    -2.4893     NaN <NA>
##
## Global goodness-of-fit:
##
##   Fisher's C = NaN with P-value = NaN and on 28 degrees of freedom
##
## ---
## Coefficients:
##
##                              Response                          Predictor
##                         scalefloating            scalesqkmtotalcrop0.5km
##                         scalefloating                        scalehafert
##   scalelogCount.of.Microhabitat.Other                      scalefloating
##                       scalelogvegmass scalelogCount.of.Microhabitat.Other
##                       scalelogvegmass                        scalehafert
##                     scalelogtotalbiom                    scalelogvegmass
##                                prop_m                    scalevillagepop
##                                prop_m                  scalelogtotalbiom
```

```
##                                prop_m                        scalefloating
##                                prop_m              scalesqkmtotalcrop0.5km
##                              ~~prop_m ~~scalelogCount.of.Microhabitat.Other
##   Estimate Std.Error DF Crit.Value P.Value Std.Estimate
##     0.6490    0.2217 12     2.9277  0.0127       0.6313   *
##     0.2490    0.2331 12     1.0684  0.3064       0.2247
##     0.3901    0.2016  3     1.9350  0.1484       0.4484
##    -0.0026    0.0818  3    -0.0313  0.9770      -0.0026
##     0.4242    0.2715 13     1.5621  0.1423       0.4489
##     0.4369    0.1989  3     2.1967  0.1155       0.4579
##     0.7220    0.0036 19   202.8701  0.0000            - ***
##    -0.6815    0.0036 19  -191.5061  0.0000            - ***
##    -0.8399    0.0036 19  -236.1206  0.0000            - ***
##    -0.0718    0.0036 19   -20.1730  0.0000            - ***
##     0.7598         - 19     4.6740  0.0001       0.7598 ***
##
##   Signif. codes:  0 '***' 0.001 '**' 0.01 '*' 0.05
##
## ---
## Individual R-squared:
##
##                              Response method Marginal Conditional
##                         scalefloating   none     0.40        0.82
##   scalelogCount.of.Microhabitat.Other   none     0.16        0.92
##                       scalelogvegmass   none     0.16        1.00
##                     scalelogtotalbiom   none     0.22        0.96
##                                prop_m  delta     0.05        0.11
```

```
#Adding correlated error does not change that floating is sig - predictor of prop_m
#what if instead we add the pathway to the model?
```

```
###################
#Child-level model:
###################

#start with same human data as in the path model, with snails and veg corresponding to FM1 and FM4 for 
dataset3s=read.csv("human_data_per_child_2017_2018.csv")
names(dataset3s)
```

```
##  [1] "Village"               "idcode"                "Intervention.type"
##  [4] "manipulation"          "net_y_n"               "manipulation_numeric"
##  [7] "exclude_timepoint"     "exclude_timepoint2"    "Intervention.time"
## [10] "Sampling.bout"         "Sh.single.test"        "Sm.single.test"
## [13] "ID"                    "School"                "Class"
## [16] "ShW"                   "SmW"                   "LogShW"
## [19] "LogSmW"                "Sh"                    "Sm"
## [22] "coinfections"          "Tx_early"              "year"
## [25] "sex"                   "sex2"                  "Pop"
## [28] "lake"                  "cluster"               "neverinfected"
## [31] "vegmass"               "totalbulinus"          "floating"
## [34] "sqkmtotalcrop0.5km"    "accessarea"
```

```r
#Scale predictors:
dataset3s$scalefloating=scale(dataset3s$floating)
dataset3s$scalesqkmtotalcrop0.5km=scale(dataset3s$sqkmtotalcrop0.5km)
dataset3s$scaletotalvegmass=scale(dataset3s$vegmass)
dataset3s$scaletotalbulinus=scale(dataset3s$totalbulinus)
dataset3s$scalelogtotalbulinus=scale(log(dataset3s$totalbulinus+1))
dataset3s$scalepopulation=scale(log(dataset3s$Pop))
dataset3s$scaleaccessarea=scale(dataset3s$accessarea)
dataset3s$sex2=factor(dataset3s$sex2)
dataset3s$class=factor(dataset3s$Class)
#dataset3s$manipulation=factor(dataset3s$manipulation)

#need to exclude only the timepoints with prawns and veg removal:
dataset5s=dataset3s[dataset3s$exclude_timepoint2 == "0", ]
dataset5s$exclude_timepoint <- dataset5s$exclude_timepoint[ , drop=TRUE]
#Scale predictors:
dataset5s$scalefloating=scale(dataset5s$floating)
dataset5s$scalesqkmtotalcrop0.5km=scale(dataset5s$sqkmtotalcrop0.5km)
dataset5s$scaletotalvegmass=scale(dataset5s$vegmass)
dataset5s$scaletotalbulinus=scale(dataset5s$totalbulinus)
dataset5s$scalelogtotalbulinus=scale(log(dataset5s$totalbulinus+1))
dataset5s$scalepopulation=scale(log(dataset5s$Pop))
dataset5s$scaleaccessarea=scale(dataset5s$accessarea)
dataset5s$sex2=factor(dataset5s$sex2)
dataset5s$class=factor(dataset5s$Class)
#Remove Na's
dataset6s<-subset(dataset5s,is.na(dataset5s$scaleaccessarea)==FALSE)

#Global model:

childmodel_lake11=glmer(Sh~ scaleaccessarea+sex2+Class+scalepopulation+scalefloating+scalesqkmtotalcrop(
summary(childmodel_lake11)
```

```
## Generalized linear mixed model fit by maximum likelihood (Laplace
##   Approximation) [glmerMod]
##  Family: binomial  ( logit )
## Formula:
## Sh ~ scaleaccessarea + sex2 + Class + scalepopulation + scalefloating +
##     scalesqkmtotalcrop0.5km + scalelogtotalbulinus + (1 | School/ID)
##    Data: dataset6s
## Control: glmerControl(optimizer = "bobyqa", optCtrl = list(maxfun = 1e+05))
##
##      AIC      BIC   logLik deviance df.resid
##   1929.7   1985.5   -954.9   1909.7     1939
##
## Scaled residuals:
##     Min      1Q  Median      3Q     Max
## -8.3144 -0.6429  0.2367  0.5669  2.3735
##
## Random effects:
##  Groups    Name        Variance Std.Dev.
##  ID:School (Intercept) 0.2460   0.4959
##  School    (Intercept) 0.9655   0.9826
```

```
## Number of obs: 1949, groups:  ID:School, 1504; School, 22
##
## Fixed effects:
##                          Estimate Std. Error z value Pr(>|z|)
## (Intercept)               0.93447    0.28218   3.312 0.000927 ***
## scaleaccessarea           0.13717    0.48052   0.285 0.775287
## sex21                     0.36463    0.12023   3.033 0.002422 **
## Class                    -0.02713    0.06609  -0.411 0.681393
## scalepopulation          -0.42953    0.21303  -2.016 0.043771 *
## scalefloating             0.57334    0.38290   1.497 0.134303
## scalesqkmtotalcrop0.5km  1.01248    0.35748   2.832 0.004622 **
## scalelogtotalbulinus      0.48730    0.21432   2.274 0.022985 *
## ---
## Signif. codes:  0 '***' 0.001 '**' 0.01 '*' 0.05 '.' 0.1 ' ' 1
##
## Correlation of Fixed Effects:
##             (Intr) sclccs sex21  Class  sclppl sclflt scl0.5
## scaleaccssr  0.051
## sex21       -0.164 -0.005
## Class       -0.496 -0.033 -0.063
## scalepopltn  0.039  0.355 -0.033  0.043
## scaleflotng  0.122 -0.690  0.017  0.026 -0.387
## sclsqkmt0.5 -0.010 -0.356  0.013 -0.019 -0.045 -0.106
## scllgttlbln -0.031  0.101  0.000 -0.019 -0.157 -0.224  0.385
```

```
#drop access area:
childmodel_lake12=glmer(Sh~ sex2+Class+scalepopulation+scalefloating+scalesqkmtotalcrop0.5km+scalelogto
summary(childmodel_lake12)
```

```
## Generalized linear mixed model fit by maximum likelihood (Laplace
##   Approximation) [glmerMod]
##  Family: binomial  ( logit )
## Formula:
## Sh ~ sex2 + Class + scalepopulation + scalefloating + scalesqkmtotalcrop0.5km +
##     scalelogtotalbulinus + (1 | School/ID)
##    Data: dataset6s
## Control: glmerControl(optimizer = "bobyqa", optCtrl = list(maxfun = 1e+05))
##
##      AIC      BIC   logLik deviance df.resid
##   1927.8   1978.0   -954.9   1909.8     1940
##
## Scaled residuals:
##     Min      1Q  Median      3Q     Max
## -8.3802 -0.6436  0.2311  0.5667  2.3926
##
## Random effects:
##  Groups    Name        Variance Std.Dev.
##  ID:School (Intercept) 0.2449   0.4949
##  School    (Intercept) 0.9690   0.9844
## Number of obs: 1949, groups:  ID:School, 1504; School, 22
##
## Fixed effects:
##                          Estimate Std. Error z value Pr(>|z|)
## (Intercept)               0.93062    0.28216   3.298 0.000973 ***
```

```
## sex21                     0.36485    0.12020   3.035 0.002402 **
## Class                    -0.02650    0.06605  -0.401 0.688232
## scalepopulation          -0.45167    0.19942  -2.265 0.023516 *
## scalefloating             0.65001    0.27885   2.331 0.019752 *
## scalesqkmtotalcrop0.5km  1.04896    0.33510   3.130 0.001746 **
## scalelogtotalbulinus      0.48241    0.21348   2.260 0.023835 *
## ---
## Signif. codes:  0 '***' 0.001 '**' 0.01 '*' 0.05 '.' 0.1 ' ' 1
##
## Correlation of Fixed Effects:
##             (Intr) sex21  Class  sclppl sclflt scl0.5
## sex21       -0.164
## Class       -0.495 -0.063
## scalepopltn  0.022 -0.033  0.058
## scaleflotng  0.219  0.018  0.005 -0.211
## sclsqkmt0.5  0.007  0.012 -0.033  0.095 -0.524
## scllgttlbln -0.037  0.000 -0.016 -0.206 -0.217  0.453
```

```
#drop non-significant terms to final model:
childmodel_lake13=glmer(Sh~ sex2+scalepopulation+scalefloating+scalesqkmtotalcrop0.5km+scalelogtotalbuli
summary(childmodel_lake13)
```

```
## Generalized linear mixed model fit by maximum likelihood (Laplace
##   Approximation) [glmerMod]
##  Family: binomial  ( logit )
## Formula:
## Sh ~ sex2 + scalepopulation + scalefloating + scalesqkmtotalcrop0.5km +
##     scalelogtotalbulinus + (1 | School/ID)
##    Data: dataset6s
## Control: glmerControl(optimizer = "bobyqa", optCtrl = list(maxfun = 1e+05))
##
##      AIC      BIC   logLik deviance df.resid
##   1925.9   1970.5   -955.0   1909.9     1941
##
## Scaled residuals:
##     Min      1Q  Median      3Q     Max
## -8.5858 -0.6398  0.2268  0.5662  2.4554
##
## Random effects:
##  Groups    Name        Variance Std.Dev.
##  ID:School (Intercept) 0.2401   0.490
##  School    (Intercept) 0.9702   0.985
## Number of obs: 1949, groups:  ID:School, 1504; School, 22
##
## Fixed effects:
##                          Estimate Std. Error z value Pr(>|z|)
## (Intercept)                0.8748     0.2453   3.566 0.000362 ***
## sex21                      0.3619     0.1199   3.019 0.002534 **
## scalepopulation           -0.4471     0.1992  -2.245 0.024773 *
## scalefloating              0.6506     0.2788   2.334 0.019608 *
## scalesqkmtotalcrop0.5km   1.0448     0.3349   3.120 0.001809 **
## scalelogtotalbulinus      0.4811     0.2134   2.255 0.024136 *
## ---
## Signif. codes:  0 '***' 0.001 '**' 0.01 '*' 0.05 '.' 0.1 ' ' 1
```

```
##
## Correlation of Fixed Effects:
##             (Intr) sex21  sclppl sclflt scl0.5
## sex21       -0.225
## scalepopltn  0.059 -0.029
## scaleflotng  0.254  0.019 -0.211
## sclsqkmt0.5 -0.010  0.010  0.097 -0.524
## scllgttlbln -0.051 -0.001 -0.206 -0.215  0.452
```

```
#try for S. mansoni per reviewer:
childmodel_Sm=glmer(Sm~ scaleaccessarea+sex2+Class+scalepopulation+scalefloating+scalesqkmtotalcrop0.5km
summary(childmodel_Sm)
```

```
## Generalized linear mixed model fit by maximum likelihood (Laplace
##   Approximation) [glmerMod]
##  Family: binomial  ( logit )
## Formula:
## Sm ~ scaleaccessarea + sex2 + Class + scalepopulation + scalefloating +
##     scalesqkmtotalcrop0.5km + scalelogtotalbulinus + (1 | School/ID)
##    Data: dataset6s
## Control: glmerControl(optimizer = "bobyqa", optCtrl = list(maxfun = 1e+05))
##
##      AIC      BIC   logLik deviance df.resid
##   1392.5   1447.0   -686.2   1372.5     1718
##
## Scaled residuals:
##     Min      1Q  Median      3Q     Max
## -1.1002 -0.3830 -0.2843 -0.1857  4.9898
##
## Random effects:
##  Groups    Name        Variance Std.Dev.
##  ID:School (Intercept) 0.3992   0.6318
##  School    (Intercept) 1.4778   1.2156
## Number of obs: 1728, groups:  ID:School, 1289; School, 15
##
## Fixed effects:
##                         Estimate Std. Error z value Pr(>|z|)
## (Intercept)             -2.16100    0.38996  -5.542    3e-08 ***
## scaleaccessarea         -0.41597    0.61380  -0.678  0.49796
## sex21                    0.53420    0.14844   3.599  0.00032 ***
## Class                   -0.06578    0.07496  -0.878  0.38019
## scalepopulation          0.08838    0.33160   0.267  0.78983
## scalefloating            0.22281    0.42003   0.530  0.59579
## scalesqkmtotalcrop0.5km  0.18062    0.49204   0.367  0.71355
## scalelogtotalbulinus     0.06944    0.29519   0.235  0.81402
## ---
## Signif. codes:  0 '***' 0.001 '**' 0.01 '*' 0.05 '.' 0.1 ' ' 1
##
## Correlation of Fixed Effects:
##             (Intr) scllccs sex21  Class  sclppl sclflt scl0.5
## scaleaccssr  0.180
## sex21       -0.214 -0.010
## Class       -0.386 -0.028 -0.077
## scalepopltn -0.069  0.146 -0.013  0.039
```

```
## scaleflotng -0.081 -0.681  0.014  0.047 -0.200
## sclsqkmt0.5 -0.109 -0.438  0.001 -0.044  0.079 -0.044
## scllgttlbln  0.028  0.052 -0.012 -0.041 -0.114 -0.252  0.392
```

```
#drop total bulinus
childmodel_Sm2=glmer(Sm~ scaleaccessarea+sex2+Class+scalepopulation+scalefloating+scalesqkmtotalcrop0.5
summary(childmodel_Sm2)
```

```
## Generalized linear mixed model fit by maximum likelihood (Laplace
##   Approximation) [glmerMod]
##  Family: binomial  ( logit )
## Formula:
## Sm ~ scaleaccessarea + sex2 + Class + scalepopulation + scalefloating +
##     scalesqkmtotalcrop0.5km + (1 | School/ID)
##    Data: dataset6s
## Control: glmerControl(optimizer = "bobyqa", optCtrl = list(maxfun = 1e+05))
##
##      AIC      BIC   logLik deviance df.resid
##   1390.5   1439.6   -686.3   1372.5     1719
##
## Scaled residuals:
##     Min      1Q  Median      3Q     Max
## -1.1009 -0.3838 -0.2846 -0.1866  4.8909
##
## Random effects:
##  Groups    Name        Variance Std.Dev.
##  ID:School (Intercept) 0.3992   0.6319
##  School    (Intercept) 1.4720   1.2133
## Number of obs: 1728, groups:  ID:School, 1289; School, 15
##
## Fixed effects:
##                         Estimate Std. Error z value Pr(>|z|)
## (Intercept)             -2.16330    0.38924  -5.558 2.73e-08 ***
## scaleaccessarea         -0.42026    0.61086  -0.688 0.491463
## sex21                    0.53469    0.14843   3.602 0.000315 ***
## Class                   -0.06504    0.07490  -0.868 0.385192
## scalepopulation          0.09731    0.32851   0.296 0.767078
## scalefloating            0.24563    0.40518   0.606 0.544364
## scalesqkmtotalcrop0.5km  0.13433    0.45192   0.297 0.766282
## ---
## Signif. codes:  0 '***' 0.001 '**' 0.01 '*' 0.05 '.' 0.1 ' ' 1
##
## Correlation of Fixed Effects:
##             (Intr) sclccs sex21  Class  sclppl sclflt
## scaleaccssr  0.179
## sex21       -0.214 -0.010
## Class       -0.386 -0.026 -0.078
## scalepopltn -0.066  0.153 -0.014  0.035
## scaleflotng -0.075 -0.690  0.012  0.038 -0.239
## sclsqkmt0.5 -0.130 -0.499  0.006 -0.031  0.135  0.060
```

```
#drop population
childmodel_Sm3=glmer(Sm~ scaleaccessarea+sex2+Class+scalefloating+scalesqkmtotalcrop0.5km+(1|School/ID)
summary(childmodel_Sm3)
```

```
## Generalized linear mixed model fit by maximum likelihood (Laplace
##   Approximation) [glmerMod]
##  Family: binomial  ( logit )
## Formula:
## Sm ~ scaleaccessarea + sex2 + Class + scalefloating + scalesqkmtotalcrop0.5km +
##     (1 | School/ID)
##    Data: dataset6s
## Control: glmerControl(optimizer = "bobyqa", optCtrl = list(maxfun = 1e+05))
##
##      AIC      BIC   logLik deviance df.resid
##   1388.6   1432.2   -686.3   1372.6     1720
##
## Scaled residuals:
##     Min      1Q  Median      3Q     Max
## -1.1047 -0.3844 -0.2834 -0.1869  4.9380
##
## Random effects:
##  Groups    Name        Variance Std.Dev.
##  ID:School (Intercept) 0.3994   0.6319
##  School    (Intercept) 1.4546   1.2061
## Number of obs: 1728, groups:  ID:School, 1289; School, 15
##
## Fixed effects:
##                          Estimate Std. Error z value Pr(>|z|)
## (Intercept)              -2.15551    0.38660  -5.576 2.47e-08 ***
## scaleaccessarea          -0.45006    0.60148  -0.748 0.454305
## sex21                     0.53540    0.14840   3.608 0.000309 ***
## Class                    -0.06580    0.07484  -0.879 0.379277
## scalefloating             0.27674    0.39285   0.704 0.481154
## scalesqkmtotalcrop0.5km  0.11621    0.44527   0.261 0.794097
## ---
## Signif. codes:  0 '***' 0.001 '**' 0.01 '*' 0.05 '.' 0.1 ' ' 1
##
## Correlation of Fixed Effects:
##             (Intr) sclccs sex21  Class  sclflt
## scaleaccssr  0.190
## sex21       -0.216 -0.007
## Class       -0.387 -0.032 -0.077
## scaleflotng -0.091 -0.682  0.008  0.048
## sclsqkmt0.5 -0.122 -0.530  0.008 -0.036  0.095
```

```
#drop crops
childmodel_Sm4=glmer(Sm~ scaleaccessarea+sex2+Class+scalefloating+(1|School/ID), family=binomial, data=
summary(childmodel_Sm4)
```

```
## Generalized linear mixed model fit by maximum likelihood (Laplace
##   Approximation) [glmerMod]
##  Family: binomial  ( logit )
## Formula: Sm ~ scaleaccessarea + sex2 + Class + scalefloating + (1 | School/ID)
##    Data: dataset6s
## Control: glmerControl(optimizer = "bobyqa", optCtrl = list(maxfun = 1e+05))
##
##      AIC      BIC   logLik deviance df.resid
##   1386.7   1424.8   -686.3   1372.7     1721
```

```
##
## Scaled residuals:
##     Min      1Q  Median      3Q     Max
## -1.1077 -0.3855 -0.2837 -0.1863  4.9091
##
## Random effects:
##  Groups    Name        Variance Std.Dev.
##  ID:School (Intercept) 0.3992   0.6318
##  School    (Intercept) 1.4600   1.2083
## Number of obs: 1728, groups:  ID:School, 1289; School, 15
##
## Fixed effects:
##                 Estimate Std. Error z value Pr(>|z|)
## (Intercept)     -2.14376    0.38410  -5.581 2.39e-08 ***
## scaleaccessarea -0.36622    0.51006  -0.718  0.47276
## sex21            0.53515    0.14838   3.607  0.00031 ***
## Class           -0.06504    0.07478  -0.870  0.38443
## scalefloating    0.26648    0.39065   0.682  0.49516
## ---
## Signif. codes:  0 '***' 0.001 '**' 0.01 '*' 0.05 '.' 0.1 ' ' 1
##
## Correlation of Fixed Effects:
##             (Intr) sclccs sex21  Class
## scaleaccssr  0.149
## sex21       -0.216 -0.003
## Class       -0.394 -0.060 -0.077
## scaleflotng -0.080 -0.748  0.008  0.052
```

```
#drop floating
childmodel_Sm5=glmer(Sm~ scaleaccessarea+sex2+Class+(1|School/ID), family=binomial, data=dataset6s,na.a
summary(childmodel_Sm5)
```

```
## Generalized linear mixed model fit by maximum likelihood (Laplace
##   Approximation) [glmerMod]
##  Family: binomial  ( logit )
## Formula: Sm ~ scaleaccessarea + sex2 + Class + (1 | School/ID)
##    Data: dataset6s
## Control: glmerControl(optimizer = "bobyqa", optCtrl = list(maxfun = 1e+05))
##
##      AIC      BIC   logLik deviance df.resid
##   1385.1   1417.8   -686.5   1373.1     1722
##
## Scaled residuals:
##     Min      1Q  Median      3Q     Max
## -1.0968 -0.3825 -0.2834 -0.1868  4.5558
##
## Random effects:
##  Groups    Name        Variance Std.Dev.
##  ID:School (Intercept) 0.4003   0.6327
##  School    (Intercept) 1.4464   1.2027
## Number of obs: 1728, groups:  ID:School, 1289; School, 15
##
## Fixed effects:
##                 Estimate Std. Error z value Pr(>|z|)
```

```
## (Intercept)     -2.12364    0.38145  -5.567 2.59e-08 ***
## scaleaccessarea -0.10820    0.33755  -0.321 0.748559
## sex21            0.53459    0.14837   3.603 0.000315 ***
## Class           -0.06780    0.07467  -0.908 0.363914
## ---
## Signif. codes:  0 '***' 0.001 '**' 0.01 '*' 0.05 '.' 0.1 ' ' 1
##
## Correlation of Fixed Effects:
##              (Intr) sclccs sex21
## scaleaccssr  0.133
## sex21       -0.217  0.003
## Class       -0.393 -0.032 -0.077
```

```
#drop access area:
childmodel_Sm6=glmer(Sm~ sex2+Class+(1|School/ID), family=binomial, data=dataset6s,na.action=na.omit,con
summary(childmodel_Sm6)
```

```
## Generalized linear mixed model fit by maximum likelihood (Laplace
##   Approximation) [glmerMod]
##  Family: binomial  ( logit )
## Formula: Sm ~ sex2 + Class + (1 | School/ID)
##    Data: dataset6s
## Control: glmerControl(optimizer = "bobyqa", optCtrl = list(maxfun = 1e+05))
##
##      AIC      BIC   logLik deviance df.resid
##   1383.2   1410.5   -686.6   1373.2     1723
##
## Scaled residuals:
##     Min      1Q  Median      3Q     Max
## -1.0958 -0.3812 -0.2828 -0.1879  4.5317
##
## Random effects:
##  Groups    Name        Variance Std.Dev.
##  ID:School (Intercept) 0.4003   0.6327
##  School    (Intercept) 1.4658   1.2107
## Number of obs: 1728, groups:  ID:School, 1289; School, 15
##
## Fixed effects:
##             Estimate Std. Error z value Pr(>|z|)
## (Intercept) -2.10847    0.37989  -5.550 2.85e-08 ***
## sex21        0.53489    0.14838   3.605 0.000312 ***
## Class       -0.06856    0.07464  -0.919 0.358332
## ---
## Signif. codes:  0 '***' 0.001 '**' 0.01 '*' 0.05 '.' 0.1 ' ' 1
##
## Correlation of Fixed Effects:
##       (Intr) sex21
## sex21 -0.219
## Class -0.391 -0.077
```

```
#drop class
childmodel_Sm7=glmer(Sm~ sex2+(1|School/ID), family=binomial, data=dataset6s,na.action=na.omit,control=g
summary(childmodel_Sm7)
```

```
## Generalized linear mixed model fit by maximum likelihood (Laplace
##   Approximation) [glmerMod]
##  Family: binomial  ( logit )
## Formula: Sm ~ sex2 + (1 | School/ID)
##    Data: dataset6s
## Control: glmerControl(optimizer = "bobyqa", optCtrl = list(maxfun = 1e+05))
##
##      AIC      BIC   logLik deviance df.resid
##   1382.0   1403.8   -687.0   1374.0     1724
##
## Scaled residuals:
##     Min      1Q  Median      3Q     Max
## -1.0792 -0.3908 -0.2896 -0.1920  4.2904
##
## Random effects:
##  Groups    Name        Variance Std.Dev.
##  ID:School (Intercept) 0.4006   0.633
##  School    (Intercept) 1.4861   1.219
## Number of obs: 1728, groups:  ID:School, 1289; School, 15
##
## Fixed effects:
##             Estimate Std. Error z value Pr(>|z|)
## (Intercept)  -2.2492     0.3516  -6.398 1.58e-10 ***
## sex21         0.5253     0.1479   3.553 0.000381 ***
## ---
## Signif. codes:  0 '***' 0.001 '**' 0.01 '*' 0.05 '.' 0.1 ' ' 1
##
## Correlation of Fixed Effects:
##       (Intr)
## sex21 -0.269
```

```
AIC(childmodel_Sm,childmodel_Sm2,childmodel_Sm3,childmodel_Sm4,childmodel_Sm5,childmodel_Sm6,childmodel_
```

```
##                 df      AIC
## childmodel_Sm  10 1392.456
## childmodel_Sm2  9 1390.509
## childmodel_Sm3  8 1388.598
## childmodel_Sm4  7 1386.666
## childmodel_Sm5  6 1385.093
## childmodel_Sm6  5 1383.194
## childmodel_Sm7  4 1381.995
```

```
#Create a figure showing agriculture at lakes versus rivers:
landp1 <- read.csv("landuse_stanford_and_our7village_weightedmeans_1_13.csv")

report1=ddply(landp1,c("waterway"),summarize,
               N=length(sqkmtotalcrop0.5km),
               avg=mean(sqkmtotalcrop0.5km,na.rm=TRUE),
               sd=sd(sqkmtotalcrop0.5km,na.rm=TRUE),
               se=sd/sqrt(N),
               ucl=avg+1.98*se,
               lcl=avg-1.98*se)
```

```
#Plot of sqkmtotalcrop0.5km  with 95%CI:
plot1 <- ggplot(data=report1, aes(x=waterway, y=avg))+
  ggtitle("")+ ylab("Avg. total crop cover (sq. km) in 0.5km w/ 95% CI")+ xlab("")+
  geom_point(position=position_dodge(w=0.1), size=3.4)+
  geom_errorbar(aes(ymax=ucl, ymin=lcl), width=0.4, position=position_dodge(w=0.1))+
  theme_bw() +
  theme(axis.line = element_line(colour = "black"),
        panel.grid.major = element_blank(),
        panel.grid.minor = element_blank(),
        panel.border = element_blank(),
        panel.background = element_blank()) +theme(axis.text=element_text(size=16), axis.title=element_
plot1
```

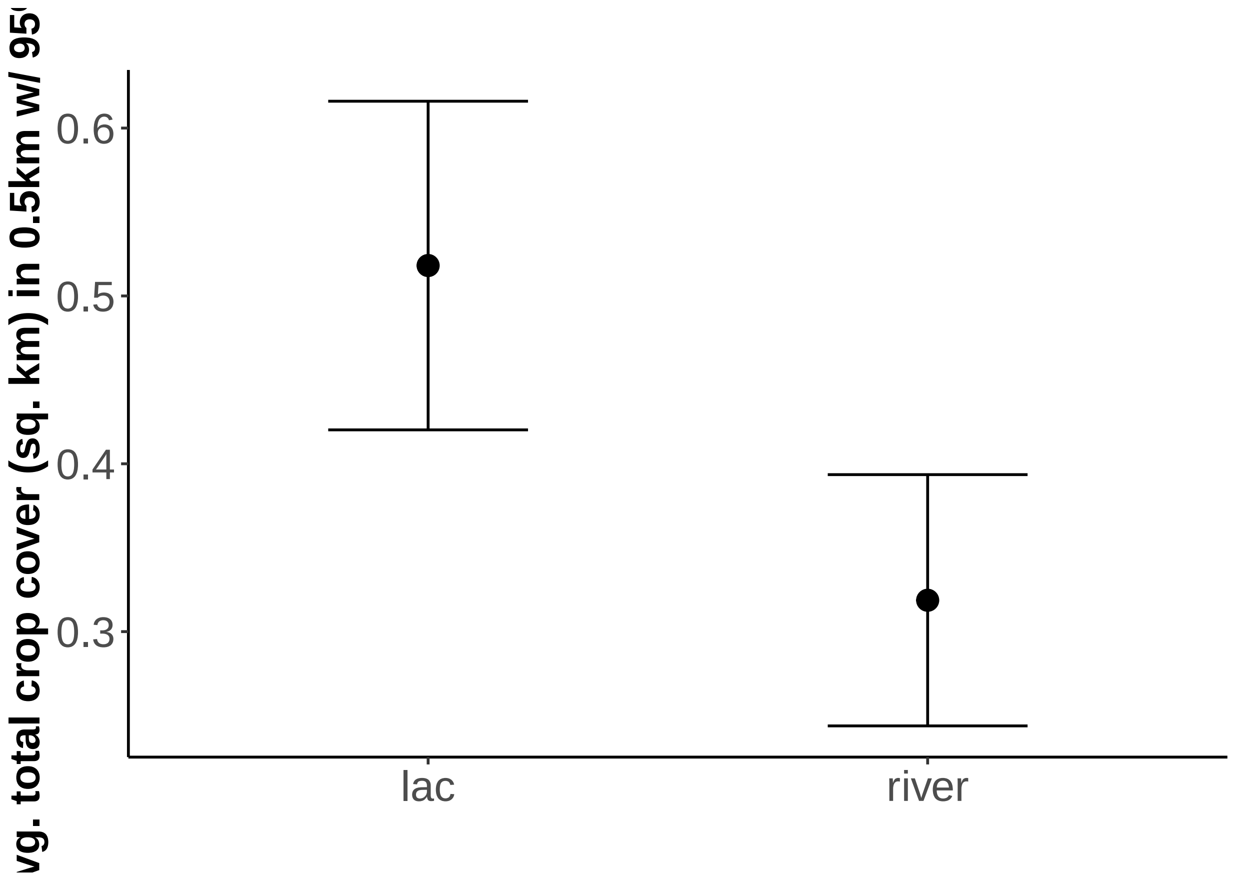


```
#try the same but for ha total of referenced fields
report1.2=ddply(landp1,c("waterway"),summarize,
               N=length(ha.total.of.referenced.fields),
               avg=mean(ha.total.of.referenced.fields,na.rm=TRUE),
               sd=sd(ha.total.of.referenced.fields,na.rm=TRUE),
               se=sd/sqrt(N),
               ucl=avg+1.98*se,
               lcl=avg-1.98*se)

#Plot of ha.total.of.referenced.fields  with 95%CI:
plot1.2 <- ggplot(data=report1.2, aes(x=waterway, y=avg))+
  ggtitle("")+ ylab("Avg. total crop cover (ha) in 0.5km w/ 95% CI")+ xlab("")+
```

```
  geom_point(position=position_dodge(w=0.1), size=3.4)+
  geom_errorbar(aes(ymax=ucl, ymin=lcl), width=0.4, position=position_dodge(w=0.1))+
  theme_bw() +
  theme(axis.line = element_line(colour = "black"),
        panel.grid.major = element_blank(),
        panel.grid.minor = element_blank(),
        panel.border = element_blank(),
        panel.background = element_blank()) +theme(axis.text=element_text(size=16),
                                                                          axis.title=element_text(size=
plot1.2
```

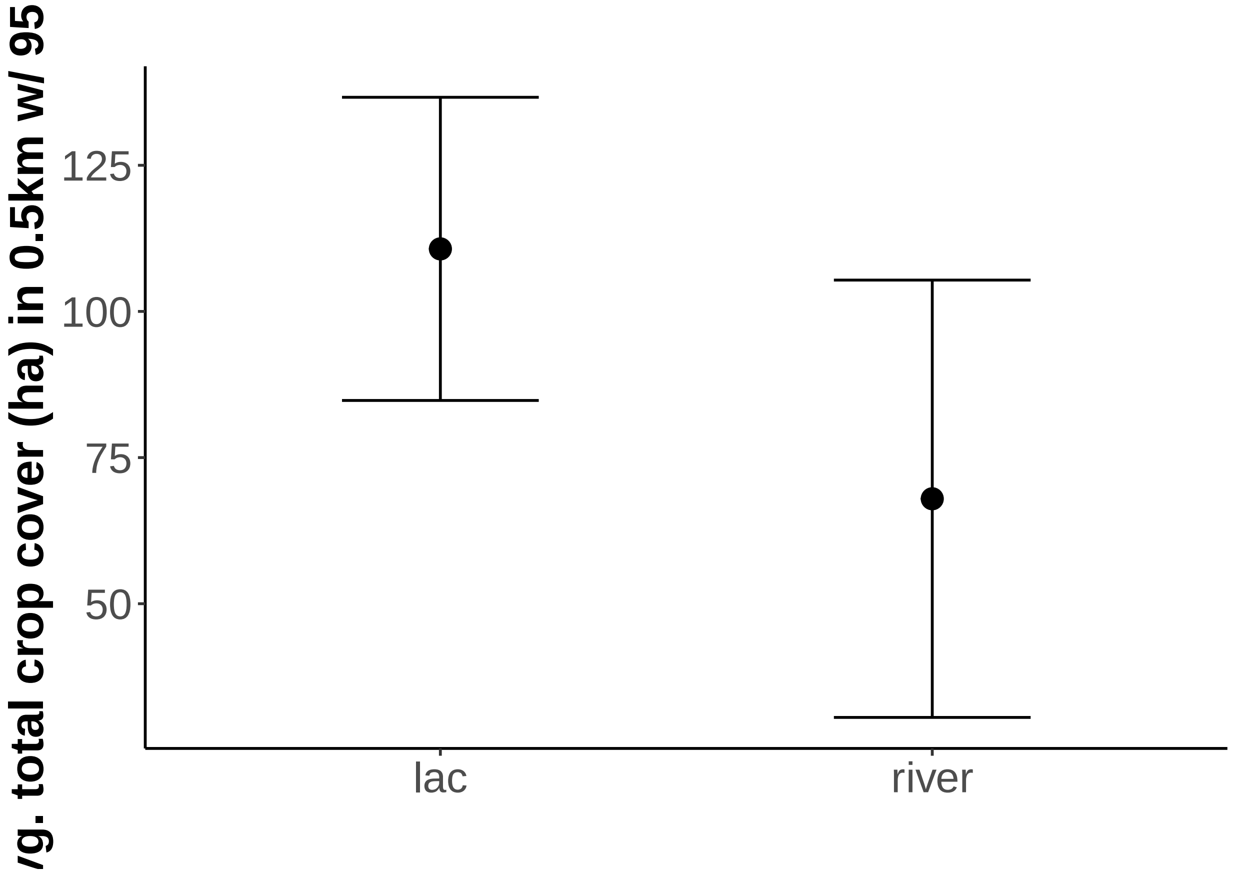


```
#Similar trend but 7 less villages = not significant.
```

```
#########################
#snail methods comparison:
#########################

#Fig S4

snailsdat=read.csv("snail_points_USF_Stanford.csv")

bul1 =ddply(snailsdat,c("id"),summarize,
            N=length(bulinus),
            avg=mean(bulinus,na.rm=TRUE),
            sd=sd(bulinus,na.rm=TRUE),
```

```
           se=sd/sqrt(N),
           ucl=avg+1.98*se,
           lcl=avg-1.98*se)

plotbul <- ggplot(data=bul1, aes(x=id, y=avg))+
  ggtitle("")+ ylab("Avg. Bul. snails w/ 95% CI")+ xlab("")+
  geom_point(position=position_dodge(w=0.1), size=3.4)+
  geom_errorbar(aes(ymax=ucl, ymin=lcl), width=0.4, position=position_dodge(w=0.1))+
  theme_bw() +
  theme(axis.line = element_line(colour = "black"),
        panel.grid.major = element_blank(),
        panel.grid.minor = element_blank(),
        panel.border = element_blank(),
        panel.background = element_blank())
plotbul
```

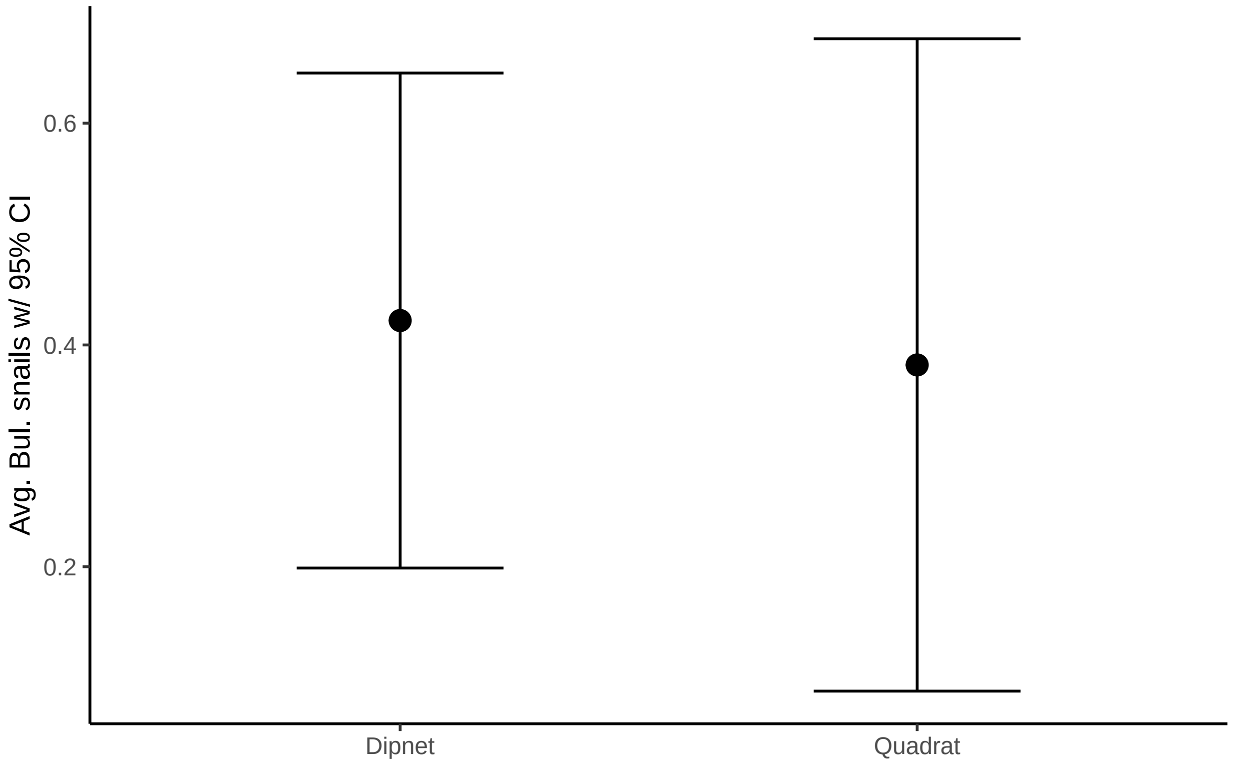


```
#no sig diff
biom1 =ddply(snailsdat,c("id"),summarize,
             N=length(Biomphalaria),
             avg=mean(Biomphalaria,na.rm=TRUE),
             sd=sd(Biomphalaria,na.rm=TRUE),
             se=sd/sqrt(N),
             ucl=avg+1.98*se,
             lcl=avg-1.98*se)
plotbiom <- ggplot(data=biom1, aes(x=id, y=avg))+
```

```
  ggtitle("")+ ylab("Avg. Biom. snails w/ 95% CI")+ xlab("")+
  geom_point(position=position_dodge(w=0.1), size=3.4)+
  geom_errorbar(aes(ymax=ucl, ymin=lcl), width=0.4, position=position_dodge(w=0.1))+
  theme_bw() +
  theme(axis.line = element_line(colour = "black"),
        panel.grid.major = element_blank(),
        panel.grid.minor = element_blank(),
        panel.border = element_blank(),
        panel.background = element_blank())
plotbiom
```

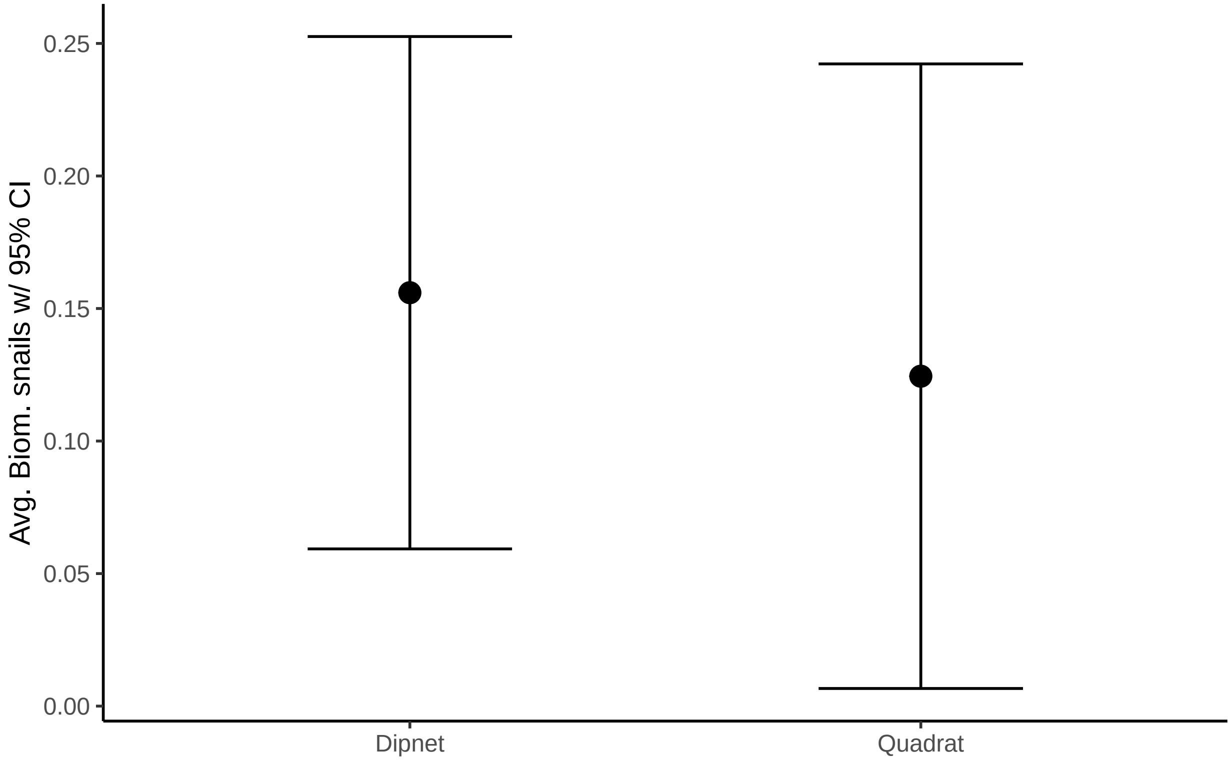


```
#no sig diff
```

```
#ZINB were favored by AIC over NB
nbulc = glmmTMB(round(bulinus) ~ id+Mass  + (1|UniqueSiteQuad), data=snailsdat, na.action=na.omit, famil
Anova(nbulc)
```

```
## Analysis of Deviance Table (Type II Wald chisquare tests)
##
## Response: round(bulinus)
##       Chisq Df Pr(>Chisq)
## id   1.4666  1     0.2259
## Mass 0.3254  1     0.5684
```

```
#re-run without zero inflation:
nbulc2 = glmmTMB(round(bulinus) ~ id+Mass+(1|UniqueSiteQuad), data=snailsdat, na.action=na.omit, family=
Anova(nbulc2)
```

```
## Analysis of Deviance Table (Type II Wald chisquare tests)
##
## Response: round(bulinus)
##        Chisq Df Pr(>Chisq)
## id    0.9175  1     0.3381
## Mass 21.8595  1  2.934e-06 ***
## ---
## Signif. codes:  0 '***' 0.001 '**' 0.01 '*' 0.05 '.' 0.1 ' ' 1
```

```
  visreg(nbulc2,"id",scale="response")
```

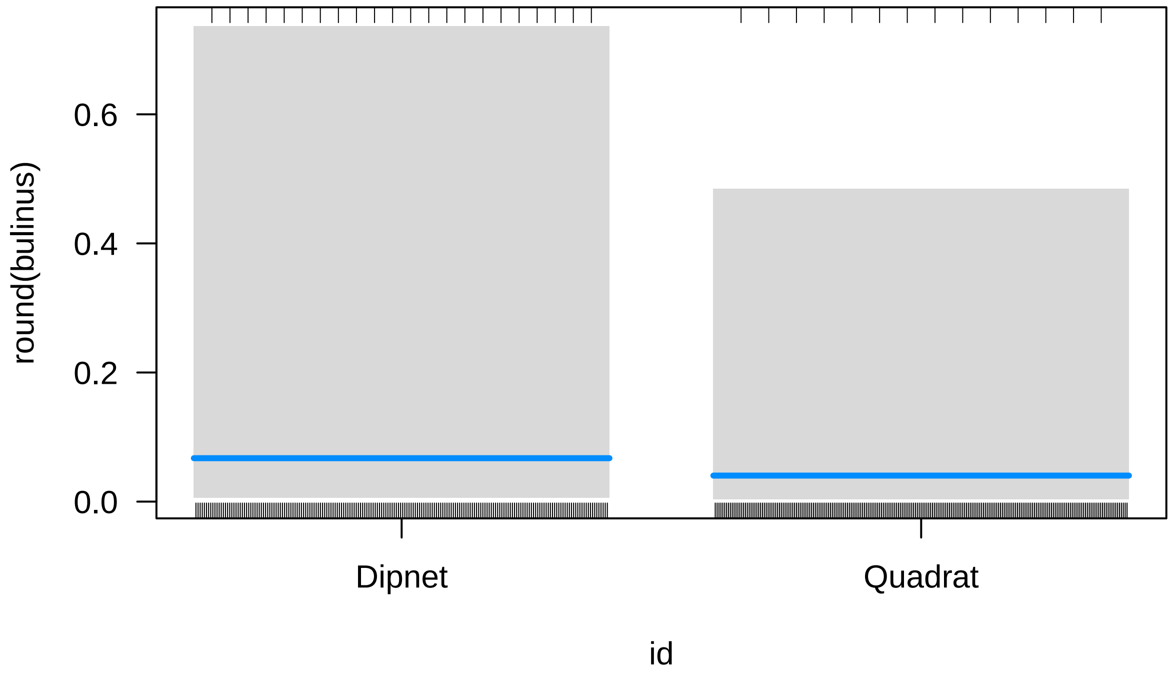


```
nbulc3 = glmmTMB(round(bulinus) ~ id+(1|Site.ID/UniqueSiteQuad), data=snailsdat, na.action=na.omit, fam
```

```
## Warning in fitTMB(TMBStruc): Model convergence problem; non-positive-definite
## Hessian matrix. See vignette('troubleshooting')
```

```
Anova(nbulc3)
```

```
## Analysis of Deviance Table (Type II Wald chisquare tests)
```

```
##
## Response: round(bulinus)
##     Chisq Df Pr(>Chisq)
## id 0.7612  1      0.383
```

```
nbiom1 = glmmTMB(round(Biomphalaria) ~ id+Mass  + (1|UniqueSiteQuad), data=snailsdat, na.action=na.omit
Anova(nbiom1)
```

```
## Analysis of Deviance Table (Type II Wald chisquare tests)
##
## Response: round(Biomphalaria)
##       Chisq Df Pr(>Chisq)
## id   0.1263  1     0.7223
## Mass 0.0466  1     0.8290
```

```
  nbiom = glmmTMB(round(Biomphalaria) ~ id+Mass  + (1|UniqueSiteQuad), data=snailsdat, na.action=na.omi
Anova(nbiom)
```

```
## Analysis of Deviance Table (Type II Wald chisquare tests)
##
## Response: round(Biomphalaria)
##       Chisq Df Pr(>Chisq)
## id   0.3383  1    0.56082
## Mass 4.2217  1    0.03991 *
## ---
## Signif. codes:  0 '***' 0.001 '**' 0.01 '*' 0.05 '.' 0.1 ' ' 1
```
